# Update of the ADNEX risk model (ADNEX2) to diagnose ovarian cancer in surgically and conservatively managed patients with an adnexal mass: multicentre prospective cohort study

**DOI:** 10.64898/2026.10.23.26365247

**Authors:** Lasai Barreñada, Wouter Froyman, Jolien Ceusters, Chiara Landolfo, Laure Wynants, Ashleigh Ledger, Povilas Sladkevicius, Antonia C Testa, Caroline Van Holsbeke, Francesca Buonomo, Marek J Kudla, Robert Fruscio, Daniela Fischerova, Dorella Franchi, Elisabeth Epstein, Valentina Chiappa, Francesco PG Leone, Joie Ensor, Maria Elisabetta Coccia, Stefano Guerriero, Juan Luis Alcazar, Fatima Alves, Ligita Jokubkiene, Federica Pozzati, Francesca Moro, Floriana Mascilini, Cristina Taliento, Tom Bourne, Lil Valentin, Dirk Timmerman, Ben Van Calster

## Abstract

**Objective:** The ADNEX ultrasound**-**based prediction model estimates the probability that an adnexal mass is benign, borderline, stage I primary invasive, stage II-IV primary invasive, or secondary metastatic. ADNEX was developed on patients managed surgically. We aimed to update ADNEX to ADNEX2 using more recent data on both surgically and conservatively managed patients. ADNEX2 is designed for use in patients in whom the benign descriptors (BDs), a set of ultrasound profiles with an estimated risk of malignancy below 1%, do not apply.

**Design:** Secondary analysis of a multicentre prospective cohort study.

**Setting:** 17 oncology and non-oncology centres from 8 European countries.

**Participants:** Patients with a newly detected adnexal mass recruited between 2012 and 2016 who were managed surgically or conservatively with clinical and ultrasound follow-up.

**Interventions:** ADNEX2 uses the same predictors as ADNEX (age, type of centre, lesion diameter, proportion of solid tissue, number of papillary projections, >10 cyst locules, ascites and acoustic shadows and CA125). Model updating involved logistic recalibration followed by Bayesian refitting. Validation was based on leave-one-centre-out cross-validation. ADNEX2 and ADNEX include CA125 but also work without CA125.

**Main outcome measures:** Model performance regarding discrimination (AUC), calibration (plots and observed:expected, O:E, ratio), and clinical utility (net benefit).

**Results:** Of 6847 patients, 4625 (68%) had a benign tumour, 1710 (25%) had a malignant tumour, while tumour outcome was uncertain for 512 (7%) patients. BDs applied in 2190 (32%) patients, of whom 0.5% (95% CI, 0.2%-0.9%) had a malignancy. A two-step strategy using BDs followed by ADNEX2 had an AUC of 0.95 (0.94-0.96) to distinguish benign from malignant tumours. When evaluated on 4657 patients in whom BDs did not apply, ADNEX2 had an AUC of 0.93 (0.91-0.94). The two-step strategy with ADNEX2 was better calibrated than the two-step strategy with ADNEX (O:E ratio of 1.05 versus 1.14) and had marginally higher clinical utility. Not including available CA125 values in ADNEX2 reduced the ability to distinguish stage II-IV primary invasive tumours from other invasive malignancies.

**Conclusions:** ADNEX2 is better calibrated than ADNEX. CA125 measurement is not crucial to estimate the risk of malignancy, but is recommended to estimate the risk of malignant subtypes.

**Trial registration:** clinicaltrials.gov NCT01698632.

**What is already known on this topic:**

- The IOTA ADNEX risk model is widely used to estimate the probability that an adnexal mass is benign or one of four malignant subtypes, and has been integrated into a two-step strategy following the application of ultrasound benign descriptors (BDs).
- Like nearly all prediction models for adnexal masses, the original ADNEX model was developed exclusively using data from patients who underwent surgery (recruited between 1999 and 2012). This introduces selection bias when the model is used to determine who should be referred for specialized care.
- Measuring serum CA125 is not essential for estimating overall malignancy risk, but is recommended when patients are referred to oncology centres to distinguish stage II-IV primary invasive cancer from stage I primary invasive cancer and secondary metastases.

**What this study adds:**

- ADNEX was updated to ADNEX2 using more recent multicentre data (IOTA5) including both surgically and conservatively managed patients with tumours to which benign descriptors did not apply.
- ADNEX2 (used alone or in a two-step strategy after benign descriptors) maintains high discrimination (AUC 0.95 for two-step strategy with ADNEX2; 0.93 when benign descriptors do not apply), improves risk calibration compared with the original ADNEX model (O:E ratio 1.05 v 1.14), and offers marginally higher clinical utility.

## 1 Introduction

When confronted with an ovarian tumour, clinical staff must decide on the appropriate management. The optimal management depends on the type of tumour. Many benign tumours can be managed conservatively with clinical and ultrasound follow-up, whereas different types of malignant tumours require different treatment [1,2]. The Assessment of Different NEoplasias in the adneXa (ADNEX) model developed by the International Ovarian Tumour Analysis (IOTA) group is a statistical model to estimate the likelihood that an adnexal mass is malignant. If malignant it subclassifies the mass as borderline malignant, stage I primary invasive, stage II-IV primary invasive, or a secondary metastasis [3]. ADNEX includes three clinical and six ultrasound variables. It has one version with serum CA125 as a predictor and another one without. The other predictors are patient’s age, type of centre at which the ultrasound examination was performed (oncology centre vs other) and the following ultrasound variables: maximum diameter of the lesion, proportion of solid tissue, number of papillary projections, more than ten cyst locules, ascites, and acoustic shadows. Type of centre is included to acknowledge differences in prevalence of malignant tumours between centres.

ADNEX is intended to be used by clinicians after ultrasound examination of a patient with an adnexal mass to assess the risk of the mass being malignant. A systematic review of the performance of ADNEX on external validation reported an area under the receiver operating characteristic (AUC) curve of 0.93 both for the version with and without CA125 and reported very good clinical utility at the 10% risk of malignancy threshold to decide which patients to refer to an oncology centre [4].

Despite its very good performance, ADNEX can potentially be improved. First, the model was developed using patients recruited between 1999 and 2012 (IOTA phases 1-3), i.e., 14 to 27 years ago. Patient populations may change over time, and this may negatively affect model performance [5]. Second, like nearly all other models in the field, ADNEX was developed using information only from patients that underwent surgery. While surgery allows tumour outcome to be based on a reliable reference standard (histopathology), it suffers from selection bias by not including ovarian masses that are followed conservatively. This is problematic, because ADNEX was developed to help clinicians decide whether conservative management without surgery could be an option (low risk of malignancy), while the development studies excluded patients where the decision was not to operate. Third, a large proportion of tumours can be classified as benign by using the IOTA Benign Descriptors (BDs) instead of a statistical model. The BDs are a set of clinical and ultrasound profiles that classify the mass as almost certainly benign (**Box**) [6]. A two-step strategy has been proposed, where the BDs are used first, followed by ADNEX if BDs do not apply [6]. It makes sense to update ADNEX using data from patients with masses to which BDs do not apply, because such an updated model should be better suited for use in a two-step strategy, where application of the BDs is the first step.

The objectives of this study were to update the ADNEX model using recent data from patients with tumours to which the BDs do not apply and using information from both surgically and conservatively managed patients (ADNEX2), to validate ADNEX2 and a two-step strategy using it, and to compare the performance of ADNEX2 with that of ADNEX whether used alone or in a two-step strategy.

### Box. The International Ovarian Tumor Analysis (IOTA) benign descriptors

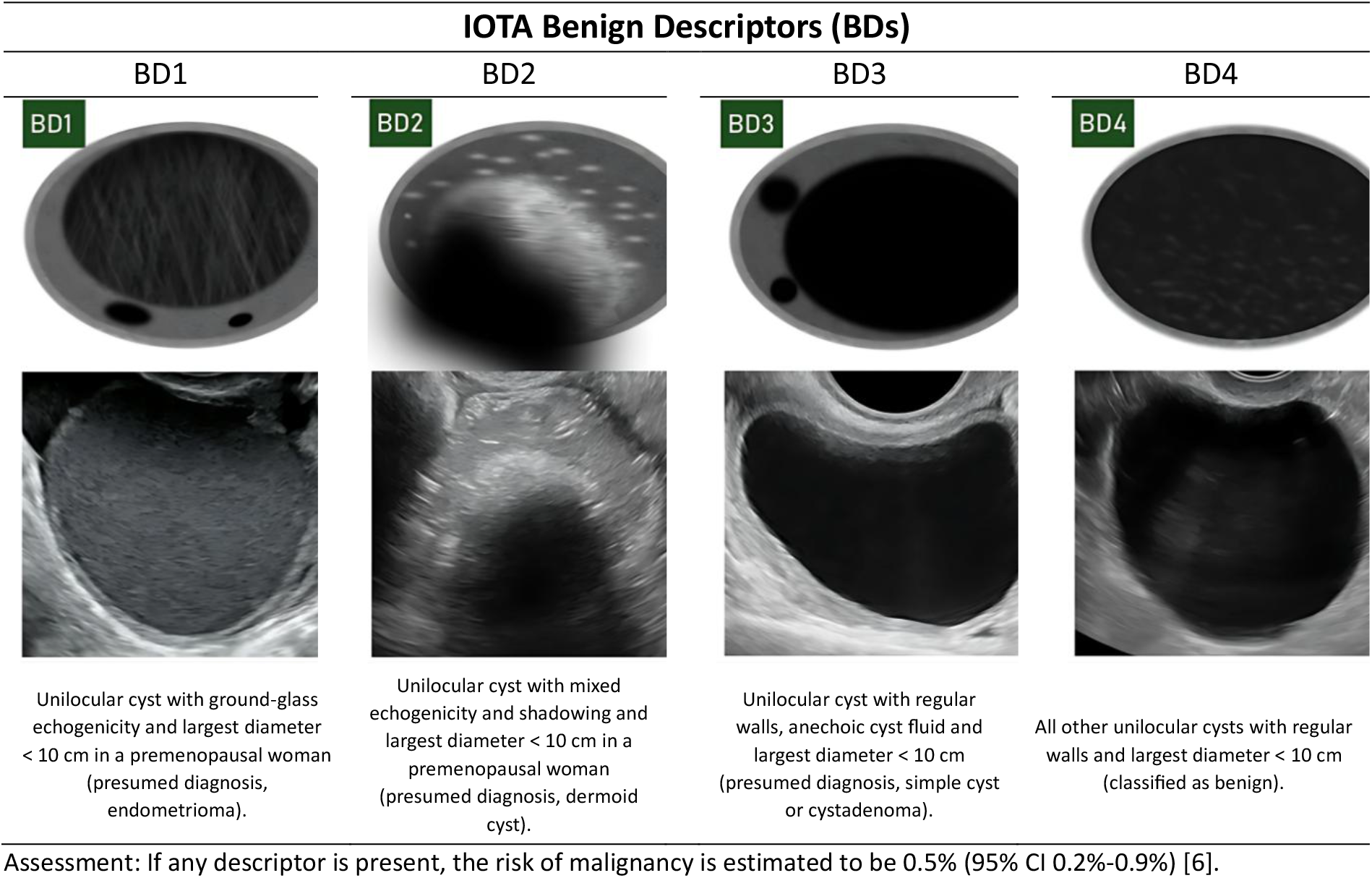

## 2 Methods

### 2.1 Design and participants

We used data from the IOTA5 study, an ongoing multicentre prospective cohort study including patients with an adnexal mass managed surgically or conservatively with clinical and ultrasound follow-up. Patients were recruited between 1^st^ January 2012 and 31^st^ December 2016. Conservatively managed patients were followed up until 31^st^ December 2022. Inclusion criteria were being at least 18 years old at recruitment and presenting with at least one adnexal mass (ovarian, para-ovarian, or tubal). Exclusion criteria were having a lesion presumed to be physiological with a largest diameter less than 3 cm, or refusal or withdrawal of informed consent. Pregnancy was not an exclusion criterion. Patients with a mass that was already in follow-up at a centre before recruitment into IOTA5 (i.e. not newly detected masses) were also excluded for this analysis.

Patients were recruited at 39 oncology and non-oncology centres across 15 countries, with oncology centres defined as centres with a dedicated gynaecologic oncology unit (**Table S1).** For this study, we use data from 6847 patients recruited at 17 centres. These 17 centres satisfied predefined quality criteria and aimed to consecutively recruit both patients selected for surgery and for management with clinical and ultrasound follow-up. The predefined quality criteria and characteristics of included and excluded centres are described in **Supplementary material S1**. The IOTA5 study was approved by the Research Ethics Committee of the University Hospitals Leuven as the coordinating centre (B32220095331/S51375) and by local ethics committees of all participating centres. We report the study according to the TRIPOD+AI and TRIPOD-Cluster guidelines (**Supplementary material S6**) [7,8].

### 2.2 Procedures and variable measurements

Patients were recruited by gynaecologists with expertise in gynaecological ultrasound. They were either IOTA trainers or had passed the IOTA certification test (https://www.gynaia.com/certified-professionals). When recruiting, the ultrasound examiners followed a standardised research protocol (**Supplementary Material S7**). Standardised clinical information was collected, and a standardised transvaginal ultrasound examination was performed supplemented with abdominal ultrasound if necessary. The recruitment examination included scanning the uterus, both adnexa, and the whole pelvis outside these organs. Grey scale and colour or power Doppler ultrasound was used to characterise the morphology and vascularity of each adnexal mass. Information on a set of predefined ultrasound variables, including those used in the ADNEX model, was collected, and the IOTA terminology was used to describe the ultrasound findings [9]. In addition, the ultrasound examiners provided a diagnosis based on their subjective assessment by classifying each lesion as benign, borderline, or malignant, and by specifying the degree of certainty of their classification (certain, probable, or uncertain). The ultrasound examiner also suggested a specific diagnosis based on pattern recognition using a predefined list of 18 histological diagnoses (11 benign, six malignant, and “not possible”) [10].

When multiple adnexal masses were found on ultrasound, the dominant mass was included in the statistical analysis. The dominant mass was the mass with the most complex ultrasound morphology. If multiple masses had similar morphology, the largest mass or the mass that was most easily accessible with ultrasound was denoted dominant. We encouraged centres to measure the patients‘ serum CA125 level, but this was not a requirement for inclusion in the study. Measurement of CA125 was left to clinical judgement and local protocols. Due to the prospective study design, predictor measurements were blinded to tumour outcome.

Based on patient history, clinical and ultrasound findings, the ultrasound examiner suggested surgery or conservative management. The final decision on management was made by the treating physician in dialogue with the patient. Conservative management included ultrasound and clinical follow-up at intervals of three months, six months, 12 months and then every 12 months thereafter. At every follow-up visit, clinical and ultrasound information was collected using same approach as for the inclusion scan. At some point during follow-up, some patients underwent surgery for a variety of reasons (e.g., suspicion of malignancy, pain or patient request) [1]. Each centre performed surgery by following local protocols. Surgically removed masses were examined histologically at the local centre. For some patients, the mass resolved spontaneously during follow-up.

Historically, ADNEX was categorized as “with” or “without” CA125. In this work we refine this terminology, using “ADNEX” and “ADNEX2” to represent models where coefficient sets depend on CA125 availability. We also replace “without CA125” with “ignoring CA125,” as some patients in our cohort actually had these measurements. To mirror clinical practice, we do not impute missing CA125 values during validation; rather, the coefficients applied in the models and the two-step strategy vary per patient based on whether CA125 data is present.

### 2.3 Outcome

The primary outcome is classification of tumours as benign or malignant at inclusion. The secondary outcome is the classification as benign, borderline, stage I primary invasive, Stage II-IV primary invasive, or secondary metastasis at inclusion. The reference standard is histology for patients who underwent surgery. If surgery was not performed, the reference standard was clinical information and subjective assessment of ultrasound images at inclusion and during follow-up until 12 (+/-2) months [11]. If there was insufficient information to make a reasonable classification of the nature of the mass at inclusion the outcome was classified as uncertain. **Table 1** shows the classification criteria.

**Table 1.** Tumour outcome for 6847 included patients.

| **Outcome and reason** | **n** |
| --- | --- |
| **Benign** | **4625** |
| - Spontaneous resolution | 2961 |
| - No surgery, no spontaneous resolution, last visit $\geq 10$ months, SA at every visit up to 10-14 months was probably benign or certainly benign | 1033 |
| - Surgery, benign histology | 631 |
| **Borderline** | **391** |
| - Surgery within 120 days, borderline histology | 378 |
| - Surgery after 120 days, borderline histology, SA at every visit up to surgery was probably borderline or certainly borderline. | 13 |
| **Stage I invasive** | **303** |
| - Surgery within 120 days, stage I invasive histology | 299 |
| - Surgery after 120 days, stage I invasive histology, SA at every visit up to surgery was probably borderline/malignant or certainly borderline/malignant. | 4 |
| **Stage II-IV invasive** | **787** |
| - Surgery within 120 days, stage II-IV invasive histology | 772 |
| - Surgery after 120 days, stage II-IV invasive histology, SA at every visit up to surgery was probably borderline/malignant or certainly borderline/malignant. | 15 |
| **Secondary metastatic** | **229** |
| - Surgery within 120 days, secondary metastatic histology | 221 |
| - Surgery after 120 days, secondary metastatic histology, SA at every visit up to surgery was probably borderline/malignant or certainly borderline/malignant | 8 |
| **Uncertain but malignant** | **5\*** |
| - No surgery, no spontaneous resolution, last visit $\geq 10$ months, SA at every visit up to 10-14 months was probably borderline/malignant or certainly borderline/malignant. | 5 |
| **Uncertain** | **507** |
| - No surgery, no spontaneous resolution, last follow-up visit was before 10 months (owing to death, withdrawal from study, or lost to follow-up) | 135 |
| - No information after the inclusion visit | 293 |
| - No surgery, no spontaneous resolution, last visit $\geq 10$ months, SA was uncertain or was inconsistent across visits up to 10-14 months | 45 |
| - Surgery after 120 days, malignant histology, SA not probably borderline/malignant or certainly borderline/malignant at every visit up to surgery | 33 |
| - Histology (surgery within 120 days), outcome unknown | 1 |

SA: Subjective assessment.
\* These tumours were classified using the most common multiple imputed outcome.

Pathologists were blinded to ultrasound predictor variables and model predictions but might have received information on subjective assessment by the ultrasound examiner when clinically relevant. Borderline tumours were classified as malignant. Central pathology review was not carried out because in a previous study we did not observe important differences in reported outcomes between local and central pathology reports [12]. The stage of the malignant tumours was determined using the criteria of the International Federation of Gynaecology and Obstetrics (FIGO) [13].

### 2.4 Data cleaning and preparation

Patient data was collected using a study-specific secure electronic platform (Astraia software, Munich, Germany) with encrypted data communication to ensure data security. From 30 May 2023, follow-up data were collected via Clinical Data Miner (CDM), which provides similar data security [14]. Data from both databases are used in our analyses. A team of biostatisticians and ultrasound examiners performed data cleaning through regular meetings and by sending queries to participating centres to retrieve missing information or to correct inconsistencies.

### 2.5 Statistical analysis

We compared the available sample size with the sample size criteria for multinomial logistic regression prediction models of Pate et al [15] (details in **Supplementary material S2.1)**. We followed a prespecified statistical analysis plan (SAP) preregistered in Open Science Framework (https://osf.io/ypn7r). Deviations from the SAP are summarized in **Supplementary Material S3**.

There were missing values for CA125 and for the tumour outcomes labelled uncertain (**Table 1**). These were handled with multiple imputation. **Supplementary material S2.2 provides** details on how we handled missing data.

ADNEX was updated on the dataset of patients with tumours to which the BDs did not apply. We used the following procedure to obtain ADNEX2. We used the same predictors and transformations (for continuous predictors) as in ADNEX. First, we fitted a multinomial logistic recalibration model to update intercepts and coefficients using slopes [16]. In the second step, we used Bayesian refitting using information from the original ADNEX model and the logistic recalibration [17,18]. In both steps, we used models with random intercepts per centre [19]. Details on the model updating procedure are presented in **supplementary material S2.3.1.** We applied this approach separately for ADNEX without CA125 and ADNEX with CA125 as a predictor.

To use the two-step strategy as a risk model, patients with tumours to which the BDs applied were also assigned risk estimates for the five outcome categories. These estimates were obtained as the prevalence of each outcome category in the subgroup of patients with tumours to which the BDs applied, averaged over 100 imputed datasets.

Whereas the final ADNEX2 model is obtained using data from all patients for whom the BDs did not apply, the two-step strategy (BDs followed by ADNEX2 if BDs do not apply) and ADNEX2 alone were validated using leave-one-center-out cross-validation to estimate how the two-step strategy and ADNEX2 alone, also ignoring CA125, would perform in new centres [20,21]. This means that the approaches to update ADNEX to ADNEX2 and to generate risk estimates when the BDs apply were implemented anew on data from 16 centres and tested on data from the remaining centre. Leave-one-center-out cross-validation mimics geographical external validation by treating every centre once as a test set for models developed on the remaining 16 centres.

We present centre-specific results of the leave-one-center-out cross-validation procedure, as well as overall results using meta-analysis of centre-specific results. 95% confidence intervals (CI) or credible intervals (CrI) are reported. Heterogeneity in performance between centres is quantified with prediction intervals (PI) [22]. We report diagnostic performance for distinguishing benign from malignant tumours as follows: area under the Receiver Operating Characteristic curve (AUC), flexible centre-specific and overall calibration plots for the overall estimated risk of malignancy, and net benefit with decision curves. In an international consensus statement it was suggested to use a decision threshold of 1% to decide between conservative management and surgery and a decision threshold of 10% to decide whether the patient should be referred for specialized oncological care [2]. To acknowledge that not everyone may agree with these thresholds, we report net benefit for decision thresholds 1 to 3% (conservative management) and 5 to 40% (specialized oncological care). Heterogeneity in clinical utility is estimated by calculating the probability of the model being useful in a hypothetical new centre (p-useful) [23]. Sensitivity, specificity, positive predictive value and negative predictive value for the same decision thresholds are also presented. We report diagnostic performance for discriminating between the five subtypes of adnexal masses as polytomous discrimination index, PDI (a multiclass AUC variant), pairwise AUCs and flexible calibration plots, all while ignoring the multicentre nature of the data due to low numbers of centre-specific events for some outcome categories [24–26]. Methodological details about the performance measures are available in **Supplementary material S2.3.**

We analysed performance for pre-specified subgroups: postmenopausal and premenopausal patients, patients recruited in oncology centres and patients recruited in other centres, and patients that were operated within 120 days after the inclusion scan without any follow-up scan. For the subgroups, we only show results for the two-step strategy.

The two key validations are the two-step strategy with ADNEX2 in all patients and ADNEX2 alone in patients for whom the BDs did not apply. Additional validations include the use of ADNEX2 ignoring CA125, using the original ADNEX model instead of ADNEX2 and the validation of both ADNEX2 and the original ADNEX in all patients (irrespective of whether BDs apply).

### 2.6 Future independent external validation

The recalibrated and refitted coefficients of ADNEX2 are not shown. ADNEX2 is a medical device that should follow medical device regulations [27]. Gynaia (www.gynaia.com) has implemented the model in an app that has obtained regulatory approval for the EU, UK, and US markets.

Researchers who aim to externally validate ADNEX2 can do it freely by using the *evaluatr* R package ( https://github.com/JoieEnsor/evaluatr) [28]. This package provides a safe environment for model developers and model evaluators where the developer does not have access to the patient data, and the evaluator does not have access to the model coefficients but can obtain a vector of predicted risks to evaluate the model’s aggregate performance. To use *evaluatr* a minimum sample size of 50 individuals is needed. Access to ADNEX2 is given by the developer, after a comprehensive description of the purpose (detailed instructions to externally validate ADNEX2 are available at https://adnex2-565575.pages.gitlab.kuleuven.be/01-independent-external-validation.html) .

### 2.7 Patient and public involvement

Patients were not involved in the study design, definition of outcome measures, recruiting plans of the study, or interpretation of study results. We discussed the study with KanActief (https://www.uzleuven.be/nl/kanactief), a cancer rehabilitation patient group at the University Hospitals Leuven.

## 3 Results

After exclusions and data cleaning, 6847 patients recruited at 17 centres in 8 countries were included in the analysis (**Figure 1, Table S1, Supplementary material S1**). Median patient age was 50 years (interquartile range 37-63) and 3302 patients (48%) were postmenopausal. The tumour was benign in 4625 (68%) patients, malignant in 1710 patients (25%), and the outcome was uncertain in 512 (7%) patients. The malignant tumours can be broken down into 391 (6%) borderline, 303 (4%) stage I primary invasive, 787 (11%) stage II-IV primary invasive, and 229 (3%) secondary metastatic tumours. (**Table 1**). The BDs applied to 2190/6847 (32%) of patients (**Table 2**). The remaining 4657 patients in whom the BDs did not apply formed the sample used to update ADNEX. Among these 4657 patients median age was 54 years (interquartile range 41–66), 2646 patients (57%) were postmenopausal, 1728 (37%) had no serum CA125 value, 2638 (57%) had a benign tumour, 1700 (36%) had a malignant tumour, and 319 (7%) had a tumour for which the outcome was uncertain (**Table 2**). Further information on tumour outcome, missing CA125 values, and management is provided in **Tables S4-S9. Table S10** provides descriptive statistics of the samples used to develop the original ADNEX model and to update ADNEX. The proportion of unilocular cysts was much smaller in the updating cohort (8% versus 30%), while the proportion of malignant tumours, multilocular cysts, and tumours with shadowing was higher.

**Figure 1.**
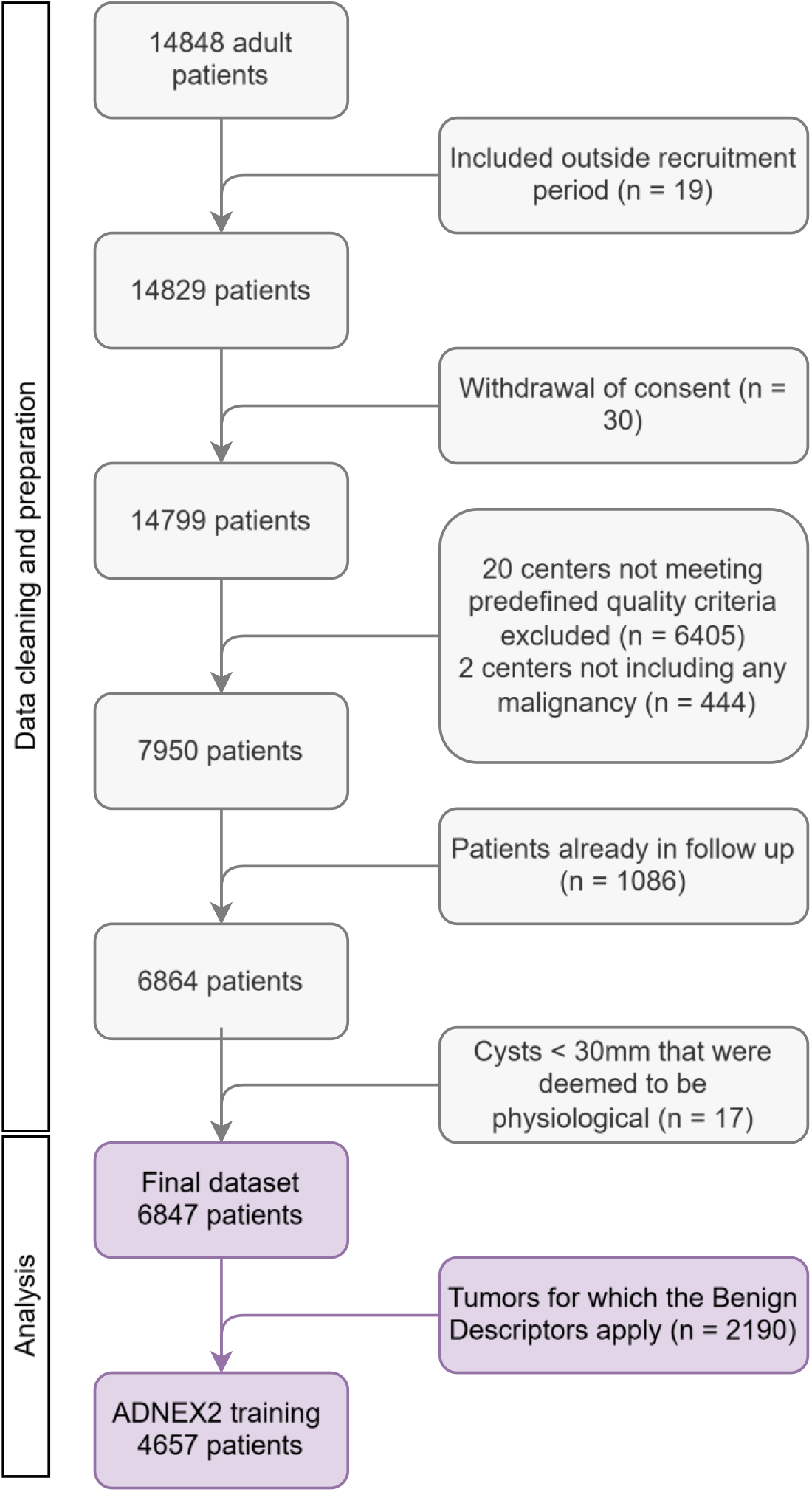
Patient flow diagram of IOTA5 5-year follow-up analysis

**Table 2.** Descriptive statistics of the study cohorts.

| Variable | Patients to whom<br>Benign Descriptors<br>apply | Patients to whom<br>Benign Descriptors do<br>not apply | All patients |
| --- | --- | --- | --- |
| Number of cases | 2190 (32%) | 4657 (68%) | 6847 |
| Benign | 1987 (91%) | 2638 (57%) | 4625 (68%) |
| Borderline | 4 (0.2%) | 387 (8%) | 391 (6%) |
| Stage I invasive cancer | 2 (0.1%) | 301 (6%) | 303 (4%) |
| Stage II-IV invasive cancer | 2 (0.1%) | 785 (17%) | 787 (11%) |
| Secondary Metastasis | 2 (0.1%) | 227 (5%) | 229 (3%) |
| Uncertain<sup>a</sup> | 193 (9%) | 319 (7%) | 512 (7%) |
| Management |  |  |  |
| Surgical | 705 (32%) | 3132 (67%) | 3837 (56%) |
| Conservative | 1332 (61%) | 1307 (28%) | 2639 (39%) |
| Unknown<sup>b</sup> | 153 (7%) | 218 (5%) | 371 (5%) |
| Patient age at recruitment (years)<sup>c</sup> | 42 (32–54) [18–98] | 54 (41–66) [18–99] | 50 (37–63) [18–99] |
| Postmenopausal | 656 (30%) | 2646 (57%) | 3302 (48%) |
| Pelvic pain | 394 (18%) | 594 (13%) | 988 (14%) |
| Presence of solid components | 0 (0%) | 2801 (60%) | 2801 (41%) |
| Proportion of solid components<sup>c</sup> | 0 (0–0) [0–0] | 0.25 (0–0.81) [0–1] | 0 (0–0.49) [0–1] |
| Maximum diameter of the lesion (mm)<sup>c</sup> | 45 (32–61) [7–99] | 65 (43–105) [7–751] | 56 (39–87) [7–751] |
| CA125 (U/ml)<sup>c</sup> | 15 (9–30) [1–7745] | 35 (14–184) [1–78200] | 27 (12–127) [1–78200] |
| CA125 missing | 1498 (68%) | 1728 (37%) | 3226 (47%) |
| Shadows<sup>c</sup> | 378 (17%) | 926 (20%) | 1304 (19%) |
| Ascites<sup>c</sup> | 8 (0.4%) | 482 (10%) | 490 (7%) |
| Number of papillary projections<sup>c</sup> |  |  |  |
| None | 2190 (100%) | 3677 (79%) | 5867 (86%) |
| 1 | 0 (0%) | 464 (10%) | 464 (7%) |
| 2 | 0 (0%) | 143 (3%) | 143 (2%) |
| 3 | 0 (0%) | 95 (2%) | 95 (1%) |
| 4 or more | 0 (0%) | 278 (6%) | 278 (4%) |
| More than 10 cyst locules<sup>c</sup> | 0 (0%) | 634 (14%) | 634 (9%) |
| Colour score of intratumoral flow |  |  |  |
| 1: No blood flow | 1574 (72%) | 1263 (27%) | 2837 (41%) |
| 2: minimal blood flow | 449 (21%) | 1314 (28%) | 1763 (26%) |
| 3: moderate blood flow | 161 (7%) | 1376 (30%) | 1537 (22%) |
| 4: very strong flow | 6 (0.3%) | 704 (15%) | 710 (10%) |
| Tumour type using IOTA terminology |  |  |  |
| Unilocular | 2190 (100%) | 388 (8%) | 2578 (38%) |
| Unilocular solid | 0 (0%) | 630 (13%) | 630 (9%) |
| Multilocular | 0 (0%) | 1438 (31%) | 1438 (21%) |
| Multilocular solid | 0 (0%) | 1111 (24%) | 1111 (16%) |
| Solid | 0 (0%) | 1060 (23%) | 1060 (15%) |
| Not possible to classify | 0 (0%) | 30 (1%) | 30 (<1%) |

Results are presented as median (IQR) [range] for continuous variables, or N (%) for categorical variables.
<sup>a</sup> The outcome for uncertain tumours was imputed with multiple imputation. Tumours are uncertain if there was insufficient information to make a reasonable classification of the nature of the mass at inclusion.
<sup>b</sup> Unknown management means that we have no information after the inclusion scan or that surgery was performed after 1 year from first scan without any follow up scan.
<sup>c</sup> ADNEX and ADNEX2 predictors

### 3.1 Benign descriptors

Among the 2190 tumours to which BDs applied, 1987 (91%) were benign, 10 (0.5%) were malignant, and 193 (9%) had an uncertain outcome (**Table S4**). The 10 histologically confirmed malignancies included 4 borderline, 2 stage I invasive, 2 stage II-IV invasive, and 2 secondary metastatic tumours. After multiple imputation of uncertain outcomes, the proportion of malignancies among patients for which the BDs applied was 0.5% (0.2%-0.9%). More specifically, 0.2% (<0.1% - 0.4%) were borderline, 0.1% (<0.1% - 0.3%) stage I primary invasive, 0.1% (<0.1% - 0.3%) stage II-IV primary invasive, and 0.1% (<0.1% - 0.3%) secondary metastatic (**Table S5**).

### 3.2 ADNEX2

Distributions of the risk predictions given by ADNEX and ADNEX2 are provided in **Figures S4-S6** (benign versus malignant) and **Figures S7-S9** (multiclass). ADNEX2 gave higher risks for malignancy than ADNEX, in particular for borderline and secondary metastatic tumours (**Figures S10-S11)**.

#### 3.2.1 Discrimination

For the two-step strategy using ADNEX2, the AUC to distinguish between benign and malignant tumours was 0.95 (95% CI 0.94-0.96, 95% PI 0.91-0.97). When validated on the patients for which the BDs did not apply, the AUC of ADNEX2 was 0.93 (0.91-0.94) (**Table 3**). The AUC values for ADNEX and ADNEX2, whether used alone or in a two-step strategy, were similar **(Table S11, Figure S12**). Centre-specific AUC values are shown as forest plots in **Figures S13-S24**.

**Table 3.**
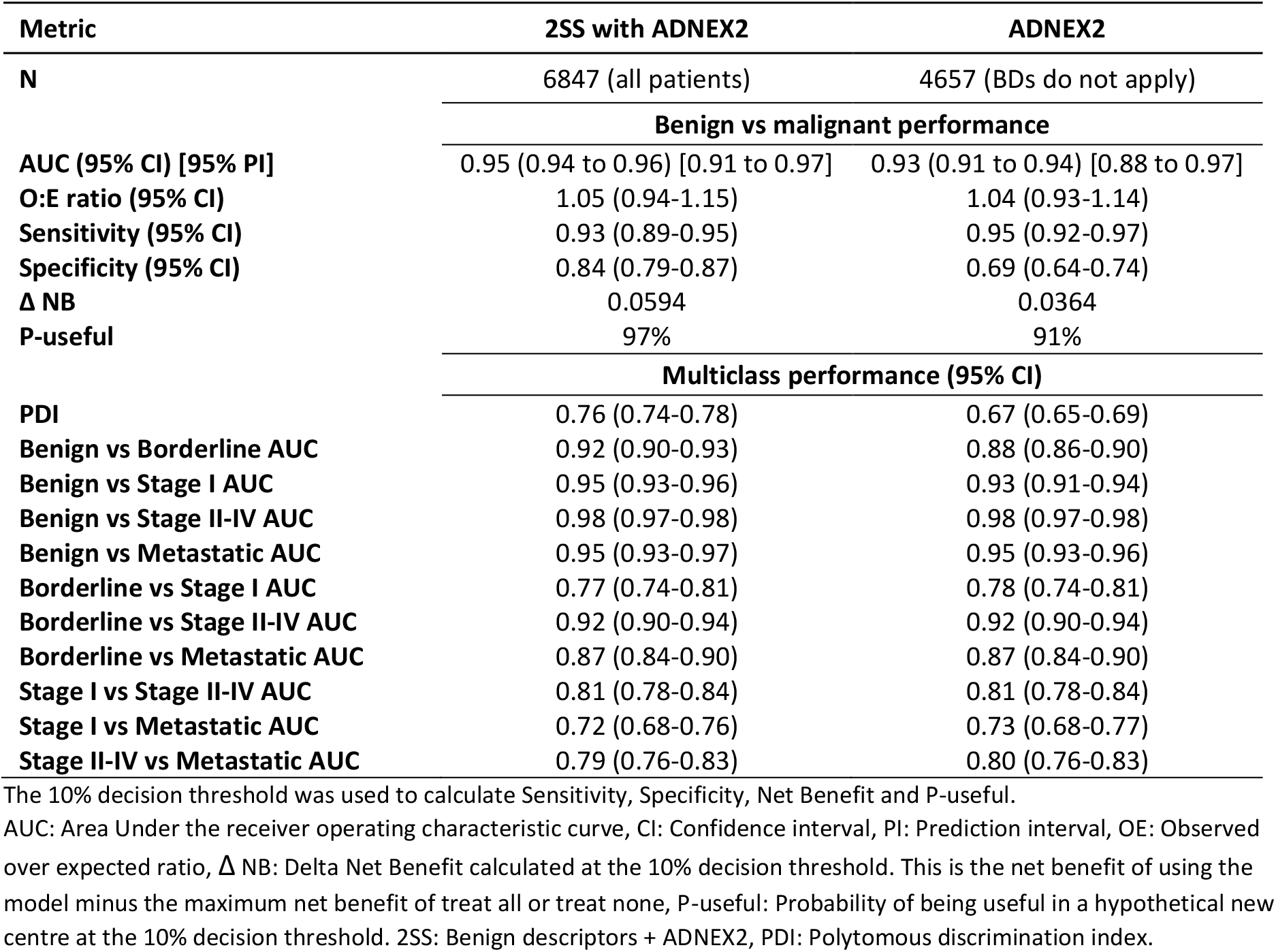
Performance of ADNEX2 in the leave-one-center-out cross-validation.

| Metric | 2SS with ADNEX2 | ADNEX2 |
| --- | --- | --- |
| N | 6847 (all patients) | 4657 (BDs do not apply) |
| **Benign vs malignant performance** |  |  |
| AUC (95% CI) [95% PI] | 0.95 (0.94 to 0.96) [0.91 to 0.97] | 0.93 (0.91 to 0.94) [0.88 to 0.97] |
| O:E ratio (95% CI) | 1.05 (0.94-1.15) | 1.04 (0.93-1.14) |
| Sensitivity (95% CI) | 0.93 (0.89-0.95) | 0.95 (0.92-0.97) |
| Specificity (95% CI) | 0.84 (0.79-0.87) | 0.69 (0.64-0.74) |
| $\Delta$ NB | 0.0594 | 0.0364 |
| P-useful | 97% | 91% |
| **Multiclass performance (95% CI)** |  |  |
| PDI | 0.76 (0.74-0.78) | 0.67 (0.65-0.69) |
| Benign vs Borderline AUC | 0.92 (0.90-0.93) | 0.88 (0.86-0.90) |
| Benign vs Stage I AUC | 0.95 (0.93-0.96) | 0.93 (0.91-0.94) |
| Benign vs Stage II-IV AUC | 0.98 (0.97-0.98) | 0.98 (0.97-0.98) |
| Benign vs Metastatic AUC | 0.95 (0.93-0.97) | 0.95 (0.93-0.96) |
| Borderline vs Stage I AUC | 0.77 (0.74-0.81) | 0.78 (0.74-0.81) |
| Borderline vs Stage II-IV AUC | 0.92 (0.90-0.94) | 0.92 (0.90-0.94) |
| Borderline vs Metastatic AUC | 0.87 (0.84-0.90) | 0.87 (0.84-0.90) |
| Stage I vs Stage II-IV AUC | 0.81 (0.78-0.84) | 0.81 (0.78-0.84) |
| Stage I vs Metastatic AUC | 0.72 (0.68-0.76) | 0.73 (0.68-0.77) |
| Stage II-IV vs Metastatic AUC | 0.79 (0.76-0.83) | 0.80 (0.76-0.83) |

The 10% decision threshold was used to calculate Sensitivity, Specificity, Net Benefit and P-useful.
AUC: Area Under the receiver operating characteristic curve, CI: Confidence interval, PI: Prediction interval, OE: Observed over expected ratio, $\Delta$ NB: Delta Net Benefit calculated at the 10% decision threshold. This is the net benefit of using the model minus the maximum net benefit of treat all or treat none, P-useful: Probability of being useful in a hypothetical new centre at the 10% decision threshold. 2SS: Benign descriptors + ADNEX2, PDI: Polytomous discrimination index.

Not using available CA125 values in ADNEX2 reduced the AUC to distinguish between benign and malignant tumours minimally (**Table S11 and Figure S12**). However, not using available CA125 values clearly reduced the AUC values to distinguish between stage II-IV primary invasive tumours and either stage I primary invasive or secondary metastatic tumours (**Table S12**).

#### 3.2.2 Calibration

ADNEX2 underestimated the risk of malignancy, but less than ADNEX, whether used alone or in the two-step strategy (**Figure 2**, **Figures S25-S36**). O:E ratios were between 1.03 and 1.06 for ADNEX2 and between 1.11 and 1.16 for ADNEX (**Table 3, Table S13**), implying that ADNEX2 underestimated risks less than ADNEX (by 3-6% versus 11-16%). Calibration plots for each of the five tumour types indicated that ADNEX2 was better calibrated than ADNEX for all tumour types and substantially better to predict risk of borderline or secondary metastatic tumours (**Figures S37-S44)**.

**Figure 2.**
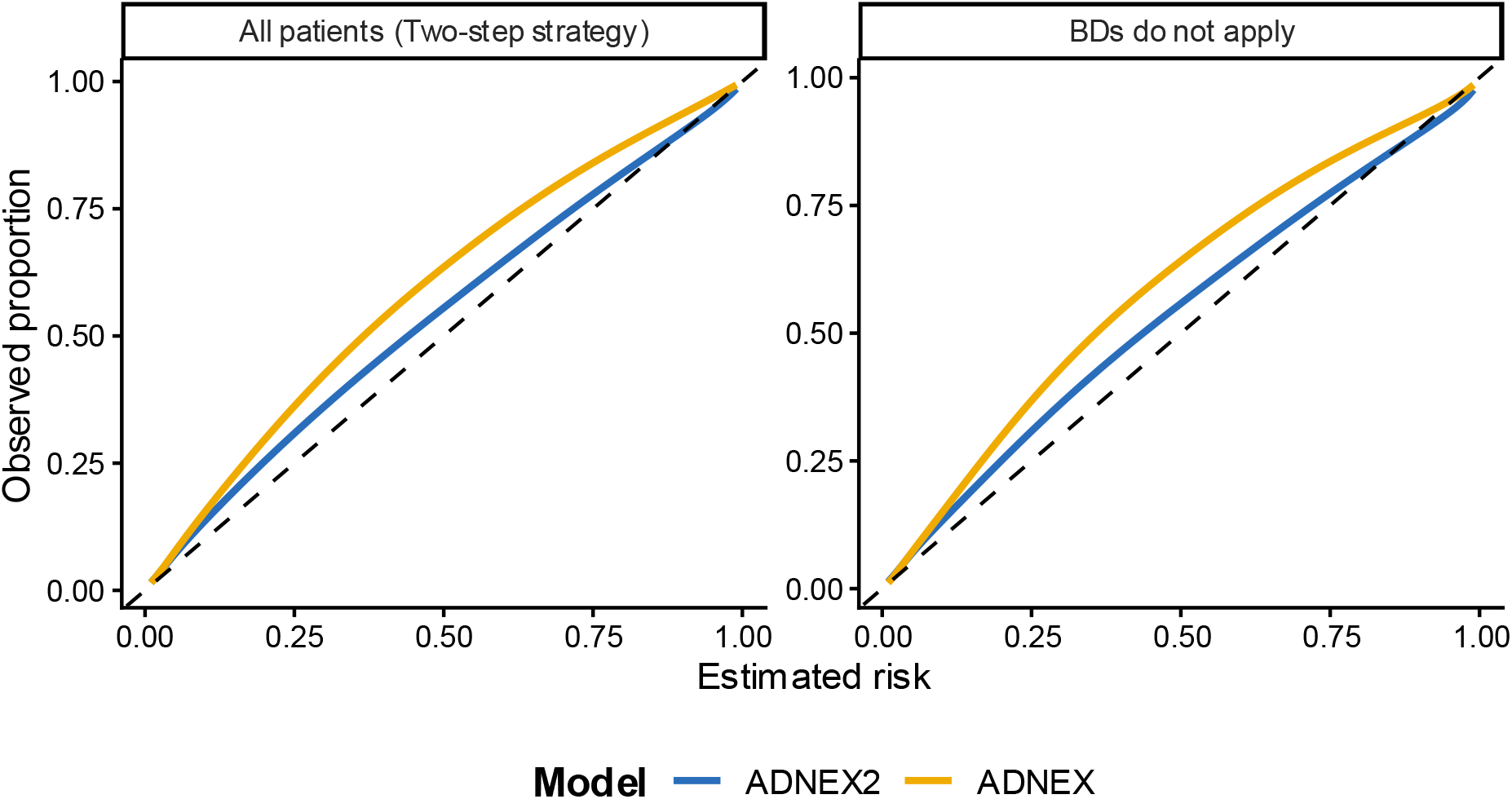
Calibration plots for ADNEX2 and ADNEX (CA125 used if available) in all patients in the two step strategy (n = 6847) and if benign descriptors (BD) do not apply (n = 4657). Black dashed line indicates perfect calibration. Patients to which BDs apply are assigned a risk of malignancy equal to the observed proportion of malignancies in the corresponding training dataset of the leave-one-center-out cross-validation.

#### 3.2.3 Classification and clinical utility

When using the two-step strategy with ADNEX2, the 10% risk of malignancy threshold yielded sensitivity 93% and specificity 84% (**Table 3**). Results were nearly identical when ignoring available CA125 values (**Table S14**). The classification performance of the two-step strategy with ADNEX was similar to that of the two-step strategy with ADNEX2 but sensitivity was slightly lower and specificity slightly higher for ADNEX at most risk thresholds.

The two-step strategy with ADNEX2 showed clinical utility at all decision thresholds and had minimally higher net benefit than the two-step strategy with ADNEX for thresholds between 1 and 30% (**Figure 3, Table S15, Figures S45-46**). Not using available CA125 values marginally reduced net benefit (**Table S15**). Based on meta-analysis, at the 10% decision threshold, the two-step strategy with ADNEX2 was shown to be useful in 97% of centres and the two-step strategy with ADNEX in 98% of centres. When available CA125 values were not used, the two-step strategy was useful in 98% (with ADNEX2) and 97% (with ADNEX) of centres (**Table S15).**

**Figure 3.**
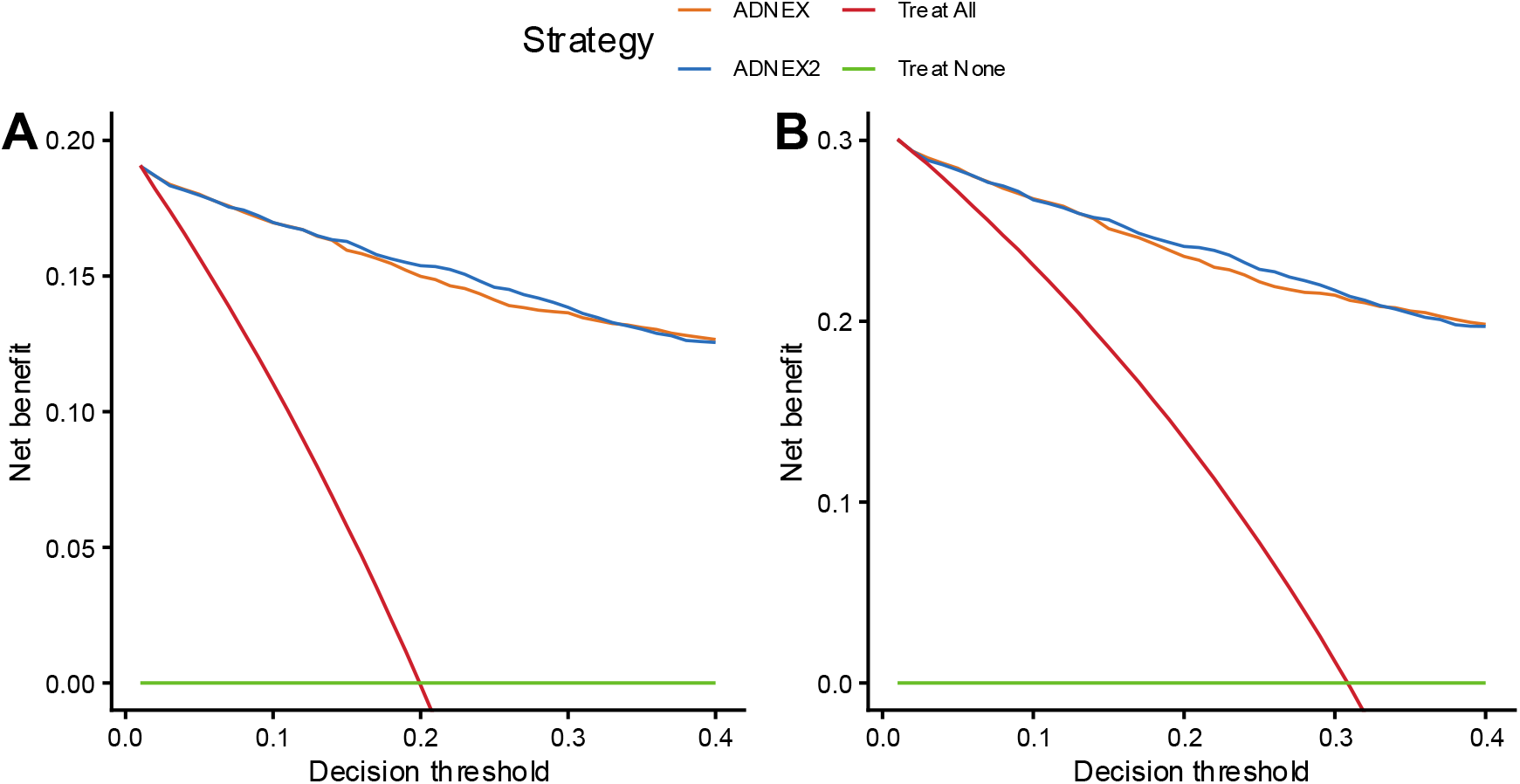
Decision curves for ADNEX2 and ADNEX (CA125 used if available) in all patients in the two-step strategy (n = 6847) (A) and where BDs did not apply (n = 4657) (B).

#### 3.2.4 Subgroup analysis (only results of two-step strategy are shown)

Characteristics of the patients in each subgroup are available in **Table S16.** The AUCs of the two-step strategy with ADNEX2 ranged from 0.92 (non-oncology centres) to 0.95 (postmenopausal patients, and patients in oncology centres) across the subgroups and were almost identical for the two-step strategy with ADNEX **(Figure S47-S57).** AUCs were higher in postmenopausal versus premenopausal patients and in oncology versus non-oncology centres. While the two-step strategy with ADNEX2 had lower AUC in patients operated within 120 days without a follow-up scan (0.93) compared to in all patients (0.95), the pairwise AUCs to distinguish between different malignant subtypes were almost identical in surgically managed patients versus all patients (**Table S17**). Predicted risks for the two-step strategy with ADNEX2 were underestimated in all subgroups. However, ADNEX2 showed better calibration than ADNEX **(Figures S58-S77)**.

At all risk thresholds sensitivity was higher and specificity lower in postmenopausal than premenopausal women and in oncology centres than non-oncology centres (**Table S18**). The two-step strategy with ADNEX2 had comparable or better utility than the two-step strategy with ADNEX in all subgroups at thresholds between 10% and 30%. Ignoring available CA125 values reduced utility mostly in postmenopausal patients (**Table S19, Figure S78**).

## 4 Discussion

### 4.1 Principal findings

We have updated ADNEX to ADNEX2 to optimize it for use in a two-step strategy, where it is used if BDs do not apply. On leave-one-center-out cross-validation, ADNEX2 had good discriminative performance both when used in all tumours and in tumours to which BDs did not apply, and a two-step strategy using ADNEX2 showed clinical utility over the whole range of decision thresholds from 1% to 40%. Although ADNEX has good performance, ADNEX2 was better calibrated and had minimally higher clinical utility than ADNEX whether used alone and in a two-step strategy.

### 4.2 Strengths and limitations

ADNEX2 is the first model for assessing ovarian masses preoperatively that has been developed using information from both surgically and conservatively managed patients. This approach reduces the selection bias that arises when using only histopathology as a reference standard (i.e. including only surgically managed patients). A second strength is large sample size and multicentre design. A third strength is that the final ADNEX2 model was based on the full dataset, i.e. using all available information, while the validation was based on leave-one-center-out cross-validation allowing evaluation of generalizability and heterogeneity of model performance [29].

A first limitation is that the inclusion of surgically managed and conservatively managed patients induces differential verification bias, since histopathology is not available for all tumours [30]. We determined the outcome of tumours without histopathology based on results of ultrasound follow-up examinations and on multiple imputation [11]. We argue that differential verification bias is less problematic than selection bias caused by excluding conservatively managed patients. A second limitation is missing CA125 values in some patients. To obtain the ADNEX2 formula that includes CA125, we handled missing CA125 values with multiple imputation. However, because it is not clinical routine to measure CA125 in all patients, when validating ADNEX2 (and ADNEX) we used ‘CA125 if available’, a realistic strategy. A third limitation is difference in validation of ADNEX2 and ADNEX. For ADNEX2, every centre-specific result can be seen as a geographically validated result due to the use of leave-one-center-out cross-validation. For ADNEX, 11 of the 17 centres in the current study also contributed patients to the original ADNEX development dataset [3]. For these 11 centres the performance of ADNEX can be seen as temporal validation. As a result, the reported ADNEX performance might be somewhat optimistic. A final limitation is that the IOTA5 study does not include information (e.g. ethnicity or socioeconomical status) to assess fairness of model performance.

### 4.3 Comparison with other studies

The discriminative performance of ADNEX2 and ADNEX that we report here is slightly superior to that reported for ADNEX without CA125 in a meta-analysis including 5465 patients examined in 29 centres in 10 countries (AUC 0.93, 95% CI 0.90-0.95) and for ADNEX with CA125 including 9989 patients examined in 44 centres in 17 countries (AUC 0.94 95% CI 0.92-0.95) [4]. However, only two of the studies in the meta-analysis included both patients managed surgically and with clinical and ultrasound follow-up [4]. Calibration and clinical utility were very rarely reported in those studies and therefore could not be meta-analysed. The discriminative performance of the two-step strategy using ADNEX as the second step has been validated in seven studies reporting AUCs of 0.91-0.95 [31–37]. The discriminative performance of the two-step strategy with ADNEX or ADNEX2 that we report here falls within this range (AUC 0.94-0.95).

### 4.4 Results in context

According to an international consensus statement, the malignancy risk calculated by ADNEX can be used to place a patient with an adnexal mass in one of four risk groups of malignancy: risk <1%, risk 1-<10%, risk 10-< 50% and risk ≥50%. The authors of the consensus statement suggest that if the risk of malignancy is <1%, the patient may be suitable for clinical and ultrasound follow-up, if the risk is 1 - <10% the patient may need surgery, and surgery can be performed in a local hospital, while if the risk of malignancy is ≥10% the patient should be referred to a centre for gynaecological oncology [2]. We suggest that the two-step strategy with ADNEX2 can be used in the same way (**Figure 4**). If a BD applies, the patient falls into the lowest risk group (malignancy risk <1%). If a BD does not apply, ADNEX2 is applied, and the calculated malignancy risk is used to place the patient in one of the four risk groups. For estimating the overall malignancy risk, CA125 is not crucial, but if a recent measurement is available at the ultrasound examination, we recommend using it, since it slightly improves discrimination between benign and malignant tumours. When referred to an oncological referral centre, the type of centre should be updated and the risks of the four subtypes of malignancy calculated by ADNEX2 can be used to guide management. In this situation, we recommend measuring CA125 if it was not yet available, because CA125 helps to distinguish Stage II-IV primary invasive cancers from Stage I primary invasive cancers and from secondary metastases. In clinical practice, ADNEX2 needs to be used through an approved medical device with regulatory approval [27].

**Figure 4.**
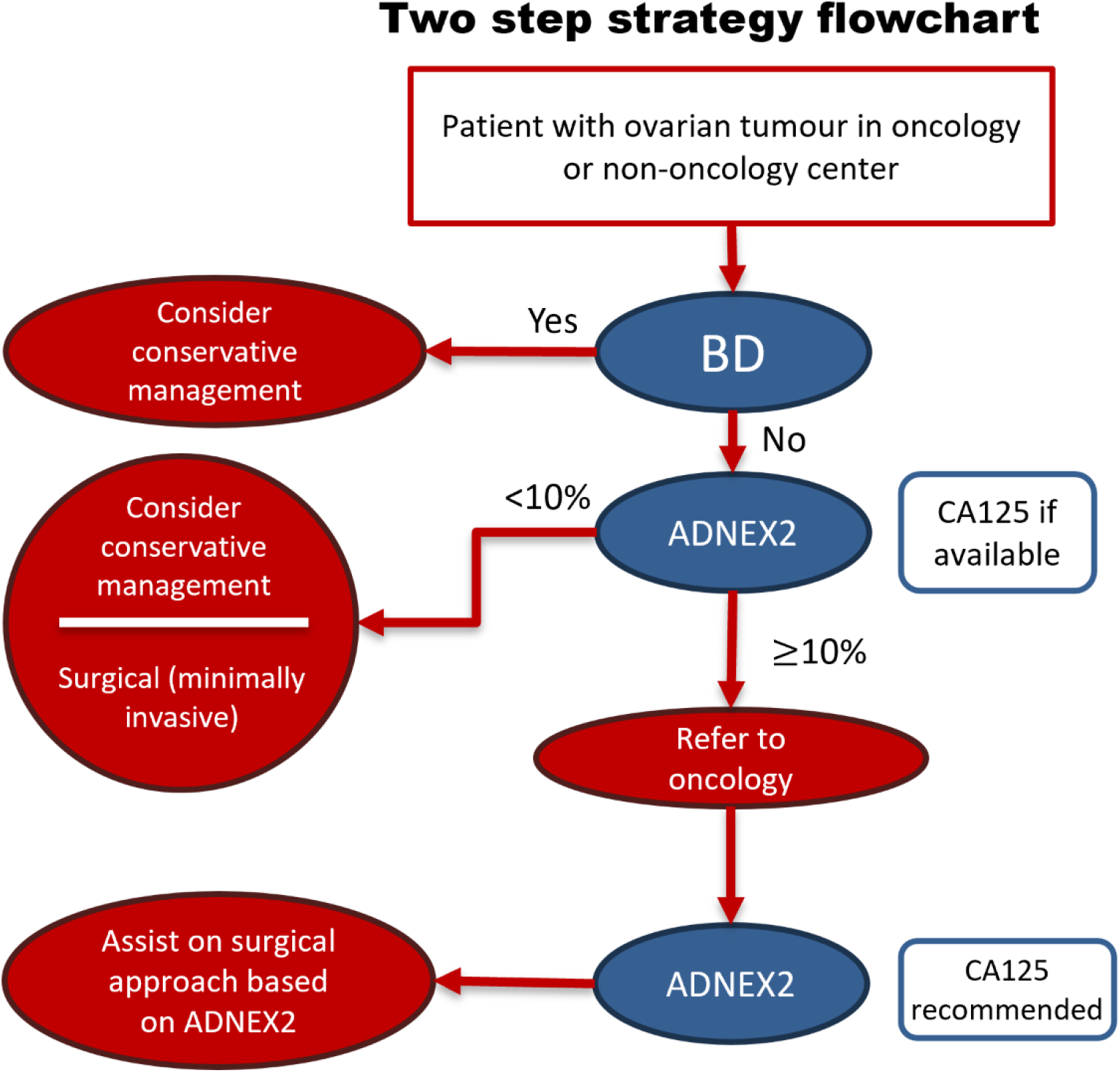
Patient flowchart in two-step strategy with ADNEX2

### 4.5 Future research

ADNEX2, preferably as part of the two-step strategy, should undergo prospective external validation in multicentre studies. If available, these validations should include both surgically and conservatively managed patients. We recommend including at least assessment of discrimination (AUC), calibration (calibration plot) and clinical utility (NB) and to consider the multicentric nature of the data by pooling centre specific results with meta-analysis and providing prediction intervals for the heterogeneity of performance.

### 4.6 Conclusion

We have updated ADNEX to ADNEX2 for use in a two-step strategy where ADNEX2 is applied only when BDs do not apply. Whilst the two-step strategy with ADNEX has very good performance, we recommend to use the two-step strategy with ADNEX2 because it is (1) better calibrated, (2) has minimally higher clinical utility, (3) it is based on more recent data and also on data from patients who were managed conservatively, and (4) it is developed to naturally fit into the two-step strategy.

## 5 Open science

### Funding

Supported by grants from the Research Foundation–Flanders (FWO) (projects G097322N/G049312N/G0B4716N/12F3114N), the Flemish anti-cancer organization Kom Op Tegen Kanker (grant 13583), and Internal Funds KU Leuven (project C24/15/037). Lil Valentin is supported by the Swedish Research Council (grant K2014-99X-22475-01-3, Dnr 2013-02282), funds administered by Malmö University Hospital and Skåne University Hospital, Allmänna Sjukhusets i Malmö Stiftelse för bekämpande av cancer (the Malmö General Hospital Foundation for fighting against cancer), and two Swedish governmental grants (Avtal om läkarutbildning och forskning (ALF)-medel and Landstingsfinansierad Regional Forskning). Tom Bourne is supported by the National Institute for Health Research (NIHR) Biomedical Research Centre based at Imperial College Healthcare NHS Trust and Imperial College London. Laure Wynants is supported by the NWO ZonMW VIDI Talent Programme (Project 09150172310023). Lasai Barreñada is supported by FWO grant 12A8F27N. The views expressed are those of the authors and not necessarily those of the NHS, NIHR or Department of Health. The sponsors had no role in study design; in the collection, analysis, and interpretation of data; in the writing of the report; and in the decision to submit the work for publication. The researchers performed this work independently of the funding sources.

### Protocol

The IOTA5 protocol was registered in clinicaltrials.gov (https://clinicaltrials.gov/study/NCT01698632, **Supplementary File S7**) and the statistical analysis plan was preregistered in an Open Science Framework (OSF) preregistration (https://osf.io/ypn7r).

### Data Sharing

Follow-up data collection for the IOTA5 study is ongoing. When the study has closed and results have been published, the study responsible may share data upon reasonable request to. The R code and a dataset that includes centre code, outcome and patient risk of malignancy for each of the different models and strategies is shared to allow readers to reproduce the results and figures.

### Code sharing

All code to produce the results and visualisations in the document is shared in a Gitlab repository (https://gitlab.kuleuven.be/sdpm/ADNEX2) as well as in an interactive website to visualize the results (https://adnex2-565575.pages.gitlab.kuleuven.be/).

### Transparency

The lead author (LB) and guarantors (DT, BVC) affirm that the manuscript is an honest, accurate, and transparent account of the study being reported; that no important aspects of the study have been omitted; and that any discrepancies from the study as originally planned (and registered) have been explained.

### Ethical approval

The IOTA5 study was approved by the Research Ethics Committee of the University Hospitals Leuven as the coordinating centre (B32220095331/S51375) and by the local ethics committees of all participating centres. All participants provided informed consent before taking part.

### Dissemination to participants and related patient and public communities

The findings of this study will be shared with patient groups (including KanActief at University Hospitals Leuven) and communicated to the public and clinical community through the IOTA website (https://iotaplus.org/), social media, and institutional press channels.

## Data Availability

Follow-up data collection for the IOTA5 study is ongoing. When the study has closed and results have been published, the study responsible may share data upon reasonable request to. All code to produce the results and visualisations in the document is shared in a Gitlab repository (https://gitlab.kuleuven.be/sdpm/ADNEX2) as well as in an interactive website to visualize the results (https://adnex2-565575.pages.gitlab.kuleuven.be/).

