## Supplementary material for "Update of the ADNEX risk model (ADNEX2) to diagnose ovarian cancer in surgically and conservatively managed patients with an adnexal mass: multicentre prospective cohort study"

**Dr Lasai Barreñada[1,2], Prof Wouter Froyman[1,3], Ms Jolien Ceusters[1,4], Dr Chiara Landolfo[5], Dr Laure Wynants[1,2,6], Ms Ashleigh Ledger[1], Dr Povilas Sladkevicius[7,8], Prof Antonia C Testa[9,10,11], Dr Caroline Van Holsbeke[12], Dr Francesca Buonomo[13], Prof Marek J Kudla[14], Prof Robert Fruscio[15,16], Prof Daniela Fischerova[17], Dr Dorella Franchi[18], Prof Elisabeth Epstein[19], Dr Valentina Chiappa[20], Dr Francesco PG Leone[21], Dr Joie Ensor[22], Prof Maria Elisabetta Coccia[23], Prof Stefano Guerriero[24,25], Prof Juan Luis Alcazar[26], Dr Fatima Alves[27], Dr Ligita Jokubkiene[7,8], Dr Federica Pozzati[9,10], Dr Francesca Moro[9,10,28], Dr Floriana Mascilini[9,10], Dr Cristina Taliento[1,29,30], Prof Tom Bourne[1,5], Prof Lil Valentin[7,8], Prof Dirk Timmerman[1,2], Prof Ben Van Calster[1,2,31]**

[1]Department of Development and Regeneration, KU Leuven, Leuven, Belgium. [2]Leuven Unit for Health Technology Assessment Research (LUHTAR), KU Leuven, Leuven, Belgium. [3]Department of Obstetrics and Gynecology, University Hospitals Leuven, Leuven, Belgium. [4]Laboratory of Tumour Immunology and Immunotherapy, Department of Oncology, Leuven Cancer Institute, KU Leuven, Leuven, Belgium. [5]Queen Charlotte's and Chelsea Hospital, Imperial College London, London, United Kingdom. [6]Department of Epidemiology, CAPHRI Care and Public Health Research Institute, Maastricht University, Maastricht, The Netherlands. [7]Department of Obstetrics and Gynecology, Skåne University Hospital, Malmö, Sweden. [8]Department of Clinical Sciences Malmö, Lund University, Malmö, Sweden. [9]Dipartimento Scienze della Salute della Donna e del Bambino, Fondazione Policlinico Universitario Agostino Gemelli, IRCCS, Rome, Italy. [10]G-AURIG – Gemelli Advanced Ultrasound Research and Innovation Group, IRCCS, Fondazione Policlinico Universitario Agostino Gemelli, Rome, Italy. [11]Gemelli Women's Health Center for Digital and Personalized Medicine, Dipartimento Scienze della Vita e Sanità Pubblica, Università Cattolica del Sacro Cuore, Rome, Italy. [12]Department of Obstetrics and Gynecology, Ziekenhuis Oost-Limburg, Genk, Belgium. [13]Institute for Maternal and Child Health-IRCCS "Burlo Garofolo", Trieste, Italy. [14]Department of Perinatology and Oncological Gynecology, Faculty of Medical Sciences, Medical University of Silesia, Katowice, Poland. [15]UO Gynecology, Fondazione IRCCS San Gerardo dei Tintori, Monza, Italy. [16]Department of Medicine and Surgery, University of Milan-Bicocca, Milan, Italy. [17]Gynecologic Oncology Centre, Department of Gynecology, Obstetrics and Neonatology, First Faculty of Medicine, Charles University and General University Hospital in Prague, Prague, Czech Republic. [18]Preventive Gynecology Unit, Division of Gynecology, European Institute of Oncology IRCCS, Milan, Italy. [19]Department of Clinical Science and Education, Karolinska Institutet, Södersjukhuset and Department of Obstetrics and Gynecology, Södersjukhuset, Stockholm, Sweden. [20]Gynecologic Oncology, Fondazione IRCCS Istituto Nazionale dei Tumori, Milan, Italy. [21]Department of Obstetrics and Gynecology, Biomedical and Clinical Sciences Institute L. Sacco, University of Milan, Milan, Italy. [22]Department of Applied Health Sciences, University of Birmingham, Birmingham, UK. [23]Department of Biomedical, Experimental and Clinical Sciences, University of Florence, Florence, Italy. [24]Centro Integrato di Procreazione Medicalmente Assistita (PMA) e Diagnostica Ostetrico-Ginecologica,Azienda Ospedaliero Universitaria. [25]University of Cagliari, Department of Surgical Sciences, Cagliari, Italy. [26]Department of Obstetrics and Gynecology, Clínica Universidad de Navarra, Pamplona, Spain. [27]Department of Gynecology, Obstetrics and Reproductive Medicine, University Hospital de Santa Maria, Lisbon, Portugal. [28]UniCamillus, International Medical University, Rome, Italy. [29]Department of Obstetrics and Gynecology, University Hospital Ferrara, Ferrara, Italy. [30]Department of Obstetrics and Gynaecology, Heidelberg University Hospital, Heidelberg, Germany. [31]Julius Center for Health Sciences and Primary Care, University Medical Center Utrecht, Utrecht, the Netherlands.

Corresponding author:

Ben Van Calster

KU Leuven, Department of Development and Regeneration

Herestraat 49 box 805 | 3000 Leuven | Belgium

T +32 16377788 | E

Figure S1. Convergence plots for CA125 and the multinomial reference standard. The plots on the top row refer to CA125 (after log-log transformation; ll_CA125), the plots on the bottom row refer to the multinomial reference standard (CD_5groups). The plots on the left show the mean value, the plots on the right show the standard deviation (sd). The x-axis refers to the iteration (1 to 50). The coloured lines refer to the 100 imputations. The multinomial reference standard was used as a nominal variable in the imputations, despite it being represented as a numerical variable in these plots. ........................................................................................................ 18

Table S9. Number of patients with tumours to which the benign descriptors did not apply with missing CA125 values by management in the cohort used to update ADNEX to ADNEX2. 32

Table S10. Characteristics of patients and tumours in the ADNEX development cohort [19] (N = 5909, 24 centres) and ADNEX to ADNEX2 update cohort. 33

S4.2 RISK DISTRIBUTIONS 34

Figure S4. Probabilities by tumour outcome in all patients (N = 6847) for two step strategy (2SS) with ADNEX2 and ADNEX. Predicted risks are averaged across imputations. 34

Figure S5. Probabilities by tumour outcome in all patients (N = 6847) for ADNEX2 and ADNEX. Predicted risks are averaged across imputations. 35

Figure S6. Probabilities calculated by ADNEX2 and original ADNEX by tumour outcome in patients where benign descriptors did not apply (N = 4657). Predicted risks are averaged across imputations. 36

Figure S7. Boxplot of predicted probability distribution by tumour subtype in all patients (N = 6847) for two step strategy (2SS) with ADNEX2 and ADNEX. Red line indicates baseline proportion per tumour subtype. Predicted risks are averaged across imputations. 37

Figure S8. Boxplot of predicted probability distribution by tumour subtype in all patients (N = 6847) for ADNEX2 and ADNEX. Red line indicates baseline proportion per tumour subtype. Predicted risks are averaged across imputations. 38

Figure S9. Boxplot of predicted probability distribution by tumour subtype in patients where BDs do not apply (N = 4657) for ADNEX2 and ADNEX. Red line indicates baseline proportion per tumour subtype. Predicted risks are averaged across imputations. 39

S4.3 ADNEX2 VS ADNEX 40

Figure S10. Scatterplot of predicted risks of ADNEX and ADNEX2 in patients where BDs do not apply (N = 4657). Each point represents a patient. Patients' specific predictions are averaged across imputations. 40

Figure S11. Scatterplot of predicted risks of ADNEX and ADNEX2 ignoring CA125 in patients where BDs do not apply (N = 4657). Each point represents a patient. Patients' specific predictions are averaged across imputations. 41

Table S11. Performance of ADNEX, ADNEX2 and two-step strategies in different populations. 42

S4.4 DISCRIMINATION 43

Table S12. Polytomous discrimination index and pairwise area under receiver operating characteristic curve of ADNEX2 and ADNEX to distinguish between outcome categories. 43

Figure S12. Forest plot showing the meta-analysed area under the receiver operating characteristic curve (AUC) for ADNEX, ADNEX2 and the two-step strategies in the different populations. Blue points and lines correspond to ADNEX2 and orange ones to ADNEX. Horizontal lines show prediction interval. CI, 95% confidence interval. PI, 95% prediction interval. 2SS, two step strategy. 44

Figure S13. Forest plot showing the area under the receiver operating characteristic curve (AUC) for two step strategy with ADNEX2 in all patients (n = 6847). CI, 95% confidence interval; PI, prediction interval 45

Figure S14. Forest plot showing the area under the receiver operating characteristic curve (AUC) for two step strategy with ADNEX2 ignoring CA125 in all patients (n = 6847). CI, 95% confidence interval; PI, prediction interval 46

Figure S15. Forest plot showing the area under the receiver operating characteristic curve (AUC) for two step strategy with ADNEX in all patients (n = 6847). CI, 95% confidence interval; PI, prediction interval 47

Figure S16. Forest plot showing the area under the receiver operating characteristic curve (AUC) for two step strategy with ADNEX ignoring CA125 in all patients (n = 6847). CI, 95% confidence interval; PI, prediction interval 48

Figure S17. Forest plot showing the area under the receiver operating characteristic curve (AUC) for ADNEX2 in all patients (n = 6847). CI, 95% confidence interval; PI, prediction interval ... 49
Figure S18. Forest plot showing the area under the receiver operating characteristic curve (AUC) for ADNEX2 ignoring CA125 in all patients (n = 6847). CI, 95% confidence interval; PI, prediction interval ... 50
Figure S19. Forest plot showing the area under the receiver operating characteristic curve (AUC) for ADNEX in all patients (n = 6847). CI, 95% confidence interval; PI, prediction interval ... 51
Figure S20. Forest plot showing the area under the receiver operating characteristic curve (AUC) for ADNEX ignoring CA125 in all patients (n = 6847). CI, 95% confidence interval; PI, prediction interval ... 52
Figure S21. Forest plot showing the area under the receiver operating characteristic curve (AUC) for ADNEX2 where benign descriptors do not apply (n = 4657). CI, 95% confidence interval; PI, prediction interval ... 53
Figure S22. Forest plot showing the area under the receiver operating characteristic curve (AUC) for ADNEX2 ignoring CA125 where benign descriptors do not apply (n = 4657). CI, 95% confidence interval; PI, prediction interval ... 54
Figure S23. Forest plot showing the area under the receiver operating characteristic curve (AUC) for ADNEX where benign descriptors do not apply (n = 4657). CI, 95%confidence interval; PI, prediction interval ... 55
Figure S24. Forest plot showing the area under the receiver operating characteristic curve (AUC) for ADNEX ignoring CA125 where benign descriptors do not apply (n = 4657). CI, 95% confidence interval; PI, prediction interval ... 56
S4.5 CALIBRATION ... 57
Table S13. Calibration metrics for ADNEX and ADNEX2 when used alone or in a two-step strategy ... 57
*S4.5.1 Overall and centre specific multicentre calibration curves with 95% prediction interval ... 58*
Figure S25. Calibration plot for two step strategy with ADNEX2 in all patients (n = 6847). Overall calibration curves calculated taking clustering into account. Shaded area indicates 95% prediction interval. Linewidth is proportional with sample size. The thick black straight line is the ideal line denoting perfect agreement between predicted risk and observed proportion. ... 58
Figure S26. Calibration plot for two step strategy with ADNEX2 ignoring CA125 in all patients (n = 6847). Overall calibration curves calculated taking clustering into account. Shaded area indicates 95% prediction interval. Linewidth is proportional with sample size. The thick black straight line is the ideal line denoting perfect agreement between predicted risk and observed proportion. ... 59
Figure S27. Calibration plot for two step strategy with ADNEX in all patients (n = 6847). Overall calibration curves calculated taking clustering into account. Shaded area indicates 95% prediction interval. Linewidth is proportional with sample size. The thick black straight line is the ideal line denoting perfect agreement between predicted risk and observed proportion. ... 60
Figure S28. Calibration plot for two step strategy with ADNEX ignoring CA125 in all patients (n = 6847). Overall calibration curves calculated taking clustering into account. Shaded area indicates 95% prediction interval. Linewidth is proportional with sample size. The thick black straight line is the ideal line denoting perfect agreement between predicted risk and observed proportion. ... 61
Figure S29. Calibration plot for ADNEX2 in all patients (n = 6847). Overall calibration curves calculated taking clustering into account. Shaded area indicates 95% prediction interval. Linewidth is proportional with sample size. The thick black straight line is the ideal line denoting perfect agreement between predicted risk and observed proportion. ... 62

### S1. EXCLUSION OF CENTRES

The data used in this study are also used to study the 5-year cumulative incidence of tumour-related complications in benign-looking tumours that were managed conservatively with clinical and ultrasound follow-up (manuscript in preparation). Predefined criteria for excluding centres from that study were 1) <50 patients recruited, 2) no information regarding actual initial management (surgical versus conservative) in more than 15% of the recruited patients or, 3) good quality follow-up information available for less than 70% of the patients with a newly detected mass that were managed conservatively. These percentages are based on 'new' patients, i.e. patients in whom the index mass was not already in follow-up at the center before recruitment into IOTA5. The thresholds of 15% and 70% are arbitrary but were considered reasonable by the International Ovarian Tumour Analysis (IOTA) Steering Committee.

'Good follow-up information' was defined as: a recorded study outcome during follow-up (spontaneous resolution of the mass, surgery at any time during follow-up or patient death), or a last follow-up visit at least 54 months after the inclusion scan.

**Table S1** provides information on included and excluded centres. Below, we provide detailed information on the decision to exclude some centres.

- Five centres were excluded because <50 patients were recruited: Catania, Cremona, Paris, Aarschot and Maurepas. The reasons for such small sample size could be that the centre joined the study late or did not recruit consecutively.
- Eleven centres were excluded because there was no information regarding actual initial management in more than 15% of the recruited patients and/or because good quality follow-up information was available for less than 70% of the patients that were followed conservatively: Bologna, Nottingham, Cairo, Lublin, Bangalore, Bari, Barcelona (BCN), Vienna, Milan (FIT), Beijing and Tienen.
- One centre (Udine) was excluded because it focussed only on recruiting surgically managed patients.
- Two centres (Lisbon (DEP) and Tampa) were excluded because they did not focus on recruiting surgically managed patients.
- One centre (Barcelona, BSP) was excluded because it is a different type of centre offering screening for gynaecological disease [1].
- One centre (Krakow) stopped participation and could not complete follow-up information.

- One centre (Athens) was excluded because there was a suspiciously high percentage (85%) of masses that spontaneously resolved among patients that were conservatively managed, and the centre was non-responsive for data cleaning.

**Table S1. Key information for centres that participated in the 5-year follow-up analysis of IOTA5**

| | | | | | | Outcome | | | Actual management | | | |
|---|---|---|---|---|---|---|---|---|---|---|---|---|
| Centre | Code | Principal investigator | N all | N new | Missing CA125 | Benign | Malignant | Uncertain | Surgery | Conservative | Unknown | Decision |
| Malmö, Sweden | MSW | Lil Valentin, MD | 1277 | 1093 | 67% | 901 | 117 | 75 | 470 | 583 | 40 | Included |
| Stockholm, Sweden | SSW | Elisabeth Epstein, MD | 629 | 509 | 18% | 266 | 211 | 32 | 379 | 116 | 14 | Included |
| Monza, Italy | OIT | Robert Fruscio, MD | 707 | 431 | 31% | 267 | 130 | 34 | 246 | 154 | 31 | Included |
| Lisbon, Portugal | DEP | Maria José dos Santos Bernardo, MD | 362 | 362 | 23% | 354 | 0 | 8 | 27 | 335 | 0 | Excluded |
| Trieste, Italy | TIT | Francesca Buonomo, MD | 275 | 273 | 25% | 227 | 42 | 4 | 121 | 151 | 1 | Included |
| Cagliari, Italy | SIT | Stefano Guerriero, MD | 265 | 247 | 15% | 202 | 39 | 6 | 190 | 53 | 4 | Included |
| Katowice, Poland | MPO | Marek Kudla, MD | 241 | 241 | 53% | 183 | 32 | 26 | 84 | 131 | 26 | Included |
| Milan, Italy (Milan 2) | NCI | Valentina Chiappa, MD | 308 | 227 | 13% | 105 | 108 | 14 | 147 | 73 | 7 | Included |
| London, UK | IUK | Tom Bourne, MD | 175 | 172 | 65% | 147 | 7 | 18 | 19 | 143 | 10 | Included |
| Milan, Italy (Milan 3) | MIT | Francesco Paolo Leone, MD | 155 | 129 | 89% | 120 | 3 | 6 | 52 | 72 | 5 | Included |
| Florence, Italy | FLI | Maria Elisabetta Coccia, MD | 95 | 85 | 94% | 73 | 2 | 10 | 31 | 49 | 5 | Included |
| Tampa, USA | TUS | Lauri Hochberg, MD | 82 | 33 | 79% | 26 | 0 | 7 | 3 | 25 | 5 | Excluded |
| Rome, Italy | RIT | Antonia Testa, MD | 1058 | 1019 | 60% | 617 | 293 | 109 | 577 | 351 | 91 | Included |
| Leuven, Belgium | LBE | Dirk Timmerman, MD | 894 | 754 | 56% | 505 | 206 | 43 | 423 | 301 | 30 | Included |
| Milan, Italy | CIT | Dorella Franchi, MD | 618 | 558 | 8% | 284 | 260 | 14 | 452 | 90 | 16 | Included |
| Genk, Belgium | GBE | Caroline Van Holsbeke, MD | 515 | 473 | 78% | 370 | 54 | 49 | 254 | 176 | 43 | Included |
| Prague, Czech Rep. | PCR | Daniela Fischerova, MD | 446 | 414 | 33% | 208 | 164 | 42 | 256 | 130 | 28 | Included |
| Pamplona, Spain | PSP | Juan Luis Alcázar, MD | 223 | 170 | 64% | 112 | 41 | 17 | 111 | 44 | 15 | Included |
| Lisbon, Portugal | LIP | Fatima Alves, MD | 52 | 52 | 15% | 38 | 6 | 8 | 25 | 22 | 5 | Included |
| Barcelona, Spain | BSP | Maria Angela Pascual, MD | 2645 | 2039 | 89% | 1682 | 49 | 308 | 314 | 1497 | 228 | Excluded |
| Udine, Italy | UDI | Alberto Rossi, MD | 153 | 153 | 0% | 112 | 40 | 1 | 151 | 0 | 2 | Excluded |
| Bologna, Italy | BIT | Luca Savelli, MD | 1133 | 992 | 75% | 437 | 106 | 449 | 282 | 381 | 329 | Excluded |
| Athens, Greece | AGR | Ekaterini Domali, MD | 727 | 715 | 55% | 509 | 97 | 109 | 494 | 123 | 98 | Excluded |
| Nottingham, UK | NUK | Nandita Deo, MD | 119 | 109 | 41% | 67 | 7 | 35 | 63 | 17 | 29 | Excluded |
| Cairo, Egypt | CEG | Mona Aboulghar, MD | 75 | 70 | 69% | 45 | 9 | 16 | 36 | 21 | 13 | Excluded |
| Lublin, Poland | LPO | Artur Czekierdowski, MD | 440 | 438 | 26% | 274 | 36 | 128 | 116 | 200 | 122 | Excluded |
| Bangalore, India | BIN | Mala Sibal, MD | 174 | 173 | 46% | 87 | 9 | 77 | 78 | 20 | 75 | Excluded |
| Bari, Italy | BAI | Doriana Scardigno, MD | 143 | 143 | 10% | 92 | 25 | 26 | 109 | 9 | 25 | Excluded |
| Barcelona, Spain | BCN | Noelia Rams, MD | 107 | 105 | 32% | 45 | 8 | 52 | 29 | 61 | 15 | Excluded |
| Krakow, Poland | KPO | Anna Knafel, MD | 175 | 96 | 65% | 48 | 41 | 7 | 86 | 4 | 6 | Excluded |
| Vienna, Austria | VAS | Samir Helmy, MD | 90 | 88 | 86% | 46 | 10 | 32 | 39 | 35 | 14 | Excluded |
| Milan, Italy | FIT | Chiara Lanzani, MD | 67 | 65 | 0% | 37 | 1 | 27 | 15 | 29 | 21 | Excluded |
| Beijing, China | BCH | Jing Zhang, MD | 64 | 64 | 2% | 26 | 19 | 19 | 39 | 4 | 21 | Excluded |
| Tienen, Belgium | RZT | Thierry Van den Bosch, MD | 60 | 46 | 85% | 4 | 0 | 42 | 1 | 10 | 35 | Excluded |
| Catania, Italy | CAI | Maria Concetta Blanco, MD | 39 | 39 | 67% | 19 | 4 | 16 | 19 | 7 | 13 | Excluded |
| Cremona, Italy | CRI | Paola Pollastri, MD | 47 | 40 | 60% | 31 | 2 | 7 | 24 | 16 | 0 | Excluded |
| Paris, France | PFR | Perrine Capmas, MD | 28 | 28 | 57% | 14 | 6 | 8 | 24 | 0 | 4 | Excluded |

| | | | | | | | | | | | | |
|---|---|---|---|---|---|---|---|---|---|---|---|---|
| **Aarschot, Belgium** | **MCA** | **Thierry Van den Bosch, MD** | **45** | **26** | **100%** | **5** | **0** | **21** | **1** | **8** | **17** | **Excluded** |
| **Maurepas, France** | **MFR** | **Ulrike Metzger, MD** | **48** | **17** | **88%** | **11** | **0** | **6** | **3** | **10** | **4** | **Excluded** |
| **Total** | | | **14756** | **12688** | **54%** | **8596** | **2184** | **1908** | **5790** | **5451** | **1447** | |

N all: all recruited patients after excluding patients recruited outside inclusion period, withdrawal of informed consent and cysts deemed to be physiological with a size smaller than 3 cm
N new: new patients after excluding patients recruited outside inclusion period, withdrawal of informed consent and cysts deemed to be physiological with a size smaller than 3 cm
Surgery: patients operated without follow-up scan within 1 year from inclusion scan.
Conservative: patients that had at least 1 follow-up scan
Unknown management means that we have no information after the inclusion scan or that surgery was performed after 1 year from first scan without follow up information.

### S2. Details on statistical analysis

#### S2.1 Sample size

For prediction model development, sample size is not about power but about precision and overfitting control. The sample size procedure for multinomial logistic regression models targets three criteria [2]:

1. **Overfitting control for every pair of outcome categories.** Operationalization: reaching a prespecified minimum shrinkage factor across the 10 pairs (10 criteria).
2. **Overfitting control for the model in general.** Operationalization: reaching a prespecified minimum difference between the observed and adjusted Nagelkerke R-squared (1 criterion)
3. **Precise estimation of the overall risk for each outcome category.** Operationalization: reaching a prespecified maximum width of the 95% confidence intervals for the estimated overall risk per outcome category (5 criteria)

The calculation requires the specification of the target values for each of the 16 criteria above, the number of coefficients per equation in the MLR model, the prevalence of each outcome category, the adjusted R-squared of the multinomial model, the adjusted R-squared values for the comparison between each pair of outcome categories (**Table S2**). The adjusted R-squared of the multinomial model is the conservative estimate from Pate et al, which is 15% of the maximum Cox-Snell R-squared value [2]. The adjusted R-squared values for each pair of outcome categories are based on c-statistics from an unpublished post hoc analysis of the data in Van Calster et al (2020) [3]. The prevalence for each outcome category is taken from **Table 2**, assuming that all uncertain outcomes are benign. The code to get the minimum sample size is available in the **Gitlab repository** (https://gitlab.kuleuven.be/sdpm/ADNEX2) and based on the code provided in Pate et al. [2]

Our available sample size (n=4657) satisfied all 16 criteria for the model with CA125 and 15 out of 16 criteria for the model ignoring CA125 (**Tables S2-S3**). The 16$^{th}$ criterion is satisfied if we target a calibration slope of 0.83 for distinction between stage II-IV primary invasive and secondary metastatic tumours.

**Table S2. Parameters for the sample size calculation.**

| Parameter | Model without CA125 | Model with CA125 |
|---|---|---|
| Minimum shrinkage for every pair of outcome categories | 0.9 | 0.9 |
| Difference between observed and adjusted R-squared | 0.05 | 0.05 |
| Confidence interval width for the overall risks | 0.05 | 0.05 |
| Number of coefficients per equation | 11 | 11 |
| Prevalence benign | 0.64 | 0.64 |
| Prevalence borderline | 0.08 | 0.08 |
| Prevalence stage I invasive | 0.06 | 0.06 |
| Prevalence stage II-IV invasive | 0.17 | 0.17 |
| Prevalence secondary metastasis | 0.05 | 0.05 |
| Adjusted $R^2$ of MLR model (conservative estimate) | 0.134 | 0.134 |
| Adjusted $R^2$ benign vs borderline | 0.170 | 0.182 |
| Adjusted $R^2$ benign vs stage I | 0.222 | 0.236 |
| Adjusted $R^2$ benign vs stage II-IV | 0.468 | 0.525 |
| Adjusted $R^2$ benign vs secondary metastasis | 0.194 | 0.208 |
| Adjusted $R^2$ borderline vs stage I | 0.227 | 0.211 |
| Adjusted $R^2$ borderline vs stage II-IV | 0.408 | 0.449 |
| Adjusted $R^2$ borderline vs secondary metastasis | 0.391 | 0.410 |
| Adjusted $R^2$ stage I vs stage II-IV | 0.115 | 0.226 |
| Adjusted $R^2$ stage I vs sec. meta | 0.183 | 0.199 |
| Adjusted $R^2$ stage II-IV vs secondary metastasis | 0.056 | 0.170 |

MLR: Multinomial logistic regression

**Table S3. Sample size requirements for the different criteria.**

| **Criterion** | **Required number for model without CA125** | **Required number for model with CA125** |
|---|---|---|
| Criterion 1 – Borderline vs benign | 705 | 656 |
| Criterion 1 – Stage I vs benign | 526 | 488 |
| Criterion 1 – Stage II-IV vs benign | 188 | 157 |
| Criterion 1 – Sec meta vs benign | 664 | 616 |
| Criterion 1 – Borderline vs stage I | 2570 | 2782 |
| Criterion 1 – Borderline vs stage II-IV | 714 | 624 |
| Criterion 1 – Borderline vs sec meta | 1484 | 1389 |
| Criterion 1 – Stage I vs stage II-IV | 3335 | 1575 |
| Criterion 1 – Stage I vs sec meta | 4293 | 3947 |
| Criterion 1 – Stage II-IV vs sec meta | 8011 | 2424 |
| Criterion 2 – Model overfitting | 895 | 895 |
| Criterion 3 – Benign | 616 | 616 |
| Criterion 3 – Borderline | 203 | 203 |
| Criterion 3 – Stage I | 161 | 161 |
| Criterion 3 – Stage II-IV | 372 | 372 |
| Criterion 3 – Sec metastasis | 124 | 124 |

Criterion 1: Overfitting control for every pair of outcome categories
Criterion 2: Overfitting control for the model in general
Criterion 3: Precise estimation of the overall risk for each outcome category

#### S2.2 MISSING DATA HANDLING

Missing values for CA125 and tumour outcome were handled by multiple imputation using the following variables in the imputation model: presumed endometrioma based on subjective assessment of the ultrasound examiner (yes/no), subjective assessment (ordinal: certainly benign, probably benign, uncertain, probably malignant, certainly malignant), age (in years), type of centre (oncology centre vs other), maximum lesion diameter in mm (logtransformed), proportion solid tissue (defined as the maximum diameter of the largest solid component divided by the maximum lesion diameter, bounded between 0 and 1), proportion of solid tissue squared, number of locules (0, 1, 2, 3, 4, 5-10, >10; treated categorically), number of papillary projections (ordinal: 0, 1, 2, 3, >3), acoustic shadows (yes/no), ascites (yes/no), bilaterality (yes/no), tender mass during examination (yes/no), personal history of ovarian cancer (yes/no), papillary height in mm, papillary projections with blood flow (yes/no), colour score (ordinal: no, minimal, moderate, very strong), echogenicity of cyst fluid (categorical), irregular cyst walls (yes/no), and whether benign descriptors apply or not (yes/no). Additionally, CA125 (IU/L, using log(log(CA125+1))) was used to impute outcome, and outcome (categorical: benign, borderline, stage I, stage II-IV, secondary metastasis) was used to impute CA125.

For the five likely malignant outcomes, we planned to pragmatically fix the outcome to the most commonly imputed malignant outcome following multiple imputation. However, due to a coding error the most likely outcome was assigned to these patients. This resulted in two patients' outcomes being fixed as benign. According to the predefined statistical plan, these patients' outcomes should have been fixed as borderline instead of benign.

Each imputation in mice was obtained after 50 iterations. Convergence plot is shown in **Figure S1.** Density and proportion plots for the two imputed characteristics (CA125 and tumour outcome) are shown in **Figures S2-S3.**

For the leave-one-centre-out validation, we generated multiply imputed datasets for each centre-specific validation. In this approach the model is trained in all centres except one left out centre and then tested in the left out centre. Therefore, 17 different multiply imputed datasets were created imputing the missing values only using information of the 16 training centres for each validation. This strategy was not pre-specified in the protocol, but we decided to adapt it to avoid leakage and overoptimistic results.

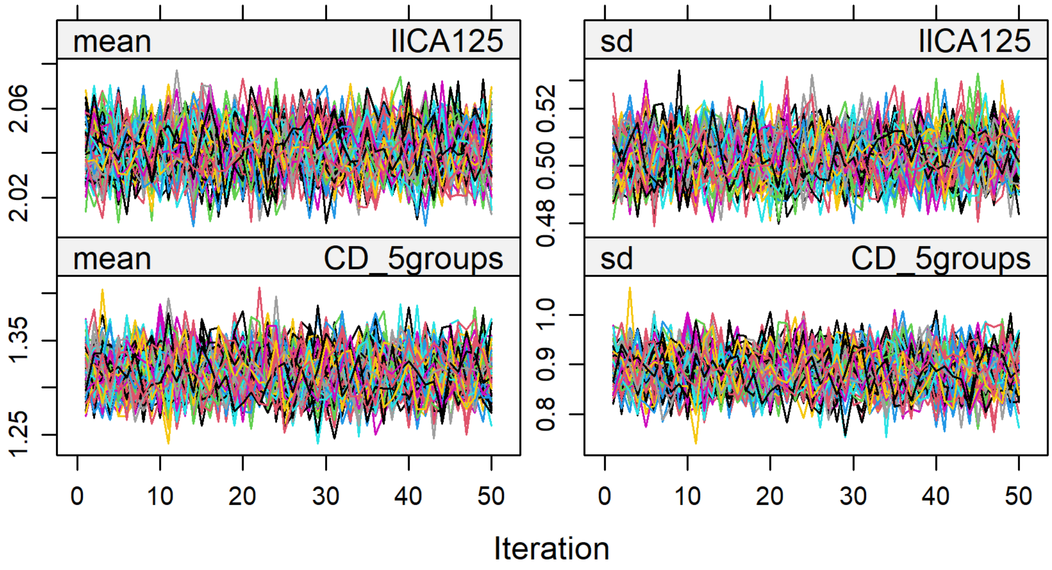

**Figure S1. Convergence plots for CA125 and the multinomial reference standard. The plots on the top row refer to CA125 (after log-log transformation; ll_CA125), the plots on the bottom row refer to the multinomial reference standard (CD_5groups). The plots on the left show the mean value, the plots on the right show the standard deviation (sd). The x-axis refers to the iteration (1 to 50). The coloured lines refer to the 100 imputations. The multinomial reference standard was used as a nominal variable in the imputations, despite it being represented as a numerical variable in these plots.**

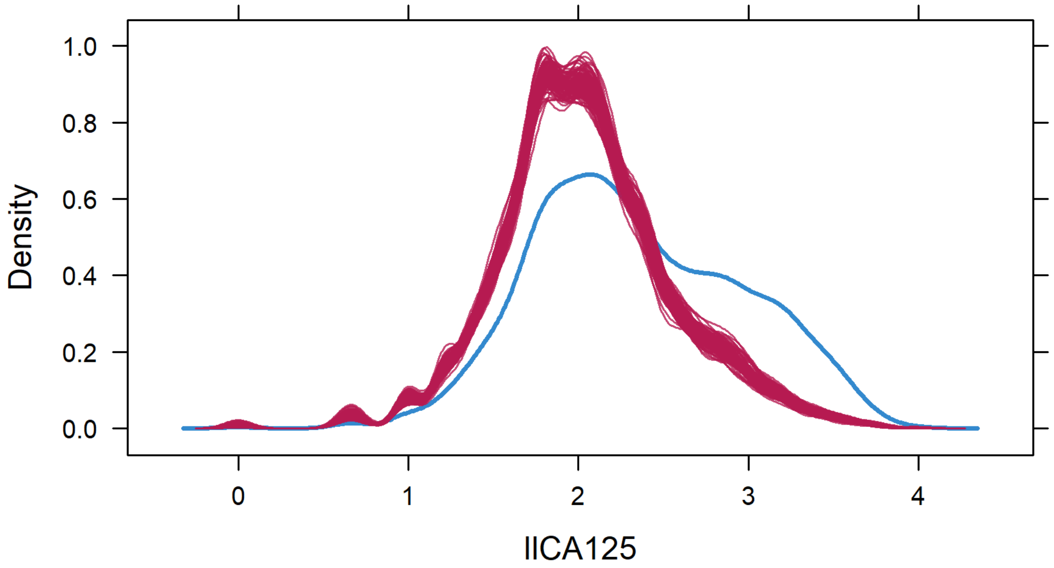

**Figure S2. Density plots of log(log(CA125+1)). The blue curve is the density of the values that were observed (i.e. were not missing), the red curves are the densities for each of the 100 imputations of the values that were missing. There were fewer high imputed than observed values. ll_CA125, log(log(CA125+1)).**

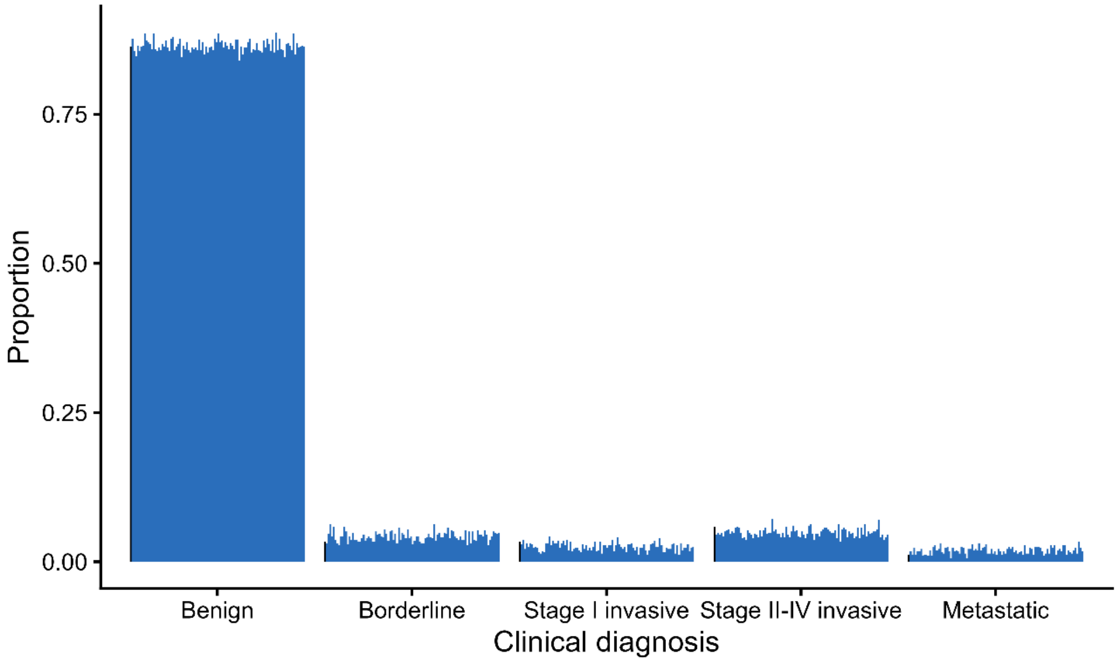

**Figure S3. Proportion plot of the clinical diagnosis. The black line refers to the observed clinical diagnosis (omitting missing values) and each of the lines represent the proportion in one imputation.**

#### S2.3 METHODOLOGICAL DETAILS

##### S2.3.1 Model updating

ADNEX2 was obtained by recalibrating the model to update intercepts and coefficients using slopes and then using these recalibrated coefficients and standard error of the original ADNEX model as priors for a Bayesian refitting using random intercepts per centre. Random intercepts were set to 0 for model validation.

This means that per linear predictor, each predictor coefficient is changed to the same degree by multiplying with the slope of the linear predictor. This fixes potential issues of ADNEX with the average estimated risk and the spread of the estimated risks. This required the estimation of 20 parameters, 1 intercept and 4 slopes per category [4]. This step was performed using the `multinom()` function of the **nnet** R package [5]. In the second step, we used Bayesian refitting [6]**.** The priors for the intercepts and coefficients were normally distributed with the updated value from step 1 as the means. The variances were the squared standard errors of the coefficients in the original ADNEX analysis for coefficients, and a large fixed value of 10 for the intercepts to avoid miscalibration-in-the-large. We realise that 10 was selected arbitrarily to ensure flat priors on the intercepts. The priors implement shrinkage towards the values obtained in step 1. This required the estimation of 40 or 44 parameters depending on if the model includes CA125 or not. We did not update the functional forms of the continuous predictors because the forms used in ADNEX are reasonable, and the data was not sufficient to properly update these complex terms. The Bayesian refitting was performed using the **brms** package with 10000 iterations and 2500 warmup rounds [7–9]. The Bayesian models in the leave-one-center-out cross validation were fitted with 5000 iterations and 2500 warmup rounds, which was sufficient to achieve convergence. For the model refitting, the resources and services used were provided by the VSC (Flemish Supercomputer Centre), funded by the Research Foundation - Flanders (FWO) and the Flemish Government.

##### S2.3.2 Confidence and prediction intervals

Centre specific performance is obtained by pooling the performance of the 100 imputations. The standard error of the centre specific performance is obtained applying Rubin's rule which is defined as $SE_{total} = \sqrt{V_B + V_W + \frac{V_B}{100}}$, where $V_B$ is the centre-specific variance between imputations and $V_W$ the within imputation variance. Then this $SE_{total}$ is used to pool the centre specific performance according to the different techniques used for each metric to obtain the confidence and prediction intervals.

##### S2.3.3 Area under the ROC curve

With respect to the area under the ROC curve (AUC), we obtained the logit(AUC) and its standard error. For this, the `auc.nonpara.mw()` function of the **auRoc** package was used. The logit(AUC) values were combined using Rubin's rules per centre to obtain centre-specific results. 95% confidence intervals for logit(AUC) were calculated, and then the point estimate and the confidence limits were back-transformed to the original scale. An overall AUC was obtained by combining centre-level logit(AUC) and its standard error using random effects meta-analysis. This meta-analysis (including the calculation of the prediction interval) was performed with Bayesian meta-analysis with the `valmeta()` function from **metamisc** package and "`REML`" estimator [10]. The process is encapsulated in the function `AUC_imp_IOTA()`.

##### S2.3.4 Polytomous discrimination index (PDI)

PDI was obtained per imputation and then averaged over imputations without accounting for centre-specific performance due to low numbers of some malignant cancer subtypes. PDI is defined in Van Calster et al. [11]. CIs are obtained using 100 bootstraps. The function to calculate PDI is `pdi5v0()`.

##### S2.3.5 Sensitivity, Specificity, Positive Predictive Value and Negative Predictive value

To deal with multiply imputed data, sensitivity and specificity values were first logit-transformed. Within each center, these logit-transformed estimates and their standard errors were combined across imputed datasets using Rubin's rules to obtain center-specific results. These center-specific estimates (on the logit scale) were then pooled using a bivariate random-effects meta-analysis to calculate the overall sensitivity and specificity[12]. Finally, all pooled estimates and their confidence intervals were back-transformed to the original probability scale. The similar process was followed for PPV/NPV. The function for Sensitivity/Specificity is `SS_imp()` and for PPV/NPV `PNPV_imp()`.

##### S2.3.6 Observed over expected ratio (O:E)

Centre specific results were obtained and pooled using Rubin's rules with within imputation variance obtained using 100 bootstraps. O:E ratio was meta-analysed in log scale as recommended by Snell et al. [13]. The function to obtain the centre specific and meta-analysed O:E is encapsulated `boot_cal_meta()` and uses `valmeta()` from **metamisc** package for O:E [10,14].

##### S2.3.7 Calibration curves

The overall calibration plot is obtained following the methodologies of Barreñada et al. [15] adapted for multiply imputed data. We first calculated the centre specific flexible calibration curves with restricted cubic splines and three knots for the 100 imputations in a grid of 100 points evenly separated from 0.01 to 0.99. Then we averaged per point in the grid the estimated observed proportions and meta-analysed per point in the grid to obtain the overall curve with prediction intervals. Then, to obtain the centre specific curves we used the MIX-C approach and simply averaged the results per imputation. Therefore, the overall curve is based on the MA2-C approach and the centre specific curves on MIX-C approach as recommended by the authors [15]. The overall curves and PI are obtained with the function `cal_meta()` and the centre specific curves with the `valProbCluster()` function from the **CalibrationCurves** package [15–17].

Multinomial calibration curves were not calculated using meta-analysis of centre-specific results due to low number of events in some subgroups. We calculated the multinomial curves per imputation using vector generalised additive models (VGAM) with smoothing splines from the **VGAM** package and then averaged over the 100 imputations. The function to perform this is `multinom_cal_MI()`.

##### S2.3.8 Clinical utility

We conducted a random effects meta-analysis of Net Benefit (NB) of ADNEX and ADNEX2 at the decision thresholds from 0.01 to 0.40 using the methodology described in Wynants et al. [18]. Relative Utility (RU) expresses NB as a percentage of the maximum possible utility: RU=1 indicates maximum possible utility, RU=0 means no utility, RU<0 means harm. No utility means that NB of the model is not higher than NB of the default of strategies of treating all patients ($NB_{TA}$) or treating no one ($NB_{TN}$, which is 0 by definition). In that case, the model is not better than simply assuming that everyone needs treatment or that no one needs treatment without the use of any model. Harm means that NB of the model is lower than $NB_{TA}$ or $NB_{TN}$. Harm means that you can make better decisions without the model by treating all or none of the patients. Delta net benefit ($\Delta NB$) is defined as the difference between the NB of the model and the NB of the best default strategy ($NB - \max(\mathrm{NB_{TN}}, NB_{TA})$). The methodology obtains the probability of being useful (p-useful)

NB, $NB_{TA}$, and RU are defined as follows:

$$NB = Se * P - w * (1 - Sp) * (1 - P),\ NB_{TA} = P - w * (1 - P),\ RU = \frac{\Delta\, NB}{P - \max(\mathrm{NB_{TN}}, NB_{TA})},$$

with $Se$ sensitivity, $P$ prevalence of malignancy, $Sp$ specificity, and $w$ the odds of the risk threshold. In our case, $w = \frac{DT}{1-DT}$ with DT being the decision threshold.

The Bayesian framework also allows to obtain the probability that the model is useful in a new centre (p-useful) by sampling hypothetical new centres from the posterior distribution of the model. P-useful is defined as $P\ [NB > \max(\mathrm{NB_{TN}}, NB_{TA})]$ and measures the centre specific heterogeneity of clinical utility.

For the Bayesian meta-analysis, we used Markov chain Monte Carlo (MCMC) sampling with 2000 samples per chain and a burn-in of 1000, and two chains. This was sufficient for convergence and to have $\left|1-\hat{R}\right| < 0.01$. We obtain performance metrics per imputation and then averaged over the 100 imputations. The function to perform this analysis is `MA_NB_tri_imp()` and the jags code for the model is included in the repository.

### S3. DEVIATIONS FROM STATISTICAL ANALYSIS PLAN (SAP)

We followed a pre-registered statistical analysis plan available in the OSF ((https://osf.io/36n8m/overview?view_only=0f1fa6d9a5e34e4d901c05922d0d0307), registered the 30th of September 2025. However, in some occasions we had to deviate from it.

1. **Sample size calculation:** There was an error in the number of coefficients per equation for the sample size calculation. The SAP said we needed 14 but we only need 11 per equation. This resulted in a smaller required sample size. Therefore it had no effect on the analysis nor results. We include in the supplementary material (S2.1) the correct sample size calculation.
2. **Subgroup and management definition change:** In the SAP we defined three possible managements. [(a) patients that were operated without a follow up scan, irrespective of the time until surgery ("immediate surgery"), (b) conservatively managed patients that have at least one follow up scan and (c) patients with unknown management, i.e. patients without follow-up information or that died before surgery or a follow up visit]. In the manuscript we redefined the immediately operated group to a surgery without follow up scan within 1 year from inclusion. Therefore, the unknown management also includes patients that were operated after 1 year without any follow up scan, because we considered it possible that there could have been unknown or unrecorded conservative follow-up visits prior to surgery when the surgery was done more than 1 year after the inclusion scan. Additionally, we redefined the subgroup of immediately operated patients to a subgroup of patients without any follow up scan that were operated within 120 days from inclusion scan. We used 120 days because this was the timeframe used to define the outcome according to **Table 1** and previous analysis on IOTA5 data.
3. **Imputation of tumours presumed to be malignant:** Five cases with uncertain outcome but presumed to be malignant were planned to be imputed with the most common malignant outcome. However, due to a coding error we imputed them with the most common imputed outcome which resulted in two of those cases being imputed as benign. The correct classification according to the SAP would have been borderline but given that the outcome for these patients is uncertain and that according to the patient's characteristics the most likely outcome was benign, we decided to keep these tumours' classification as benign. Given the sample size of the study, the impact of this change on the final coefficients of ADNEX2 or in the results of the validations is negligible.
4. **Prior variance of the intercepts:** To refit ADNEX2 we used as variance of the priors in a Bayesian framework the standard errors of original ADNEX coefficients. In the SAP we wrote

that this would apply to coefficients and intercepts. However, we realised that setting such an informative prior in the intercepts was too strong and would result in an incorrect model. Therefore, we reduced the informativeness of the priors and set the variance to 10 to allow the model to estimate the intercepts from the data.

### S4. Figures and Tables

**Most of the figures and tables included below can be explored in a interactive website that also includes code to generate the different tables and figures (https://adnex2-565575.pages.gitlab.kuleuven.be/).**

#### S4.1 Descriptives

**Table S4. Tumour outcome for tumours to which a benign descriptor applied**

| Benign Descriptor | N | Benign, n (%) | Borderline, n (%) | Stage I, n (%) | Stage II – IV, n (%) | Sec. meta, n (%) | Uncertain, n (%) |
|---|---|---|---|---|---|---|---|
| Any descriptor | 2190 | 1987 (90.7) | 4 (0.2) | 2 (0.1) | 2 (0.1) | 2 (0.1) | 193 (8.8) |
| Descriptor 1 | 615 | 553 (89.9) | 0 (0) | 0 (0) | 0 (0) | 0 (0) | 62 (10.1) |
| Descriptor 2 | 281 | 267 (95.0) | 0 (0) | 0 (0) | 1 (0.4) | 0 (0) | 13 (4.6) |
| Descriptor 3 | 836 | 755 (90.3) | 2 (0.2) | 0 (0) | 1 (0.1) | 1 (0.1) | 77 (9.2) |
| Descriptor 4 | 458 | 412 (90.0) | 2 (0.4) | 2 (0.4) | 0 (0) | 1 (0.2) | 41 (9.0) |
| No descriptor | 4657 | 2638 (56.6) | 387 (8.3) | 301 (6.5) | 785 (16.9) | 227 (4.9) | 319 (6.9) |

Sec.meta, secondary metastasis

**Table S5. Tumour outcome for tumours to which a benign descriptor applied (multiple imputation results)**

| Benign Descriptor | N | Benign, % (95% CI) | Borderline, % (95% CI) | Stage I, % (95% CI) | Stage II – IV, % (95% CI) | Sec. meta, % (95% CI) | Malignant % (95% CI) |
|---|---|---|---|---|---|---|---|
| Any descriptor | 2190 | 99.5 (99.1-99.8) | 0.2 (<0.1-0.4) | 0.1 (<0.1-0.3) | 0.1 (<0.1-0.3) | 0.1 (<0.1-0.3) | 0.5 (0.2-0.9) |
| Descriptor 1 | 615 | 99.9 (99.7->99.9) | <0.1 (<0.1-0.2) | <0.1 (<0.1-0.1) | <0.1 (<0.1-0.2) | <0.1 (<0.1-0.1) | 0.1 (<0.1-0.3) |
| Descriptor 2 | 281 | 99.6 (98.9->99.9) | <0.1 (<0.1-<0.1) | <0.1 (<0.1-0.1) | 0.4 (<0.1-1.1) | <0.1 (<0.1-0.1) | 0.4 (<0.1-1.1) |
| Descriptor 3 | 836 | 99.4 (98.8->99.9) | 0.3 (<0.1-0.7) | <0.1 (<0.1-0.1) | 0.2 (<0.1-0.5) | 0.1 (<0.1-0.4) | 0.6 (<0.1-1.2) |
| Descriptor 4 | 458 | 98.8 (97.8-99.8) | 0.5 (<0.1-1.2) | 0.5 (<0.1-1.1) | <0.1 (<0.1-0.1) | 0.2 (<0.1-0.7) | 1.2 (0.2-2.2) |
| No descriptor | 4657 | 62.0 (60.6-63.5) | 8.8 (7.9-9.6) | 6.7 (6.0-7.5) | 17.4 (16.3-18.5) | 5.1 (4.4-5.7) | 38.0 (36.5-39.4) |

CI: Confidence interval; Sec.meta, secondary metastasis

**Table S6. Number of patients by tumour outcome in each participating centre**

| Centre | Code | N | Benign | Borderline | Stage I | Stage II-IV | Metastasis | Uncertain |
|---|---|---|---|---|---|---|---|---|
| Malmö, Sweden | MSW | 1093 | 901 | 33 | 22 | 45 | 16 | 76 |
| Rome, Italy* | RIT | 1019 | 617 | 56 | 54 | 144 | 39 | 109 |
| Leuven, Belgium* | LBE | 754 | 505 | 57 | 30 | 95 | 24 | 43 |
| Milan 1, Italy* | CIT | 558 | 284 | 38 | 48 | 160 | 14 | 14 |
| Stockholm, Sweden* | SSW | 509 | 266 | 43 | 41 | 89 | 38 | 32 |
| Genk, Belgium | GBE | 473 | 370 | 16 | 10 | 17 | 11 | 49 |
| Monza, Italy* | OIT | 431 | 267 | 33 | 27 | 55 | 11 | 38 |
| Prague, Czech Rep* | PCR | 414 | 208 | 50 | 30 | 59 | 25 | 42 |
| Trieste, Italy | TIT | 273 | 227 | 18 | 4 | 12 | 8 | 4 |
| Cagliari, Italy | SIT | 247 | 202 | 14 | 1 | 15 | 9 | 6 |
| Katowice, Poland* | MPO | 241 | 183 | 10 | 8 | 14 | 0 | 26 |
| Milan 2, Italy* | NCI | 227 | 105 | 14 | 22 | 53 | 19 | 14 |
| London, UK | IUK | 172 | 147 | 2 | 1 | 4 | 0 | 18 |
| Pamplona, Spain* | PSP | 170 | 112 | 4 | 4 | 21 | 12 | 17 |
| Milan 3, Italy | MIT | 129 | 120 | 1 | 0 | 1 | 1 | 6 |
| Florence, Italy | FLI | 85 | 73 | 1 | 1 | 0 | 0 | 10 |
| Lisbon, Portugal* | LIP | 52 | 38 | 1 | 0 | 3 | 2 | 8 |
| Oncology centres | | 4375 | 2585 (59%) | 306 (7%) | 264 (6%) | 693 (16%) | 184 (4%) | 343 (8%) |
| Other centres | | 2472 | 2040 (83%) | 85 (3%) | 39 (2%) | 94 (4%) | 45 (2%) | 169 (7%) |
| TOTAL | | 6847 | 4625 (68%) | 391 (6%) | 303[a] (4%) | 787 (11%) | 229 (3%) | 512 (7%) |

[a] This includes one patient with missing FIGO stage, as of 24 February 2025.* These centres are oncology centres. Percentages are row percentages

**Table S7. Number of patients with tumours to which benign descriptors did not apply by tumour outcome in the cohort used to update ADNEX to ADNEX2.**

| Centre | N | Benign | Borderline | Stage I | Stage II-IV | Metastasis | Uncertain |
|---|---|---|---|---|---|---|---|
| Malmö | 715 | 547 | 33 | 22 | 45 | 16 | 52 |
| Rome* | 733 | 379 | 54 | 53 | 144 | 39 | 64 |
| Leuven* | 533 | 301 | 57 | 30 | 94 | 24 | 27 |
| Milan 1* | 461 | 192 | 38 | 48 | 160 | 13 | 10 |
| Stockholm* | 465 | 223 | 43 | 41 | 89 | 38 | 31 |
| Genk | 254 | 173 | 16 | 10 | 17 | 10 | 28 |
| Monza* | 298 | 148 | 33 | 27 | 55 | 11 | 24 |
| Prague* | 318 | 123 | 50 | 30 | 59 | 25 | 31 |
| Trieste | 173 | 128 | 17 | 4 | 12 | 8 | 4 |
| Cagliari | 129 | 88 | 14 | 1 | 15 | 9 | 2 |
| Katowice* | 95 | 54 | 10 | 7 | 14 | 0 | 10 |
| Milan 2* | 184 | 67 | 14 | 22 | 52 | 19 | 10 |
| London | 71 | 55 | 2 | 1 | 4 | 0 | 9 |
| Pamplona* | 79 | 34 | 4 | 4 | 21 | 12 | 4 |
| Milan 3 | 78 | 72 | 1 | 0 | 1 | 1 | 3 |
| Florence | 31 | 26 | 0 | 1 | 0 | 0 | 4 |
| Lisbon* | 40 | 28 | 1 | 0 | 3 | 2 | 6 |
| Oncology centres | 3206 | 1549 (48%) | 304 (9%) | 262 (8%) | 691 (22%) | 183 (6%) | 217 (7%) |
| Other centres | 1451 | 1089 (75%) | 83 (6%) | 39 (3%) | 94 (6%) | 44 (3%) | 102 (7%) |
| TOTAL | 4657 | 2638 (57%) | 387 (8%) | 301[a] (6%) | 785 (17%) | 227 (5%) | 319 (7%) |

[a] This includes one patient with missing FIGO stage, as of 24 February 2025.

* These centres are oncology centres. Percentages are row percentages

**Table S8. Number of patients with missing CA125 values by management.**

| | Total | | Surgically managed | | Conservatively managed | | Unknown management | |
|---|---|---|---|---|---|---|---|---|
| Centre | N | CA125 missing, n (%) | N | CA125 missing, n (%) | N | CA125 missing, n, (%) | N | CA125 missing, n (%) |
| Malmö | 1093 | 736 (67%) | 470 | 132 (28%) | 583 | 568 (97%) | 40 | 36 (90%) |
| Rome* | 1019 | 615 (60%) | 577 | 283 (49%) | 351 | 270 (77%) | 91 | 62 (68%) |
| Leuven* | 754 | 422 (56%) | 423 | 124 (29%) | 301 | 279 (93%) | 30 | 19 (63%) |
| Milan 1* | 558 | 44 (8%) | 452 | 3 (0.7%) | 90 | 39 (43%) | 16 | 2 (13%) |
| Stockholm* | 509 | 90 (18%) | 379 | 28 (7%) | 116 | 56 (48%) | 14 | 6 (43%) |
| Genk | 473 | 367 (78%) | 254 | 175 (69%) | 176 | 156 (89%) | 43 | 36 (84%) |
| Monza* | 431 | 132 (31%) | 246 | 30 (12%) | 154 | 84 (55%) | 31 | 18 (58%) |
| Prague* | 414 | 135 (33%) | 256 | 57 (22%) | 130 | 63 (48%) | 28 | 15 (54%) |
| Trieste | 273 | 67 (25%) | 121 | 6 (5%) | 151 | 61 (40%) | 1 | 0 (0%) |
| Cagliari | 247 | 37 (15%) | 190 | 2 (1%) | 53 | 34 (64%) | 4 | 1 (25%) |
| Katowice* | 241 | 128 (53%) | 84 | 31 (37%) | 131 | 82 (63%) | 26 | 15 (58%) |
| Milan 2* | 227 | 29 (13%) | 147 | 3 (2%) | 73 | 25 (34%) | 7 | 1 (14%) |
| London | 172 | 112 (65%) | 19 | 5 (26%) | 143 | 100 (70%) | 10 | 7 (70%) |
| Pamplona* | 170 | 109 (64%) | 111 | 52 (47%) | 44 | 43 (98%) | 15 | 14 (93%) |
| Milan 3 | 129 | 115 (89%) | 52 | 43 (83%) | 72 | 67 (93%) | 5 | 5 (100%) |
| Florence | 85 | 80 (94%) | 31 | 31 (100%) | 49 | 44 (90%) | 5 | 5 (100%) |
| Lisbon* | 52 | 8 (15%) | 25 | 1 (4%) | 22 | 5 (23%) | 5 | 2 (40%) |
| Oncology centres | 4375 | 1712 (39%) | 2700 | 612 (23%) | 1412 | 946 (67%) | 263 | 154 (59%) |
| Other centres | 2472 | 1514 (61%) | 1137 | 394 (35%) | 1227 | 1030 (84%) | 108 | 90 (83%) |
| TOTAL | 6847 | 3226 (47%) | 3837 | 1006 (26%) | 2639 | 1976 (75%) | 371 | 244 (66%) |

* These centres are oncology centres.
Unknown management means that we have no information after the inclusion scan or that surgery was performed after 1 year from first scan without any follow up scan.

**Table S9. Number of patients with tumours to which the benign descriptors did not apply with missing CA125 values by management in the cohort used to update ADNEX to ADNEX2.**

| | Total | | Surgically managed | | Conservatively managed | | Unknown management | |
|---|---|---|---|---|---|---|---|---|
| Centre | N | CA125 missing, n (%) | N | CA125 missing, n (%) | N | CA125 missing, n (%) | N | CA125 missing, n (%) |
| Malmö | 715 | 426 (60%) | 372 | 97 (26%) | 318 | 308 (97%) | 25 | 21 (84%) |
| Rome* | 733 | 404 (55%) | 495 | 231 (47%) | 187 | 142 (76%) | 51 | 31 (61%) |
| Leuven* | 533 | 212 (40%) | 377 | 83 (22%) | 133 | 117 (88%) | 23 | 12 (52%) |
| Milan 1* | 461 | 23 (5%) | 398 | 2 (0.5%) | 51 | 21 (41%) | 12 | 0 (0%) |
| Stockholm* | 465 | 70 (15%) | 356 | 20 (6%) | 96 | 44 (46%) | 13 | 6 (46%) |
| Genk | 254 | 175 (69%) | 155 | 88 (57%) | 73 | 64 (88%) | 26 | 23 (88%) |
| Monza* | 298 | 65 (22%) | 198 | 18 (9%) | 85 | 38 (45%) | 15 | 9 (60%) |
| Prague* | 318 | 84 (26%) | 228 | 44 (19%) | 69 | 30 (43%) | 21 | 10 (48%) |
| Trieste | 173 | 34 (20%) | 103 | 6 (6%) | 69 | 28 (41%) | 1 | 0 (0%) |
| Cagliari | 129 | 8 (6%) | 112 | 0 (0%) | 15 | 8 (53%) | 2 | 0 (0%) |
| Katowice* | 95 | 40 (42%) | 57 | 23 (40%) | 28 | 14 (50%) | 10 | 3 (30%) |
| Milan 2* | 184 | 19 (10%) | 134 | 3 (2%) | 47 | 16 (34%) | 3 | 0 (0%) |
| London | 71 | 37 (52%) | 16 | 3 (19%) | 51 | 32 (63%) | 4 | 2 (50%) |
| Pamplona* | 79 | 29 (37%) | 67 | 17 (25%) | 8 | 8 (100%) | 4 | 4 (100%) |
| Milan 3 | 78 | 68 (87%) | 32 | 24 (75%) | 42 | 40 (95%) | 4 | 4 (100%) |
| Florence | 31 | 29 (94%) | 11 | 11 (100%) | 18 | 16 (89%) | 2 | 2 (100%) |
| Lisbon* | 40 | 5 (13%) | 21 | 0 (0%) | 17 | 4 (24%) | 2 | 1 (50%) |
| Oncology centres | 3206 | 951 (30%) | 2331 | 441 (19%) | 721 | 434 (60%) | 154 | 76 (49%) |
| Other centres | 1451 | 777 (54%) | 801 | 229 (29%) | 586 | 496 (85%) | 64 | 52 (81%) |
| TOTAL | 4657 | 1728 (37%) | 3132 | 670 (21%) | 1307 | 930 (71%) | 218 | 128 (59%) |

* These centres are oncology centres.
Unknown management means that we have no information after the inclusion scan or that surgery was performed after 1 year from first scan without any follow up scan.

**Table S10. Characteristics of patients and tumours in the ADNEX development cohort [19] (N = 5909, 24 centres) and ADNEX to ADNEX2 update cohort.**

| Variable | ADNEX | ADNEX2 |
|---|---|---|
| Number of cases | 5909 | 4657 |
| Benign | 3980 (67%) | 2638 (57%) [a]<br>2890 (62%)[b] |
| Borderline | 339 (6%) | 387 (8%) [a]<br>408 (9%)[b] |
| Stage I invasive cancer | 356 (6%) | 301 (6%) [a]<br>313 (7%)[b] |
| Stage II-IV invasive cancer | 988 (17%) | 785 (17%) [a]<br>810 (17%)[b] |
| Secondary Metastasis | 246 (4%) | 227 (5%) [a]<br>236 (5%)[b] |
| Uncertain | 0 (0%) | 319 (7%) [a] |
| Surgically managed | 5909 (100%) | 3132 (67%) |
| Conservatively managed | 0 (0%) | 1307 (28%) |
| Unknown management | 0 (0%) | 218 (5%) |
| Patient age at recruitment (years)* | 47 (35-60) [8-96] | 54 (41–66) [18–99] |
| Postmenopausal | 2426 (41%) | 2646 (57%) |
| Pelvic pain | 1092 (18%) | 594 (13%) |
| Presence of solid components | 3119 (53%) | 2801 (60%) |
| Proportion of solid components * | 0.11 (0-0.46) [0-1] | 0.25 (0–0.81) [0–1] |
| Maximum diameter of the lesion (mm)* | 69 (48-100) [8-550] | 65 (43–105) [7–751] |
| CA125 (U/ml)* | 30 (14-140) [1-40140] | 35 (14–184) [1–78200] |
| CA125 missing | 1805 (31%) | 1728 (37%) |
| Shadows* | 742 (13%) | 926 (20%) |
| Ascites* | 720 (12%) | 482 (10%) |
| Number of papillations* | | |
| None | 4771 (81%) | 3677 (79%) |
| 1 | 495 (8%) | 464 (10%) |
| 2 | 148 (3%) | 143 (3%) |
| 3 | 137 (2%) | 95 (2%) |
| 4 or more | 358 (6%) | 278 (6%) |
| More than 10 cyst locules* | 471 (8%) | 634 (14%) |
| Colour score of intratumoral flow | | |
| 1: No blood flow | 1635 (28%) | 1263 (27%) |
| 2: minimal blood flow | 1904 (32%) | 1314 (28%) |
| 3: moderate blood flow | 1637 (28%) | 1376 (30%) |
| 4: very strong flow | 733 (12%) | 704 (15%) |
| Tumour type using IOTA terminology | | |
| Unilocular | 1746 (30%) | 388 (8%) |
| Unilocular solid | 689 (12%) | 630 (13%) |
| Multilocular | 1036 (18%) | 1438 (31%) |
| Multilocular solid | 1252 (21%) | 1111 (24%) |
| Solid | 1177 (20%) | 1060 (23%) |
| Not possible to classify | 9 (<1%) | 30 (1%) |
| Number of oncology centres | 13/24 (54%) | 10/17 (59%) |

Results are presented as median (IQR) [range] for continuous variables, or N (%) for categorical variables.

[a] Outcome before imputing uncertain tumours

[b] Outcome after imputing uncertain tumours

*Predictors of ADNEX and ADNEX2

#### S4.2 Risk distributions

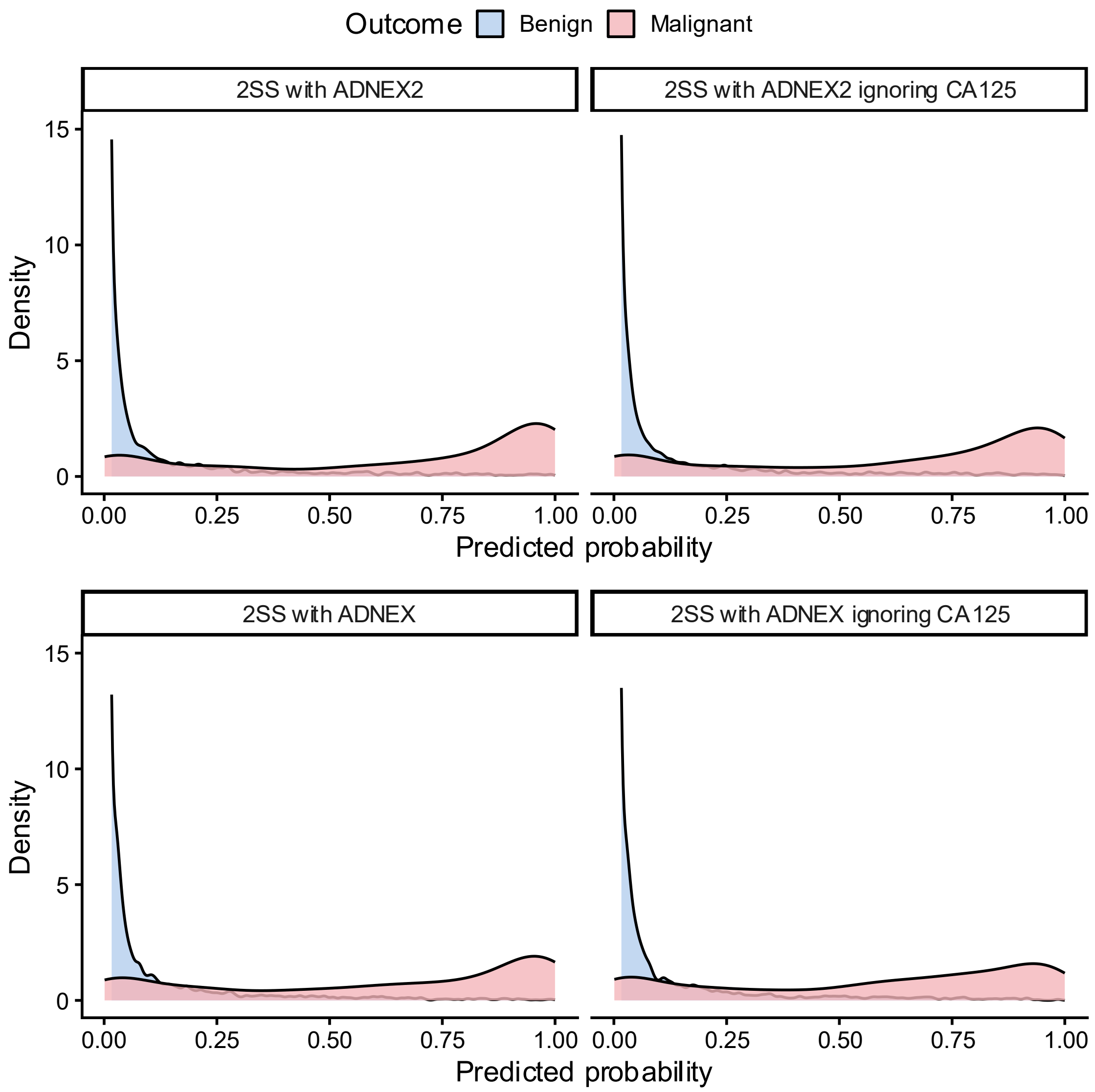

**Figure S4. Probabilities by tumour outcome in all patients (N = 6847) for two step strategy (2SS) with ADNEX2 and ADNEX. Predicted risks are averaged across imputations.**

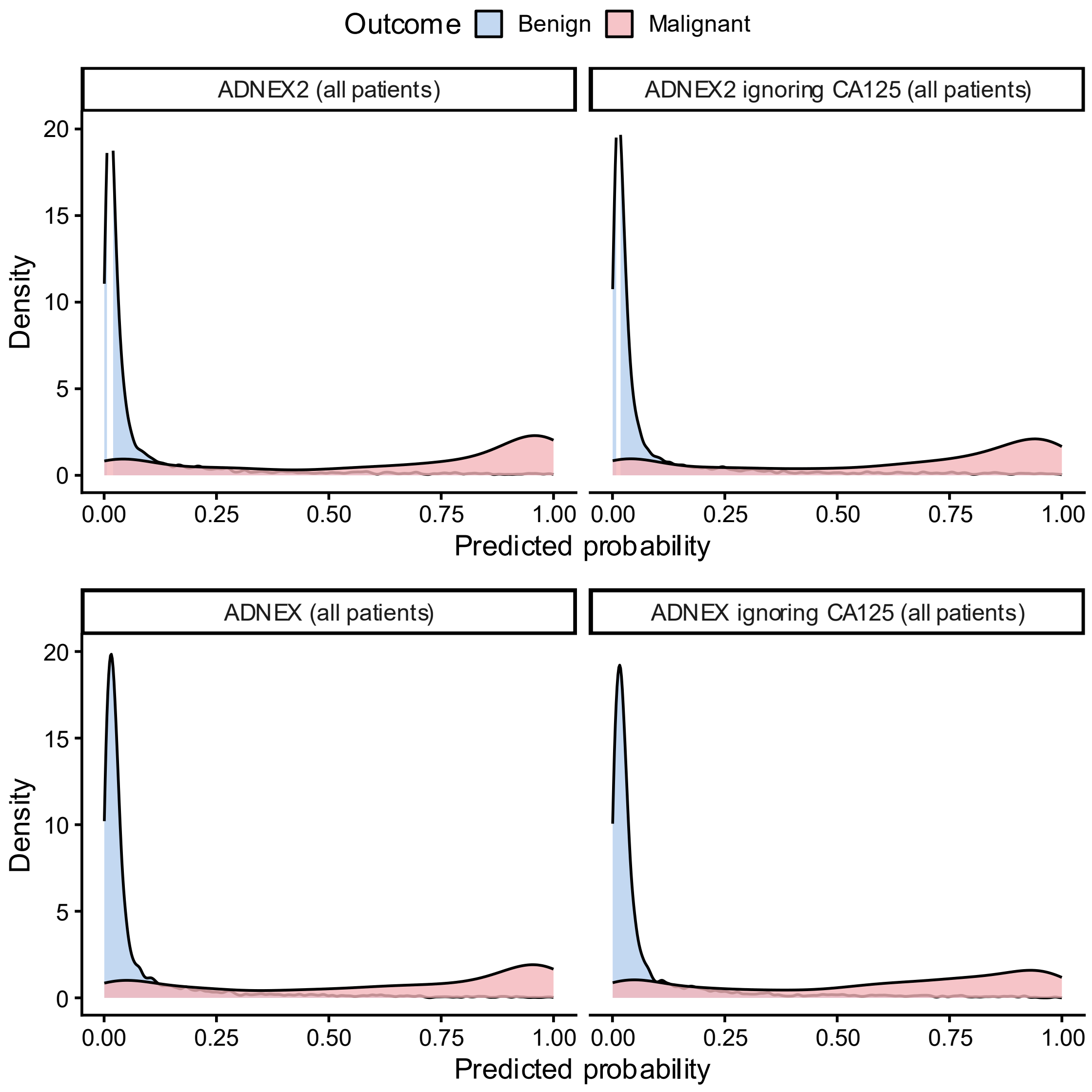

**Figure S5. Probabilities by tumour outcome in all patients (N = 6847) for ADNEX2 and ADNEX. Predicted risks are averaged across imputations.**

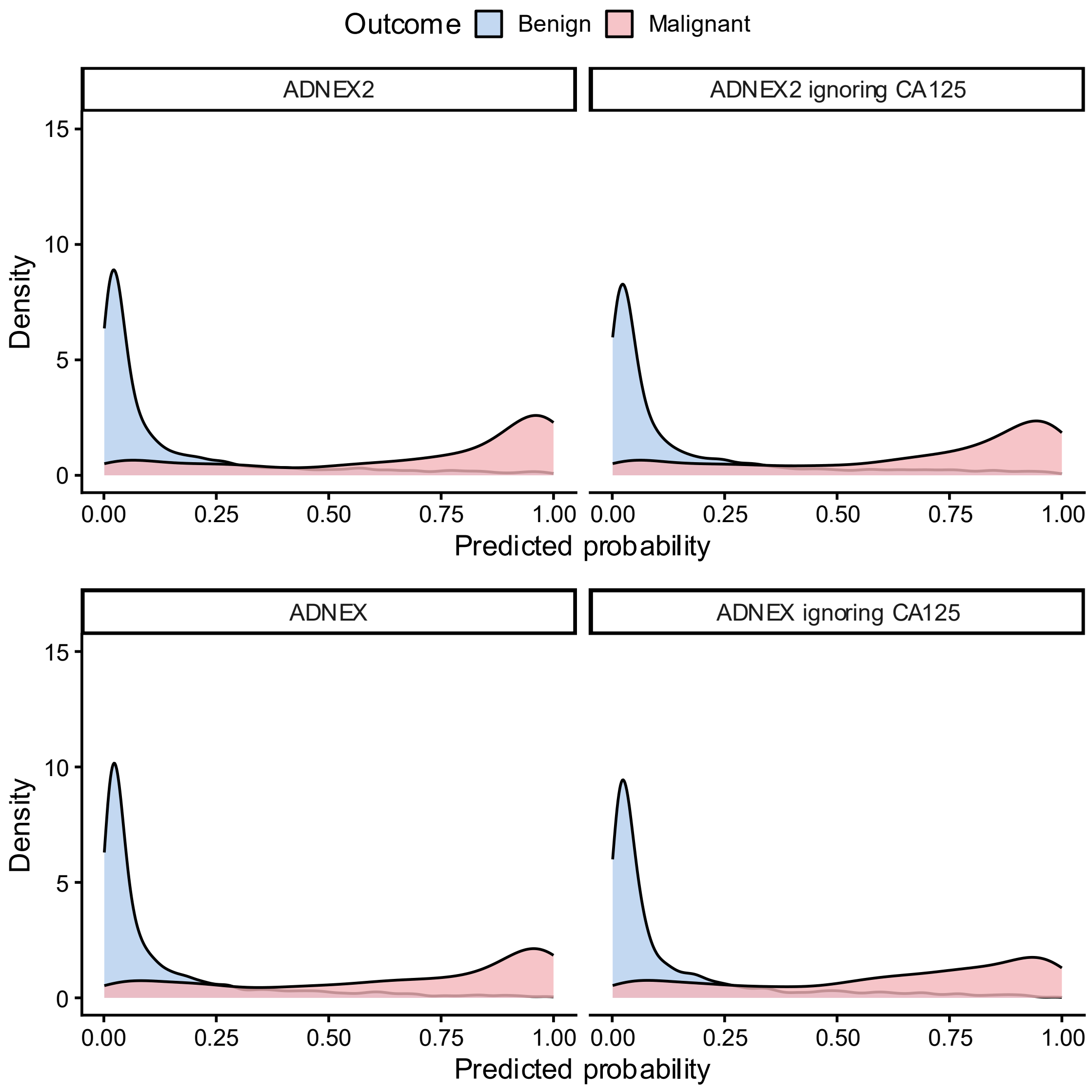

**Figure S6. Probabilities calculated by ADNEX2 and original ADNEX by tumour outcome in patients where benign descriptors did not apply (N = 4657). Predicted risks are averaged across imputations.**

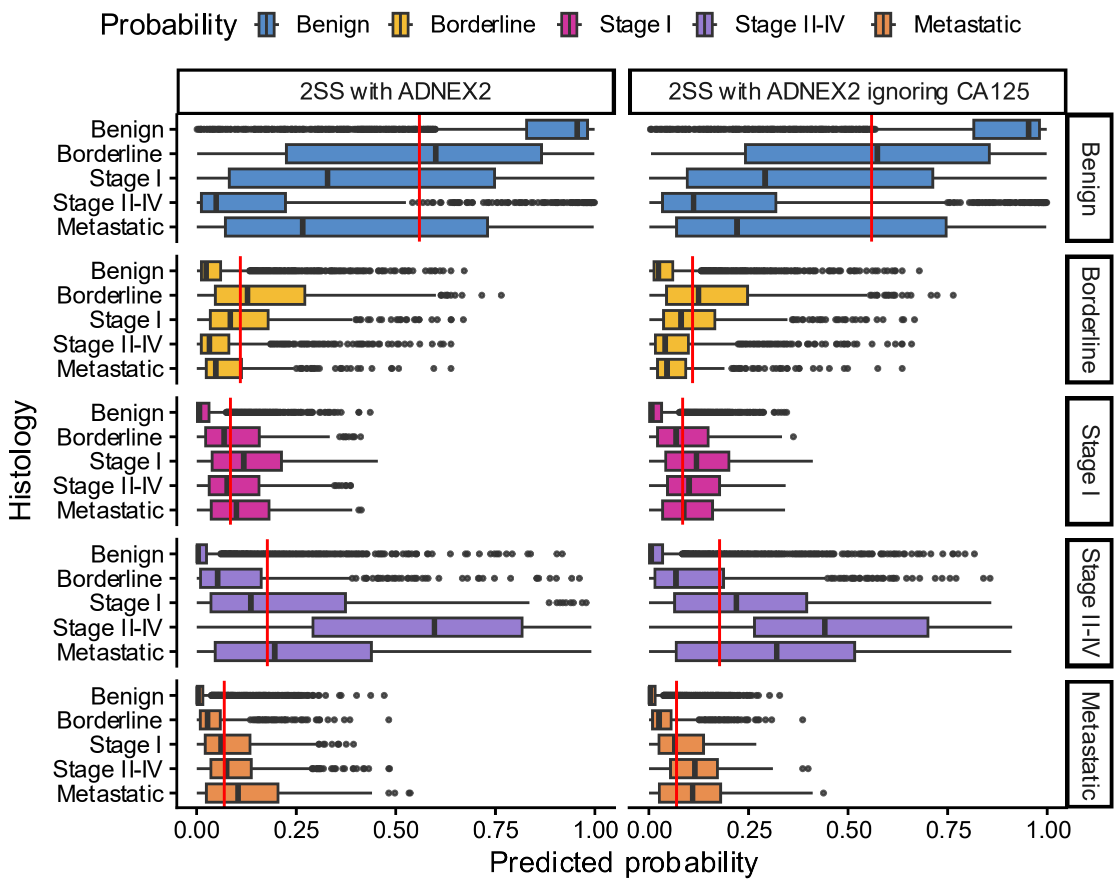

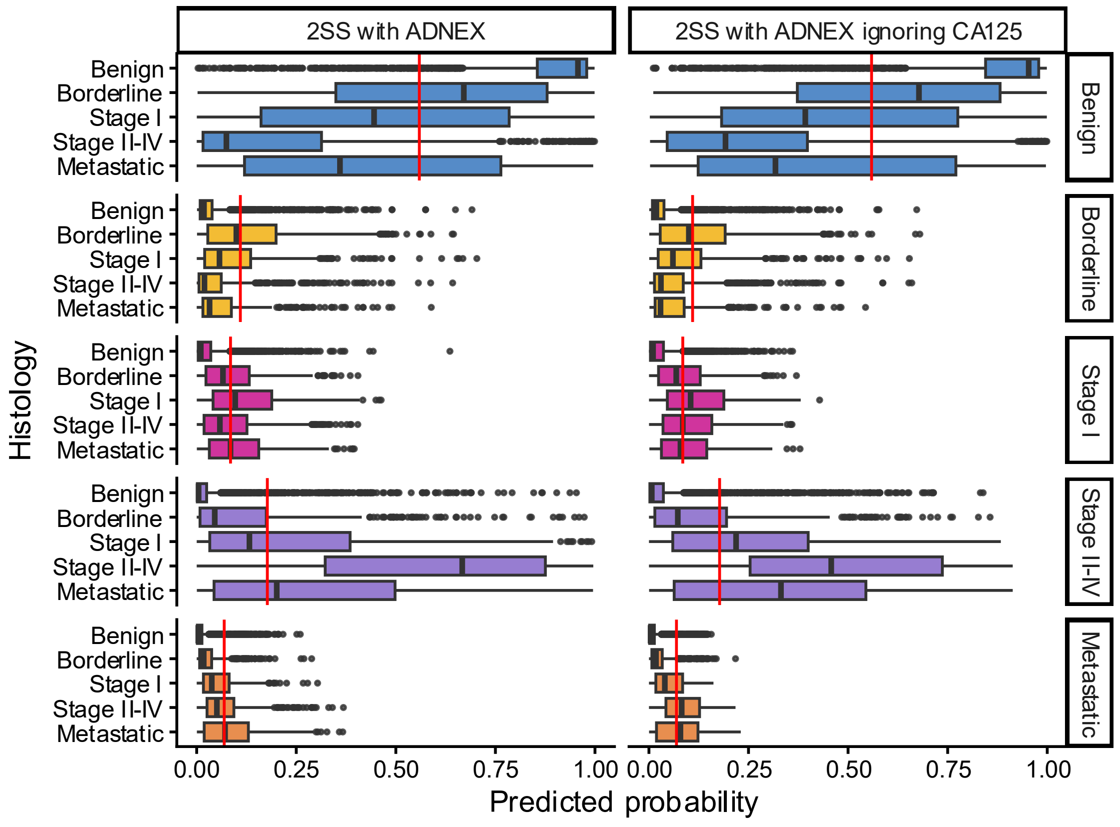

**Figure S7. Boxplot of predicted probability distribution by tumour subtype in all patients (N = 6847) for two step strategy (2SS) with ADNEX2 and ADNEX. Red line indicates baseline proportion per tumour subtype. Predicted risks are averaged across imputations.**

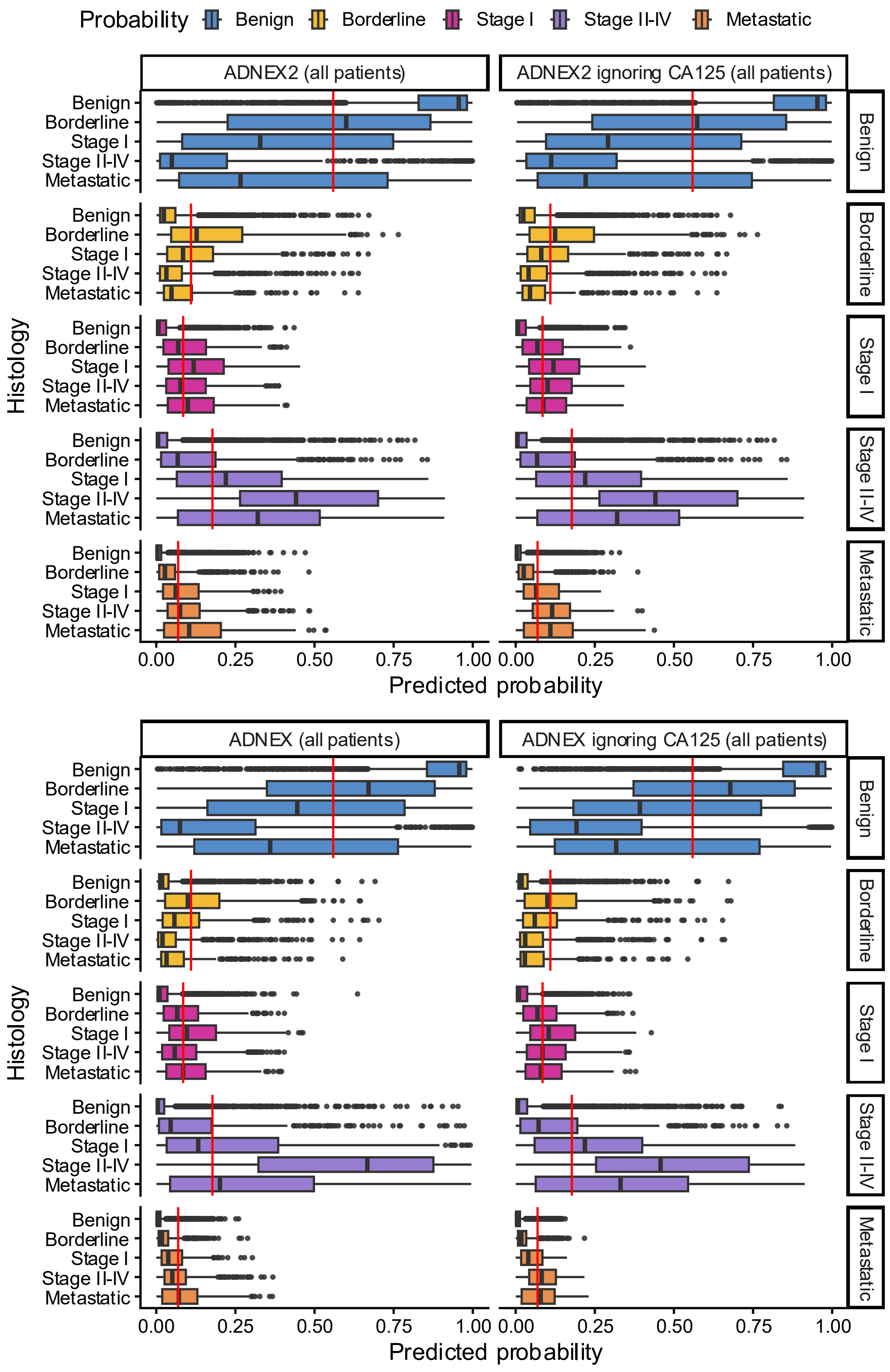

**Figure S8. Boxplot of predicted probability distribution by tumour subtype in all patients (N = 6847) for ADNEX2 and ADNEX. Red line indicates baseline proportion per tumour subtype. Predicted risks are averaged across imputations.**

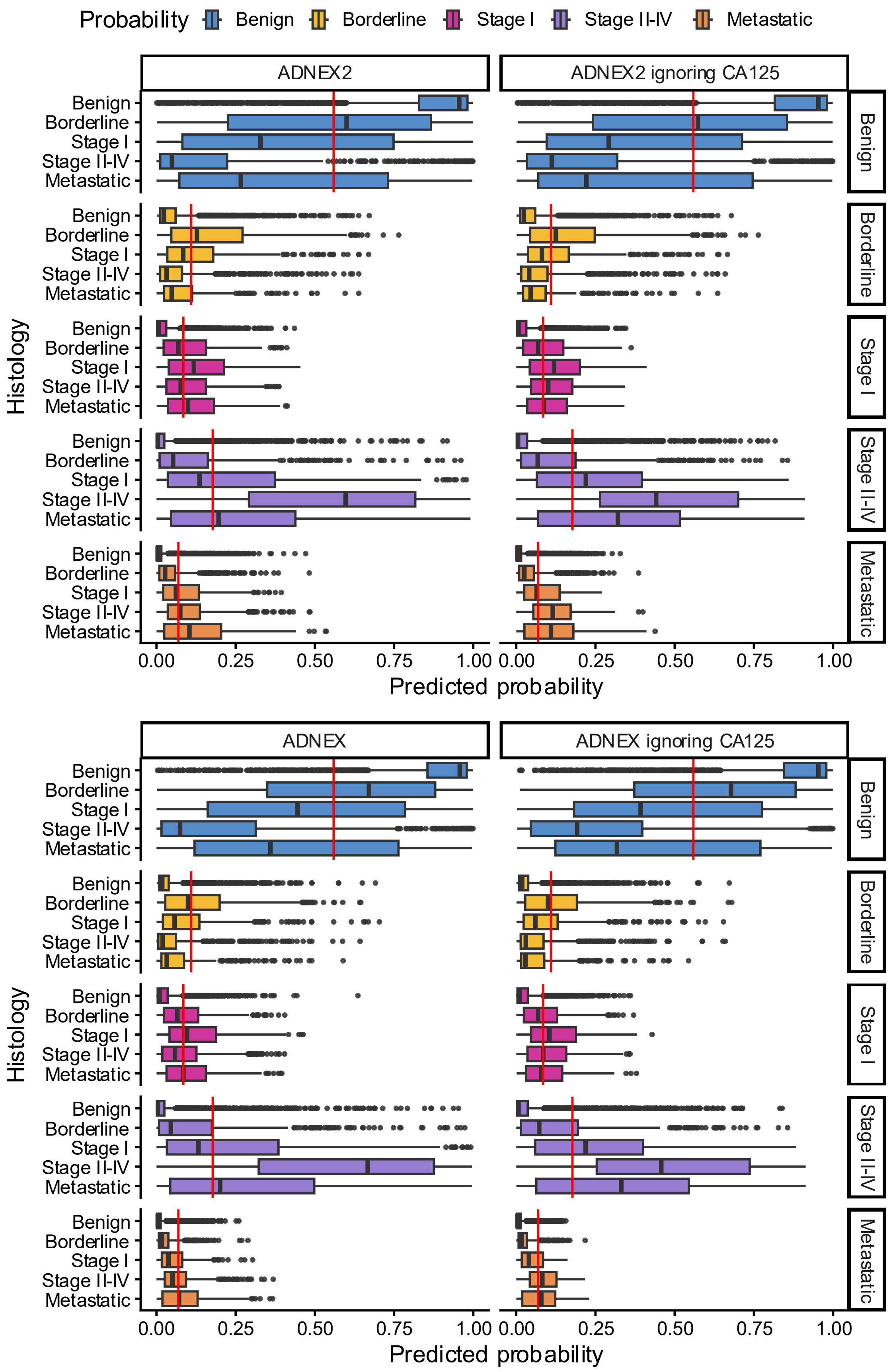

**Figure S9. Boxplot of predicted probability distribution by tumour subtype in patients where BDs do not apply (N = 4657) for ADNEX2 and ADNEX. Red line indicates baseline proportion per tumour subtype. Predicted risks are averaged across imputations.**

#### S4.3 ADNEX2 vs ADNEX

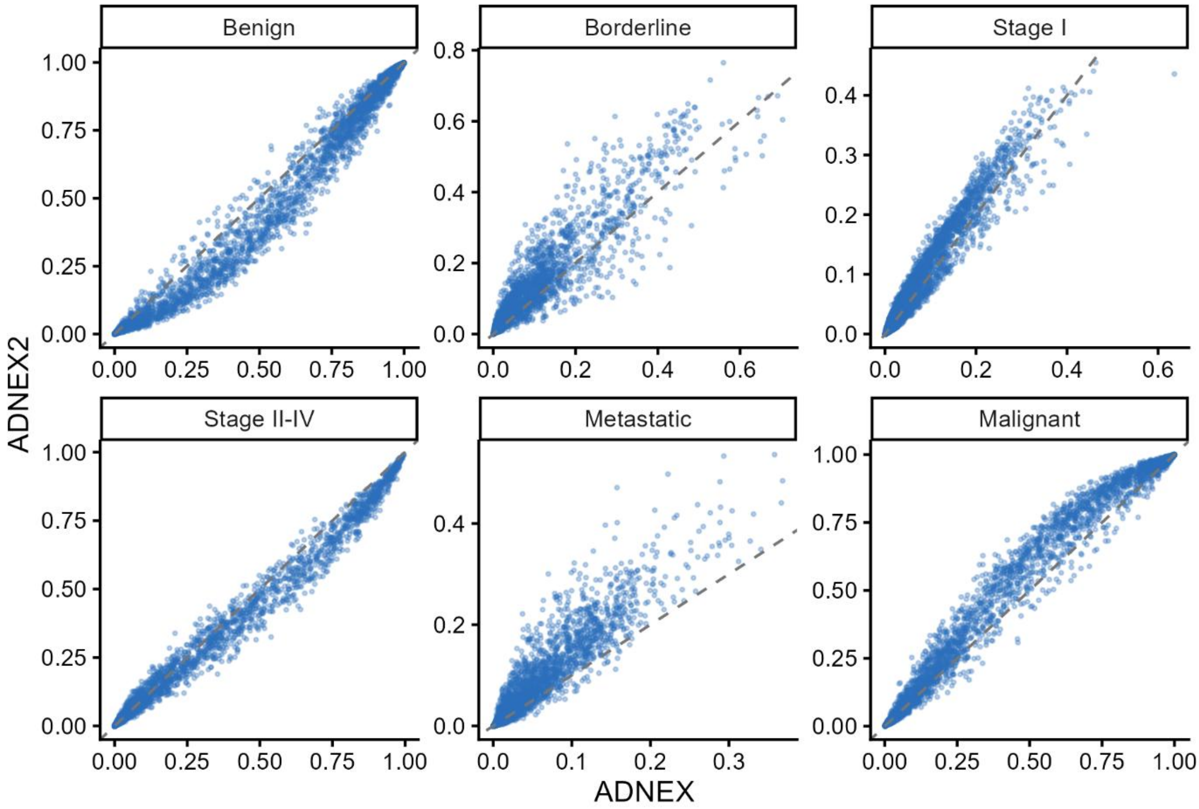

**Figure S10. Scatterplot of predicted risks of ADNEX and ADNEX2 in patients where BDs do not apply (N = 4657). Each point represents a patient. Patients' specific predictions are averaged across imputations.**

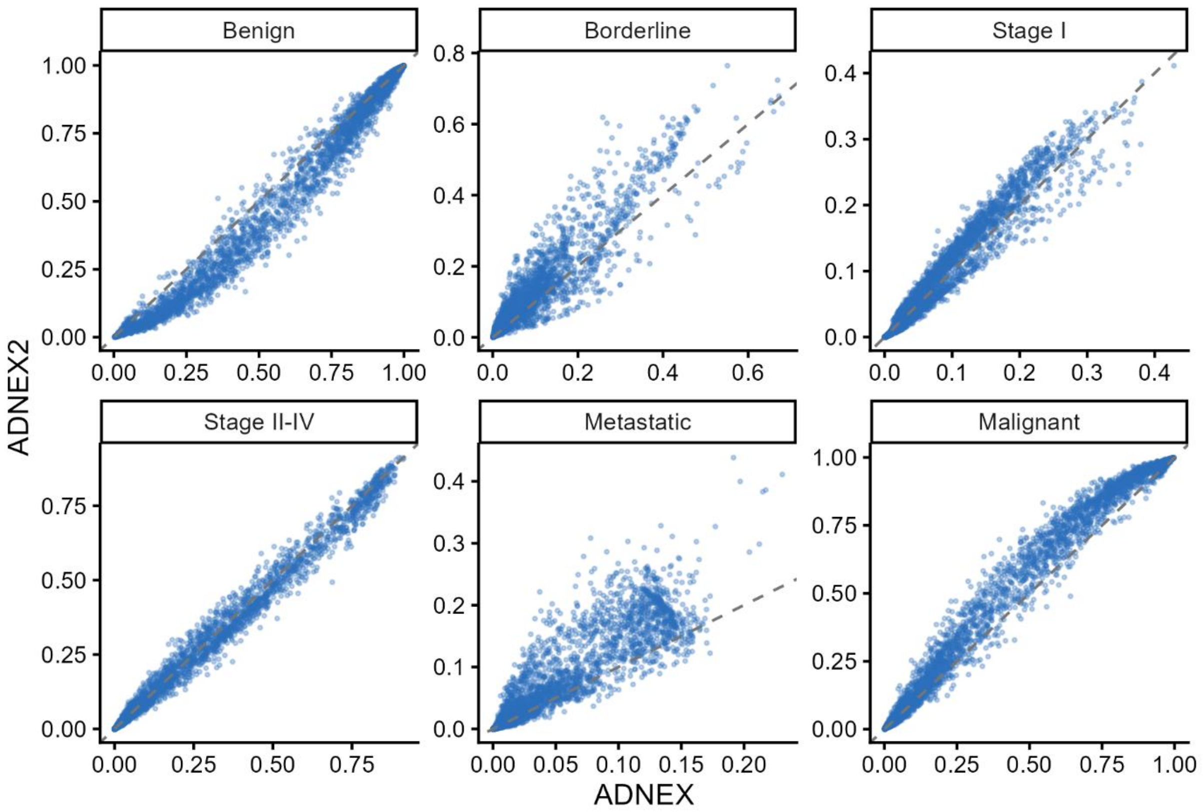

**Figure S11. Scatterplot of predicted risks of ADNEX and ADNEX2 ignoring CA125 in patients where BDs do not apply (N = 4657). Each point represents a patient. Patients' specific predictions are averaged across imputations.**

**Table S11. Performance of ADNEX, ADNEX2 and two-step strategies in different populations.**

| Model | AUC (CI) | PI | O:E (CI) | Sensitivity (CI) | Specificity (CI) | Δ NB | P-useful |
|---|---|---|---|---|---|---|---|
| **All patients (N=6847)** | | | | | | | |
| 2SS with ADNEX2 | 0.95 (0.94 to 0.96) | (0.91 to 0.97) | 1.05 (0.94-1.15) | 0.93 (0.89-0.95) | 0.84 (0.79-0.87) | 0.0594 | 97% |
| 2SS with ADNEX2 ignoring CA125 | 0.94 (0.93 to 0.95) | (0.90 to 0.97) | 1.06 (0.95-1.15) | 0.93 (0.89-0.95) | 0.83 (0.78-0.87) | 0.0596 | 98% |
| 2SS with ADNEX | 0.95 (0.94 to 0.96) | (0.92 to 0.97) | 1.14 (1.05-1.23) | 0.92 (0.87-0.95) | 0.85 (0.80-0.89) | 0.0592 | 98% |
| 2SS with ADNEX ignoring CA125 | 0.94 (0.93 to 0.95) | (0.91 to 0.97) | 1.16 (1.08-1.24) | 0.92 (0.87-0.95) | 0.84 (0.79-0.88) | 0.0594 | 97% |
| ADNEX2 | 0.95 (0.93 to 0.96) | (0.90 to 0.98) | 1.03 (0.92-1.13) | 0.92 (0.89-0.95) | 0.84 (0.79-0.87) | 0.0593 | 97% |
| ADNEX2 ignoring CA125 | 0.94 (0.93 to 0.95) | (0.90 to 0.98) | 1.04 (0.94-1.12) | 0.93 (0.89-0.95) | 0.83 (0.78-0.87) | 0.0596 | 98% |
| ADNEX | 0.94 (0.93 to 0.96) | (0.91 to 0.98) | 1.11 (1.03-1.20) | 0.92 (0.87-0.95) | 0.85 (0.80-0.89) | 0.0593 | 98% |
| ADNEX ignoring CA125 | 0.94 (0.93 to 0.95) | (0.90 to 0.97) | 1.14 (1.05-1.22) | 0.92 (0.87-0.95) | 0.84 (0.79-0.88) | 0.0594 | 97% |
| **Where benign descriptors do not apply (N = 4657)** | | | | | | | |
| ADNEX2 | 0.93 (0.91 to 0.94) | (0.88 to 0.97) | 1.04 (0.93-1.14) | 0.95 (0.92-0.97) | 0.69 (0.64-0.74) | 0.0364 | 91% |
| ADNEX2 ignoring CA125 | 0.92 (0.90 to 0.93) | (0.87 to 0.96) | 1.05 (0.96-1.14) | 0.95 (0.92-0.97) | 0.68 (0.63-0.73) | 0.0365 | 92% |
| ADNEX | 0.93 (0.91 to 0.94) | (0.88 to 0.96) | 1.13 (1.05-1.22) | 0.94 (0.90-0.96) | 0.71 (0.65-0.76) | 0.0365 | 90% |
| ADNEX ignoring CA125 | 0.92 (0.90 to 0.93) | (0.87 to 0.96) | 1.16 (1.08-1.24) | 0.94 (0.90-0.96) | 0.70 (0.64-0.75) | 0.0368 | 89% |

The 10% decision threshold was used to calculate Sensitivity, Specificity, Net Benefit and P-useful.
AUC: Area Under the receiver operating characteristic curve, CI: Confidence interval, PI: Prediction interval, OE: Observed over expected ratio, 2SS: Two step strategy, Δ NB: Delta Net Benefit calculated at the 10% decision threshold. This is the net benefit of using the model minus the maximum net benefit of treat all or treat none, P-useful: Probability of being useful in a hypothetical new centre at the 10% decision threshold. BDs: Benign descriptors

#### S4.4 Discrimination

**Table S12. Polytomous discrimination index and pairwise area under receiver operating characteristic curve of ADNEX2 and ADNEX to distinguish between outcome categories.**

| | All patients (N = 6847) | | | | Patients where BDs do not apply (N = 4657) | | | |
|---|---|---|---|---|---|---|---|---|
| | 2SS with ADNEX2 | 2SS with ADNEX2 ignoring CA125 | 2SS with ADNEX | 2SS with ADNEX ignoring CA125 | ADNEX2 | ADNEX2 ignoring CA125 | ADNEX | ADNEX ignoring CA125 |
| | **Polytomous discrimination index (95% confidence interval)** | | | | | | | |
| | 0.76 (0.74-0.78) | 0.74 (0.72-0.76) | 0.76 (0.74-0.78) | 0.74 (0.72-0.76) | 0.67 (0.65-0.69) | 0.66 (0.64-0.68) | 0.69 (0.67-0.71) | 0.66 (0.64-0.68) |
| | **Area under receiver operating characteristic curve (95% confidence interval)** | | | | | | | |
| Benign vs Borderline | 0.92 (0.90-0.93) | 0.92 (0.90-0.93) | 0.92 (0.90-0.93) | 0.91 (0.90-0.93) | 0.88 (0.86-0.90) | 0.88 (0.86-0.89) | 0.88 (0.86-0.90) | 0.87 (0.85-0.89) |
| Benign vs Stage I | 0.95 (0.93-0.96) | 0.95 (0.93-0.96) | 0.95 (0.93-0.96) | 0.95 (0.93-0.96) | 0.93 (0.91-0.94) | 0.93 (0.91-0.94) | 0.93 (0.91-0.94) | 0.93 (0.91-0.94) |
| Benign vs Stage II-IV | 0.98 (0.97-0.98) | 0.97 (0.96-0.98) | 0.98 (0.97-0.99) | 0.97 (0.96-0.98) | 0.98 (0.97-0.98) | 0.96 (0.96-0.97) | 0.98 (0.97-0.98) | 0.96 (0.96-0.97) |
| Benign vs Metastatic | 0.95 (0.93-0.97) | 0.95 (0.93-0.96) | 0.95 (0.93-0.97) | 0.95 (0.93-0.96) | 0.95 (0.93-0.96) | 0.94 (0.92-0.96) | 0.95 (0.93-0.96) | 0.94 (0.92-0.96) |
| Borderline vs Stage I | 0.77 (0.74-0.81) | 0.78 (0.74-0.81) | 0.75 (0.71-0.79) | 0.76 (0.72-0.79) | 0.78 (0.74-0.81) | 0.78 (0.74-0.81) | 0.75 (0.72-0.79) | 0.76 (0.72-0.80) |
| Borderline vs Stage II-IV | 0.92 (0.90-0.94) | 0.90 (0.88-0.92) | 0.92 (0.90-0.93) | 0.89 (0.87-0.91) | 0.92 (0.90-0.94) | 0.90 (0.88-0.92) | 0.92 (0.90-0.93) | 0.90 (0.88-0.92) |
| Borderline vs Metastatic | 0.87 (0.84-0.90) | 0.87 (0.84-0.90) | 0.86 (0.83-0.89) | 0.87 (0.83-0.89) | 0.87 (0.84-0.90) | 0.87 (0.84-0.90) | 0.87 (0.84-0.90) | 0.87 (0.84-0.90) |
| Stage I vs Stage II-IV | 0.81 (0.78-0.84) | 0.73 (0.70-0.76) | 0.81 (0.79-0.84) | 0.73 (0.70-0.76) | 0.81 (0.78-0.84) | 0.73 (0.70-0.77) | 0.82 (0.79-0.84) | 0.74 (0.70-0.77) |
| Stage I vs Metastatic | 0.72 (0.68-0.76) | 0.72 (0.68-0.76) | 0.71 (0.66-0.75) | 0.71 (0.66-0.75) | 0.73 (0.68-0.77) | 0.73 (0.68-0.77) | 0.71 (0.67-0.75) | 0.71 (0.67-0.76) |
| Stage II-IV vs Metastatic | 0.79 (0.76-0.83) | 0.65 (0.61-0.69) | 0.80 (0.77-0.83) | 0.66 (0.62-0.70) | 0.80 (0.76-0.83) | 0.65 (0.61-0.69) | 0.80 (0.77-0.83) | 0.66 (0.62-0.70) |

2SS: Two-step strategy

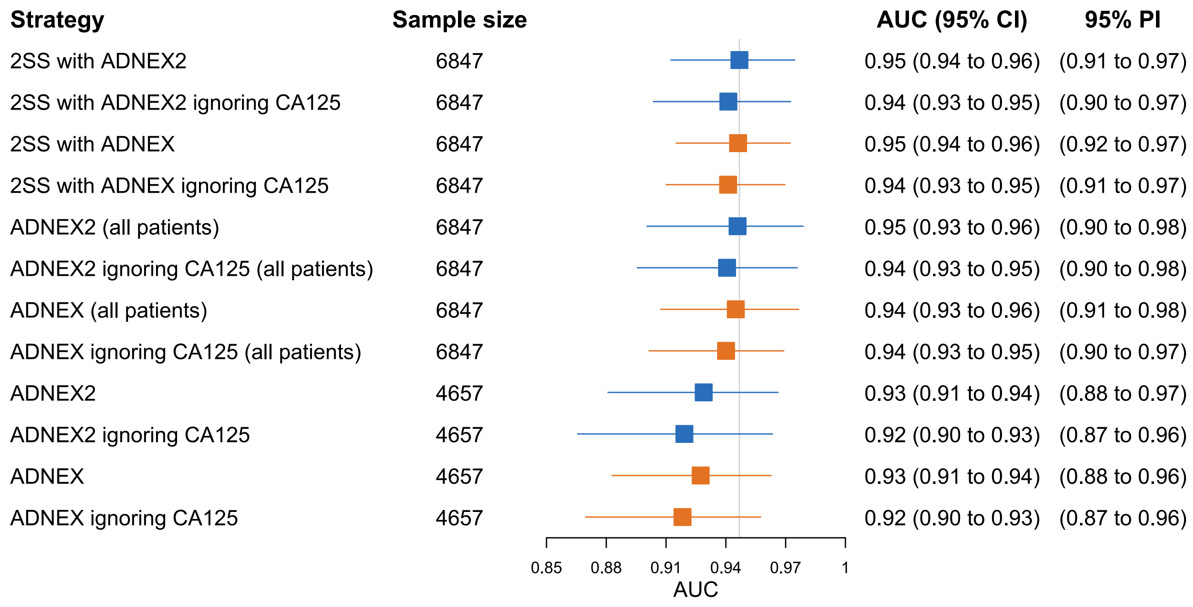

**Figure S12. Forest plot showing the meta-analysed area under the receiver operating characteristic curve (AUC) for ADNEX, ADNEX2 and the two-step strategies in the different populations. Blue points and lines correspond to ADNEX2 and orange ones to ADNEX. Horizontal lines show prediction interval. CI, 95% confidence interval. PI, 95% prediction interval. 2SS, two step strategy.**

| Centre | AUC (95% CI) | N |
|---|---|---|
| Malmö, Sweden | 0.93 (0.89 to 0.95) | 1093 |
| Rome, Italy | 0.95 (0.93 to 0.97) | 1019 |
| Leuven, Belgium | 0.96 (0.94 to 0.97) | 754 |
| Milan, Italy | 0.97 (0.95 to 0.98) | 558 |
| Stockholm, Sweden | 0.93 (0.90 to 0.95) | 509 |
| Genk, Belgium | 0.93 (0.87 to 0.96) | 473 |
| Monza, Italy | 0.94 (0.90 to 0.96) | 431 |
| Prague, Czech Rep. | 0.96 (0.93 to 0.97) | 414 |
| Trieste, Italy | 0.95 (0.89 to 0.98) | 273 |
| Cagliari, Italy | 0.97 (0.92 to 0.99) | 247 |
| Katowice, Poland | 0.94 (0.86 to 0.97) | 241 |
| Milan 2, Italy | 0.96 (0.92 to 0.98) | 227 |
| London, UK | 0.98 (0.56 to 1.00) | 172 |
| Pamplona, Spain | 0.99 (0.93 to 1.00) | 170 |
| Milan 3, Italy | 0.71 (0.34 to 0.92) | 129 |
| Florence, Italy | 0.67 (0.28 to 0.92) | 85 |
| Lisbon, Portugal | 0.98 (0.54 to 1.00) | 52 |
| **Meta-analysis** | **0.95 (0.94 to 0.96)** | **6847** |
| **AUC (95% PI)** | **(0.91 to 0.97)** | |

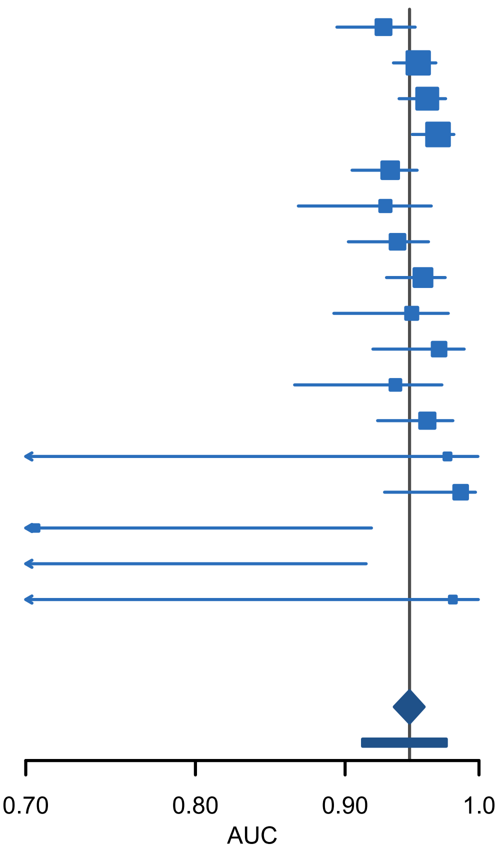

**Figure S13. Forest plot showing the area under the receiver operating characteristic curve (AUC) for two step strategy with ADNEX2 in all patients (n = 6847). CI, 95% confidence interval; PI, prediction interval**

| Centre | AUC (95% CI) | N |
|---|---|---|
| Malmö, Sweden | 0.92 (0.89 to 0.95) | 1093 |
| Rome, Italy | 0.95 (0.93 to 0.96) | 1019 |
| Leuven, Belgium | 0.96 (0.94 to 0.97) | 754 |
| Milan, Italy | 0.96 (0.94 to 0.98) | 558 |
| Stockholm, Sweden | 0.92 (0.90 to 0.95) | 509 |
| Genk, Belgium | 0.93 (0.87 to 0.96) | 473 |
| Monza, Italy | 0.92 (0.89 to 0.95) | 431 |
| Prague, Czech Rep. | 0.95 (0.93 to 0.97) | 414 |
| Trieste, Italy | 0.94 (0.89 to 0.97) | 273 |
| Cagliari, Italy | 0.96 (0.91 to 0.98) | 247 |
| Katowice, Poland | 0.93 (0.86 to 0.97) | 241 |
| Milan 2, Italy | 0.95 (0.92 to 0.98) | 227 |
| London, UK | 0.96 (0.65 to 1.00) | 172 |
| Pamplona, Spain | 0.98 (0.93 to 1.00) | 170 |
| Milan 3, Italy | 0.70 (0.33 to 0.91) | 129 |
| Florence, Italy | 0.67 (0.28 to 0.91) | 85 |
| Lisbon, Portugal | 0.97 (0.60 to 1.00) | 52 |
| **Meta-analysis** | **0.94 (0.93 to 0.95)** | **6847** |
| **AUC (95% PI)** | **(0.9 to 0.97)** | |

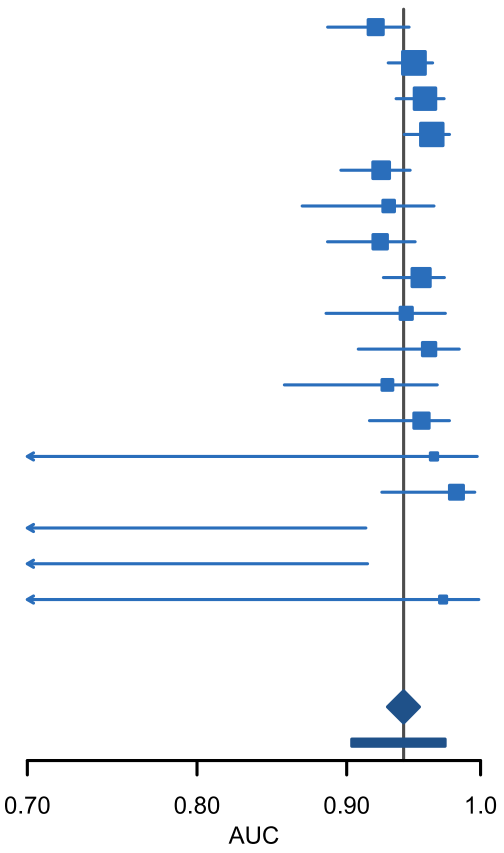

**Figure S14. Forest plot showing the area under the receiver operating characteristic curve (AUC) for two step strategy with ADNEX2 ignoring CA125 in all patients (n = 6847). CI, 95% confidence interval; PI, prediction interval**

| Centre | AUC (95% CI) | N |
|---|---|---|
| Malmö, Sweden | 0.93 (0.89 to 0.95) | 1093 |
| Rome, Italy | 0.95 (0.93 to 0.97) | 1019 |
| Leuven, Belgium | 0.96 (0.94 to 0.97) | 754 |
| Milan, Italy | 0.97 (0.95 to 0.98) | 558 |
| Stockholm, Sweden | 0.93 (0.90 to 0.95) | 509 |
| Genk, Belgium | 0.93 (0.87 to 0.96) | 473 |
| Monza, Italy | 0.94 (0.91 to 0.96) | 431 |
| Prague, Czech Rep. | 0.96 (0.93 to 0.97) | 414 |
| Trieste, Italy | 0.95 (0.89 to 0.98) | 273 |
| Cagliari, Italy | 0.97 (0.92 to 0.99) | 247 |
| Katowice, Poland | 0.94 (0.86 to 0.97) | 241 |
| Milan 2, Italy | 0.96 (0.92 to 0.98) | 227 |
| London, UK | 0.97 (0.59 to 1.00) | 172 |
| Pamplona, Spain | 0.99 (0.93 to 1.00) | 170 |
| Milan 3, Italy | 0.71 (0.34 to 0.92) | 129 |
| Florence, Italy | 0.67 (0.28 to 0.92) | 85 |
| Lisbon, Portugal | 0.98 (0.54 to 1.00) | 52 |
| **Meta-analysis** | **0.95 (0.94 to 0.96)** | **6847** |
| **AUC (95% PI)** | **(0.92 to 0.97)** | |

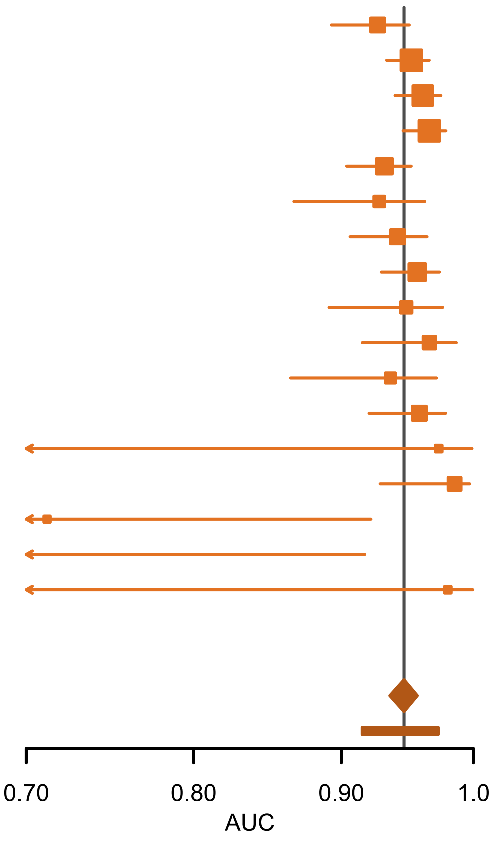

**Figure S15. Forest plot showing the area under the receiver operating characteristic curve (AUC) for two step strategy with ADNEX in all patients (n = 6847). CI, 95% confidence interval; PI, prediction interval**

| Centre | AUC (95% CI) | N |
|---|---|---|
| Malmö, Sweden | 0.92 (0.89 to 0.94) | 1093 |
| Rome, Italy | 0.95 (0.93 to 0.96) | 1019 |
| Leuven, Belgium | 0.96 (0.93 to 0.97) | 754 |
| Milan, Italy | 0.96 (0.94 to 0.97) | 558 |
| Stockholm, Sweden | 0.93 (0.90 to 0.95) | 509 |
| Genk, Belgium | 0.93 (0.87 to 0.96) | 473 |
| Monza, Italy | 0.93 (0.89 to 0.95) | 431 |
| Prague, Czech Rep. | 0.95 (0.93 to 0.97) | 414 |
| Trieste, Italy | 0.94 (0.89 to 0.97) | 273 |
| Cagliari, Italy | 0.96 (0.90 to 0.98) | 247 |
| Katowice, Poland | 0.93 (0.86 to 0.97) | 241 |
| Milan 2, Italy | 0.95 (0.91 to 0.97) | 227 |
| London, UK | 0.96 (0.65 to 1.00) | 172 |
| Pamplona, Spain | 0.98 (0.93 to 1.00) | 170 |
| Milan 3, Italy | 0.71 (0.34 to 0.92) | 129 |
| Florence, Italy | 0.67 (0.28 to 0.92) | 85 |
| Lisbon, Portugal | 0.98 (0.53 to 1.00) | 52 |
| **Meta-analysis** | **0.94 (0.93 to 0.95)** | **6847** |
| **AUC (95% PI)** | **(0.91 to 0.97)** | |

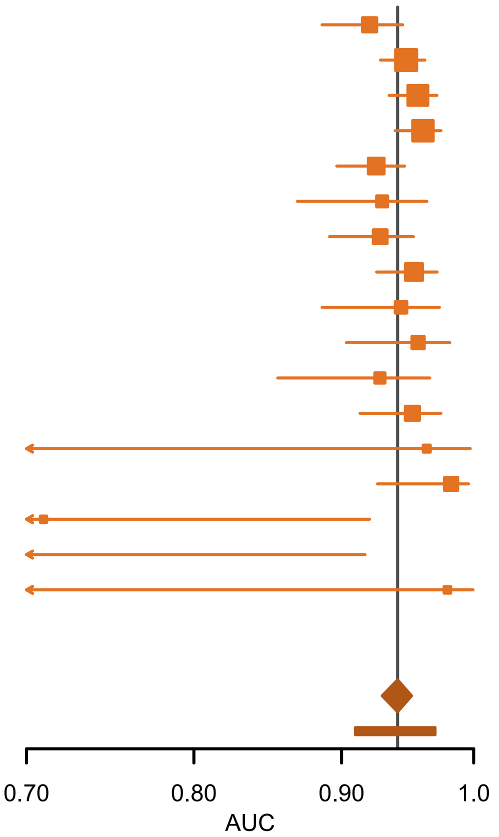

**Figure S16. Forest plot showing the area under the receiver operating characteristic curve (AUC) for two step strategy with ADNEX ignoring CA125 in all patients (n = 6847). CI, 95% confidence interval; PI, prediction interval**

| Centre | AUC (95% CI) | N |
|---|---|---|
| Malmö, Sweden | 0.92 (0.89 to 0.95) | 1093 |
| Rome, Italy | 0.96 (0.94 to 0.97) | 1019 |
| Leuven, Belgium | 0.96 (0.94 to 0.97) | 754 |
| Milan, Italy | 0.97 (0.95 to 0.98) | 558 |
| Stockholm, Sweden | 0.93 (0.90 to 0.95) | 509 |
| Genk, Belgium | 0.93 (0.86 to 0.96) | 473 |
| Monza, Italy | 0.94 (0.90 to 0.96) | 431 |
| Prague, Czech Rep. | 0.96 (0.93 to 0.97) | 414 |
| Trieste, Italy | 0.95 (0.90 to 0.98) | 273 |
| Cagliari, Italy | 0.97 (0.92 to 0.99) | 247 |
| Katowice, Poland | 0.94 (0.87 to 0.97) | 241 |
| Milan 2, Italy | 0.96 (0.92 to 0.98) | 227 |
| London, UK | 0.98 (0.63 to 1.00) | 172 |
| Pamplona, Spain | 0.99 (0.93 to 1.00) | 170 |
| Milan 3, Italy | 0.62 (0.29 to 0.87) | 129 |
| Florence, Italy | 0.74 (0.34 to 0.94) | 85 |
| Lisbon, Portugal | 0.98 (0.56 to 1.00) | 52 |
| **Meta-analysis** | **0.95 (0.93 to 0.96)** | **6847** |
| **AUC (95% PI)** | **(0.9 to 0.98)** | |

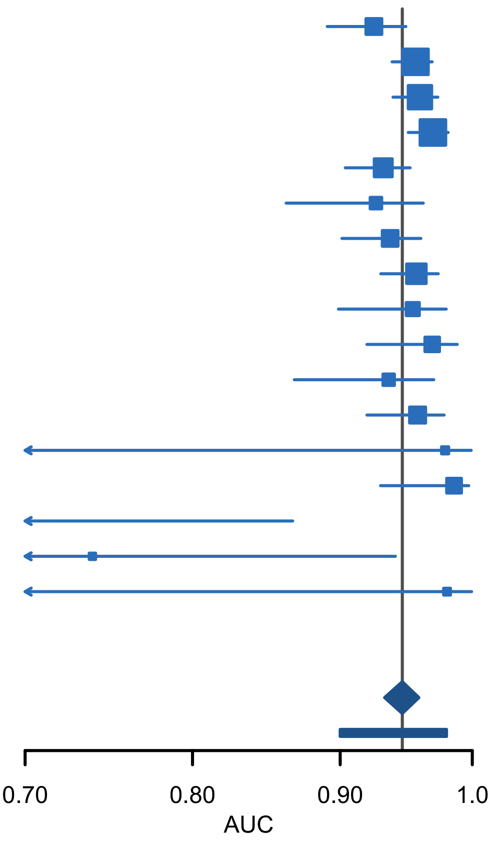

**Figure S17. Forest plot showing the area under the receiver operating characteristic curve (AUC) for ADNEX2 in all patients (n = 6847). CI, 95% confidence interval; PI, prediction interval**

| Centre | AUC (95% CI) | N |
|---|---|---|
| Malmö, Sweden | 0.92 (0.89 to 0.95) | 1093 |
| Rome, Italy | 0.95 (0.93 to 0.96) | 1019 |
| Leuven, Belgium | 0.96 (0.93 to 0.97) | 754 |
| Milan, Italy | 0.96 (0.94 to 0.98) | 558 |
| Stockholm, Sweden | 0.92 (0.90 to 0.95) | 509 |
| Genk, Belgium | 0.93 (0.86 to 0.96) | 473 |
| Monza, Italy | 0.92 (0.88 to 0.95) | 431 |
| Prague, Czech Rep. | 0.95 (0.93 to 0.97) | 414 |
| Trieste, Italy | 0.95 (0.89 to 0.98) | 273 |
| Cagliari, Italy | 0.96 (0.91 to 0.98) | 247 |
| Katowice, Poland | 0.92 (0.85 to 0.96) | 241 |
| Milan 2, Italy | 0.95 (0.91 to 0.97) | 227 |
| London, UK | 0.97 (0.70 to 1.00) | 172 |
| Pamplona, Spain | 0.98 (0.93 to 1.00) | 170 |
| Milan 3, Italy | 0.61 (0.28 to 0.86) | 129 |
| Florence, Italy | 0.74 (0.34 to 0.94) | 85 |
| Lisbon, Portugal | 0.97 (0.61 to 1.00) | 52 |
| **Meta-analysis** | **0.94 (0.93 to 0.95)** | **6847** |
| **AUC (95% PI)** | **(0.89 to 0.98)** | |

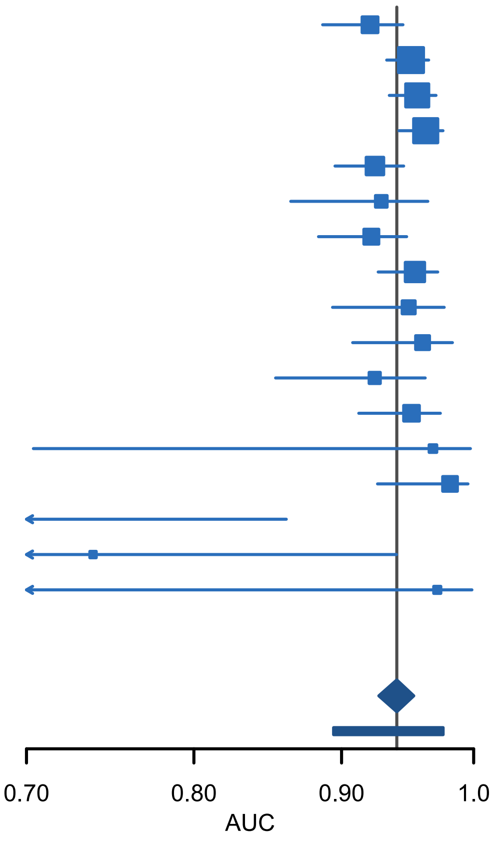

**Figure S18. Forest plot showing the area under the receiver operating characteristic curve (AUC) for ADNEX2 ignoring CA125 in all patients (n = 6847). CI, 95% confidence interval; PI, prediction interval**

| Centre | AUC (95% CI) | N |
|---|---|---|
| Malmö, Sweden | 0.92 (0.89 to 0.95) | 1093 |
| Rome, Italy | 0.95 (0.94 to 0.97) | 1019 |
| Leuven, Belgium | 0.96 (0.94 to 0.97) | 754 |
| Milan, Italy | 0.97 (0.95 to 0.98) | 558 |
| Stockholm, Sweden | 0.93 (0.90 to 0.95) | 509 |
| Genk, Belgium | 0.93 (0.87 to 0.96) | 473 |
| Monza, Italy | 0.94 (0.91 to 0.96) | 431 |
| Prague, Czech Rep. | 0.96 (0.93 to 0.97) | 414 |
| Trieste, Italy | 0.95 (0.90 to 0.98) | 273 |
| Cagliari, Italy | 0.96 (0.91 to 0.99) | 247 |
| Katowice, Poland | 0.93 (0.86 to 0.97) | 241 |
| Milan 2, Italy | 0.95 (0.92 to 0.98) | 227 |
| London, UK | 0.97 (0.63 to 1.00) | 172 |
| Pamplona, Spain | 0.99 (0.93 to 1.00) | 170 |
| Milan 3, Italy | 0.62 (0.28 to 0.87) | 129 |
| Florence, Italy | 0.74 (0.33 to 0.94) | 85 |
| Lisbon, Portugal | 0.98 (0.56 to 1.00) | 52 |
| **Meta-analysis** | **0.95 (0.93 to 0.96)** | **6847** |
| **AUC (95% PI)** | **(0.91 to 0.97)** | |

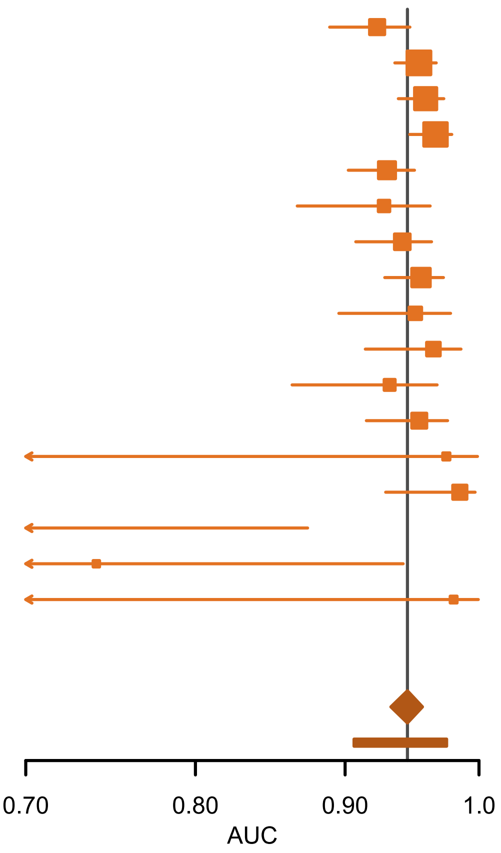

**Figure S19. Forest plot showing the area under the receiver operating characteristic curve (AUC) for ADNEX in all patients (n = 6847). CI, 95% confidence interval; PI, prediction interval**

| Centre | AUC (95% CI) | N |
|---|---|---|
| Malmö, Sweden | 0.92 (0.89 to 0.94) | 1093 |
| Rome, Italy | 0.95 (0.93 to 0.96) | 1019 |
| Leuven, Belgium | 0.95 (0.93 to 0.97) | 754 |
| Milan, Italy | 0.96 (0.94 to 0.97) | 558 |
| Stockholm, Sweden | 0.92 (0.90 to 0.95) | 509 |
| Genk, Belgium | 0.93 (0.87 to 0.97) | 473 |
| Monza, Italy | 0.93 (0.89 to 0.95) | 431 |
| Prague, Czech Rep. | 0.95 (0.93 to 0.97) | 414 |
| Trieste, Italy | 0.95 (0.89 to 0.98) | 273 |
| Cagliari, Italy | 0.96 (0.90 to 0.98) | 247 |
| Katowice, Poland | 0.92 (0.85 to 0.96) | 241 |
| Milan 2, Italy | 0.95 (0.91 to 0.97) | 227 |
| London, UK | 0.97 (0.68 to 1.00) | 172 |
| Pamplona, Spain | 0.98 (0.93 to 1.00) | 170 |
| Milan 3, Italy | 0.62 (0.28 to 0.87) | 129 |
| Florence, Italy | 0.74 (0.33 to 0.94) | 85 |
| Lisbon, Portugal | 0.98 (0.54 to 1.00) | 52 |
| **Meta-analysis** | **0.94 (0.93 to 0.95)** | **6847** |
| **AUC (95% PI)** | **(0.9 to 0.97)** | |

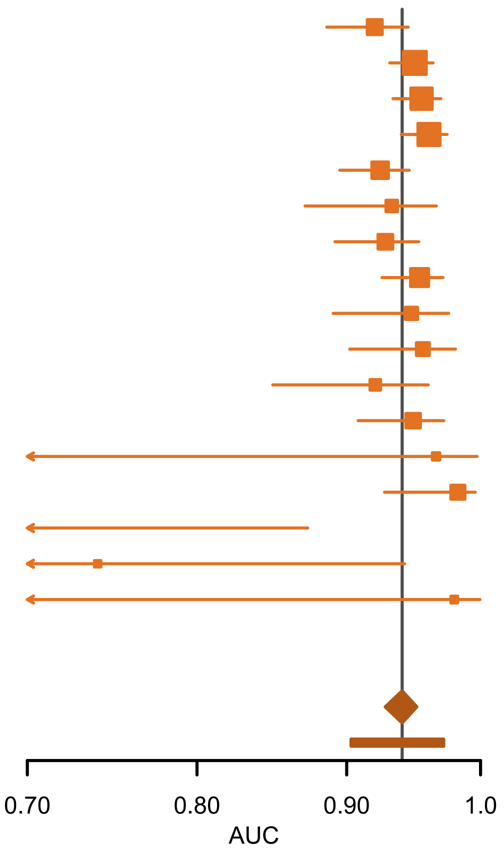

**Figure S20. Forest plot showing the area under the receiver operating characteristic curve (AUC) for ADNEX ignoring CA125 in all patients (n = 6847). CI, 95% confidence interval; PI, prediction interval**

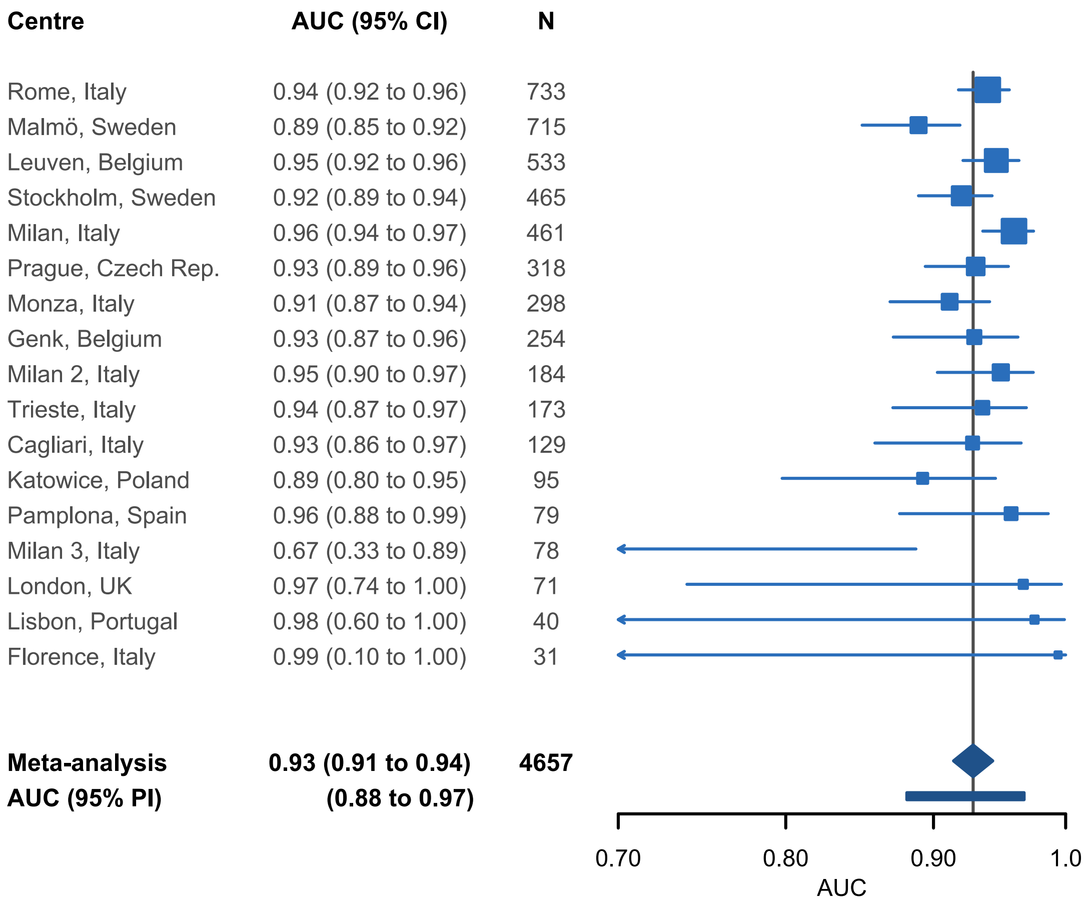

**Figure S21. Forest plot showing the area under the receiver operating characteristic curve (AUC) for ADNEX2 where benign descriptors do not apply (n = 4657). CI, 95% confidence interval; PI, prediction interval**

| Centre | AUC (95% CI) | N |
|---|---|---|
| Rome, Italy | 0.93 (0.91 to 0.95) | 733 |
| Malmö, Sweden | 0.88 (0.84 to 0.91) | 715 |
| Leuven, Belgium | 0.94 (0.92 to 0.96) | 533 |
| Stockholm, Sweden | 0.91 (0.88 to 0.94) | 465 |
| Milan, Italy | 0.95 (0.93 to 0.97) | 461 |
| Prague, Czech Rep. | 0.93 (0.89 to 0.95) | 318 |
| Monza, Italy | 0.89 (0.84 to 0.92) | 298 |
| Genk, Belgium | 0.93 (0.87 to 0.96) | 254 |
| Milan 2, Italy | 0.94 (0.89 to 0.97) | 184 |
| Trieste, Italy | 0.93 (0.86 to 0.96) | 173 |
| Cagliari, Italy | 0.91 (0.83 to 0.95) | 129 |
| Katowice, Poland | 0.87 (0.77 to 0.93) | 95 |
| Pamplona, Spain | 0.94 (0.86 to 0.98) | 79 |
| Milan 3, Italy | 0.65 (0.32 to 0.88) | 78 |
| London, UK | 0.94 (0.74 to 0.99) | 71 |
| Lisbon, Portugal | 0.96 (0.64 to 1.00) | 40 |
| Florence, Italy | 0.99 (0.10 to 1.00) | 31 |
| **Meta-analysis** | **0.92 (0.90 to 0.93)** | **4657** |
| **AUC (95% PI)** | **(0.87 to 0.96)** | |

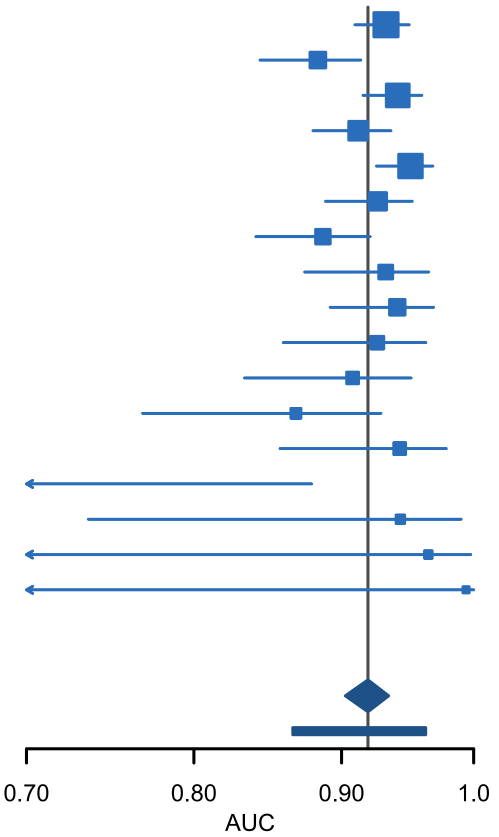

**Figure S22. Forest plot showing the area under the receiver operating characteristic curve (AUC) for ADNEX2 ignoring CA125 where benign descriptors do not apply (n = 4657). CI, 95% confidence interval; PI, prediction interval**

| Centre | AUC (95% CI) | N |
|---|---|---|
| Rome, Italy | 0.94 (0.92 to 0.95) | 733 |
| Malmö, Sweden | 0.89 (0.85 to 0.92) | 715 |
| Leuven, Belgium | 0.95 (0.92 to 0.96) | 533 |
| Stockholm, Sweden | 0.92 (0.89 to 0.94) | 465 |
| Milan, Italy | 0.96 (0.93 to 0.97) | 461 |
| Prague, Czech Rep. | 0.93 (0.89 to 0.95) | 318 |
| Monza, Italy | 0.92 (0.88 to 0.95) | 298 |
| Genk, Belgium | 0.93 (0.87 to 0.96) | 254 |
| Milan 2, Italy | 0.95 (0.90 to 0.97) | 184 |
| Trieste, Italy | 0.93 (0.87 to 0.97) | 173 |
| Cagliari, Italy | 0.92 (0.85 to 0.96) | 129 |
| Katowice, Poland | 0.89 (0.80 to 0.94) | 95 |
| Pamplona, Spain | 0.96 (0.87 to 0.99) | 79 |
| Milan 3, Italy | 0.67 (0.34 to 0.89) | 78 |
| London, UK | 0.96 (0.74 to 1.00) | 71 |
| Lisbon, Portugal | 0.98 (0.60 to 1.00) | 40 |
| Florence, Italy | 0.99 (0.12 to 1.00) | 31 |
| **Meta-analysis** | **0.93 (0.91 to 0.94)** | **4657** |
| **AUC (95% PI)** | **(0.89 to 0.96)** | |

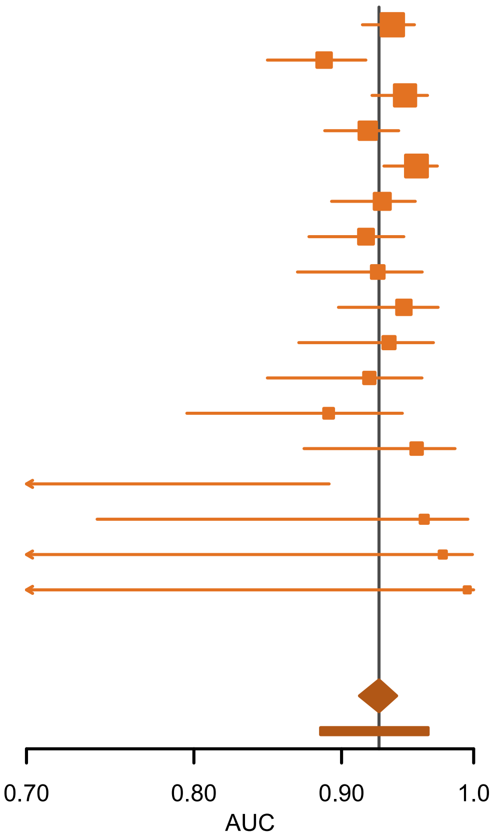

**Figure S23. Forest plot showing the area under the receiver operating characteristic curve (AUC) for ADNEX where benign descriptors do not apply (n = 4657). CI, 95%confidence interval; PI, prediction interval**

| Centre | AUC (95% CI) | N |
|---|---|---|
| Rome, Italy | 0.93 (0.91 to 0.95) | 733 |
| Malmö, Sweden | 0.88 (0.84 to 0.91) | 715 |
| Leuven, Belgium | 0.94 (0.91 to 0.96) | 533 |
| Stockholm, Sweden | 0.91 (0.88 to 0.94) | 465 |
| Milan, Italy | 0.95 (0.92 to 0.97) | 461 |
| Prague, Czech Rep. | 0.93 (0.89 to 0.95) | 318 |
| Monza, Italy | 0.89 (0.85 to 0.93) | 298 |
| Genk, Belgium | 0.93 (0.87 to 0.96) | 254 |
| Milan 2, Italy | 0.94 (0.89 to 0.97) | 184 |
| Trieste, Italy | 0.93 (0.86 to 0.96) | 173 |
| Cagliari, Italy | 0.90 (0.82 to 0.95) | 129 |
| Katowice, Poland | 0.86 (0.76 to 0.93) | 95 |
| Pamplona, Spain | 0.95 (0.86 to 0.98) | 79 |
| Milan 3, Italy | 0.67 (0.34 to 0.89) | 78 |
| London, UK | 0.94 (0.73 to 0.99) | 71 |
| Lisbon, Portugal | 0.98 (0.59 to 1.00) | 40 |
| Florence, Italy | 0.99 (0.12 to 1.00) | 31 |
| **Meta-analysis** | **0.92 (0.90 to 0.93)** | **4657** |
| **AUC (95% PI)** | **(0.87 to 0.96)** | |

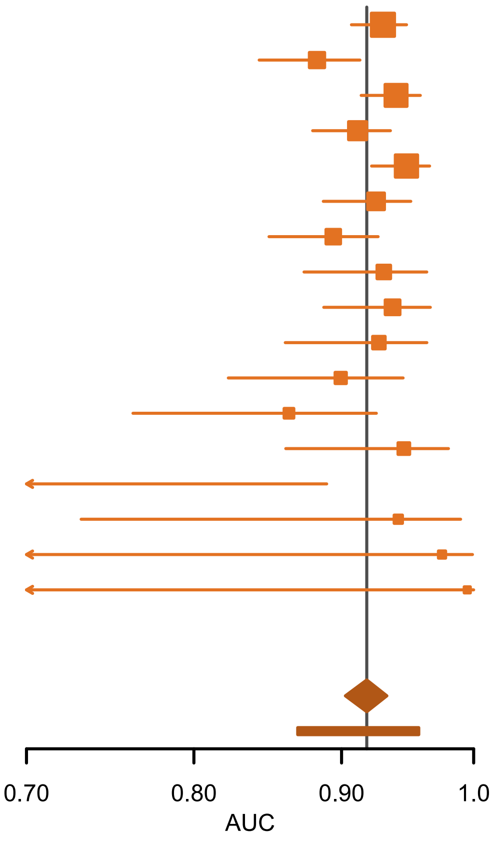

**Figure S24. Forest plot showing the area under the receiver operating characteristic curve (AUC) for ADNEX ignoring CA125 where benign descriptors do not apply (n = 4657). CI, 95% confidence interval; PI, prediction interval**

#### S4.5 CALIBRATION

**Table S13. Calibration metrics for ADNEX and ADNEX2 when used alone or in a two-step strategy**

| Model | O:E (CI;PI) |
|---|---|
| **All patients (n = 6847)** | |
| 2SS with ADNEX2 | 1.05 (0.94-1.15;0.70-1.44) |
| 2SS with ADNEX2 ignoring CA125 | 1.06 (0.96-1.15;0.74-1.40) |
| 2SS with ADNEX | 1.14 (1.06-1.23;0.87-1.45) |
| 2SS with ADNEX ignoring CA125 | 1.16 (1.08-1.24;0.89-1.45) |
| ADNEX2 | 1.03 (0.92-1.13;0.68-1.43) |
| ADNEX2 ignoring CA125 | 1.04 (0.94-1.12;0.71-1.37) |
| ADNEX | 1.11 (1.03-1.20;0.83-1.42) |
| ADNEX ignoring CA125 | 1.14 (1.05-1.22;0.87-1.44) |
| **BDs do not apply (n = 4657)** | |
| ADNEX2 | 1.04 (0.93-1.14;0.70-1.46) |
| ADNEX2 ignoring CA125 | 1.05 (0.95-1.14;0.73-1.40) |
| ADNEX | 1.13 (1.05-1.22;0.86-1.45) |
| ADNEX ignoring CA125 | 1.16 (1.08-1.24;0.89-1.45) |

OE: Observed to expected ratio; CI: Confidence interval; PI: prediction interval.

##### S4.5.1 Overall and centre specific multicentre calibration curves with 95% prediction interval

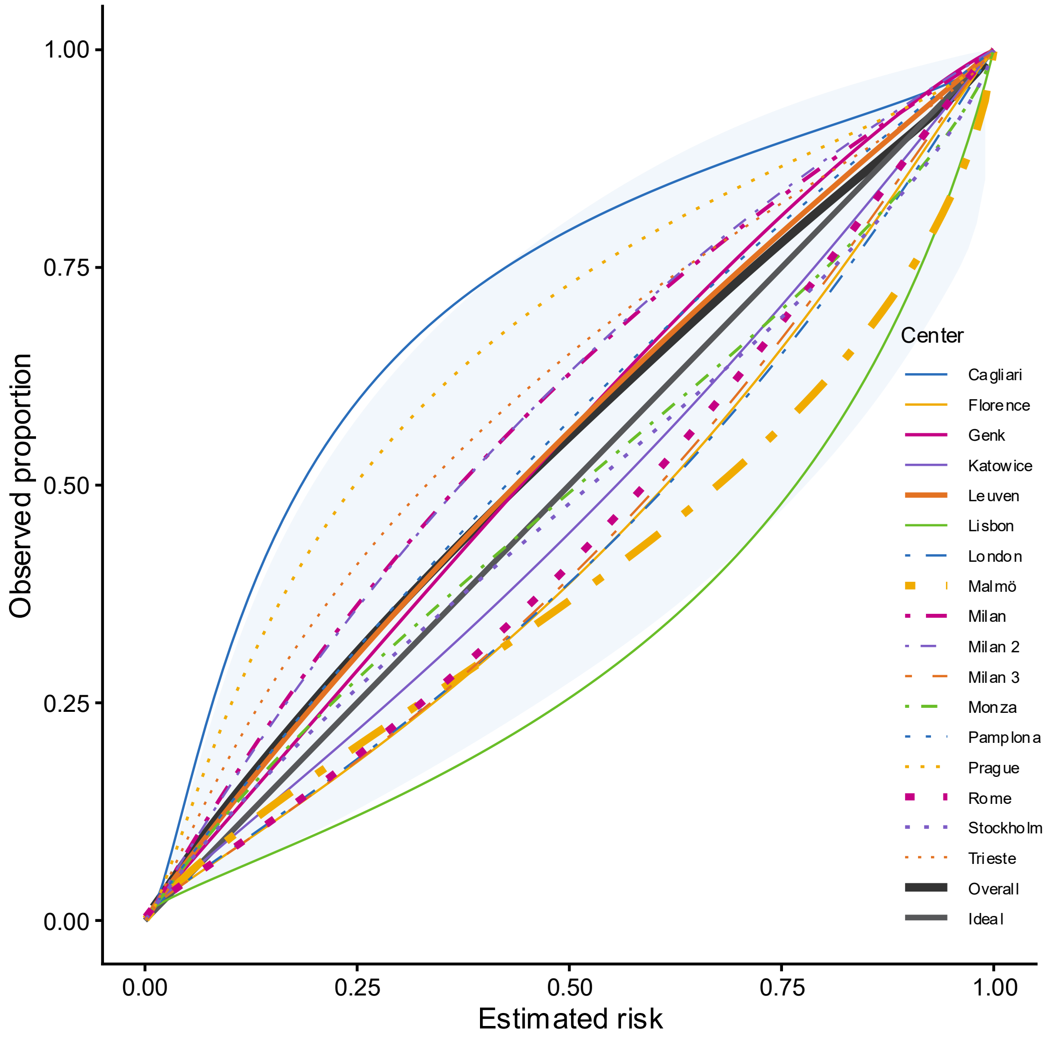

**Figure S25. Calibration plot for two step strategy with ADNEX2 in all patients (n = 6847). Overall calibration curves calculated taking clustering into account. Shaded area indicates 95% prediction interval. Linewidth is proportional with sample size. The thick black straight line is the ideal line denoting perfect agreement between predicted risk and observed proportion.**

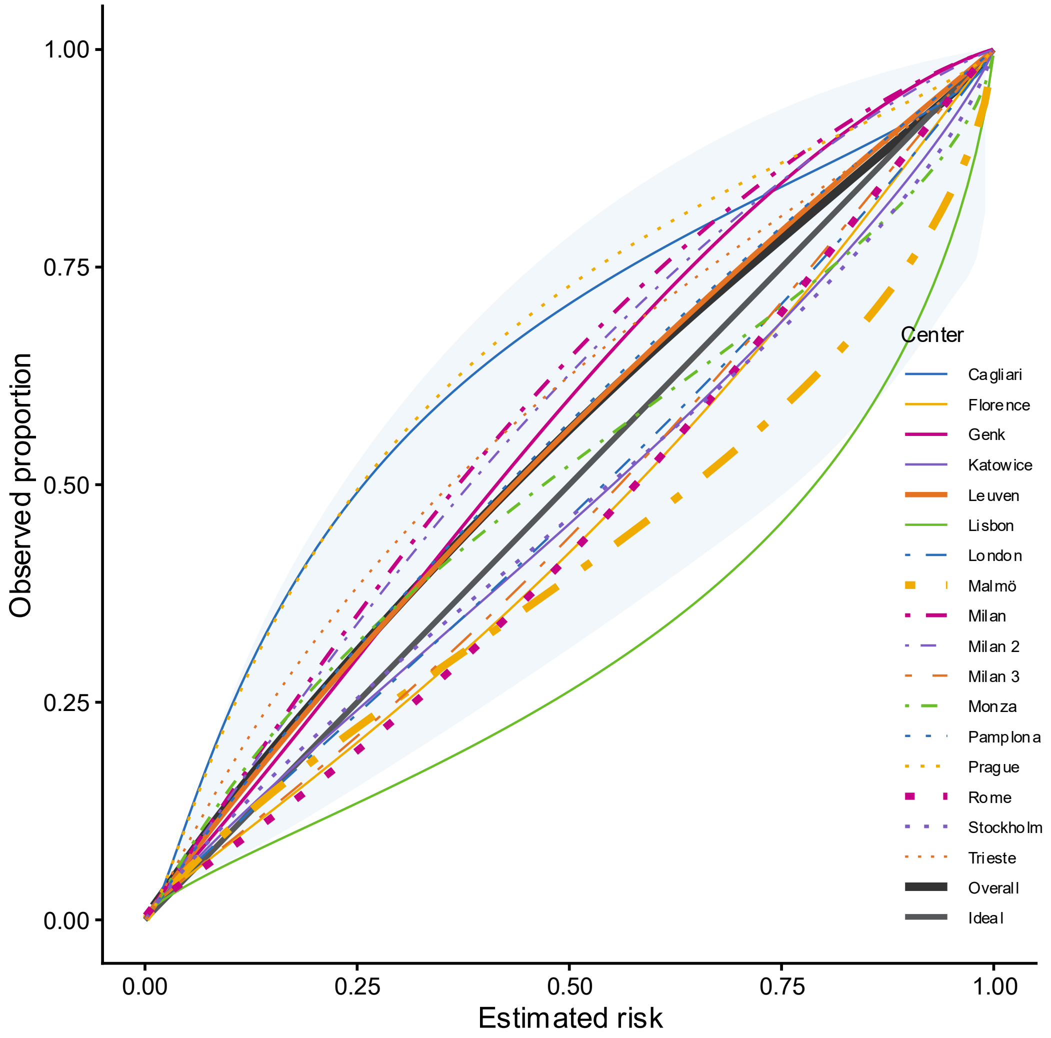

**Figure S26. Calibration plot for two step strategy with ADNEX2 ignoring CA125 in all patients (n = 6847). Overall calibration curves calculated taking clustering into account. Shaded area indicates 95% prediction interval. Linewidth is proportional with sample size. The thick black straight line is the ideal line denoting perfect agreement between predicted risk and observed proportion.**

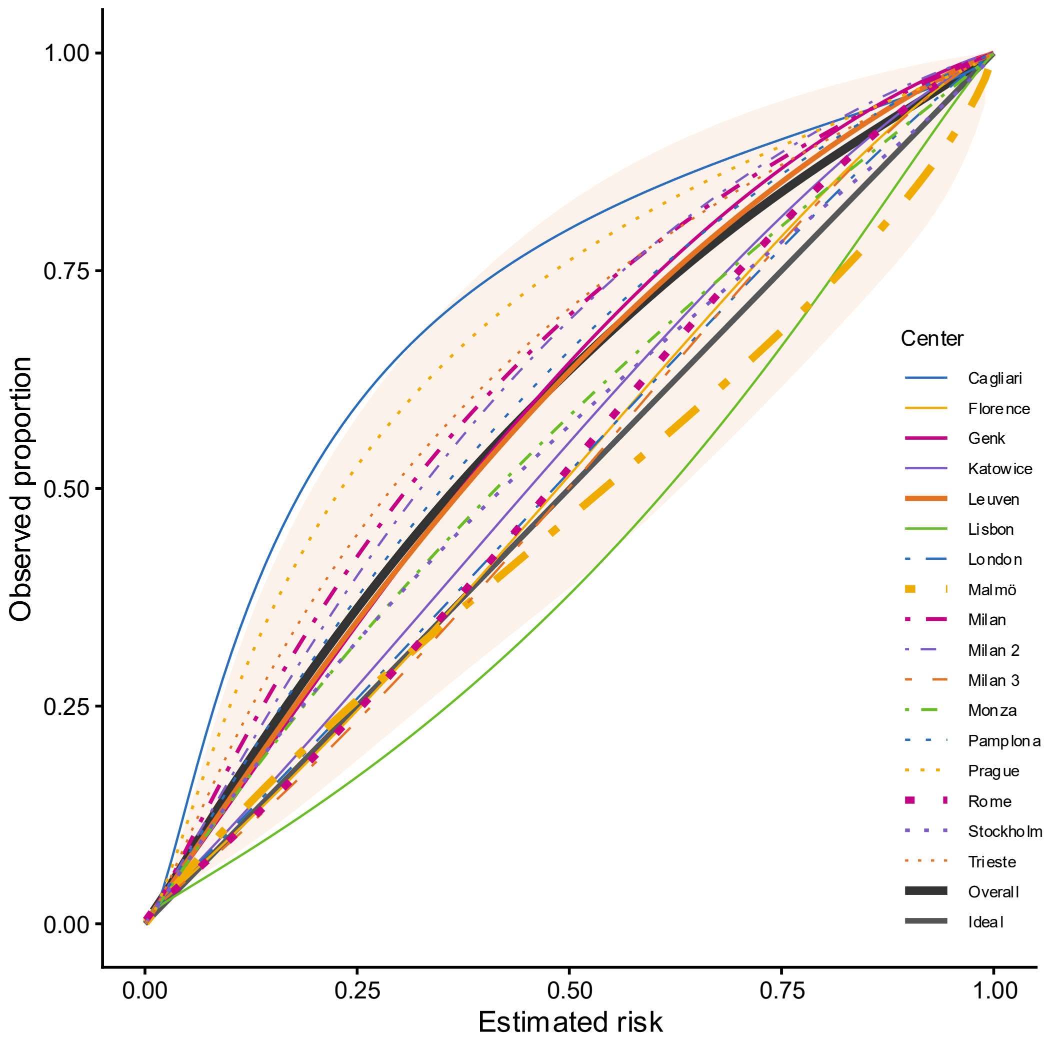

**Figure S27. Calibration plot for two step strategy with ADNEX in all patients (n = 6847). Overall calibration curves calculated taking clustering into account. Shaded area indicates 95% prediction interval. Linewidth is proportional with sample size. The thick black straight line is the ideal line denoting perfect agreement between predicted risk and observed proportion.**

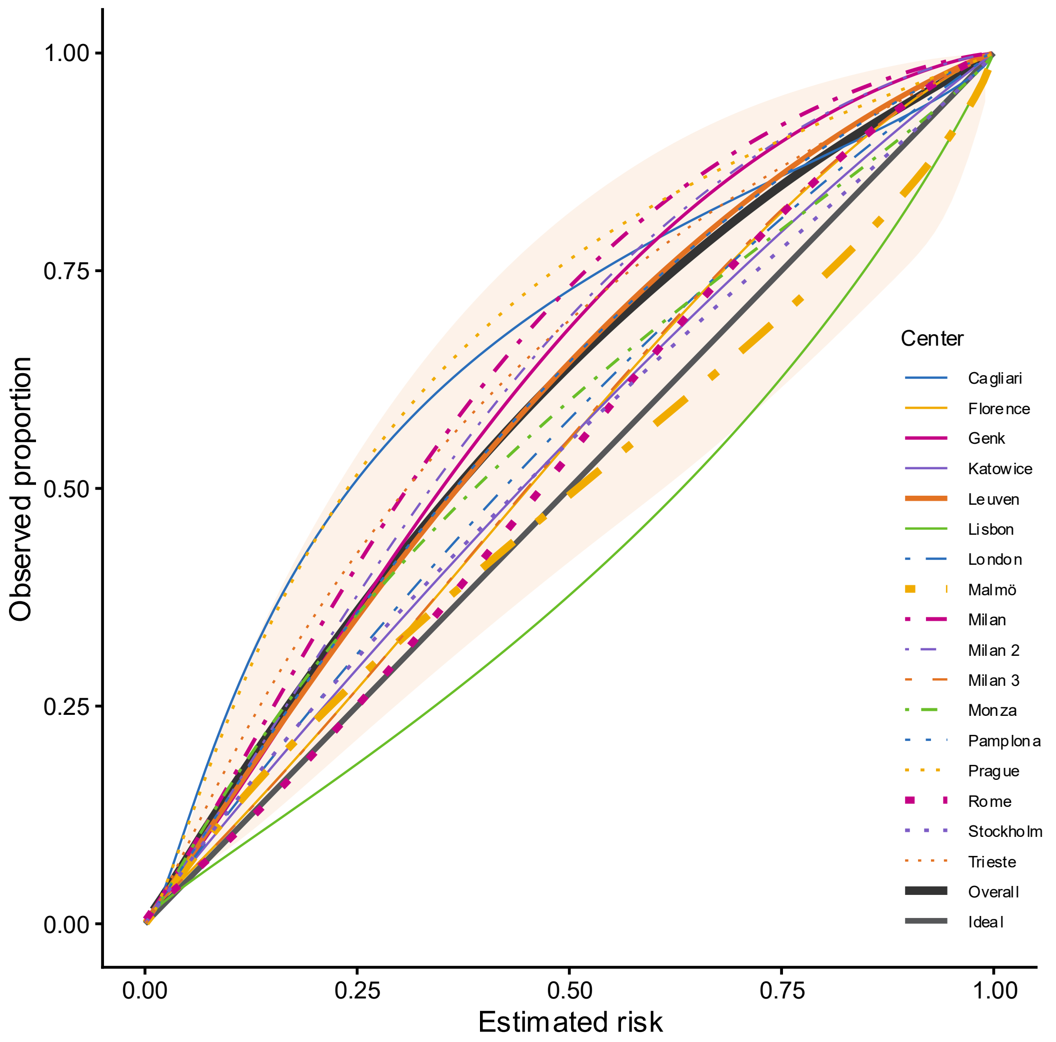

**Figure S28. Calibration plot for two step strategy with ADNEX ignoring CA125 in all patients (n = 6847). Overall calibration curves calculated taking clustering into account. Shaded area indicates 95% prediction interval. Linewidth is proportional with sample size. The thick black straight line is the ideal line denoting perfect agreement between predicted risk and observed proportion.**

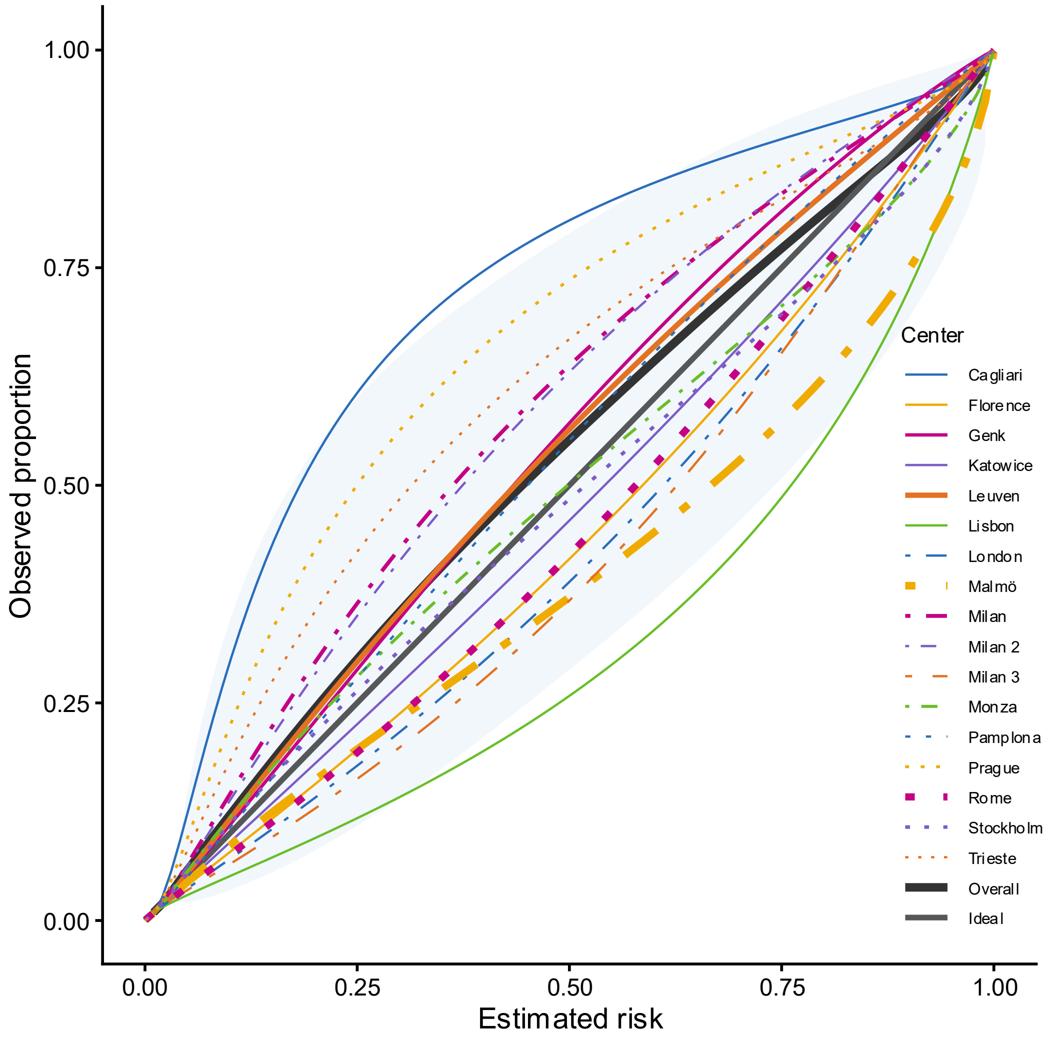

**Figure S29. Calibration plot for ADNEX2 in all patients (n = 6847). Overall calibration curves calculated taking clustering into account. Shaded area indicates 95% prediction interval. Linewidth is proportional with sample size. The thick black straight line is the ideal line denoting perfect agreement between predicted risk and observed proportion.**

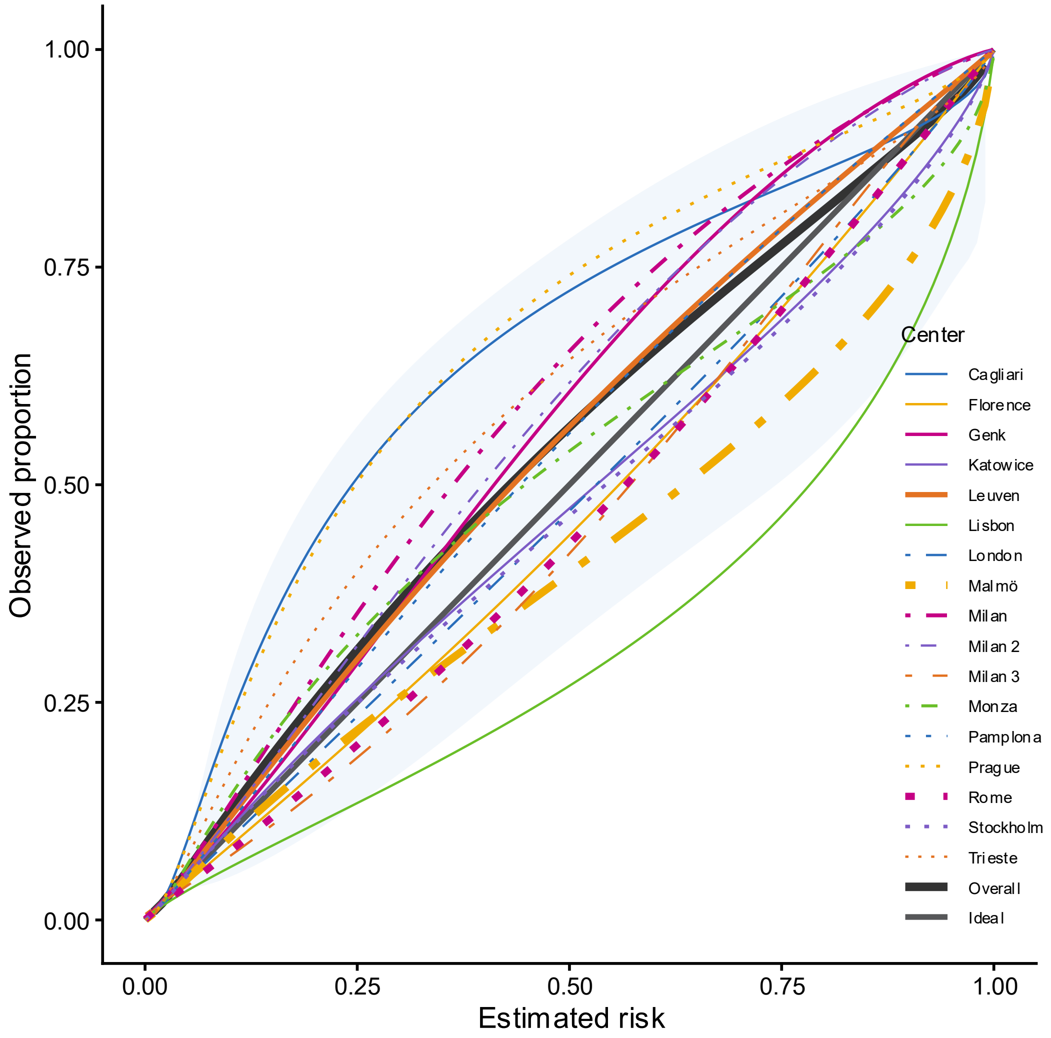

**Figure S30. Calibration plot for ADNEX2 ignoring CA125 in all patients (n = 6847). Overall calibration curves calculated taking clustering into account. Shaded area indicates 95% prediction interval. Linewidth is proportional with sample size. The thick black straight line is the ideal line denoting perfect agreement between predicted risk and observed proportion.**

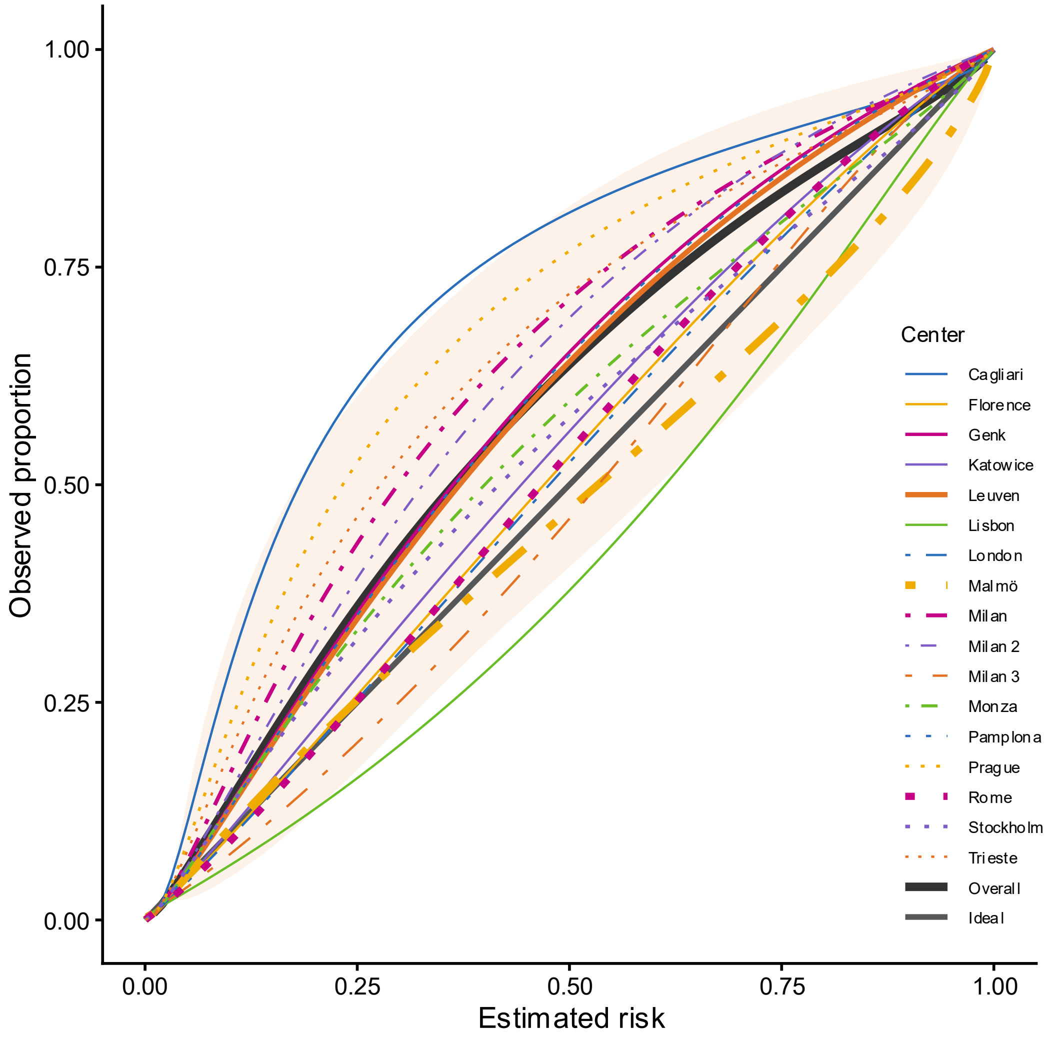

**Figure S31. Calibration plot for ADNEX in all patients (n = 6847). Overall calibration curves calculated taking clustering into account. Shaded area indicates 95% prediction interval. Linewidth is proportional with sample size. The thick black straight line is the ideal line denoting perfect agreement between predicted risk and observed proportion.**

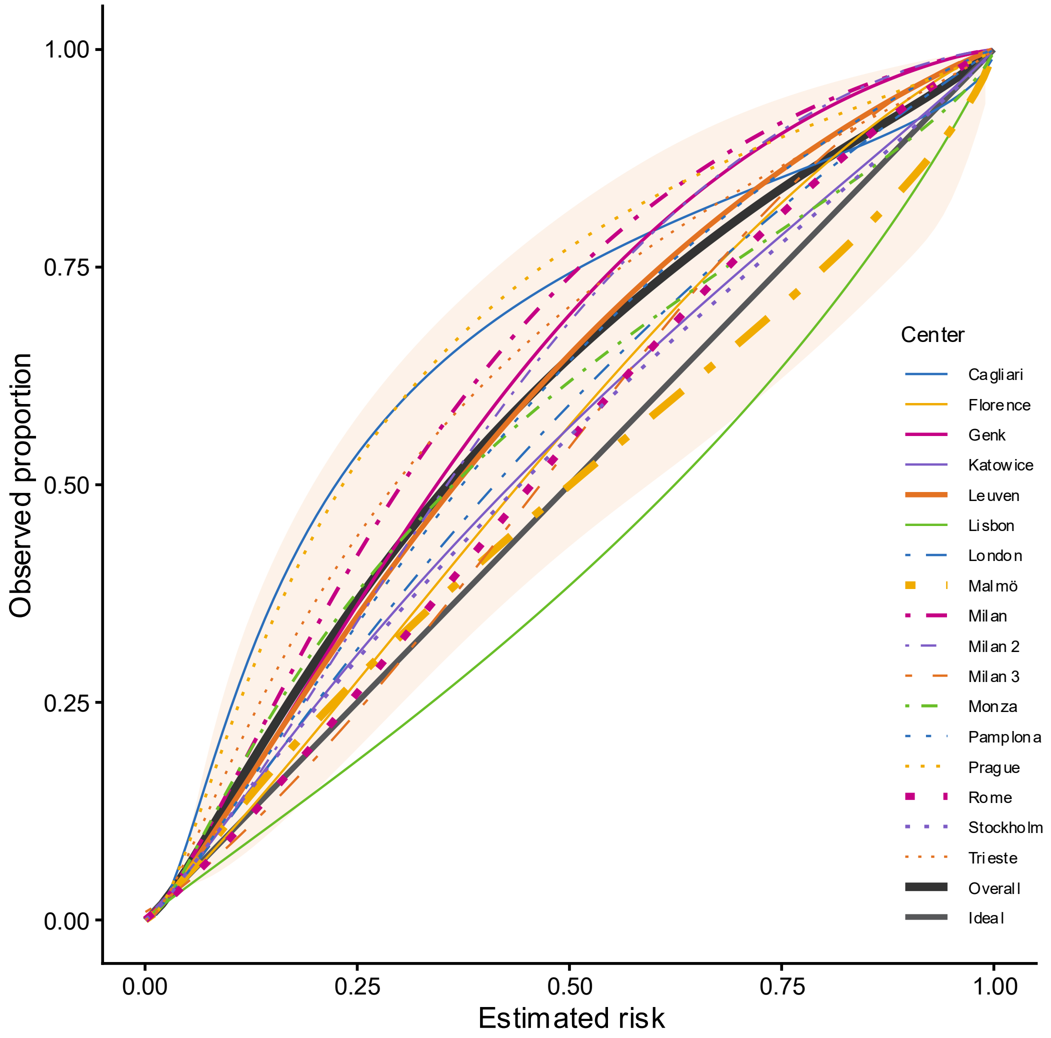

**Figure S32. Calibration plot for ADNEX ignoring CA125 in all patients (n = 6847). Overall calibration curves calculated taking clustering into account. Shaded area indicates 95% prediction interval. Linewidth is proportional with sample size. The thick black straight line is the ideal line denoting perfect agreement between predicted risk and observed proportion.**

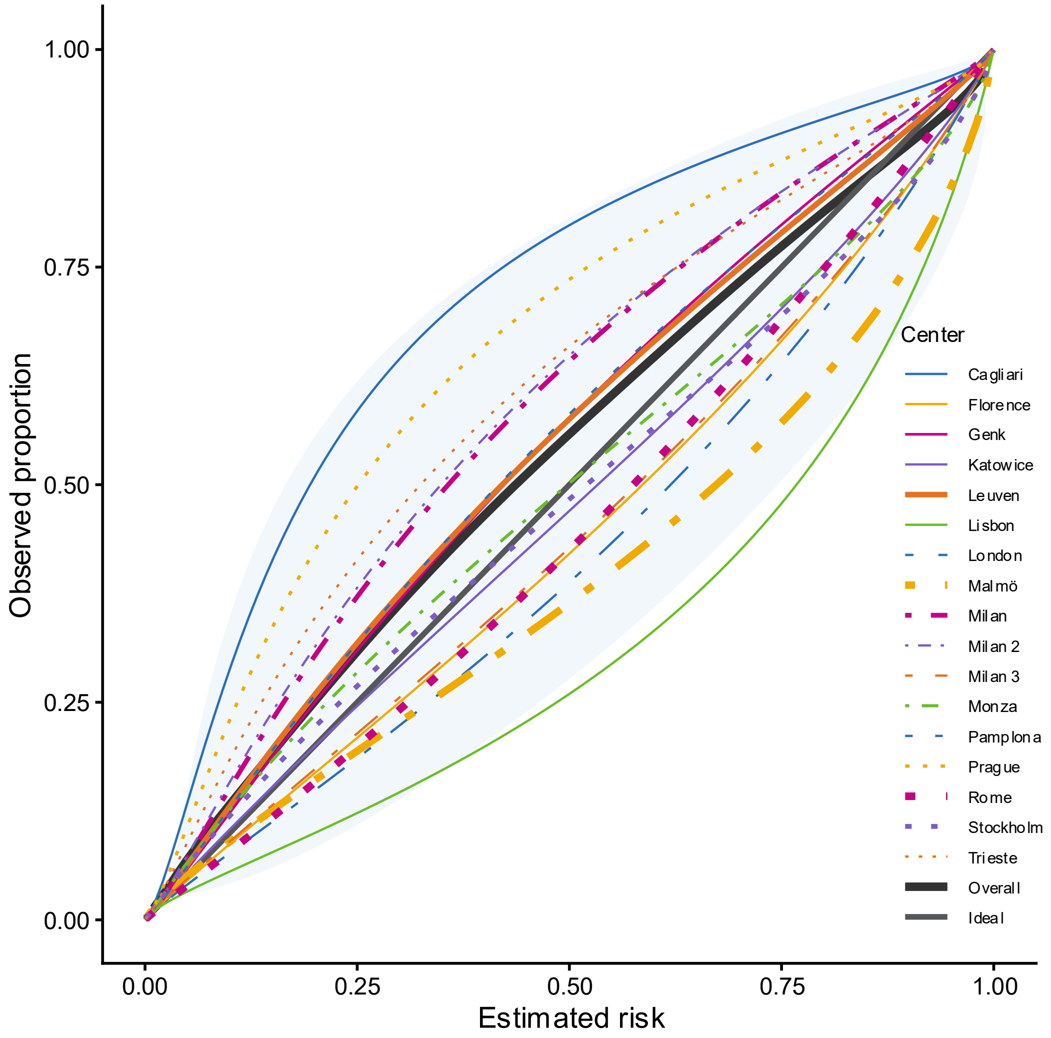

**Figure S33. Calibration plot for ADNEX2 where benign descriptors do not apply (n = 4657). Overall calibration curves calculated taking clustering into account. Shaded area indicates 95% prediction interval. Linewidth is proportional with sample size. The thick black straight line is the ideal line denoting perfect agreement between predicted risk and observed proportion.**

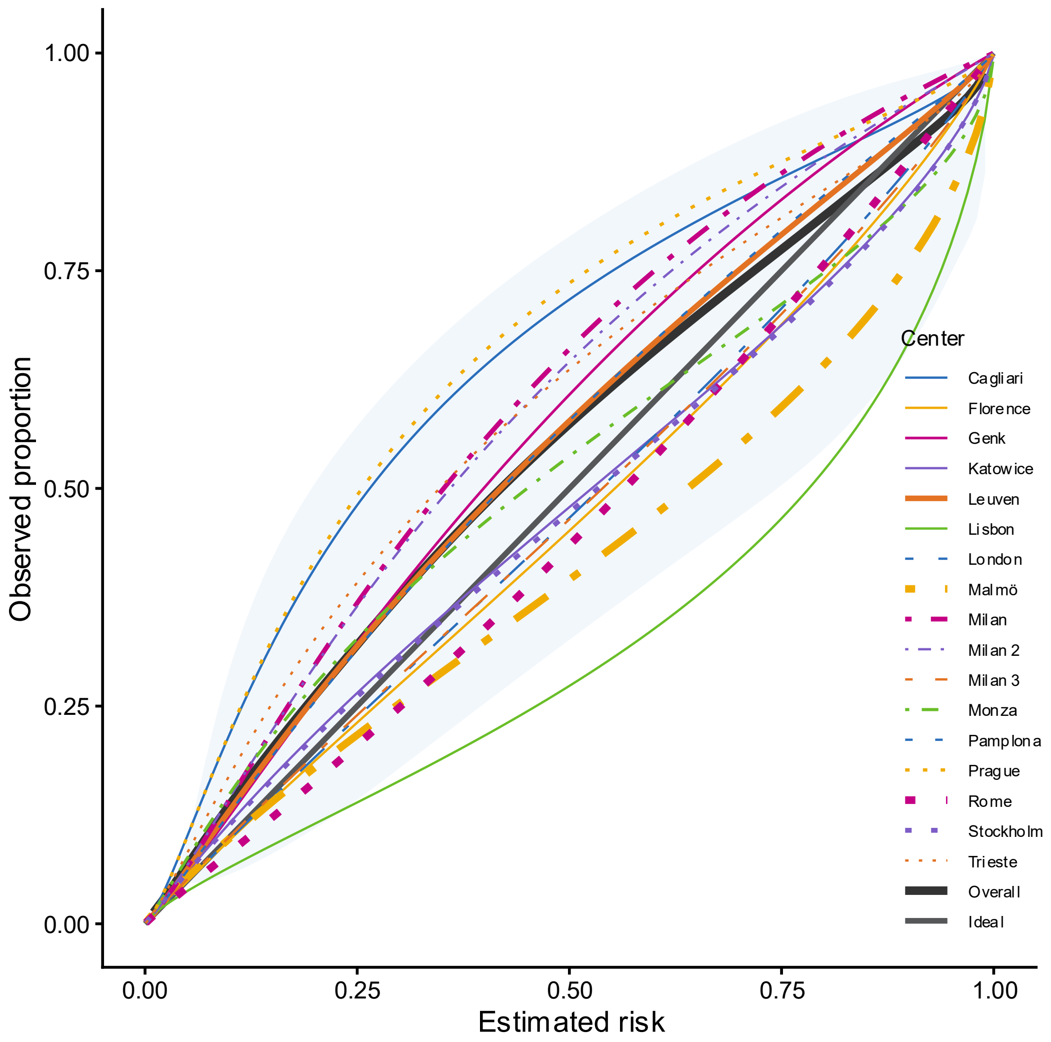

**Figure S34. Calibration plot for ADNEX2 ignoring CA125 if benign descriptors do not apply (n = 4657). Overall calibration curves calculated taking clustering into account. Shaded area indicates 95% prediction interval. Linewidth is proportional with sample size. The thick black straight line is the ideal line denoting perfect agreement between predicted risk and observed proportion.**

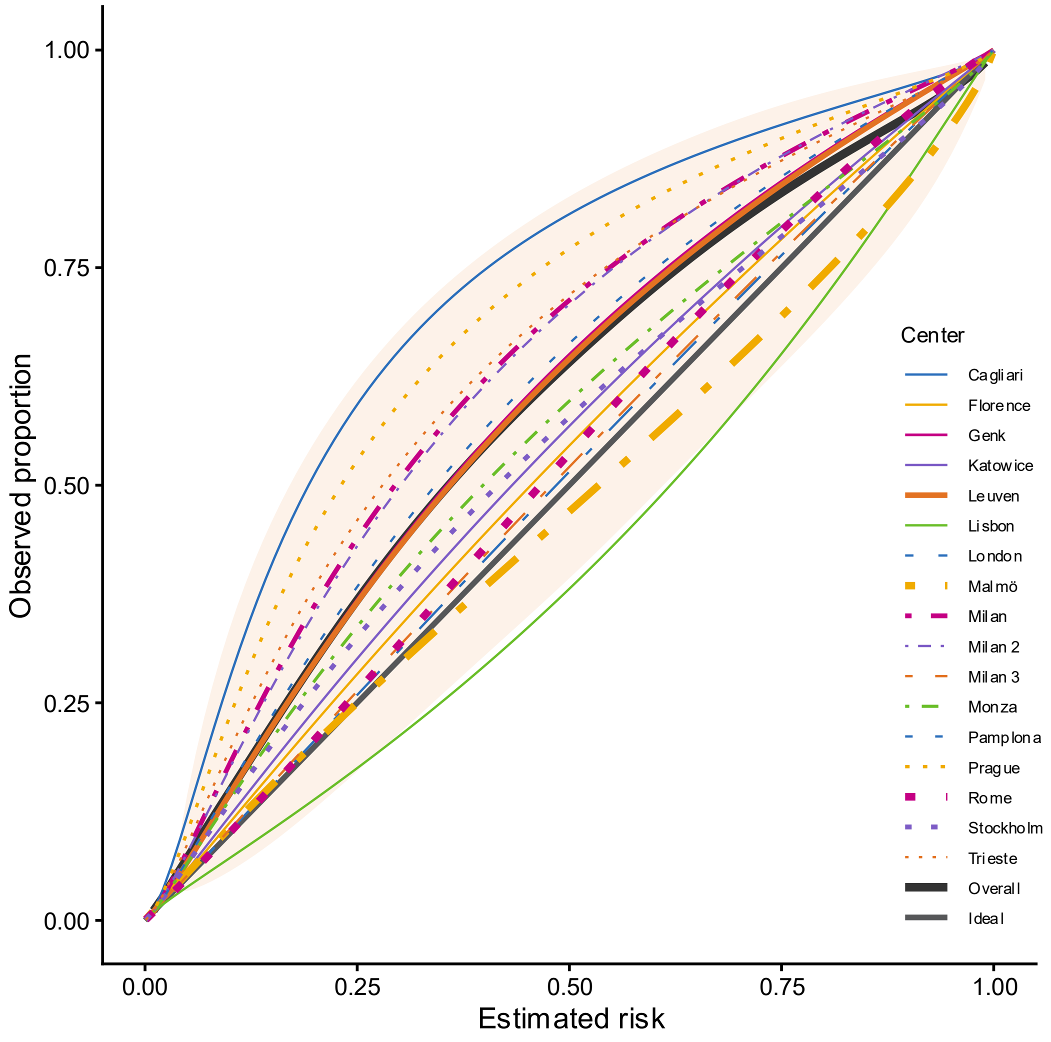

**Figure S35. Calibration plot for ADNEX in patients where benign descriptors do not apply (n = 4657). Overall calibration curves calculated taking clustering into account. Shaded area indicates 95% prediction interval. Linewidth is proportional with sample size. The thick black straight line is the ideal line denoting perfect agreement between predicted risk and observed proportion.**

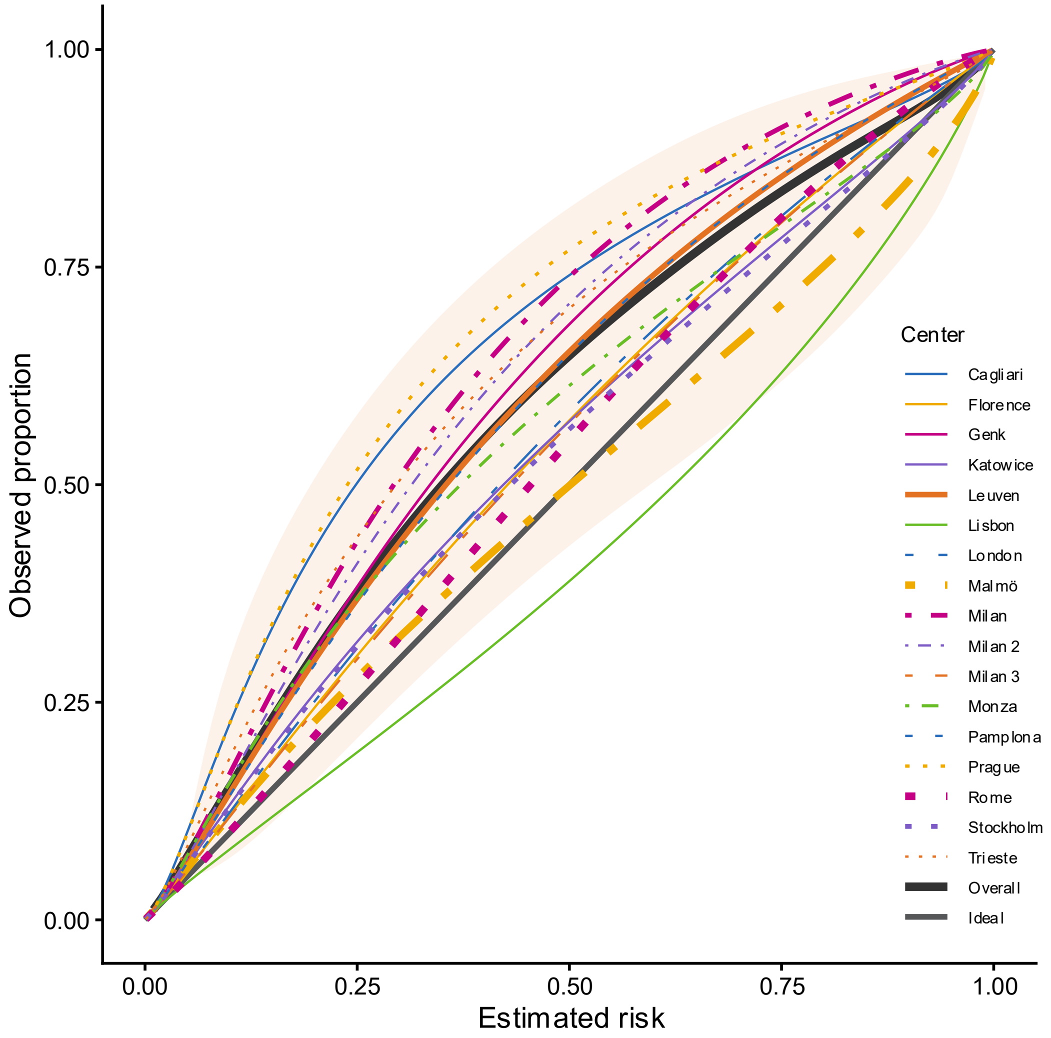

**Figure S36. Calibration plot for ADNEX ignoring CA125 in patients where benign descriptors do not apply (n = 4657). Overall calibration curves calculated taking clustering into account. Shaded area indicates 95% prediction interval. Linewidth is proportional with sample size. The thick black straight line is the ideal line denoting perfect agreement between predicted risk and observed proportion.**

##### S4.5.2 Multinomial calibration of ADNEX2 and ADNEX

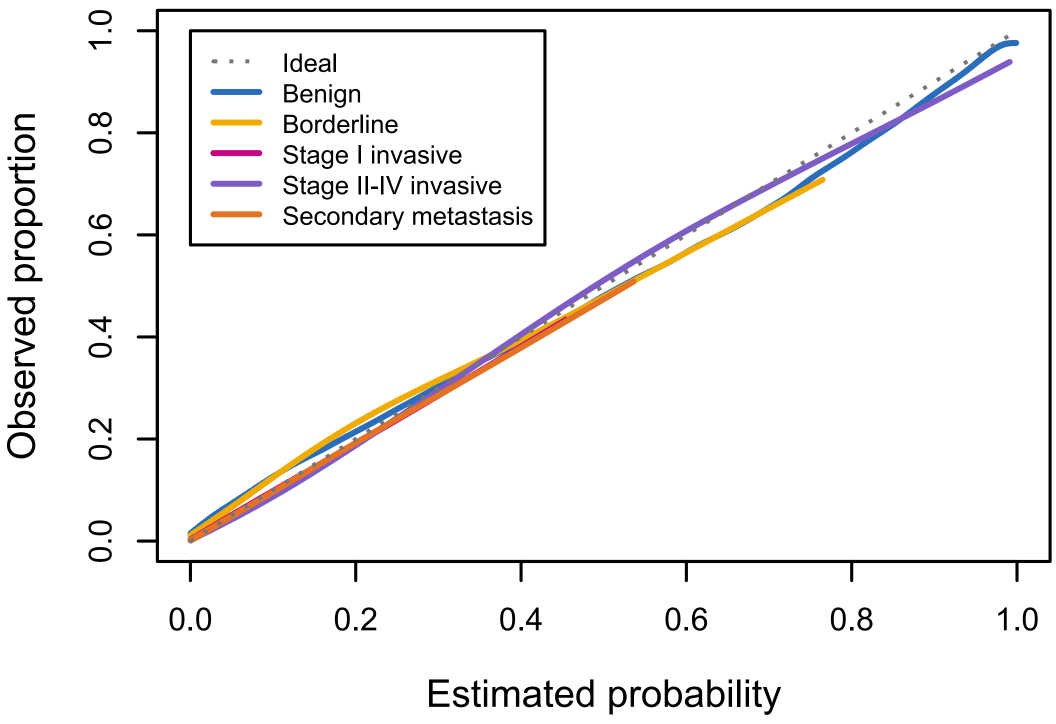

**Figure S37. Multinomial calibration plot for the two-step strategy with ADNEX2 in all patients (pooled analysis, n = 6847)**

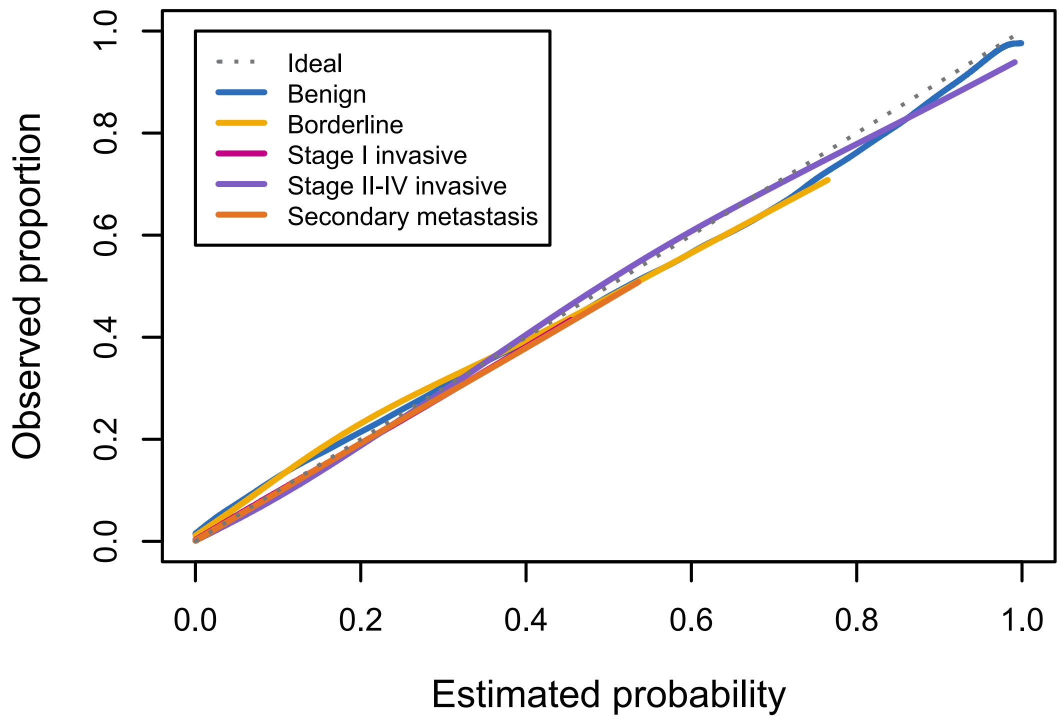

**Figure S38. Multinomial calibration plot for the two-step strategy with ADNEX2 ignoring CA125 in all patients (pooled analysis, n = 6847)**

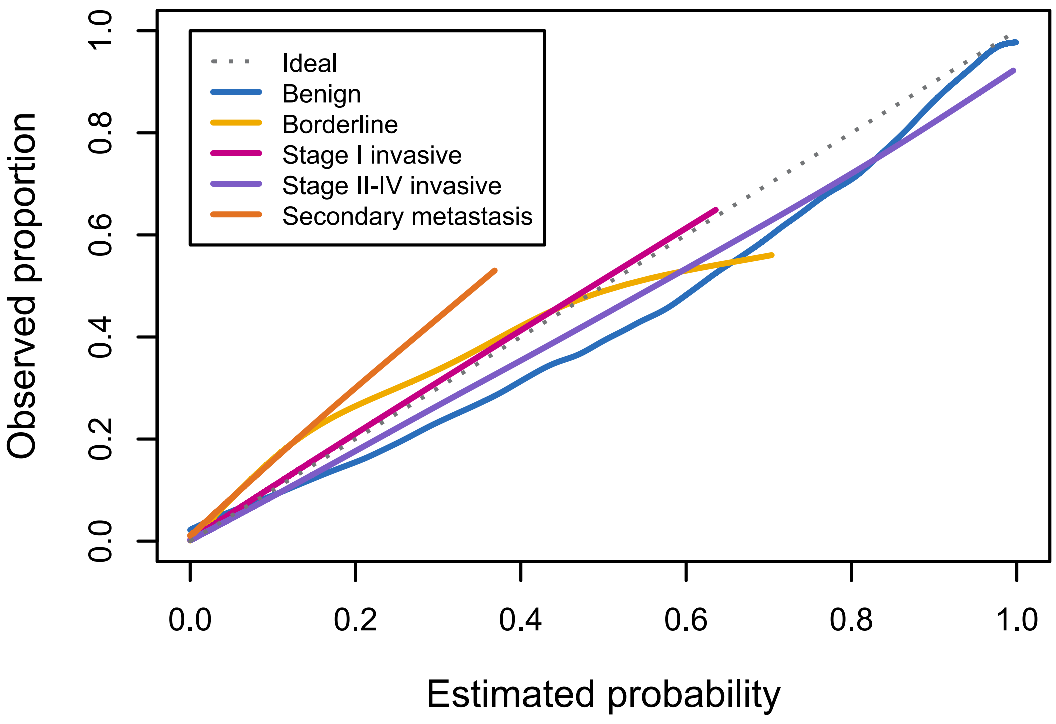

**Figure S39. Multinomial calibration plot for the two-step strategy with ADNEX in all patients (pooled analysis, n = 6847)**

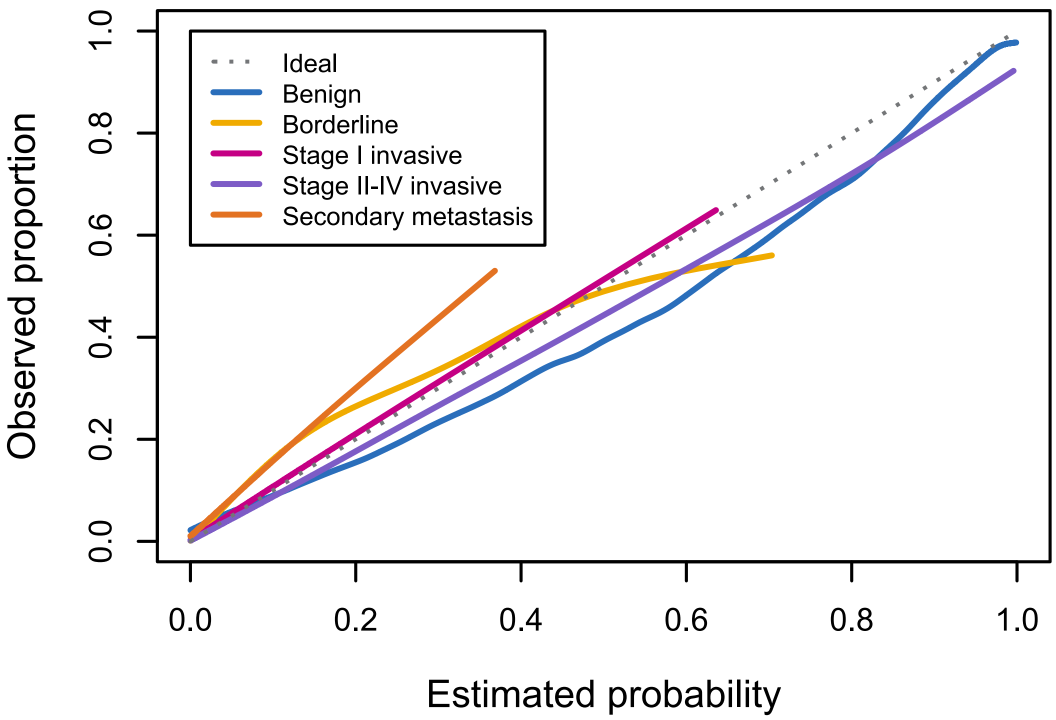

**Figure S40. Multinomial calibration plot for the two-step strategy with ADNEX  ignoring CA125 in all patients (pooled analysis, n = 6847)**

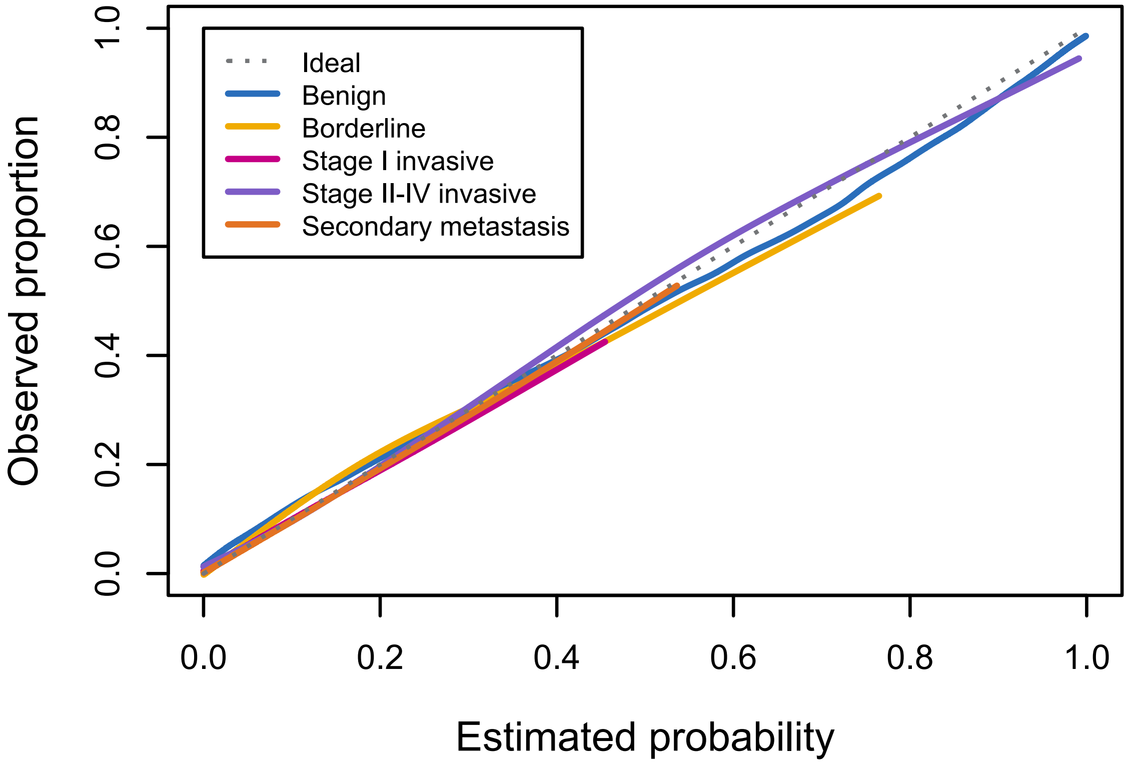

**Figure S41. Multinomial calibration plot for ADNEX2 where benign descriptors did not apply (pooled analysis, n = 4657)**

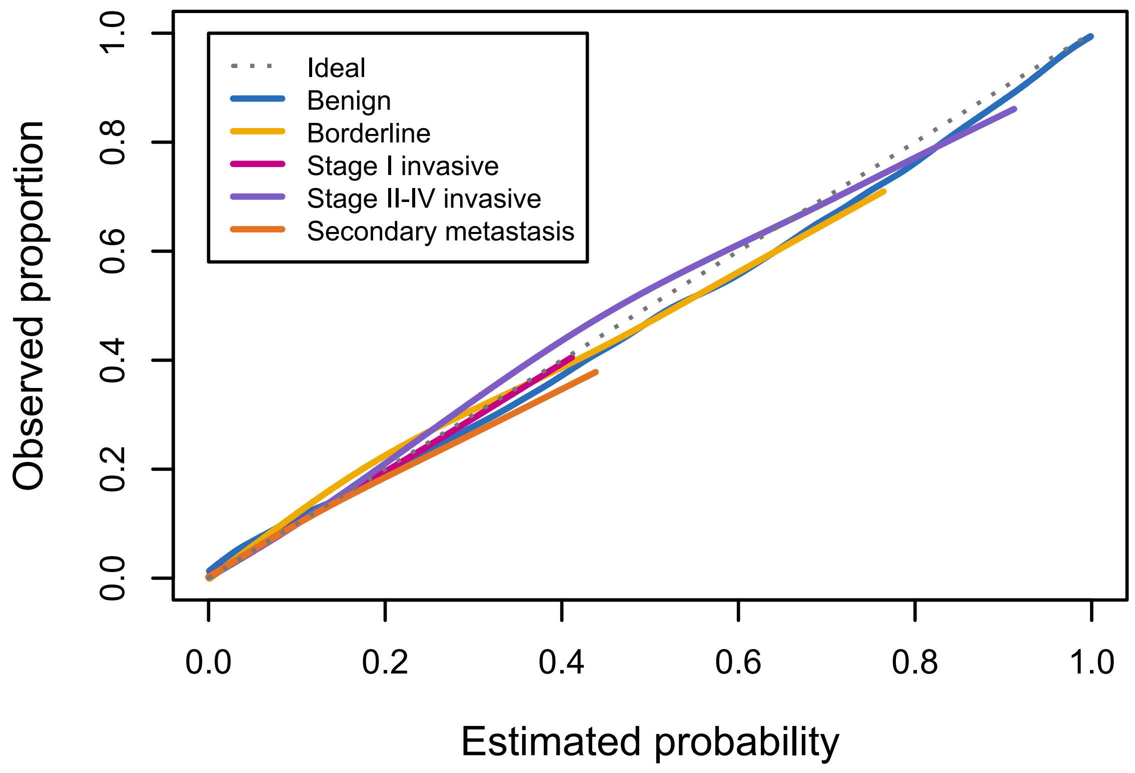

**Figure S42. Multinomial calibration plot for ADNEX2 ignoring CA125 where benign descriptors did not apply (pooled analysis, n = 4657)**

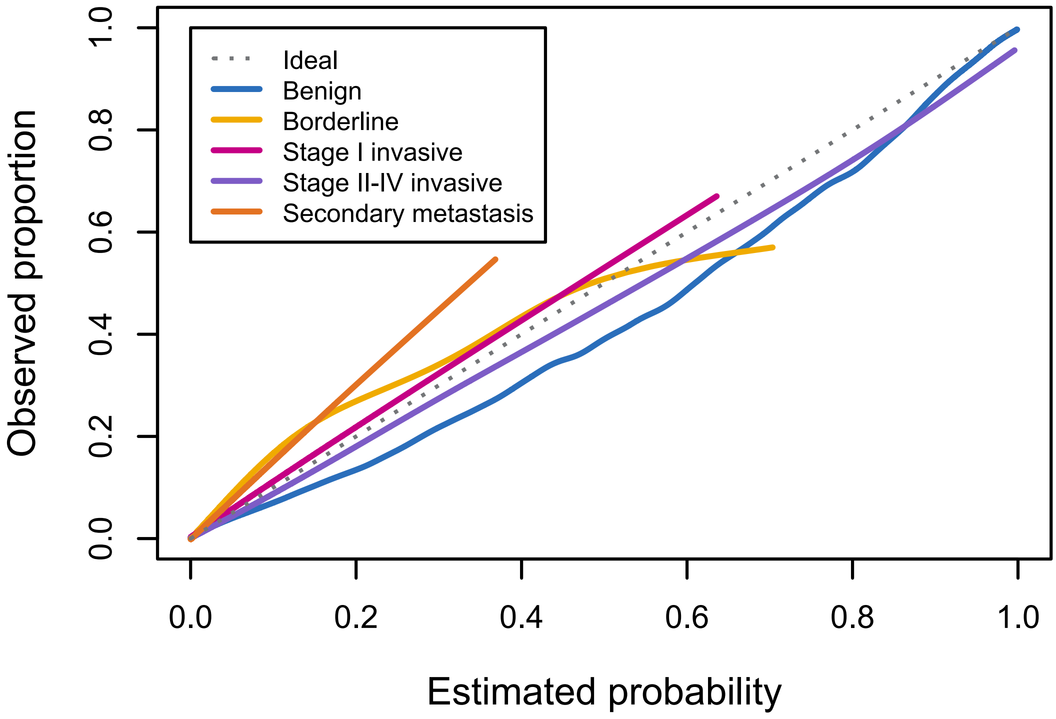

**Figure S43. Multinomial calibration plot for ADNEX where benign descriptors did not apply (pooled analysis, n = 4657)**

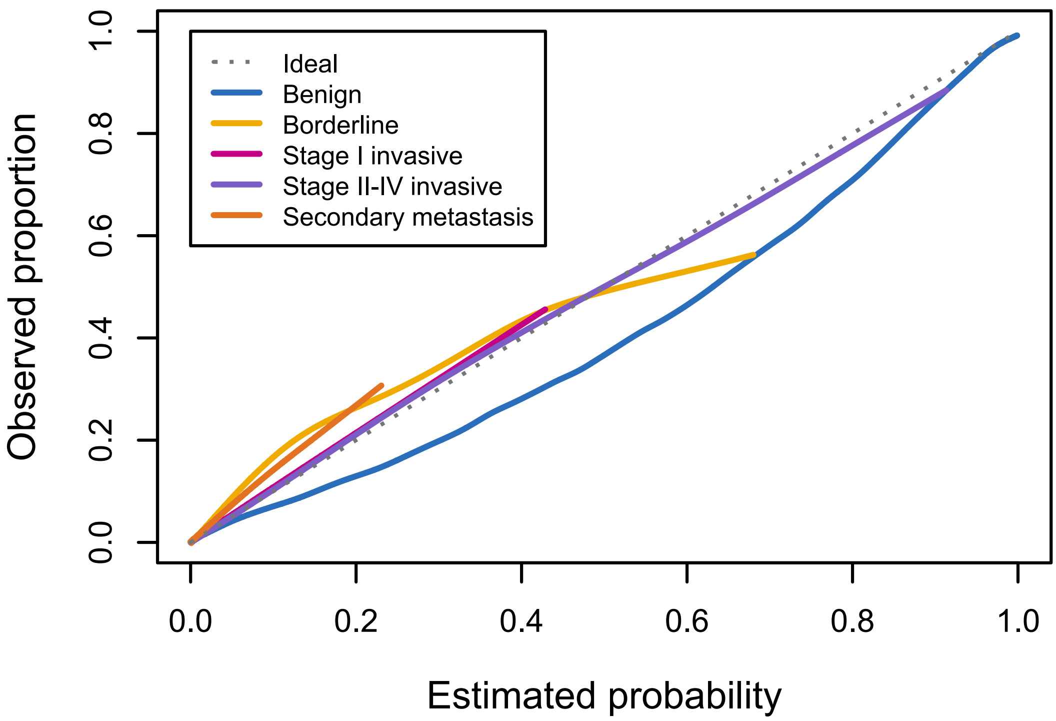

**Figure S44. Multinomial calibration plot for ADNEX ignoring CA125 where benign descriptors did not apply (pooled analysis, n = 4657)**

#### S4.6 CLASSIFICATION

**Table S14. Classification metrics for ADNEX2 and ADNEX**

| Strategy | Threshold | Sensitivity (95% CI) | Specificity (95% CI) | PPV (95% CI) | NPV (95% CI) |
|---|---|---|---|---|---|
| **All patients (N = 6847)** | | | | | |
| **Two step strategy with ADNEX2** | 0.01 | 0.97 (0.94-0.99) | 0.52 (0.45-0.60) | 0.35 (0.26-0.45) | 0.99 (0.98-0.99) |
| | 0.02 | 0.96 (0.93-0.98) | 0.63 (0.55-0.70) | 0.41 (0.33-0.50) | 0.98 (0.98-0.99) |
| | 0.03 | 0.96 (0.93-0.97) | 0.70 (0.63-0.76) | 0.46 (0.37-0.55) | 0.98 (0.98-0.99) |
| | 0.04 | 0.95 (0.92-0.97) | 0.74 (0.68-0.79) | 0.49 (0.40-0.58) | 0.98 (0.98-0.99) |
| | 0.05 | 0.95 (0.92-0.97) | 0.77 (0.71-0.81) | 0.52 (0.43-0.61) | 0.98 (0.98-0.99) |
| | **0.10** | **0.93 (0.89-0.95)** | **0.84 (0.79-0.87)** | **0.60 (0.51-0.69)** | **0.98 (0.97-0.98)** |
| | 0.15 | 0.90 (0.85-0.94) | 0.88 (0.83-0.91) | 0.66 (0.57-0.74) | 0.97 (0.96-0.98) |
| | 0.20 | 0.87 (0.81-0.91) | 0.90 (0.86-0.93) | 0.70 (0.61-0.77) | 0.96 (0.95-0.97) |
| | 0.25 | 0.84 (0.77-0.89) | 0.92 (0.88-0.94) | 0.73 (0.65-0.79) | 0.95 (0.94-0.96) |
| | 0.30 | 0.81 (0.74-0.87) | 0.93 (0.90-0.95) | 0.76 (0.69-0.81) | 0.95 (0.93-0.96) |
| | 0.35 | 0.79 (0.71-0.85) | 0.94 (0.91-0.96) | 0.77 (0.70-0.82) | 0.94 (0.92-0.96) |
| | 0.40 | 0.77 (0.70-0.84) | 0.94 (0.92-0.96) | 0.79 (0.73-0.84) | 0.94 (0.92-0.95) |
| **Two step strategy with ADNEX2 ignoring CA125** | 0.01 | 0.97 (0.95-0.98) | 0.52 (0.44-0.59) | 0.35 (0.26-0.44) | 0.99 (0.98-0.99) |
| | 0.02 | 0.96 (0.93-0.98) | 0.61 (0.53-0.69) | 0.40 (0.32-0.49) | 0.98 (0.98-0.99) |
| | 0.03 | 0.96 (0.93-0.98) | 0.68 (0.61-0.74) | 0.44 (0.36-0.53) | 0.98 (0.98-0.99) |
| | 0.04 | 0.96 (0.92-0.97) | 0.72 (0.66-0.78) | 0.48 (0.40-0.57) | 0.98 (0.98-0.99) |
| | 0.05 | 0.95 (0.92-0.97) | 0.76 (0.70-0.80) | 0.51 (0.42-0.60) | 0.98 (0.98-0.99) |
| | **0.10** | **0.93 (0.89-0.95)** | **0.83 (0.78-0.87)** | **0.59 (0.50-0.68)** | **0.98 (0.97-0.98)** |
| | 0.15 | 0.90 (0.85-0.94) | 0.87 (0.82-0.90) | 0.65 (0.56-0.72) | 0.97 (0.96-0.98) |
| | 0.20 | 0.87 (0.81-0.92) | 0.89 (0.85-0.92) | 0.68 (0.60-0.75) | 0.96 (0.95-0.97) |
| | 0.25 | 0.85 (0.78-0.90) | 0.91 (0.87-0.94) | 0.71 (0.64-0.78) | 0.95 (0.94-0.96) |
| | 0.30 | 0.82 (0.74-0.88) | 0.92 (0.89-0.94) | 0.74 (0.66-0.80) | 0.95 (0.93-0.96) |
| | 0.35 | 0.79 (0.71-0.86) | 0.93 (0.90-0.95) | 0.76 (0.69-0.82) | 0.94 (0.93-0.96) |
| | 0.40 | 0.78 (0.71-0.84) | 0.94 (0.92-0.96) | 0.78 (0.72-0.84) | 0.94 (0.92-0.95) |
| **Two step strategy with ADNEX** | 0.01 | 0.97 (0.95-0.98) | 0.51 (0.44-0.58) | 0.34 (0.26-0.44) | 0.99 (0.98-0.99) |
| | 0.02 | 0.96 (0.93-0.98) | 0.60 (0.51-0.69) | 0.40 (0.33-0.48) | 0.98 (0.98-0.99) |
| | 0.03 | 0.95 (0.92-0.97) | 0.68 (0.61-0.75) | 0.46 (0.38-0.53) | 0.98 (0.98-0.99) |
| | 0.04 | 0.95 (0.92-0.97) | 0.74 (0.68-0.80) | 0.50 (0.42-0.58) | 0.98 (0.98-0.99) |
| | 0.05 | 0.95 (0.92-0.97) | 0.77 (0.72-0.82) | 0.53 (0.44-0.61) | 0.98 (0.98-0.99) |
| | **0.10** | **0.92 (0.87-0.95)** | **0.85 (0.80-0.89)** | **0.62 (0.54-0.70)** | **0.97 (0.97-0.98)** |
| | 0.15 | 0.88 (0.82-0.92) | 0.89 (0.85-0.92) | 0.68 (0.60-0.75) | 0.96 (0.96-0.97) |
| | 0.20 | 0.84 (0.77-0.90) | 0.91 (0.88-0.94) | 0.72 (0.64-0.78) | 0.95 (0.94-0.96) |
| | 0.25 | 0.80 (0.73-0.86) | 0.93 (0.90-0.95) | 0.75 (0.69-0.81) | 0.94 (0.93-0.96) |
| | 0.30 | 0.78 (0.71-0.84) | 0.94 (0.92-0.96) | 0.79 (0.73-0.84) | 0.94 (0.92-0.95) |
| | 0.35 | 0.77 (0.69-0.83) | 0.95 (0.92-0.96) | 0.81 (0.76-0.85) | 0.93 (0.92-0.95) |
| | 0.40 | 0.75 (0.67-0.81) | 0.95 (0.93-0.97) | 0.83 (0.78-0.87) | 0.93 (0.91-0.94) |
| **Two step strategy with ADNEX ignoring CA125** | 0.01 | 0.97 (0.95-0.98) | 0.51 (0.44-0.58) | 0.34 (0.26-0.44) | 0.99 (0.98-0.99) |
| | 0.02 | 0.96 (0.93-0.98) | 0.59 (0.50-0.67) | 0.40 (0.32-0.48) | 0.98 (0.98-0.99) |
| | 0.03 | 0.96 (0.93-0.98) | 0.67 (0.59-0.74) | 0.45 (0.38-0.52) | 0.98 (0.98-0.99) |
| | 0.04 | 0.95 (0.92-0.97) | 0.72 (0.65-0.78) | 0.48 (0.40-0.56) | 0.98 (0.98-0.99) |
| | 0.05 | 0.95 (0.91-0.97) | 0.76 (0.70-0.81) | 0.52 (0.43-0.60) | 0.98 (0.98-0.99) |
| | **0.10** | **0.92 (0.87-0.95)** | **0.84 (0.79-0.88)** | **0.62 (0.54-0.69)** | **0.97 (0.97-0.98)** |
| | 0.15 | 0.88 (0.83-0.92) | 0.88 (0.84-0.92) | 0.67 (0.60-0.74) | 0.96 (0.96-0.97) |
| | 0.20 | 0.84 (0.77-0.89) | 0.91 (0.87-0.93) | 0.71 (0.63-0.77) | 0.95 (0.94-0.96) |
| | 0.25 | 0.81 (0.73-0.87) | 0.92 (0.89-0.95) | 0.74 (0.67-0.80) | 0.94 (0.93-0.96) |
| | 0.30 | 0.79 (0.71-0.85) | 0.93 (0.91-0.95) | 0.77 (0.70-0.82) | 0.94 (0.92-0.95) |
| | 0.35 | 0.76 (0.69-0.82) | 0.94 (0.92-0.96) | 0.80 (0.74-0.84) | 0.93 (0.92-0.95) |
| | 0.40 | 0.73 (0.65-0.80) | 0.95 (0.93-0.96) | 0.81 (0.76-0.85) | 0.92 (0.90-0.94) |
| **ADNEX2** | 0.01 | 0.99 (0.97->.99) | 0.20 (0.15-0.26) | 0.24 (0.17-0.33) | 0.98 (0.97-0.99) |
| | 0.02 | 0.97 (0.95-0.99) | 0.47 (0.37-0.58) | 0.34 (0.26-0.42) | 0.99 (0.98-0.99) |

| Strategy | Threshold | Sensitivity (95% CI) | Specificity (95% CI) | PPV (95% CI) | NPV (95% CI) |
|---|---|---|---|---|---|
| | 0.03 | 0.96 (0.94-0.98) | 0.63 (0.54-0.71) | 0.42 (0.34-0.50) | 0.98 (0.98-0.99) |
| | 0.04 | 0.96 (0.93-0.98) | 0.71 (0.64-0.77) | 0.47 (0.38-0.56) | 0.98 (0.98-0.99) |
| | 0.05 | 0.95 (0.93-0.97) | 0.75 (0.69-0.80) | 0.51 (0.42-0.60) | 0.98 (0.98-0.99) |
| | **0.10** | **0.92 (0.89-0.95)** | **0.84 (0.79-0.87)** | **0.60 (0.51-0.69)** | **0.98 (0.97-0.98)** |
| | 0.15 | 0.90 (0.85-0.94) | 0.88 (0.83-0.91) | 0.66 (0.57-0.74) | 0.97 (0.96-0.98) |
| | 0.20 | 0.87 (0.81-0.91) | 0.90 (0.86-0.93) | 0.70 (0.61-0.77) | 0.96 (0.95-0.97) |
| | 0.25 | 0.84 (0.77-0.89) | 0.92 (0.88-0.94) | 0.73 (0.65-0.79) | 0.95 (0.94-0.96) |
| | 0.30 | 0.81 (0.74-0.87) | 0.93 (0.90-0.95) | 0.76 (0.69-0.81) | 0.95 (0.93-0.96) |
| | 0.35 | 0.79 (0.71-0.85) | 0.94 (0.91-0.96) | 0.77 (0.70-0.82) | 0.94 (0.92-0.96) |
| | 0.40 | 0.77 (0.70-0.84) | 0.94 (0.92-0.96) | 0.79 (0.73-0.84) | 0.94 (0.92-0.95) |
| **ADNEX2 ignoring CA125** | 0.01 | 0.99 (0.97-0.99) | 0.19 (0.14-0.24) | 0.24 (0.17-0.33) | 0.98 (0.96-0.99) |
| | 0.02 | 0.98 (0.95-0.99) | 0.44 (0.34-0.55) | 0.33 (0.25-0.41) | 0.99 (0.98-0.99) |
| | 0.03 | 0.97 (0.94-0.98) | 0.61 (0.52-0.70) | 0.41 (0.33-0.49) | 0.99 (0.98-0.99) |
| | 0.04 | 0.96 (0.93-0.98) | 0.70 (0.62-0.76) | 0.46 (0.38-0.54) | 0.98 (0.98-0.99) |
| | 0.05 | 0.95 (0.92-0.97) | 0.74 (0.68-0.80) | 0.50 (0.42-0.58) | 0.98 (0.98-0.99) |
| | **0.10** | **0.93 (0.89-0.95)** | **0.83 (0.78-0.87)** | **0.59 (0.50-0.68)** | **0.98 (0.97-0.98)** |
| | 0.15 | 0.90 (0.85-0.94) | 0.87 (0.82-0.90) | 0.65 (0.56-0.72) | 0.97 (0.96-0.98) |
| | 0.20 | 0.87 (0.81-0.92) | 0.89 (0.85-0.92) | 0.68 (0.60-0.75) | 0.96 (0.95-0.97) |
| | 0.25 | 0.85 (0.78-0.90) | 0.91 (0.87-0.94) | 0.71 (0.64-0.78) | 0.95 (0.94-0.96) |
| | 0.30 | 0.82 (0.74-0.88) | 0.92 (0.89-0.94) | 0.74 (0.66-0.80) | 0.95 (0.93-0.96) |
| | 0.35 | 0.79 (0.71-0.86) | 0.93 (0.90-0.95) | 0.76 (0.69-0.82) | 0.94 (0.93-0.96) |
| | 0.40 | 0.78 (0.71-0.84) | 0.94 (0.92-0.96) | 0.78 (0.72-0.84) | 0.94 (0.92-0.95) |
| **ADNEX** | 0.01 | 0.99 (0.97->.99) | 0.16 (0.11-0.22) | 0.24 (0.17-0.32) | 0.98 (0.96-0.99) |
| | 0.02 | 0.98 (0.95-0.99) | 0.40 (0.28-0.53) | 0.32 (0.26-0.40) | 0.98 (0.97-0.99) |
| | 0.03 | 0.96 (0.93-0.98) | 0.60 (0.50-0.70) | 0.41 (0.34-0.48) | 0.98 (0.98-0.99) |
| | 0.04 | 0.96 (0.92-0.97) | 0.71 (0.63-0.77) | 0.48 (0.40-0.55) | 0.98 (0.98-0.99) |
| | 0.05 | 0.95 (0.92-0.97) | 0.76 (0.70-0.81) | 0.52 (0.43-0.60) | 0.98 (0.98-0.99) |
| | **0.10** | **0.92 (0.87-0.95)** | **0.85 (0.80-0.89)** | **0.62 (0.54-0.70)** | **0.97 (0.97-0.98)** |
| | 0.15 | 0.88 (0.82-0.92) | 0.89 (0.85-0.92) | 0.68 (0.60-0.75) | 0.96 (0.96-0.97) |
| | 0.20 | 0.84 (0.77-0.90) | 0.91 (0.88-0.94) | 0.72 (0.64-0.78) | 0.95 (0.94-0.96) |
| | 0.25 | 0.80 (0.73-0.86) | 0.93 (0.90-0.95) | 0.75 (0.69-0.81) | 0.94 (0.93-0.96) |
| | 0.30 | 0.78 (0.71-0.84) | 0.94 (0.92-0.96) | 0.79 (0.73-0.84) | 0.94 (0.92-0.95) |
| | 0.35 | 0.77 (0.69-0.83) | 0.95 (0.92-0.96) | 0.81 (0.76-0.85) | 0.93 (0.92-0.95) |
| | 0.40 | 0.75 (0.67-0.81) | 0.95 (0.93-0.97) | 0.83 (0.78-0.87) | 0.93 (0.91-0.94) |
| **ADNEX ignoring CA125** | 0.01 | 0.99 (0.98->.99) | 0.16 (0.11-0.22) | 0.24 (0.17-0.32) | 0.98 (0.96-0.99) |
| | 0.02 | 0.98 (0.95-0.99) | 0.38 (0.27-0.51) | 0.32 (0.25-0.39) | 0.98 (0.97-0.99) |
| | 0.03 | 0.97 (0.94-0.98) | 0.58 (0.47-0.68) | 0.40 (0.34-0.47) | 0.99 (0.98-0.99) |
| | 0.04 | 0.96 (0.92-0.98) | 0.69 (0.60-0.76) | 0.46 (0.39-0.53) | 0.98 (0.98-0.99) |
| | 0.05 | 0.95 (0.91-0.97) | 0.74 (0.68-0.80) | 0.50 (0.42-0.58) | 0.98 (0.98-0.99) |
| | **0.10** | **0.92 (0.87-0.95)** | **0.84 (0.79-0.88)** | **0.61 (0.54-0.69)** | **0.97 (0.97-0.98)** |
| | 0.15 | 0.88 (0.83-0.92) | 0.88 (0.84-0.92) | 0.67 (0.60-0.74) | 0.96 (0.96-0.97) |
| | 0.20 | 0.84 (0.77-0.89) | 0.91 (0.87-0.93) | 0.71 (0.63-0.77) | 0.95 (0.94-0.96) |
| | 0.25 | 0.81 (0.73-0.87) | 0.92 (0.89-0.95) | 0.74 (0.67-0.80) | 0.94 (0.93-0.96) |
| | 0.30 | 0.79 (0.71-0.85) | 0.93 (0.91-0.95) | 0.77 (0.70-0.82) | 0.94 (0.92-0.95) |
| | 0.35 | 0.76 (0.69-0.82) | 0.94 (0.92-0.96) | 0.80 (0.74-0.84) | 0.93 (0.92-0.95) |
| | 0.40 | 0.73 (0.65-0.80) | 0.95 (0.93-0.96) | 0.81 (0.76-0.85) | 0.92 (0.90-0.94) |
| **BDs do not apply (N = 4657)** | | | | | |
| **ADNEX2** | 0.01 | 0.99 (0.98->.99) | 0.11 (0.07-0.16) | 0.35 (0.26-0.45) | 0.97 (0.94-0.99) |
| | 0.02 | 0.98 (0.97-0.99) | 0.29 (0.22-0.37) | 0.41 (0.32-0.50) | 0.97 (0.95-0.98) |
| | 0.03 | 0.98 (0.96-0.99) | 0.42 (0.36-0.50) | 0.46 (0.37-0.55) | 0.97 (0.96-0.98) |
| | 0.04 | 0.97 (0.95-0.98) | 0.50 (0.44-0.56) | 0.49 (0.40-0.58) | 0.98 (0.96-0.98) |
| | 0.05 | 0.97 (0.95-0.98) | 0.56 (0.51-0.61) | 0.52 (0.43-0.61) | 0.97 (0.96-0.98) |
| | **0.10** | **0.95 (0.92-0.97)** | **0.69 (0.64-0.74)** | **0.60 (0.51-0.69)** | **0.96 (0.95-0.97)** |
| | 0.15 | 0.92 (0.88-0.95) | 0.76 (0.71-0.81) | 0.66 (0.57-0.74) | 0.95 (0.93-0.96) |
| | 0.20 | 0.90 (0.84-0.93) | 0.80 (0.75-0.84) | 0.70 (0.62-0.77) | 0.94 (0.92-0.95) |
| | 0.25 | 0.87 (0.81-0.91) | 0.84 (0.79-0.87) | 0.73 (0.65-0.80) | 0.92 (0.90-0.94) |
| | 0.30 | 0.84 (0.77-0.89) | 0.86 (0.82-0.90) | 0.76 (0.69-0.82) | 0.91 (0.89-0.93) |
| | 0.35 | 0.82 (0.74-0.87) | 0.88 (0.84-0.91) | 0.77 (0.71-0.83) | 0.90 (0.88-0.92) |

| Strategy | Threshold | Sensitivity (95% CI) | Specificity (95% CI) | PPV (95% CI) | NPV (95% CI) |
|---|---|---|---|---|---|
| | 0.40 | 0.80 (0.72-0.86) | 0.89 (0.86-0.92) | 0.80 (0.74-0.85) | 0.89 (0.87-0.92) |
| **ADNEX2 ignoring CA125** | 0.01 | 0.99 (0.98->.99) | 0.10 (0.07-0.14) | 0.35 (0.26-0.44) | 0.96 (0.92-0.98) |
| | 0.02 | 0.98 (0.97-0.99) | 0.26 (0.19-0.34) | 0.40 (0.32-0.49) | 0.97 (0.95-0.99) |
| | 0.03 | 0.98 (0.96-0.99) | 0.39 (0.32-0.47) | 0.44 (0.36-0.53) | 0.98 (0.96-0.98) |
| | 0.04 | 0.98 (0.96-0.99) | 0.48 (0.42-0.54) | 0.48 (0.40-0.57) | 0.98 (0.96-0.98) |
| | 0.05 | 0.97 (0.95-0.98) | 0.54 (0.49-0.59) | 0.51 (0.42-0.60) | 0.97 (0.96-0.98) |
| | **0.10** | **0.95 (0.92-0.97)** | **0.68 (0.63-0.73)** | **0.59 (0.50-0.68)** | **0.96 (0.95-0.97)** |
| | 0.15 | 0.92 (0.88-0.95) | 0.75 (0.70-0.80) | 0.65 (0.56-0.73) | 0.95 (0.93-0.96) |
| | 0.20 | 0.90 (0.84-0.93) | 0.79 (0.74-0.83) | 0.68 (0.60-0.75) | 0.93 (0.92-0.95) |
| | 0.25 | 0.87 (0.82-0.91) | 0.82 (0.78-0.86) | 0.72 (0.64-0.78) | 0.92 (0.90-0.94) |
| | 0.30 | 0.85 (0.78-0.90) | 0.85 (0.81-0.88) | 0.74 (0.67-0.80) | 0.91 (0.89-0.93) |
| | 0.35 | 0.82 (0.74-0.88) | 0.87 (0.83-0.90) | 0.76 (0.70-0.82) | 0.90 (0.88-0.92) |
| | 0.40 | 0.81 (0.73-0.86) | 0.88 (0.85-0.91) | 0.79 (0.73-0.84) | 0.90 (0.87-0.92) |
| **ADNEX** | 0.01 | 0.99 (0.98->.99) | 0.10 (0.07-0.14) | 0.34 (0.26-0.44) | 0.97 (0.93-0.98) |
| | 0.02 | 0.99 (0.98-0.99) | 0.22 (0.14-0.33) | 0.40 (0.32-0.48) | 0.97 (0.95-0.98) |
| | 0.03 | 0.98 (0.96-0.99) | 0.40 (0.32-0.48) | 0.45 (0.38-0.53) | 0.97 (0.96-0.98) |
| | 0.04 | 0.97 (0.95-0.98) | 0.51 (0.44-0.57) | 0.50 (0.41-0.58) | 0.97 (0.96-0.98) |
| | 0.05 | 0.97 (0.94-0.98) | 0.57 (0.51-0.62) | 0.53 (0.44-0.61) | 0.97 (0.96-0.98) |
| | **0.10** | **0.94 (0.90-0.96)** | **0.71 (0.65-0.76)** | **0.62 (0.54-0.70)** | **0.96 (0.94-0.96)** |
| | 0.15 | 0.90 (0.86-0.94) | 0.79 (0.74-0.83) | 0.68 (0.60-0.76) | 0.94 (0.92-0.96) |
| | 0.20 | 0.87 (0.80-0.91) | 0.83 (0.79-0.87) | 0.72 (0.65-0.79) | 0.92 (0.90-0.94) |
| | 0.25 | 0.83 (0.76-0.88) | 0.86 (0.82-0.89) | 0.76 (0.69-0.81) | 0.91 (0.89-0.92) |
| | 0.30 | 0.81 (0.74-0.86) | 0.88 (0.85-0.91) | 0.79 (0.73-0.84) | 0.90 (0.87-0.92) |
| | 0.35 | 0.79 (0.72-0.85) | 0.90 (0.87-0.92) | 0.81 (0.76-0.86) | 0.89 (0.86-0.91) |
| | 0.40 | 0.77 (0.70-0.82) | 0.91 (0.88-0.93) | 0.84 (0.79-0.88) | 0.87 (0.85-0.89) |
| **ADNEX ignoring CA125** | 0.01 | 0.99 (0.98->.99) | 0.10 (0.07-0.14) | 0.34 (0.26-0.44) | 0.96 (0.92-0.98) |
| | 0.02 | 0.99 (0.98-0.99) | 0.20 (0.13-0.30) | 0.39 (0.32-0.48) | 0.97 (0.94-0.98) |
| | 0.03 | 0.98 (0.97-0.99) | 0.36 (0.28-0.45) | 0.45 (0.37-0.52) | 0.97 (0.96-0.98) |
| | 0.04 | 0.97 (0.95-0.99) | 0.47 (0.41-0.54) | 0.48 (0.40-0.56) | 0.97 (0.96-0.98) |
| | 0.05 | 0.97 (0.94-0.98) | 0.54 (0.49-0.60) | 0.51 (0.43-0.60) | 0.97 (0.96-0.98) |
| | **0.10** | **0.94 (0.90-0.96)** | **0.70 (0.64-0.75)** | **0.62 (0.54-0.69)** | **0.96 (0.94-0.96)** |
| | 0.15 | 0.91 (0.86-0.94) | 0.78 (0.72-0.82) | 0.67 (0.60-0.74) | 0.94 (0.92-0.95) |
| | 0.20 | 0.87 (0.80-0.91) | 0.82 (0.78-0.86) | 0.71 (0.63-0.78) | 0.92 (0.90-0.94) |
| | 0.25 | 0.84 (0.76-0.89) | 0.85 (0.81-0.88) | 0.75 (0.68-0.80) | 0.91 (0.89-0.93) |
| | 0.30 | 0.81 (0.74-0.87) | 0.87 (0.84-0.90) | 0.77 (0.71-0.82) | 0.90 (0.87-0.92) |
| | 0.35 | 0.79 (0.72-0.84) | 0.89 (0.86-0.92) | 0.81 (0.75-0.85) | 0.88 (0.86-0.90) |
| | 0.40 | 0.75 (0.68-0.81) | 0.90 (0.88-0.93) | 0.82 (0.77-0.86) | 0.87 (0.84-0.89) |

CI: Confidence interval; PPV: Positive predictive value; NPV: Negative Predictive value

#### S4.7 CLINICAL UTILITY

**Table S15. Clinical utility metrics for ADNEX2 and ADNEX**

| Strategy | Threshold | NB (95% Crl) | Delta NB | P-useful |
|---|---|---|---|---|
| **BDs alone** | 0.01 | 0.191 (0.12-0.28) | 0.0007 | 58% |
| **All patients (N = 6847)** | | | | |
| **2SS with ADNEX2** | 0.01 | 0.191 (0.12-0.28) | 0.0002 | 52% |
| | 0.02 | 0.187 (0.12-0.28) | 0.0049 | 78% |
| | 0.03 | 0.183 (0.11-0.27) | 0.0097 | 88% |
| | 0.04 | 0.182 (0.11-0.27) | 0.0163 | 94% |
| | 0.05 | 0.180 (0.11-0.27) | 0.0233 | 96% |
| | **0.10** | **0.170 (0.10-0.26)** | **0.0594** | **97%** |
| | 0.15 | 0.163 (0.09-0.25) | 0.1042 | 97% |
| | 0.20 | 0.154 (0.09-0.24) | 0.1538 | 96% |
| | 0.25 | 0.146 (0.08-0.23) | 0.1459 | 96% |
| | 0.30 | 0.138 (0.07-0.22) | 0.1385 | 96% |
| | 0.35 | 0.130 (0.07-0.21) | 0.1304 | 95% |
| | 0.40 | 0.126 (0.07-0.20) | 0.1255 | 96% |
| **2SS with ADNEX2 ignoring CA125** | 0.01 | 0.190 (0.12-0.28) | 0 | 50% |
| | 0.02 | 0.187 (0.12-0.27) | 0.0046 | 78% |
| | 0.03 | 0.184 (0.11-0.27) | 0.0097 | 87% |
| | 0.04 | 0.181 (0.11-0.27) | 0.0162 | 94% |
| | 0.05 | 0.179 (0.11-0.27) | 0.023 | 96% |
| | **0.10** | **0.170 (0.10-0.26)** | **0.0596** | **98%** |
| | 0.15 | 0.161 (0.09-0.25) | 0.1033 | 97% |
| | 0.20 | 0.153 (0.09-0.24) | 0.1528 | 97% |
| | 0.25 | 0.145 (0.08-0.23) | 0.1448 | 96% |
| | 0.30 | 0.137 (0.07-0.22) | 0.1371 | 95% |
| | 0.35 | 0.130 (0.07-0.21) | 0.1301 | 95% |
| | 0.40 | 0.126 (0.07-0.20) | 0.1263 | 95% |
| **2SS with ADNEX** | 0.01 | 0.190 (0.12-0.28) | 0 | 50% |
| | 0.02 | 0.187 (0.12-0.27) | 0.0043 | 76% |
| | 0.03 | 0.184 (0.11-0.27) | 0.0094 | 88% |
| | 0.04 | 0.182 (0.11-0.27) | 0.0163 | 94% |
| | 0.05 | 0.180 (0.11-0.27) | 0.0232 | 97% |
| | **0.10** | **0.170 (0.10-0.26)** | **0.0592** | **98%** |
| | 0.15 | 0.160 (0.09-0.25) | 0.102 | 98% |
| | 0.20 | 0.150 (0.08-0.23) | 0.1499 | 97% |
| | 0.25 | 0.141 (0.08-0.22) | 0.1412 | 97% |
| | 0.30 | 0.136 (0.07-0.22) | 0.1365 | 97% |
| | 0.35 | 0.131 (0.07-0.21) | 0.131 | 98% |
| | 0.40 | 0.127 (0.07-0.20) | 0.1266 | 98% |
| **2SS with ADNEX ignoring CA125** | 0.01 | 0.191 (0.12-0.28) | 0 | 50% |
| | 0.02 | 0.187 (0.12-0.27) | 0.004 | 74% |
| | 0.03 | 0.184 (0.11-0.27) | 0.0095 | 86% |
| | 0.04 | 0.181 (0.11-0.27) | 0.0156 | 93% |
| | 0.05 | 0.180 (0.11-0.27) | 0.0228 | 96% |
| | **0.10** | **0.170 (0.10-0.25)** | **0.0594** | **97%** |
| | 0.15 | 0.159 (0.09-0.24) | 0.1018 | 98% |
| | 0.20 | 0.149 (0.08-0.23) | 0.1488 | 97% |
| | 0.25 | 0.142 (0.08-0.22) | 0.1417 | 97% |
| | 0.30 | 0.135 (0.07-0.21) | 0.135 | 97% |
| | 0.35 | 0.130 (0.07-0.21) | 0.13 | 97% |
| | 0.40 | 0.121 (0.06-0.19) | 0.1207 | 97% |
| **ADNEX2** | 0.01 | 0.192 (0.12-0.29) | 0.0004 | 61% |
| | 0.02 | 0.186 (0.11-0.27) | 0.0041 | 77% |
| | 0.03 | 0.183 (0.11-0.27) | 0.0094 | 88% |
| | 0.04 | 0.181 (0.11-0.27) | 0.0161 | 94% |
| | 0.05 | 0.180 (0.11-0.27) | 0.0232 | 96% |
| | **0.10** | **0.170 (0.10-0.26)** | **0.0593** | **97%** |
| | 0.15 | 0.163 (0.09-0.25) | 0.1042 | 97% |
| | 0.20 | 0.154 (0.09-0.24) | 0.1537 | 96% |
| | 0.25 | 0.146 (0.08-0.23) | 0.1457 | 96% |
| | 0.30 | 0.138 (0.07-0.22) | 0.1385 | 96% |

| Strategy | Threshold | NB (95% Crl) | Delta NB | P-useful |
|---|---|---|---|---|
| | 0.35 | 0.130 (0.07-0.21) | 0.1304 | 95% |
| | 0.40 | 0.126 (0.07-0.20) | 0.1256 | 96% |
| **ADNEX2 ignoring CA125** | 0.01 | 0.191 (0.12-0.29) | 0.0002 | 54% |
| | 0.02 | 0.186 (0.11-0.27) | 0.0035 | 76% |
| | 0.03 | 0.183 (0.11-0.27) | 0.0092 | 86% |
| | 0.04 | 0.181 (0.11-0.27) | 0.016 | 93% |
| | 0.05 | 0.179 (0.11-0.27) | 0.0229 | 96% |
| | **0.10** | **0.170 (0.10-0.26)** | **0.0596** | **98%** |
| | 0.15 | 0.161 (0.09-0.25) | 0.1033 | 97% |
| | 0.20 | 0.153 (0.09-0.24) | 0.1528 | 97% |
| | 0.25 | 0.145 (0.08-0.23) | 0.1447 | 96% |
| | 0.30 | 0.137 (0.07-0.22) | 0.1371 | 95% |
| | 0.35 | 0.130 (0.07-0.21) | 0.13 | 95% |
| | 0.40 | 0.126 (0.07-0.20) | 0.1263 | 95% |
| **ADNEX** | 0.01 | 0.191 (0.12-0.28) | 0.0001 | 52% |
| | 0.02 | 0.185 (0.11-0.27) | 0.0032 | 72% |
| | 0.03 | 0.183 (0.11-0.27) | 0.0087 | 87% |
| | 0.04 | 0.182 (0.11-0.27) | 0.016 | 94% |
| | 0.05 | 0.180 (0.11-0.27) | 0.0234 | 97% |
| | **0.10** | **0.170 (0.10-0.26)** | **0.0593** | **98%** |
| | 0.15 | 0.160 (0.09-0.24) | 0.102 | 97% |
| | 0.20 | 0.150 (0.08-0.23) | 0.15 | 97% |
| | 0.25 | 0.141 (0.08-0.22) | 0.1411 | 97% |
| | 0.30 | 0.137 (0.08-0.22) | 0.1366 | 97% |
| | 0.35 | 0.131 (0.07-0.21) | 0.131 | 98% |
| | 0.40 | 0.127 (0.07-0.20) | 0.1266 | 98% |
| **ADNEX ignoring CA125** | 0.01 | 0.191 (0.12-0.29) | 0.0001 | 51% |
| | 0.02 | 0.185 (0.11-0.27) | 0.0027 | 69% |
| | 0.03 | 0.183 (0.11-0.27) | 0.0088 | 86% |
| | 0.04 | 0.181 (0.11-0.27) | 0.0151 | 92% |
| | 0.05 | 0.180 (0.11-0.27) | 0.0228 | 96% |
| | **0.10** | **0.170 (0.10-0.26)** | **0.0594** | **97%** |
| | 0.15 | 0.159 (0.09-0.24) | 0.1018 | 98% |
| | 0.20 | 0.149 (0.08-0.23) | 0.1487 | 97% |
| | 0.25 | 0.142 (0.08-0.22) | 0.1418 | 97% |
| | 0.30 | 0.135 (0.07-0.22) | 0.1351 | 97% |
| | 0.35 | 0.130 (0.07-0.21) | 0.1299 | 97% |
| | 0.40 | 0.121 (0.06-0.19) | 0.1206 | 97% |
| **BDs do not apply (N = 4657)** | | | | |
| **ADNEX2** | 0.01 | 0.301 (0.20-0.41) | -0.0003 | 39% |
| | 0.02 | 0.294 (0.19-0.41) | 0.0012 | 62% |
| | 0.03 | 0.289 (0.19-0.40) | 0.0032 | 70% |
| | 0.04 | 0.287 (0.19-0.40) | 0.0074 | 83% |
| | 0.05 | 0.284 (0.18-0.40) | 0.0122 | 89% |
| | **0.10** | **0.267 (0.17-0.38)** | **0.0364** | **91%** |
| | 0.15 | 0.256 (0.15-0.37) | 0.0701 | 92% |
| | 0.20 | 0.241 (0.14-0.36) | 0.1069 | 92% |
| | 0.25 | 0.229 (0.13-0.34) | 0.1513 | 93% |
| | 0.30 | 0.217 (0.12-0.33) | 0.206 | 93% |
| | 0.35 | 0.204 (0.11-0.31) | 0.2045 | 92% |
| | 0.40 | 0.197 (0.11-0.30) | 0.1972 | 94% |
| **ADNEX2 ignoring CA125** | 0.01 | 0.300 (0.20-0.41) | -0.0005 | 34% |
| | 0.02 | 0.294 (0.20-0.40) | 0.0007 | 58% |
| | 0.03 | 0.289 (0.19-0.40) | 0.0032 | 69% |
| | 0.04 | 0.286 (0.19-0.40) | 0.0074 | 83% |
| | 0.05 | 0.283 (0.18-0.39) | 0.0116 | 88% |
| | **0.10** | **0.267 (0.17-0.38)** | **0.0365** | **92%** |
| | 0.15 | 0.253 (0.15-0.37) | 0.0679 | 93% |
| | 0.20 | 0.240 (0.14-0.35) | 0.1052 | 92% |
| | 0.25 | 0.227 (0.13-0.34) | 0.1502 | 93% |
| | 0.30 | 0.215 (0.12-0.33) | 0.2033 | 93% |
| | 0.35 | 0.204 (0.11-0.31) | 0.204 | 92% |
| | 0.40 | 0.198 (0.11-0.30) | 0.1979 | 93% |
| **ADNEX** | 0.01 | 0.300 (0.20-0.41) | -0.0006 | 28% |
| | 0.02 | 0.294 (0.19-0.40) | 0.0001 | 50% |
| | 0.03 | 0.290 (0.19-0.40) | 0.0026 | 66% |

| Strategy | Threshold | NB (95% Crl) | Delta NB | P-useful |
|---|---|---|---|---|
| | 0.04 | 0.287 (0.19-0.40) | 0.0076 | 82% |
| | 0.05 | 0.285 (0.19-0.39) | 0.0124 | 88% |
| | **0.10** | **0.268 (0.17-0.38)** | **0.0364** | **90%** |
| | 0.15 | 0.251 (0.15-0.36) | 0.0659 | 93% |
| | 0.20 | 0.236 (0.14-0.35) | 0.1004 | 94% |
| | 0.25 | 0.222 (0.13-0.33) | 0.144 | 94% |
| | 0.30 | 0.214 (0.12-0.32) | 0.2025 | 94% |
| | 0.35 | 0.206 (0.12-0.31) | 0.2057 | 95% |
| | 0.40 | 0.198 (0.12-0.29) | 0.1983 | 96% |
| **ADNEX ignoring CA125** | 0.01 | 0.300 (0.20-0.42) | -0.0005 | 32% |
| | 0.02 | 0.294 (0.19-0.40) | -0.0004 | 44% |
| | 0.03 | 0.291 (0.19-0.40) | 0.0029 | 66% |
| | 0.04 | 0.287 (0.19-0.39) | 0.0066 | 79% |
| | 0.05 | 0.283 (0.19-0.39) | 0.0116 | 88% |
| | **0.10** | **0.268 (0.17-0.38)** | **0.0367** | **90%** |
| | 0.15 | 0.251 (0.15-0.36) | 0.0655 | 93% |
| | 0.20 | 0.234 (0.14-0.34) | 0.0989 | 93% |
| | 0.25 | 0.223 (0.13-0.33) | 0.1445 | 93% |
| | 0.30 | 0.212 (0.12-0.32) | 0.1988 | 94% |
| | 0.35 | 0.204 (0.12-0.30) | 0.2042 | 95% |
| | 0.40 | 0.189 (0.11-0.29) | 0.1892 | 95% |

BDs: Benign Descriptors; NB: Net Benefit; Delta NB: Model net benefit minus the NB of the best default strategy.

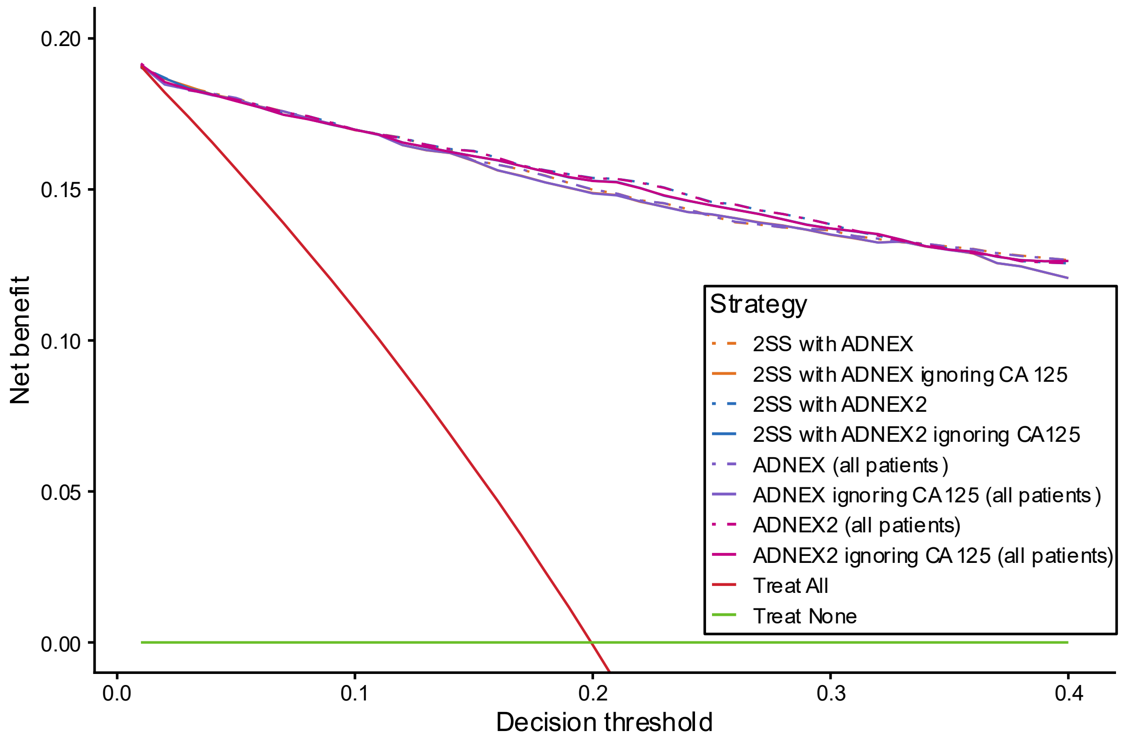

**Figure S45. Decision curves for ADNEX2 and ADNEX alone or in a two-step strategy in all patients (n = 6847). 2SS: Two step strategy.**

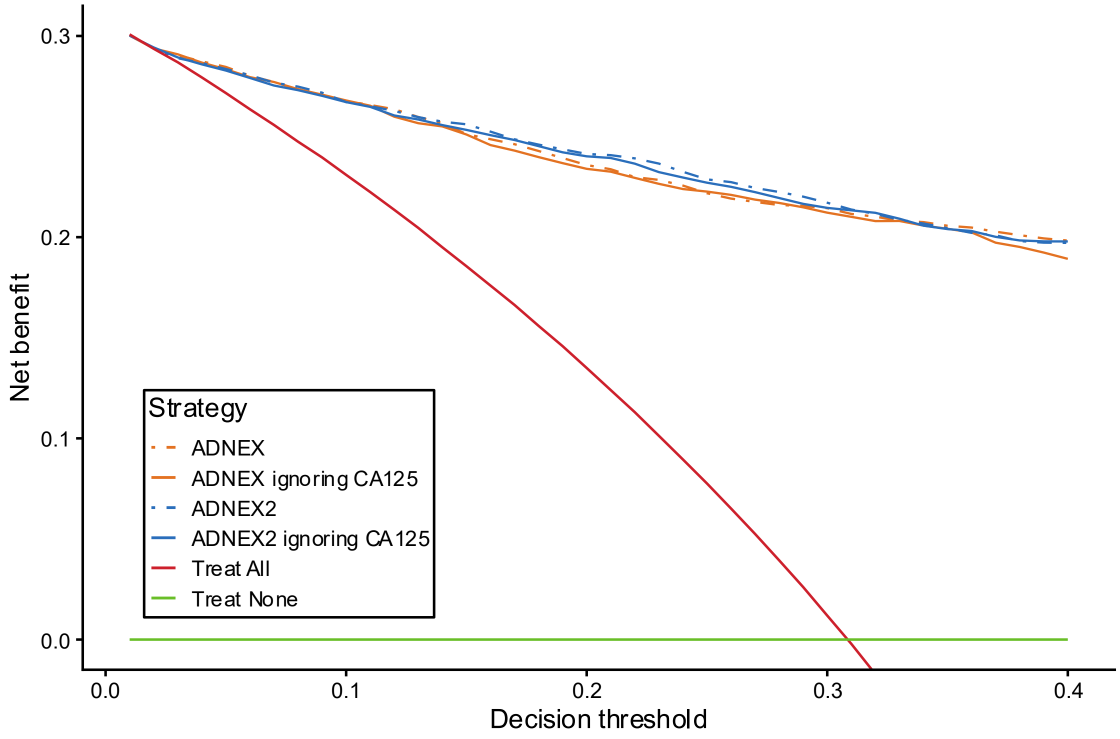

**Figure S46. Decision curves for ADNEX2 and ADNEX in patients where benign descriptors do not apply (n = 4657).**

### S5. SUBGROUP RESULTS

#### S5.1 DESCRIPTIVE STATISTICS

**Table S16. Characteristics of patients and tumours in subgroups**

| Variable | Surgery within 120 days | Non-Oncology center | Oncology center | Postmenopausal | Premenopausal |
|---|---|---|---|---|---|
| Number of cases | 3679 | 2472 | 4375 | 3302 | 3545 |
| Benign | 2028 (55%) | 2040 (83%) | 2585 (59%) | 1945 (59%) | 2680 (76%) |
| Borderline | 368 (10%) | 85 (3.4%) | 306 (7.0%) | 153 (4.6%) | 238 (6.7%) |
| Stage I invasive cancer | 295 (8.0%) | 39 (1.6%) | 264 (6.0%) | 183 (5.5%) | 120 (3.4%) |
| Stage II-IV invasive cancer | 769 (21%) | 94 (3.8%) | 693 (16%) | 603 (18%) | 184 (5.2%) |
| Secondary Metastasis | 218 (5.9%) | 45 (1.8%) | 184 (4.2%) | 159 (4.8%) | 70 (2.0%) |
| Uncertain | 1 (<0.1%) | 169 (6.8%) | 343 (7.8%) | 259 (7.8%) | 253 (7.1%) |
| Surgery within 120 days after first scan | 3679 (100%) | 1075 (43%) | 2604 (60%) | 1937 (59%) | 1742 (49%) |
| Patient age at recruitment (years) | 52 (39-64) | 48 (35-64) | 50 (38-63) | 64 (57-71) | 38 (30-45) |
| Postmenopausal | 1937 (53%) | 1118 (45%) | 2184 (50%) | 3302 (100%) | 0 (0%) |
| Pelvic pain | 577 (16%) | 419 (17%) | 569 (13%) | 297 (9.0%) | 691 (19%) |
| Presence of solid components | 2162 (59%) | 705 (29%) | 2096 (48%) | 1767 (54%) | 1034 (29%) |
| Proportion of solid components | 0.25 (0.00-0.80) | 0.00 (0.00-0.16) | 0.00 (0.00-0.64) | 0.16 (0.00-0.78) | 0.00 (0.00-0.21) |
| Maximum diameter of the lesion (mm) | 74 (51-111) | 54 (40-77) | 58 (38-92) | 59 (40-94) | 54 (37-81) |
| CA125 (U/ml) | 36 (14-194) | 18 (10-39) | 35 (13-177) | 29 (11-189) | 26 (13-76) |
| CA125 missing | 946 (26%) | 1514 (61%) | 1712 (39%) | 1297 (39%) | 1929 (54%) |
| Shadows | 698 (19%) | 539 (22%) | 765 (17%) | 656 (20%) | 648 (18%) |
| Ascites | 450 (12%) | 64 (2.6%) | 426 (9.7%) | 359 (11%) | 131 (3.7%) |
| Number of papillations | | | | | |
| None | 2968 (81%) | 2180 (88%) | 3687 (84%) | 2769 (84%) | 3098 (87%) |
| 1 | 291 (7.9%) | 131 (5.3%) | 333 (7.6%) | 240 (7.3%) | 224 (6.3%) |
| 2 | 97 (2.6%) | 57 (2.3%) | 86 (2.0%) | 98 (3.0%) | 45 (1.3%) |
| 3 | 71 (1.9%) | 38 (1.5%) | 57 (1.3%) | 53 (1.6%) | 42 (1.2%) |
| 4 or more | 252 (6.8%) | 66 (2.7%) | 212 (4.8%) | 142 (4.3%) | 136 (3.8%) |
| More than 10 cyst locules | 541 (15%) | 163 (6.6%) | 471 (11%) | 396 (12%) | 238 (6.7%) |
| Colour score of intratumoral flow | | | | | |
| 1: No blood flow | 908 (25%) | 1173 (47%) | 1664 (38%) | 1259 (38%) | 1578 (45%) |
| 2: minimal blood flow | 966 (26%) | 603 (24%) | 1160 (27%) | 807 (24%) | 956 (27%) |
| 3: moderate blood flow | 1152 (31%) | 568 (23%) | 969 (22%) | 770 (23%) | 767 (22%) |
| 4: very strong flow | 653 (18%) | 128 (5.2%) | 582 (13%) | 466 (14%) | 244 (6.9%) |
| Tumour type using IOTA terminology | | | | | |
| Unilocular | 852 (23%) | 1173 (47%) | 1405 (32%) | 826 (25%) | 1752 (49%) |
| Unilocular solid | 423 (11%) | 183 (7.4%) | 447 (10%) | 275 (8.3%) | 355 (10%) |
| Multilocular | 649 (18%) | 573 (23%) | 865 (20%) | 698 (21%) | 740 (21%) |
| Multilocular solid | 922 (25%) | 279 (11%) | 832 (19%) | 746 (23%) | 365 (10%) |
| Solid | 817 (22%) | 243 (9.8%) | 817 (19%) | 746 (23%) | 314 (8.9%) |
| Not possible to classify | 16 (0.4%) | 21 (0.8%) | 9 (0.2%) | 11 (0.3%) | 19 (0.5%) |

Results are presented as median (IQR) for continuous variables, or N (%) for categorical variables.

#### S5.2 DISCRIMINATION

| Strategy | Sample size | Prevalence | AUC (95% CI) | 95% PI |
|---|---|---|---|---|
| 2SS with ADNEX2 (Postmenopausal) | 3302 | 35% | 0.948 (0.93 to 0.96) | (0.88 to 0.99) |
| 2SS with ADNEX2 ignoring CA125 (Postmenopausal) | 3302 | 35% | 0.938 (0.92 to 0.96) | (0.87 to 0.98) |
| 2SS with ADNEX (Postmenopausal) | 3302 | 35% | 0.947 (0.93 to 0.96) | (0.89 to 0.99) |
| 2SS with ADNEX ignoring CA125 (Postmenopausal) | 3302 | 35% | 0.936 (0.92 to 0.95) | (0.87 to 0.98) |
| 2SS with ADNEX2 (Premenopausal) | 3545 | 18% | 0.939 (0.92 to 0.95) | (0.91 to 0.97) |
| 2SS with ADNEX2 ignoring CA125 (Premenopausal) | 3545 | 18% | 0.938 (0.92 to 0.95) | (0.91 to 0.97) |
| 2SS with ADNEX (Premenopausal) | 3545 | 18% | 0.941 (0.93 to 0.95) | (0.91 to 0.96) |
| 2SS with ADNEX ignoring CA125 (Premenopausal) | 3545 | 18% | 0.939 (0.92 to 0.95) | (0.91 to 0.96) |
| 2SS with ADNEX2 (Surgery within 120 days) | 3679 | 45% | 0.928 (0.91 to 0.94) | (0.89 to 0.97) |
| 2SS with ADNEX2 ignoring CA125 (Surgery within 120 days) | 3679 | 45% | 0.918 (0.90 to 0.93) | (0.87 to 0.96) |
| 2SS with ADNEX (Surgery within 120 days) | 3679 | 45% | 0.929 (0.92 to 0.94) | (0.89 to 0.96) |
| 2SS with ADNEX ignoring CA125 (Surgery within 120 days) | 3679 | 45% | 0.919 (0.90 to 0.93) | (0.88 to 0.96) |
| 2SS with ADNEX2 (Oncology center) | 4375 | 35% | 0.953 (0.94 to 0.96) | (0.93 to 0.98) |
| 2SS with ADNEX2 ignoring CA125 (Oncology center) | 4375 | 35% | 0.947 (0.93 to 0.96) | (0.92 to 0.97) |
| 2SS with ADNEX (Oncology center) | 4375 | 35% | 0.952 (0.94 to 0.96) | (0.93 to 0.97) |
| 2SS with ADNEX ignoring CA125 (Oncology center) | 4375 | 35% | 0.947 (0.94 to 0.96) | (0.92 to 0.97) |
| 2SS with ADNEX2 (Non-Oncology center) | 2472 | 12% | 0.922 (0.87 to 0.96) | (0.81 to 0.99) |
| 2SS with ADNEX2 ignoring CA125 (Non-Oncology center) | 2472 | 12% | 0.918 (0.87 to 0.95) | (0.81 to 0.99) |
| 2SS with ADNEX (Non-Oncology center) | 2472 | 12% | 0.921 (0.87 to 0.96) | (0.81 to 0.99) |
| 2SS with ADNEX ignoring CA125 (Non-Oncology center) | 2472 | 12% | 0.918 (0.87 to 0.95) | (0.82 to 0.99) |

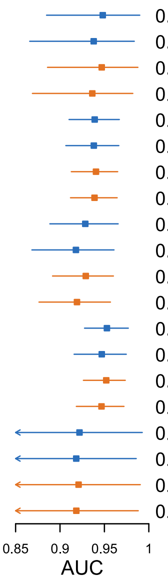

**Figure S47. Forest plot showing area under the receiver operating characteristic curve (AUC) for the two-step strategy with ADNEX2 (blue) or with ADNEX (orange) with and ignoring CA125 in subgroups. Results of meta-analysis are shown. Horizontal lines indicate 95% prediction interval (PI). CI, confidence interval; Prevalence means prevalence of malignancy**

| Centre | AUC (95% CI) | N |
|---|---|---|
| Malmö, Sweden | 0.89 (0.85 to 0.93) | 519 |
| Rome, Italy | 0.97 (0.94 to 0.98) | 492 |
| Leuven, Belgium | 0.95 (0.92 to 0.97) | 415 |
| Milan, Italy | 0.97 (0.94 to 0.99) | 323 |
| Stockholm, Sweden | 0.94 (0.90 to 0.96) | 302 |
| Prague, Czech Rep. | 0.97 (0.93 to 0.99) | 222 |
| Genk, Belgium | 0.93 (0.84 to 0.97) | 213 |
| Monza, Italy | 0.95 (0.89 to 0.98) | 156 |
| Trieste, Italy | 0.97 (0.89 to 0.99) | 145 |
| Milan 2, Italy | 0.97 (0.92 to 0.99) | 135 |
| Cagliari, Italy | 0.96 (0.87 to 0.99) | 97 |
| Milan 3, Italy | 0.55 (0.20 to 0.85) | 73 |
| Katowice, Poland | 0.91 (0.79 to 0.97) | 61 |
| Pamplona, Spain | 0.97 (0.87 to 1.00) | 56 |
| London, UK | 1.00 (0.43 to 1.00) | 42 |
| Florence, Italy | 1.00 (0.16 to 1.00) | 29 |
| Lisbon, Portugal | 0.97 (0.52 to 1.00) | 22 |
| **Meta-analysis** | **0.95 (0.93 to 0.96)** | **3302** |
| **AUC (95% PI)** | **(0.88 to 0.99)** | |

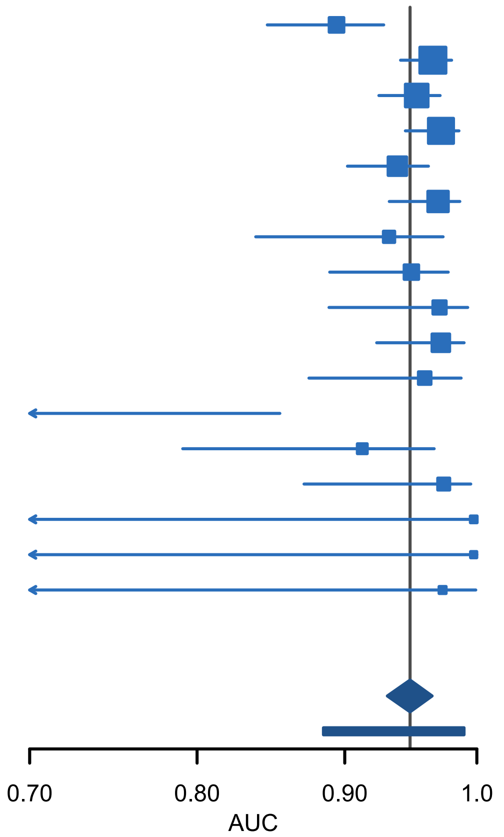

**Figure S48. Forest plot showing the area under the receiver operating characteristic curve (AUC) for two step strategy with ADNEX2 in postmenopausal patients (n = 3302). CI, confidence interval; PI, prediction interval**

| Centre | AUC (95% CI) | N |
|---|---|---|
| Malmö, Sweden | 0.88 (0.83 to 0.92) | 519 |
| Rome, Italy | 0.96 (0.93 to 0.97) | 492 |
| Leuven, Belgium | 0.95 (0.92 to 0.96) | 415 |
| Milan, Italy | 0.97 (0.94 to 0.98) | 323 |
| Stockholm, Sweden | 0.93 (0.89 to 0.96) | 302 |
| Prague, Czech Rep. | 0.96 (0.92 to 0.98) | 222 |
| Genk, Belgium | 0.93 (0.84 to 0.97) | 213 |
| Monza, Italy | 0.92 (0.85 to 0.95) | 156 |
| Trieste, Italy | 0.96 (0.88 to 0.99) | 145 |
| Milan 2, Italy | 0.97 (0.91 to 0.99) | 135 |
| Cagliari, Italy | 0.95 (0.86 to 0.98) | 97 |
| Milan 3, Italy | 0.52 (0.18 to 0.84) | 73 |
| Katowice, Poland | 0.90 (0.77 to 0.96) | 61 |
| Pamplona, Spain | 0.96 (0.86 to 0.99) | 56 |
| London, UK | 0.97 (0.44 to 1.00) | 42 |
| Florence, Italy | 1.00 (0.16 to 1.00) | 29 |
| Lisbon, Portugal | 0.95 (0.59 to 1.00) | 22 |
| **Meta-analysis** | **0.94 (0.92 to 0.96)** | **3302** |
| **AUC (95% PI)** | **(0.87 to 0.98)** | |

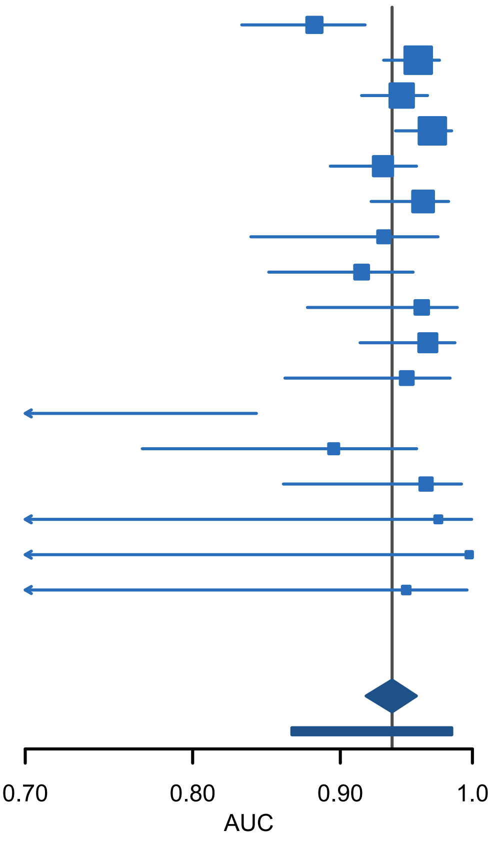

**Figure S49. Forest plot showing the area under the receiver operating characteristic curve (AUC) for two step strategy with ADNEX2 ignoring CA125 in postmenopausal patients (n = 3302). CI, confidence interval; PI, prediction interval**

| Centre | AUC (95% CI) | N |
|---|---|---|
| Malmö, Sweden | 0.96 (0.89 to 0.98) | 574 |
| Rome, Italy | 0.94 (0.91 to 0.96) | 527 |
| Leuven, Belgium | 0.96 (0.91 to 0.99) | 339 |
| Monza, Italy | 0.92 (0.86 to 0.95) | 275 |
| Genk, Belgium | 0.92 (0.80 to 0.97) | 260 |
| Milan, Italy | 0.97 (0.93 to 0.98) | 235 |
| Stockholm, Sweden | 0.93 (0.88 to 0.96) | 207 |
| Prague, Czech Rep. | 0.94 (0.88 to 0.97) | 192 |
| Katowice, Poland | 0.93 (0.78 to 0.98) | 180 |
| Cagliari, Italy | 0.98 (0.88 to 1.00) | 150 |
| London, UK | 0.97 (0.38 to 1.00) | 130 |
| Trieste, Italy | 0.93 (0.82 to 0.97) | 128 |
| Pamplona, Spain | 0.98 (0.79 to 1.00) | 114 |
| Milan 2, Italy | 0.94 (0.86 to 0.98) | 92 |
| Florence, Italy | 0.35 (0.05 to 0.84) | 56 |
| Milan 3, Italy | 0.98 (0.02 to 1.00) | 56 |
| Lisbon, Portugal | 1.00 (0.16 to 1.00) | 30 |
| **Meta-analysis** | **0.94 (0.92 to 0.95)** | **3545** |
| **AUC (95% PI)** | **(0.91 to 0.97)** | |

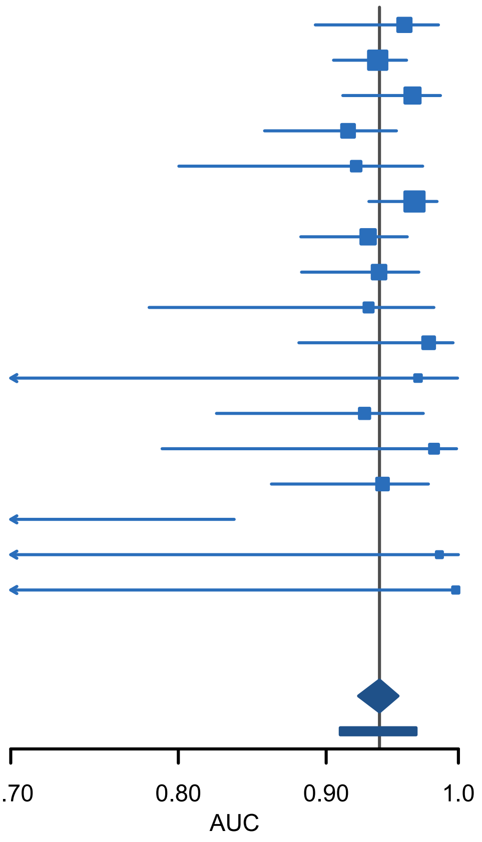

**Figure S50. Forest plot showing the area under the receiver operating characteristic curve (AUC) for two step strategy with ADNEX2 in premenopausal patients (n = 3545). CI, confidence interval; PI, prediction interval**

| Centre | AUC (95% CI) | N |
|---|---|---|
| Malmö, Sweden | 0.96 (0.89 to 0.99) | 574 |
| Rome, Italy | 0.94 (0.90 to 0.96) | 527 |
| Leuven, Belgium | 0.97 (0.92 to 0.99) | 339 |
| Monza, Italy | 0.91 (0.85 to 0.95) | 275 |
| Genk, Belgium | 0.93 (0.80 to 0.97) | 260 |
| Milan, Italy | 0.96 (0.92 to 0.98) | 235 |
| Stockholm, Sweden | 0.93 (0.88 to 0.96) | 207 |
| Prague, Czech Rep. | 0.94 (0.89 to 0.97) | 192 |
| Katowice, Poland | 0.93 (0.78 to 0.98) | 180 |
| Cagliari, Italy | 0.97 (0.87 to 0.99) | 150 |
| London, UK | 0.96 (0.45 to 1.00) | 130 |
| Trieste, Italy | 0.93 (0.82 to 0.97) | 128 |
| Pamplona, Spain | 0.98 (0.79 to 1.00) | 114 |
| Milan 2, Italy | 0.94 (0.86 to 0.98) | 92 |
| Florence, Italy | 0.35 (0.05 to 0.84) | 56 |
| Milan 3, Italy | 0.98 (0.02 to 1.00) | 56 |
| Lisbon, Portugal | 1.00 (0.14 to 1.00) | 30 |
| **Meta-analysis** | **0.94 (0.92 to 0.95)** | **3545** |
| **AUC (95% PI)** | **(0.91 to 0.97)** | |

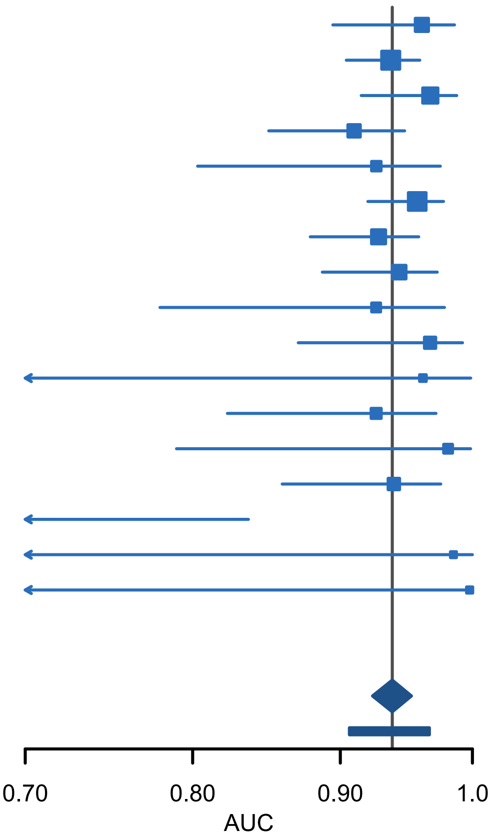

**Figure S51. Forest plot showing the area under the receiver operating characteristic curve (AUC) for two step strategy with ADNEX2 ignoring CA125 in premenopausal patients (n = 3545). CI, confidence interval; PI, prediction interval**

| Centre | AUC (95% CI) | N |
|---|---|---|
| Rome, Italy | 0.93 (0.90 to 0.95) | 545 |
| Milan, Italy | 0.96 (0.94 to 0.98) | 449 |
| Malmö, Sweden | 0.89 (0.85 to 0.93) | 429 |
| Leuven, Belgium | 0.93 (0.90 to 0.95) | 406 |
| Stockholm, Sweden | 0.91 (0.88 to 0.94) | 359 |
| Prague, Czech Rep. | 0.93 (0.89 to 0.96) | 248 |
| Genk, Belgium | 0.95 (0.90 to 0.97) | 246 |
| Monza, Italy | 0.93 (0.89 to 0.96) | 241 |
| Cagliari, Italy | 0.97 (0.91 to 0.99) | 185 |
| Milan 2, Italy | 0.92 (0.85 to 0.96) | 145 |
| Trieste, Italy | 0.91 (0.84 to 0.96) | 118 |
| Pamplona, Spain | 0.98 (0.92 to 1.00) | 111 |
| Katowice, Poland | 0.92 (0.82 to 0.96) | 81 |
| Milan 3, Italy | 0.76 (0.37 to 0.94) | 52 |
| Florence, Italy | 0.66 (0.25 to 0.92) | 28 |
| Lisbon, Portugal | 0.94 (0.35 to 1.00) | 19 |
| London, UK | 0.97 (0.60 to 1.00) | 17 |
| **Meta-analysis** | **0.93 (0.91 to 0.94)** | **3679** |
| **AUC (95% PI)** | **(0.89 to 0.97)** | |

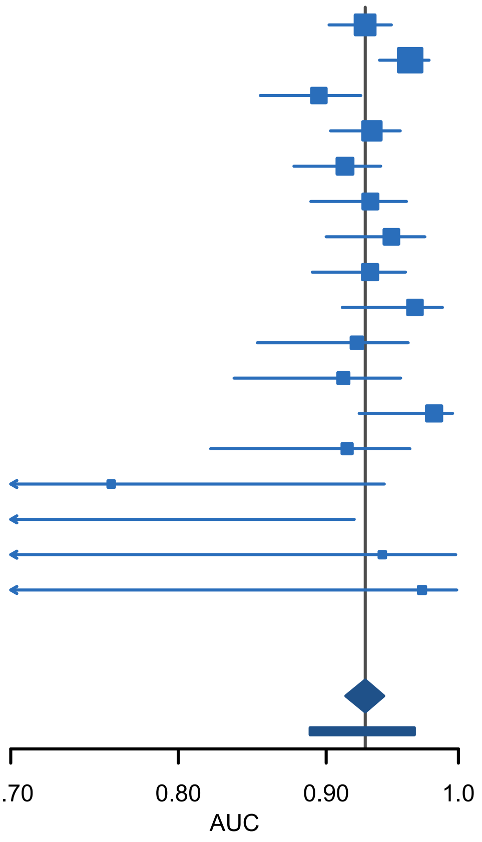

**Figure S52. Forest plot showing the area under the receiver operating characteristic curve (AUC) for two step strategy with ADNEX2 in patients operated within 120 days after inclusion scan (n = 3679). CI, confidence interval; PI, prediction interval**

| Centre | AUC (95% CI) | N |
|---|---|---|
| Rome, Italy | 0.92 (0.89 to 0.94) | 545 |
| Milan, Italy | 0.96 (0.93 to 0.97) | 449 |
| Malmö, Sweden | 0.88 (0.84 to 0.92) | 429 |
| Leuven, Belgium | 0.93 (0.89 to 0.95) | 406 |
| Stockholm, Sweden | 0.90 (0.86 to 0.93) | 359 |
| Prague, Czech Rep. | 0.93 (0.88 to 0.96) | 248 |
| Genk, Belgium | 0.95 (0.90 to 0.97) | 246 |
| Monza, Italy | 0.90 (0.86 to 0.94) | 241 |
| Cagliari, Italy | 0.95 (0.90 to 0.98) | 185 |
| Milan 2, Italy | 0.91 (0.83 to 0.95) | 145 |
| Trieste, Italy | 0.90 (0.82 to 0.95) | 118 |
| Pamplona, Spain | 0.97 (0.91 to 0.99) | 111 |
| Katowice, Poland | 0.89 (0.79 to 0.94) | 81 |
| Milan 3, Italy | 0.75 (0.37 to 0.94) | 52 |
| Florence, Italy | 0.66 (0.25 to 0.92) | 28 |
| Lisbon, Portugal | 0.94 (0.35 to 1.00) | 19 |
| London, UK | 0.93 (0.62 to 0.99) | 17 |
| **Meta-analysis** | **0.92 (0.90 to 0.93)** | **3679** |
| **AUC (95% PI)** | **(0.87 to 0.96)** | |

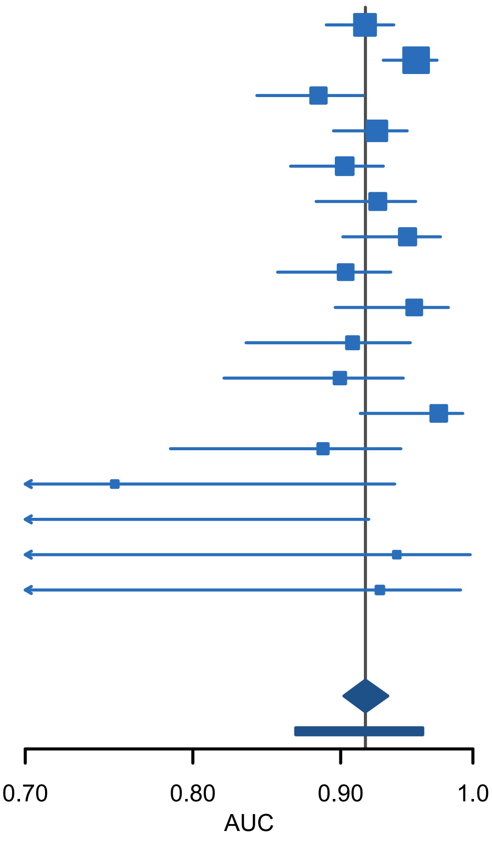

**Figure S53. Forest plot showing the area under the receiver operating characteristic curve (AUC) for two step strategy with ADNEX2 ignoring CA125 in patients operated within 120 days after inclusion scan (n = 3679). CI, confidence interval; PI, prediction interval**

| Centre | AUC (95% CI) | N |
|---|---|---|
| Rome, Italy | 0.95 (0.93 to 0.97) | 1019 |
| Leuven, Belgium | 0.96 (0.94 to 0.97) | 754 |
| Milan, Italy | 0.97 (0.95 to 0.98) | 558 |
| Stockholm, Sweden | 0.93 (0.90 to 0.95) | 509 |
| Monza, Italy | 0.94 (0.90 to 0.96) | 431 |
| Prague, Czech Rep. | 0.96 (0.93 to 0.97) | 414 |
| Katowice, Poland | 0.94 (0.86 to 0.97) | 241 |
| Milan 2, Italy | 0.96 (0.92 to 0.98) | 227 |
| Pamplona, Spain | 0.99 (0.93 to 1.00) | 170 |
| Lisbon, Portugal | 0.98 (0.54 to 1.00) | 52 |
| **Meta-analysis** | **0.95 (0.94 to 0.96)** | **4375** |
| **AUC (95% PI)** | **(0.93 to 0.98)** | |

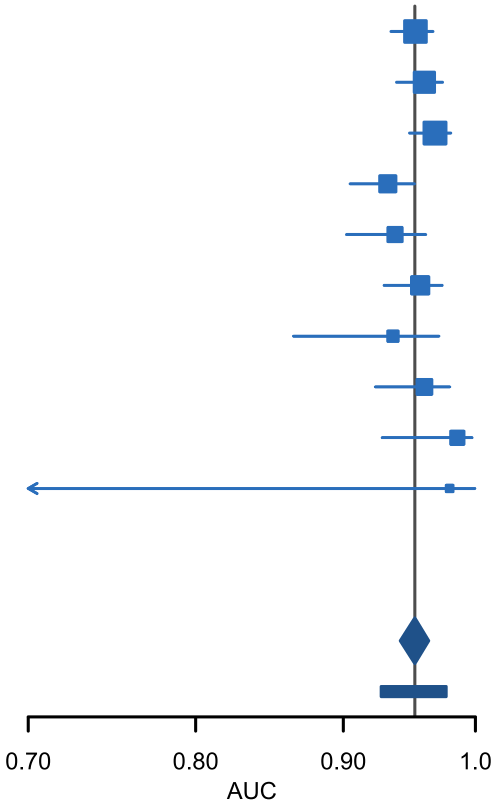

**Figure S54. Forest plot showing the area under the receiver operating characteristic curve (AUC) for two step strategy with ADNEX2 in oncology centres (n = 4375). CI, confidence interval; PI, prediction interval**

| Centre | AUC (95% CI) | N |
|---|---|---|
| Rome, Italy | 0.95 (0.93 to 0.96) | 1019 |
| Leuven, Belgium | 0.96 (0.94 to 0.97) | 754 |
| Milan, Italy | 0.96 (0.94 to 0.98) | 558 |
| Stockholm, Sweden | 0.92 (0.90 to 0.95) | 509 |
| Monza, Italy | 0.92 (0.89 to 0.95) | 431 |
| Prague, Czech Rep. | 0.95 (0.93 to 0.97) | 414 |
| Katowice, Poland | 0.93 (0.86 to 0.97) | 241 |
| Milan 2, Italy | 0.95 (0.92 to 0.98) | 227 |
| Pamplona, Spain | 0.98 (0.93 to 1.00) | 170 |
| Lisbon, Portugal | 0.97 (0.60 to 1.00) | 52 |
| **Meta-analysis** | **0.95 (0.93 to 0.96)** | **4375** |
| **AUC (95% PI)** | **(0.92 to 0.97)** | |

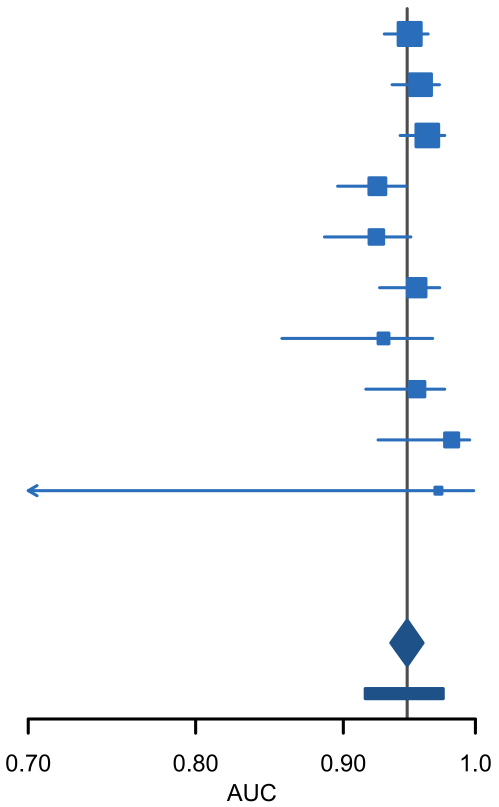

**Figure S55. Forest plot showing the area under the receiver operating characteristic curve (AUC) for two step strategy with ADNEX2 ignoring CA125 in oncology centres (n = 4375). CI, confidence interval; PI, prediction interval**

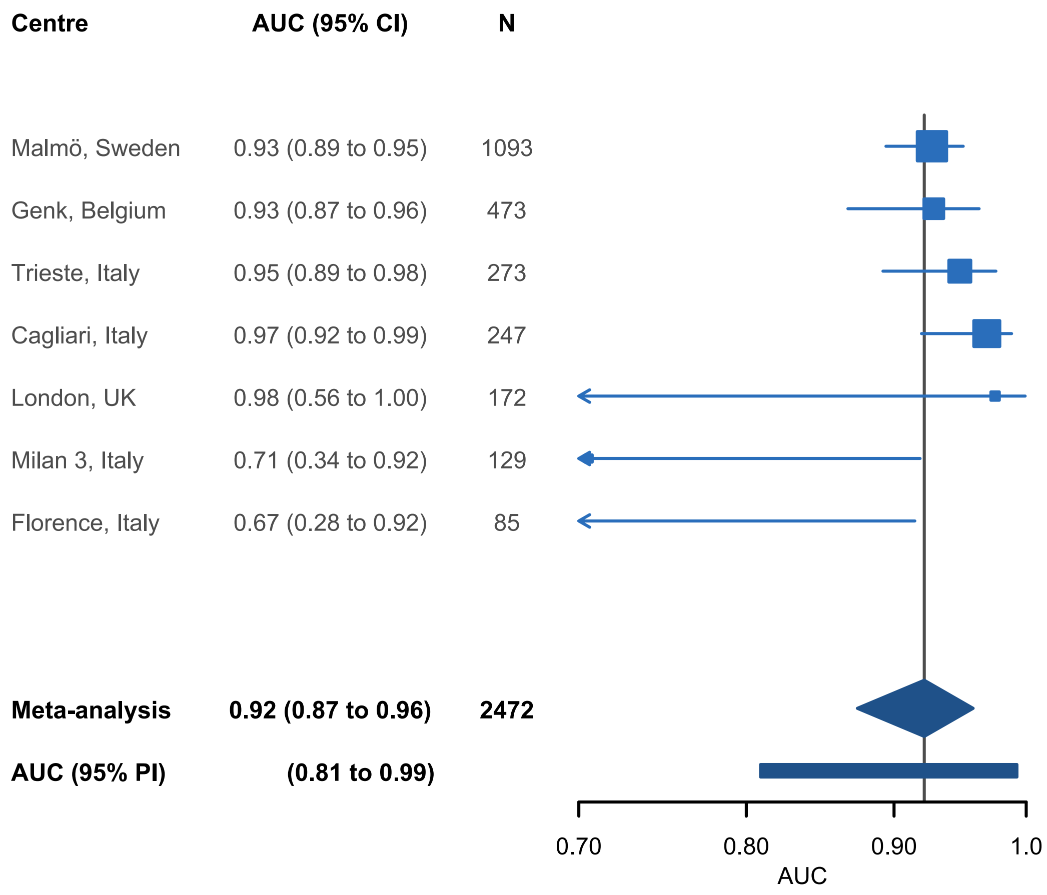

**Figure S56. Forest plot showing the area under the receiver operating characteristic curve (AUC) for two step strategy with ADNEX2 in non-oncology centres (n = 2472). CI, confidence interval; PI, prediction interval**

| Centre | AUC (95% CI) | N |
|---|---|---|
| Malmö, Sweden | 0.92 (0.89 to 0.95) | 1093 |
| Genk, Belgium | 0.93 (0.87 to 0.96) | 473 |
| Trieste, Italy | 0.94 (0.89 to 0.97) | 273 |
| Cagliari, Italy | 0.96 (0.91 to 0.98) | 247 |
| London, UK | 0.96 (0.65 to 1.00) | 172 |
| Milan 3, Italy | 0.70 (0.33 to 0.91) | 129 |
| Florence, Italy | 0.67 (0.28 to 0.91) | 85 |
| **Meta-analysis** | **0.92 (0.87 to 0.95)** | **2472** |
| **AUC (95% PI)** | **(0.81 to 0.99)** | |

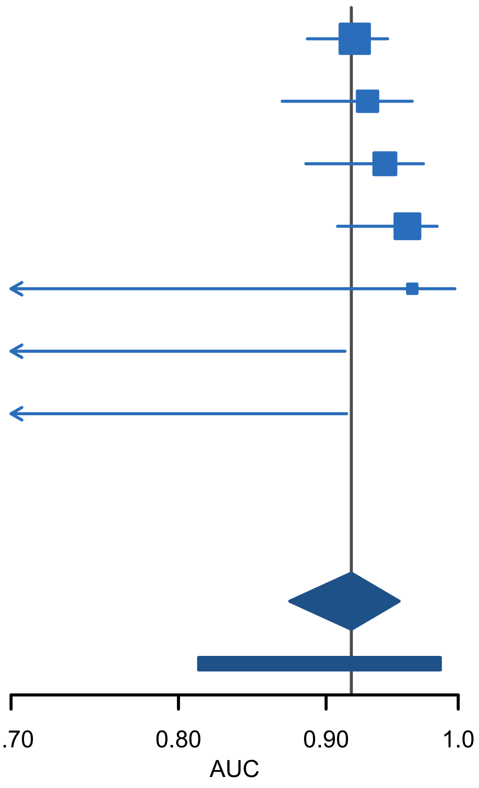

**Figure S57. Forest plot showing the area under the receiver operating characteristic curve (AUC) for two step strategy with ADNEX2 ignoring CA125 in non-oncology centres (n = 2472). CI, confidence interval; PI, prediction interval**

**Table S17. Pairwise AUCs in patients who underwent surgery within 120 days of the inclusion scan (n = 3679) for the two-step strategies (2SS) with ADNEX2 and ADNEX.**

| Pair | 2SS with ADNEX2 | 2SS with ADNEX2 ignoring CA125 | 2SS with ADNEX | 2SS with ADNEX ignoring CA125 |
|---|---|---|---|---|
| Benign vs Borderline | 0.88 (0.86-0.90) | 0.87 (0.85-0.89) | 0.88 (0.86-0.90) | 0.88 (0.85-0.89) |
| Benign vs Stage I | 0.93 (0.91-0.94) | 0.93 (0.91-0.94) | 0.93 (0.91-0.94) | 0.93 (0.91-0.94) |
| Benign vs Stage II-IV | 0.98 (0.97-0.98) | 0.96 (0.95-0.97) | 0.98 (0.97-0.98) | 0.96 (0.95-0.97) |
| Benign vs Metastatic | 0.94 (0.92-0.96) | 0.94 (0.92-0.95) | 0.95 (0.93-0.96) | 0.94 (0.92-0.96) |
| Borderline vs Stage I | 0.77 (0.74-0.81) | 0.78 (0.74-0.81) | 0.75 (0.71-0.78) | 0.76 (0.72-0.79) |
| Borderline vs Stage II-IV | 0.92 (0.90-0.94) | 0.90 (0.88-0.92) | 0.91 (0.89-0.93) | 0.90 (0.87-0.91) |
| Borderline vs Metastatic | 0.87 (0.83-0.90) | 0.87 (0.83-0.90) | 0.87 (0.83-0.89) | 0.87 (0.83-0.90) |
| Stage I vs Stage II-IV | 0.81 (0.78-0.84) | 0.73 (0.70-0.76) | 0.82 (0.79-0.85) | 0.73 (0.70-0.77) |
| Stage I vs Metastatic | 0.73 (0.69-0.77) | 0.73 (0.69-0.78) | 0.72 (0.67-0.76) | 0.72 (0.67-0.76) |
| Stage II-IV vs Metastatic | 0.80 (0.77-0.83) | 0.65 (0.61-0.69) | 0.81 (0.77-0.84) | 0.66 (0.62-0.70) |

#### S5.3 Calibration

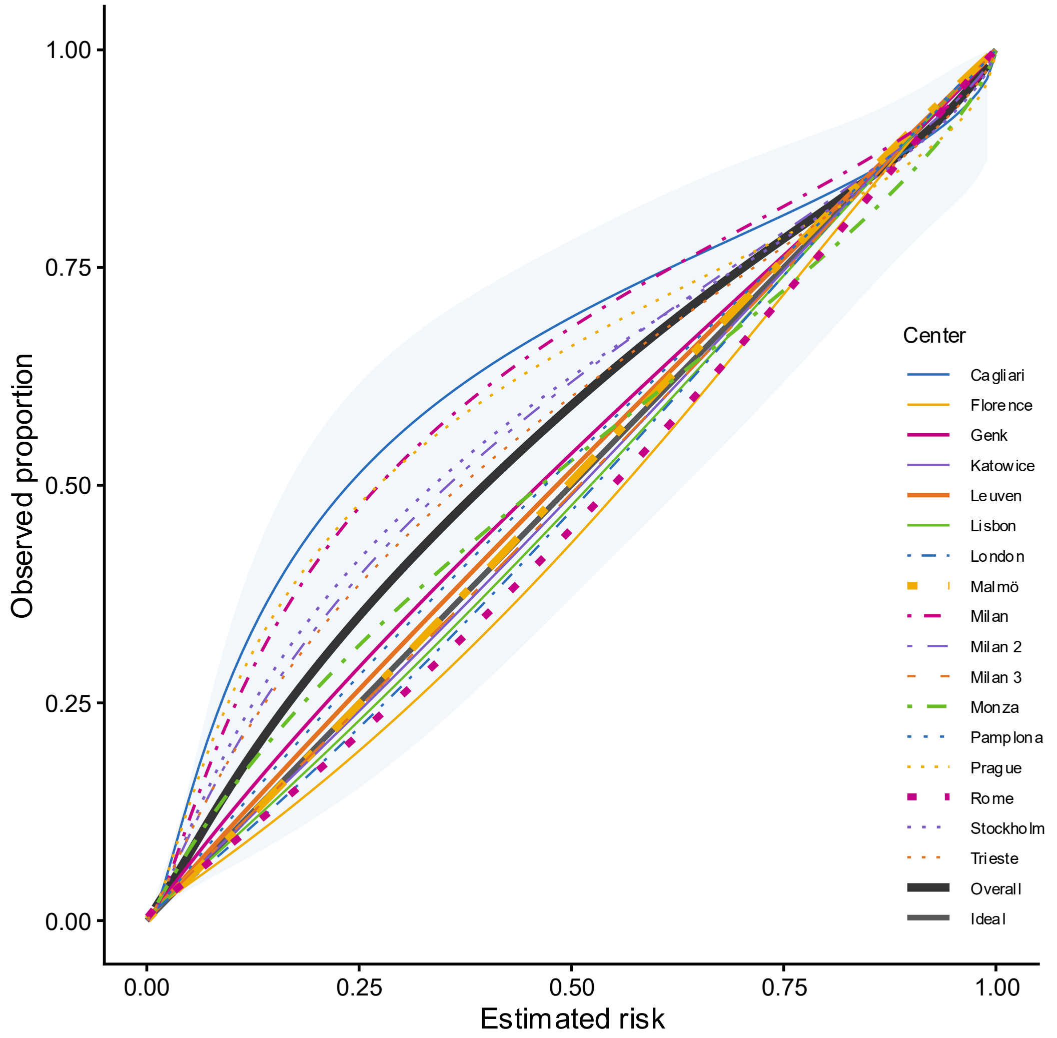

**Figure S58. Calibration plot for two step strategy with ADNEX2 in postmenopausal patients (n = 3302). Overall calibration curves calculated taking clustering into account and shaded area indicating 95% prediction interval. Line width proportional to sample size.**

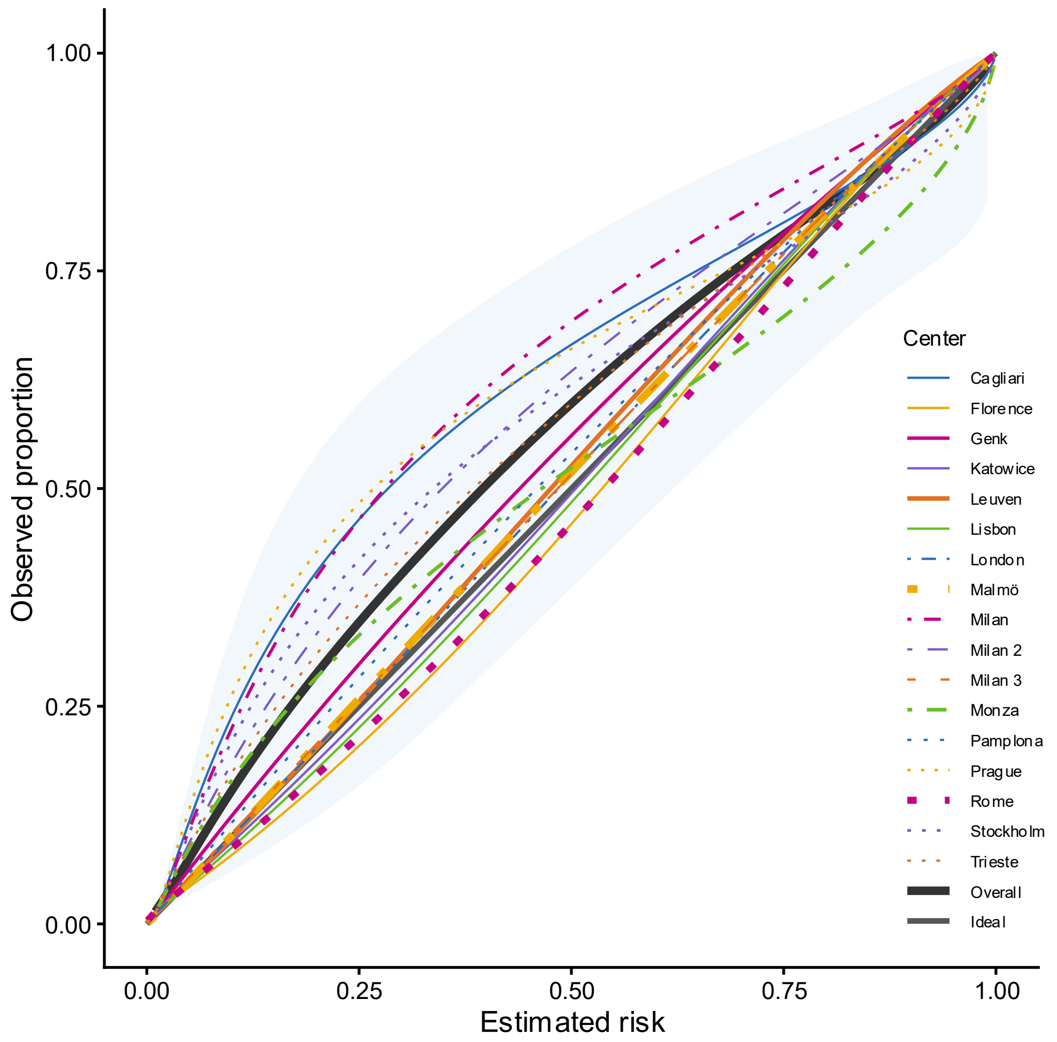

**Figure S59. Calibration plot for two step strategy with ADNEX2 ignoring CA125 in postmenopausal patients (n = 3302). Overall calibration curves calculated taking clustering into account and shaded area indicating 95% prediction interval. Line width proportional to sample size.**

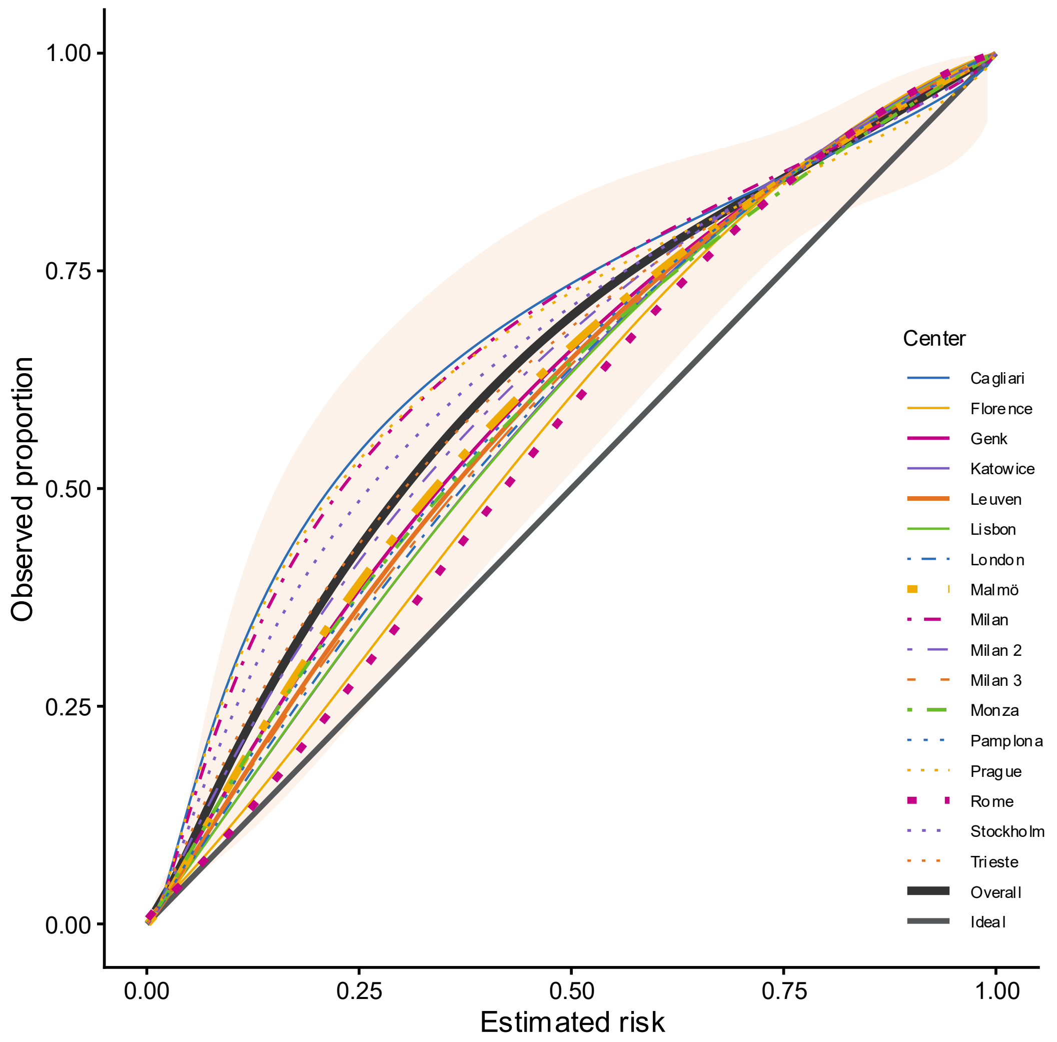

**Figure S60. Calibration plot for two step strategy with ADNEX in postmenopausal patients (n = 3302). Overall calibration curves calculated taking clustering into account and shaded area indicating 95% prediction interval. Line width proportional to sample size.**

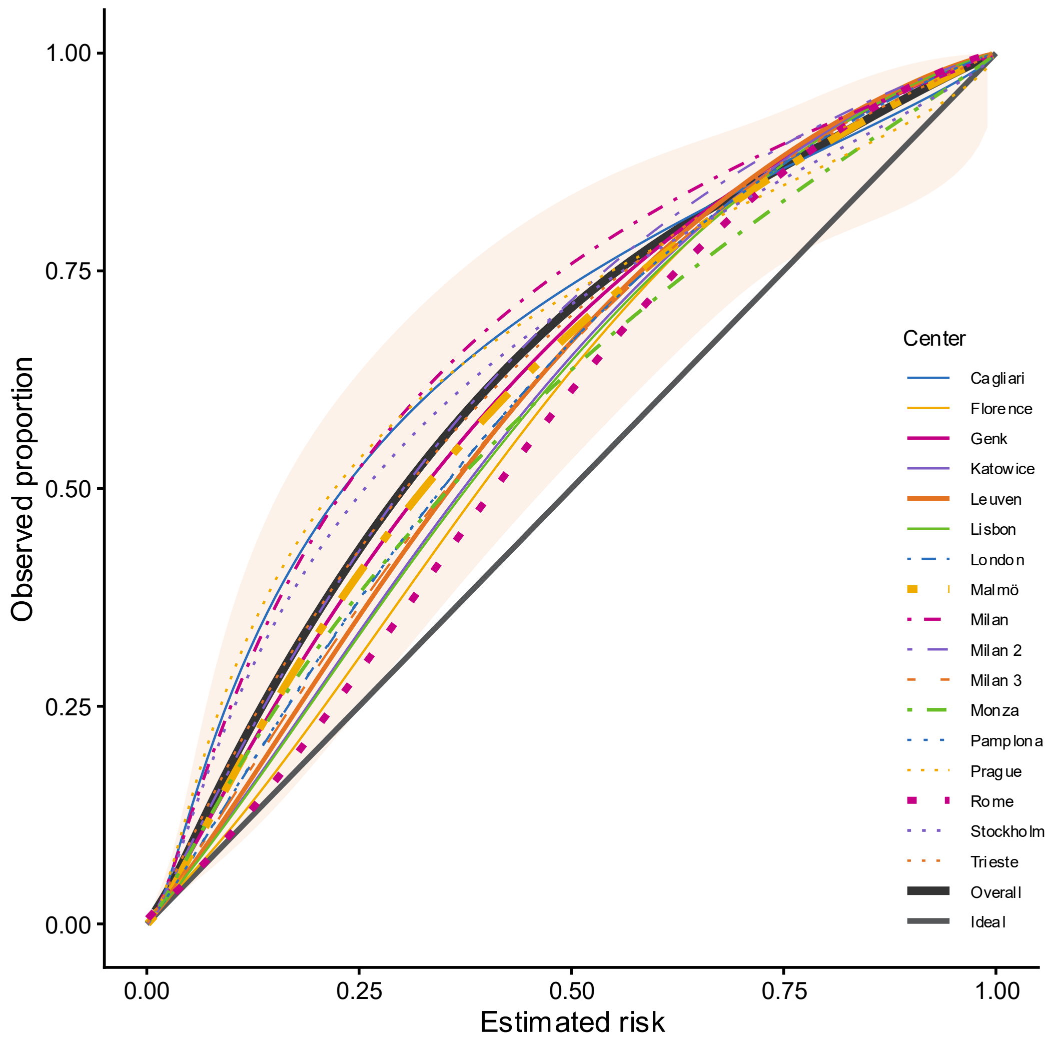

**Figure S61. Calibration plot for two step strategy with ADNEX ignoring CA125 in postmenopausal patients (n = 3302). Overall calibration curves calculated taking clustering into account and shaded area indicating 95% prediction interval. Line width proportional to sample size.**

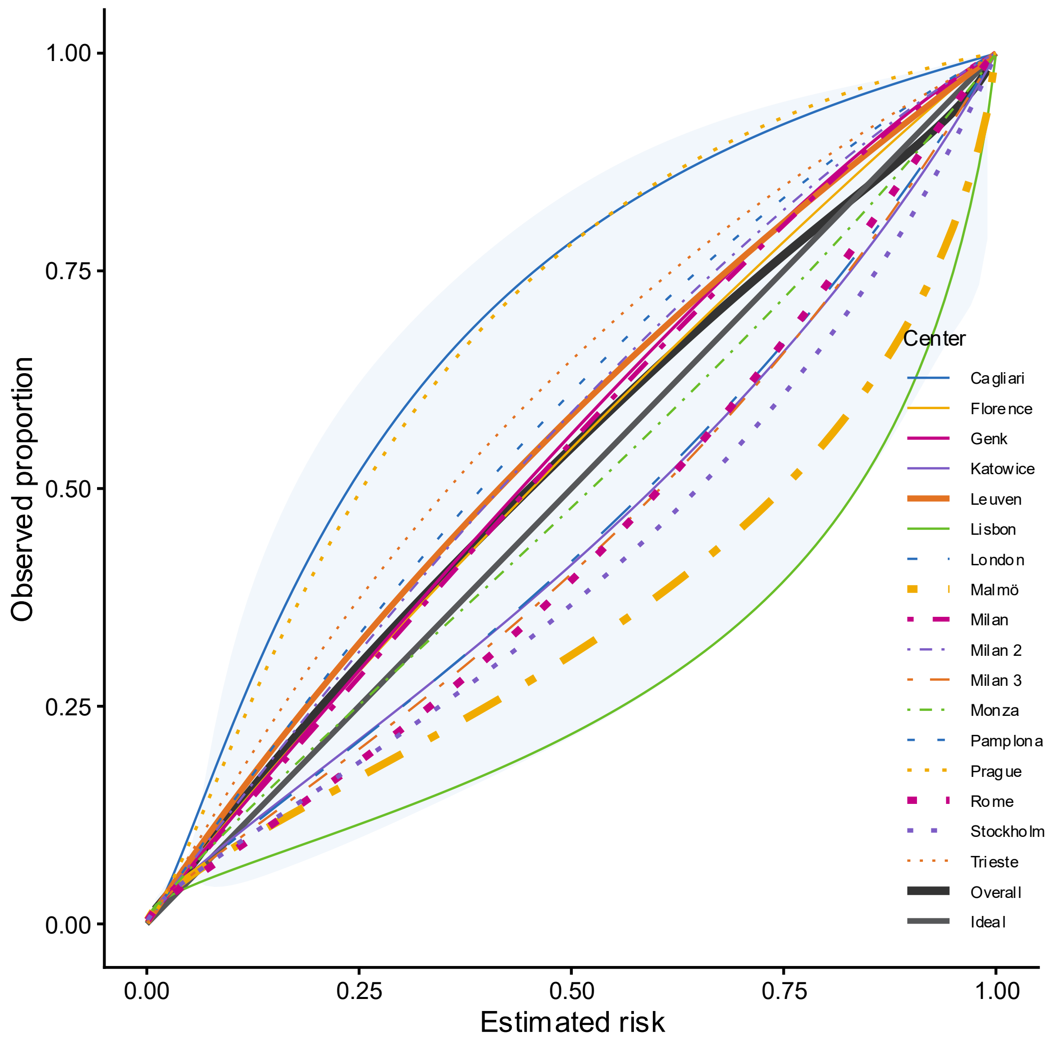

**Figure S62. Calibration plot for two step strategy with ADNEX2 in premenopausal patients (n = 3545). Overall calibration curves calculated taking clustering into account and shaded area indicating 95% prediction interval. Line width proportional to sample size.**

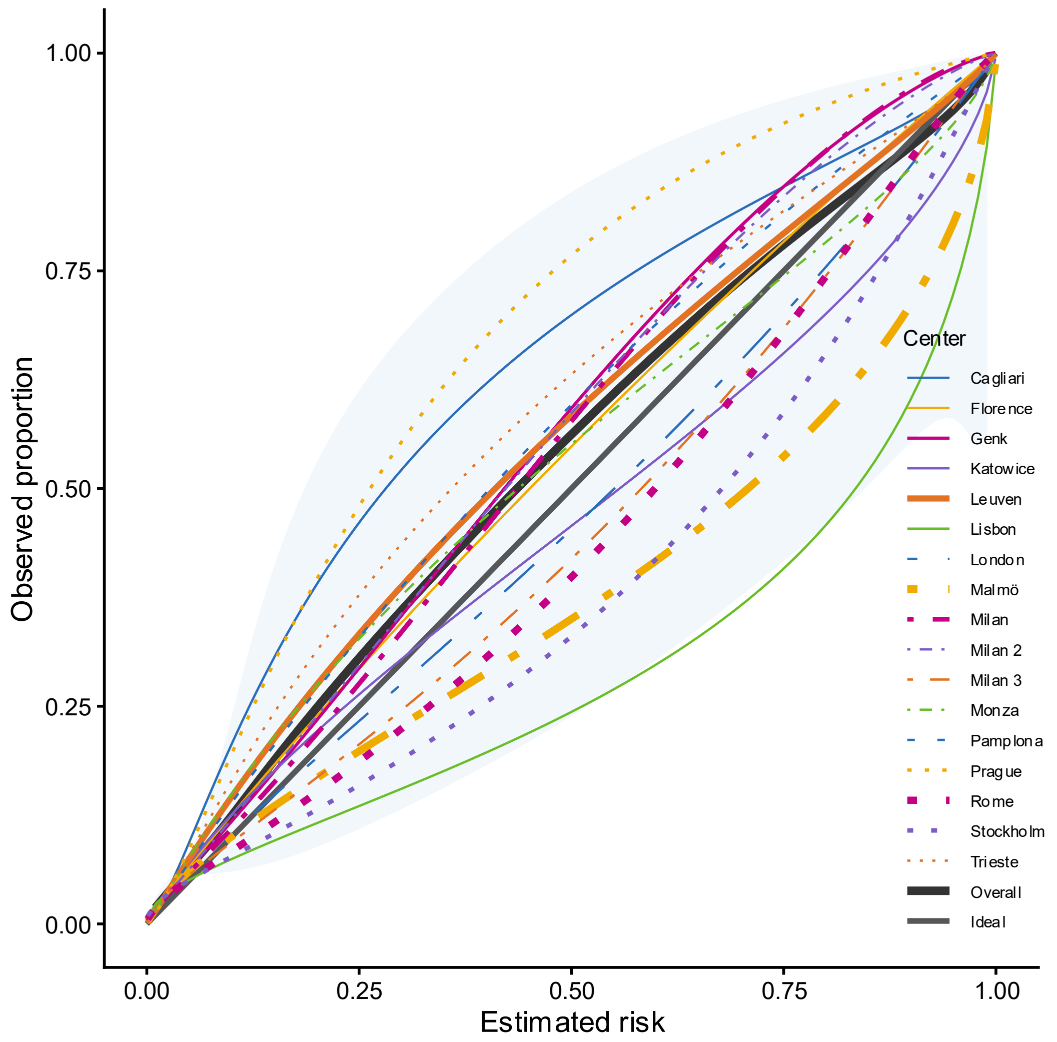

**Figure S63. Calibration plot for two step strategy with ADNEX2 ignoring CA125 in premenopausal patients (n = 3545). Overall calibration curves calculated taking clustering into account and shaded area indicating 95% prediction interval. Line width proportional to sample size.**

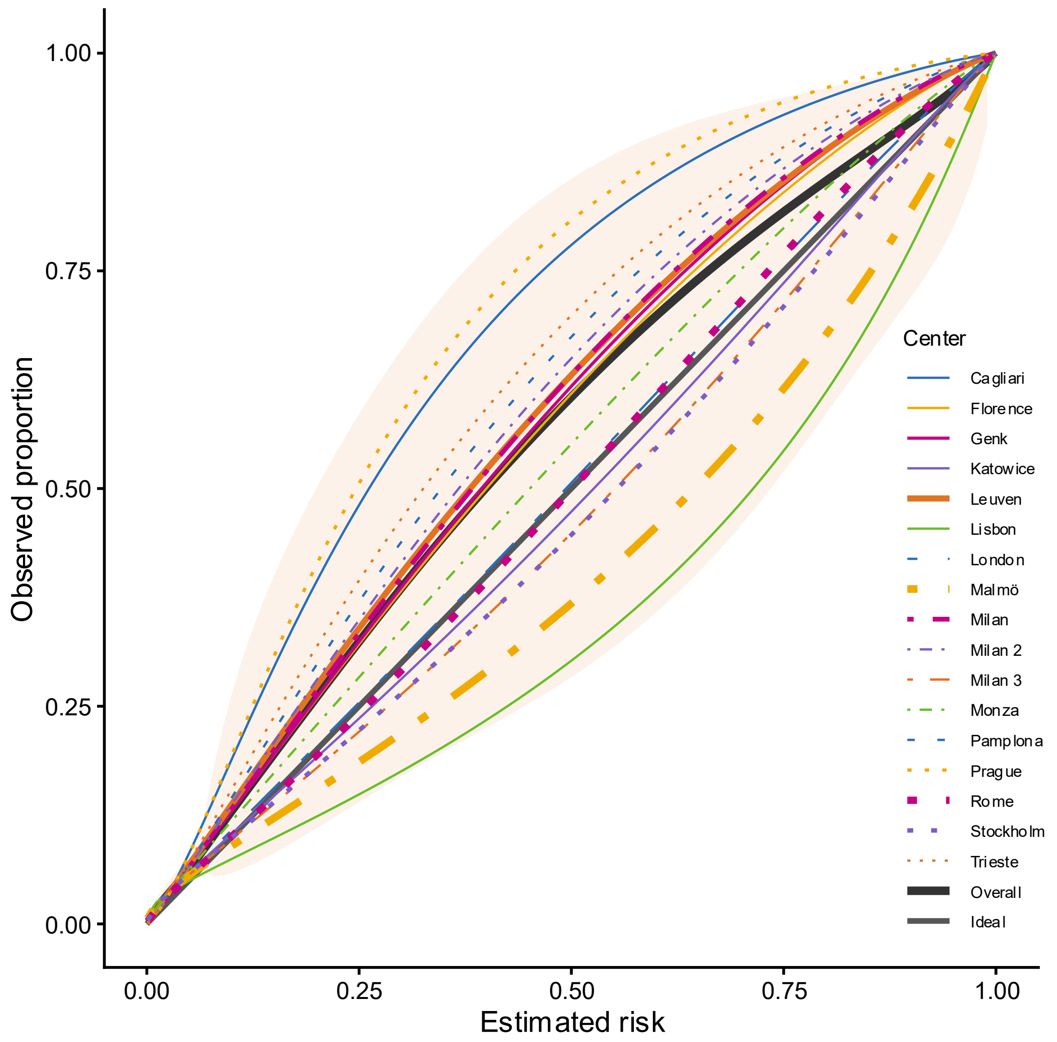

**Figure S64. Calibration plot for two step strategy with ADNEX in premenopausal patients (n = 3545). Overall calibration curves calculated taking clustering into account and shaded area indicating 95% prediction interval. Line width proportional to sample size.**

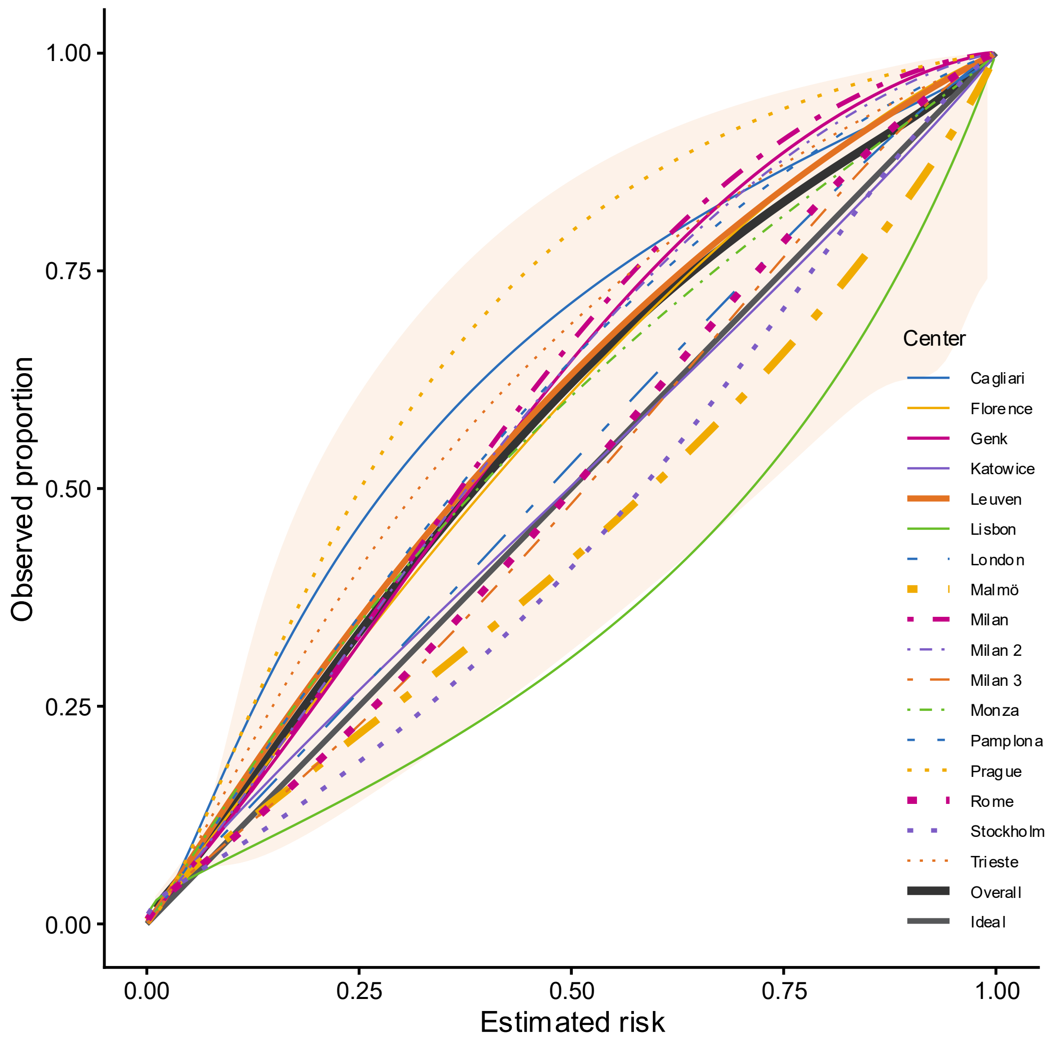

**Figure S65. Calibration plot for two step strategy with ADNEX ignoring CA125 in premenopausal patients (n = 3545). Overall calibration curves calculated taking clustering into account and shaded area indicating 95% prediction interval. Line width proportional to sample size.**

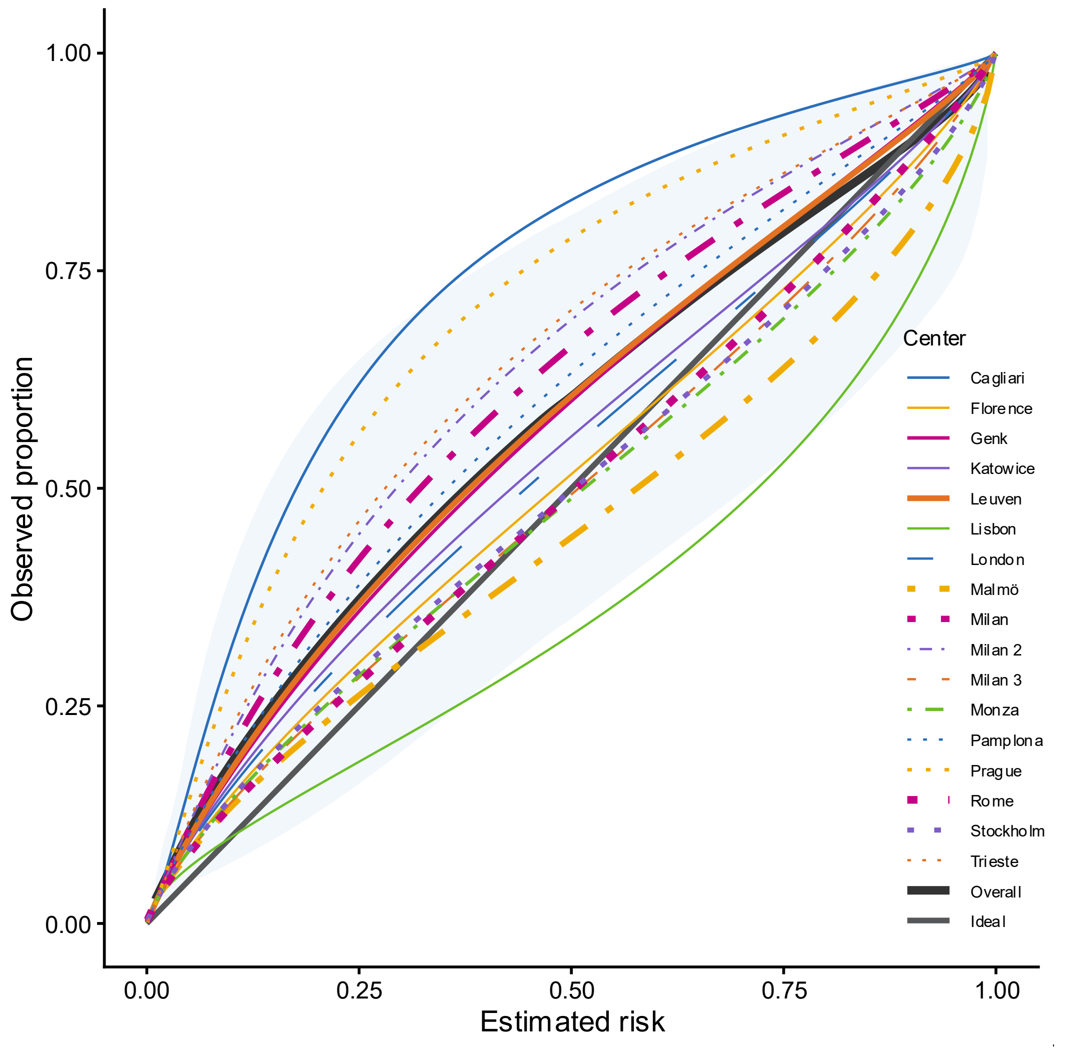

**Figure S66. Calibration plot for two step strategy with ADNEX2 in patients operated within 120 days after inclusion scan (n = 3679). Overall calibration curves calculated taking clustering into account and shaded area indicates 95% prediction interval. Line width is proportional to sample size.**

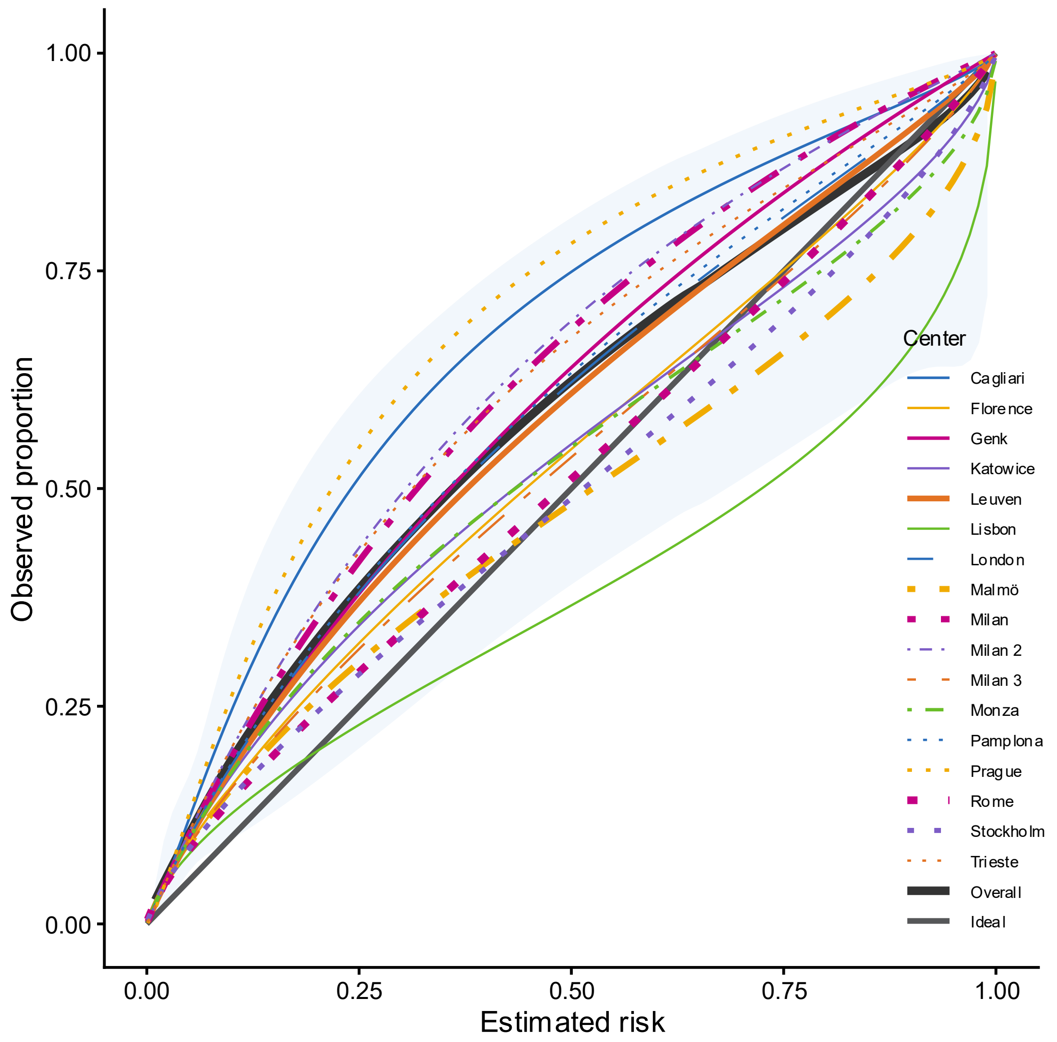

**Figure S67. Calibration plot for two step strategy with ADNEX2 ignoring CA125 in patients operated within 120 days after inclusion scan (n = 3679). Overall calibration curves calculated taking clustering into account and shaded area indicates 95% prediction interval. Line width is proportional to sample size.**

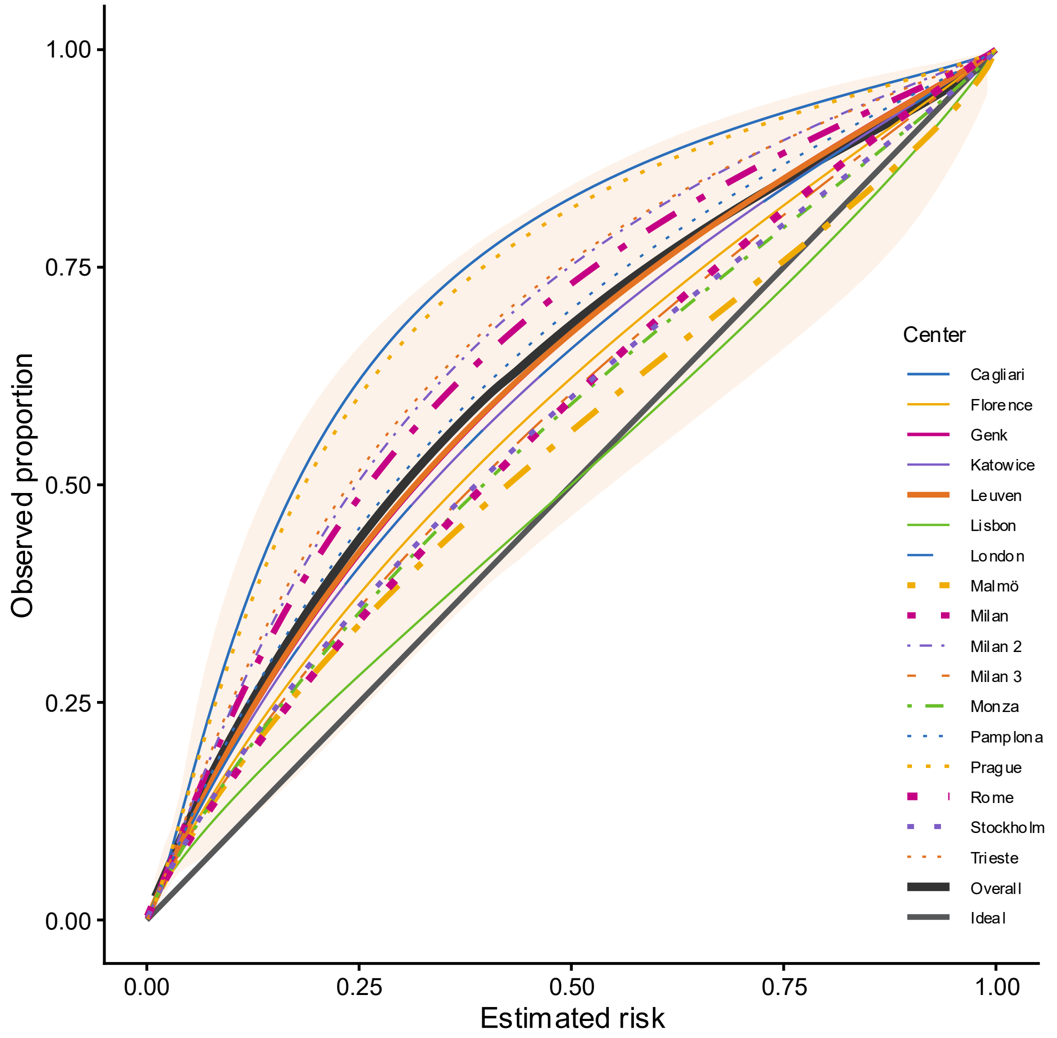

**Figure S68. Calibration plot for two step strategy with ADNEX in patients operated within 120 days after inclusion scan (n = 3679). Overall calibration curves calculated taking clustering into account and shaded area indicates 95% prediction interval. Line width is proportional to sample size.**

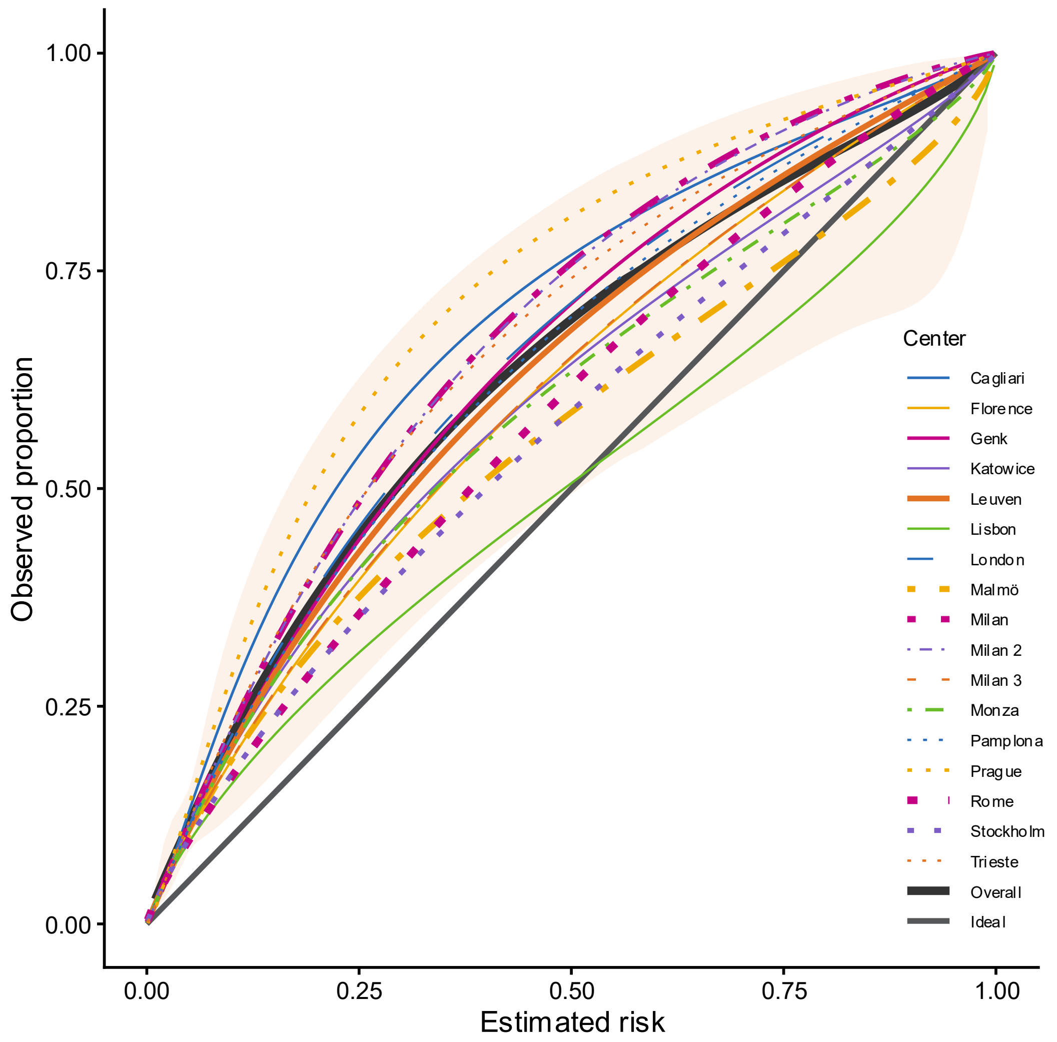

**Figure S69. Calibration plot for two step strategy with ADNEX ignoring CA125 in patients operated within 120 days after inclusion scan (n = 3679). Overall calibration curves calculated taking clustering into account and shaded area indicates 95% prediction interval. Line width is proportional to sample size.**

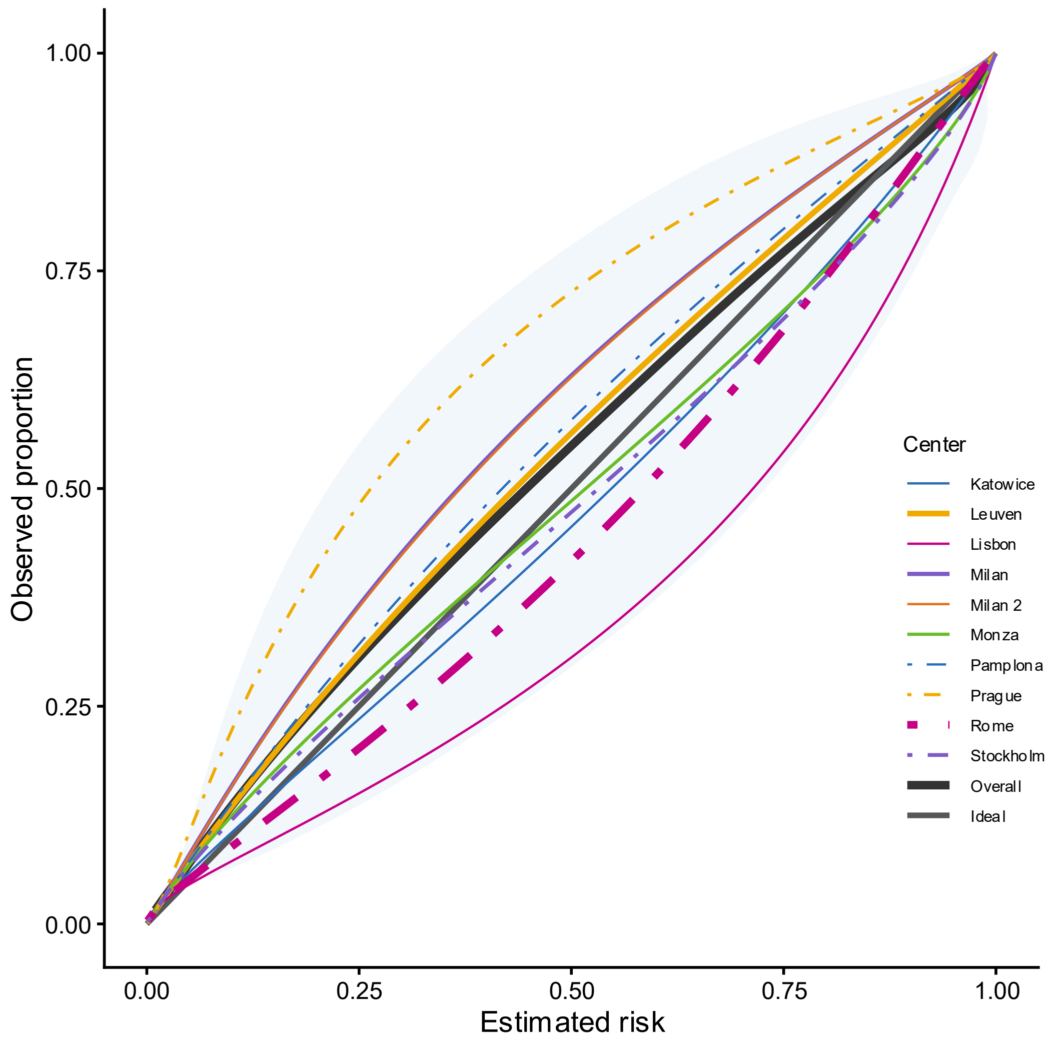

**Figure S70. Calibration plot for two step strategy with ADNEX2 withCA125 if available in oncology centres (n = 4375). Overall calibration curves calculated taking clustering into account and shaded area indicating 95% prediction interval. Line width is proportional to sample size.**

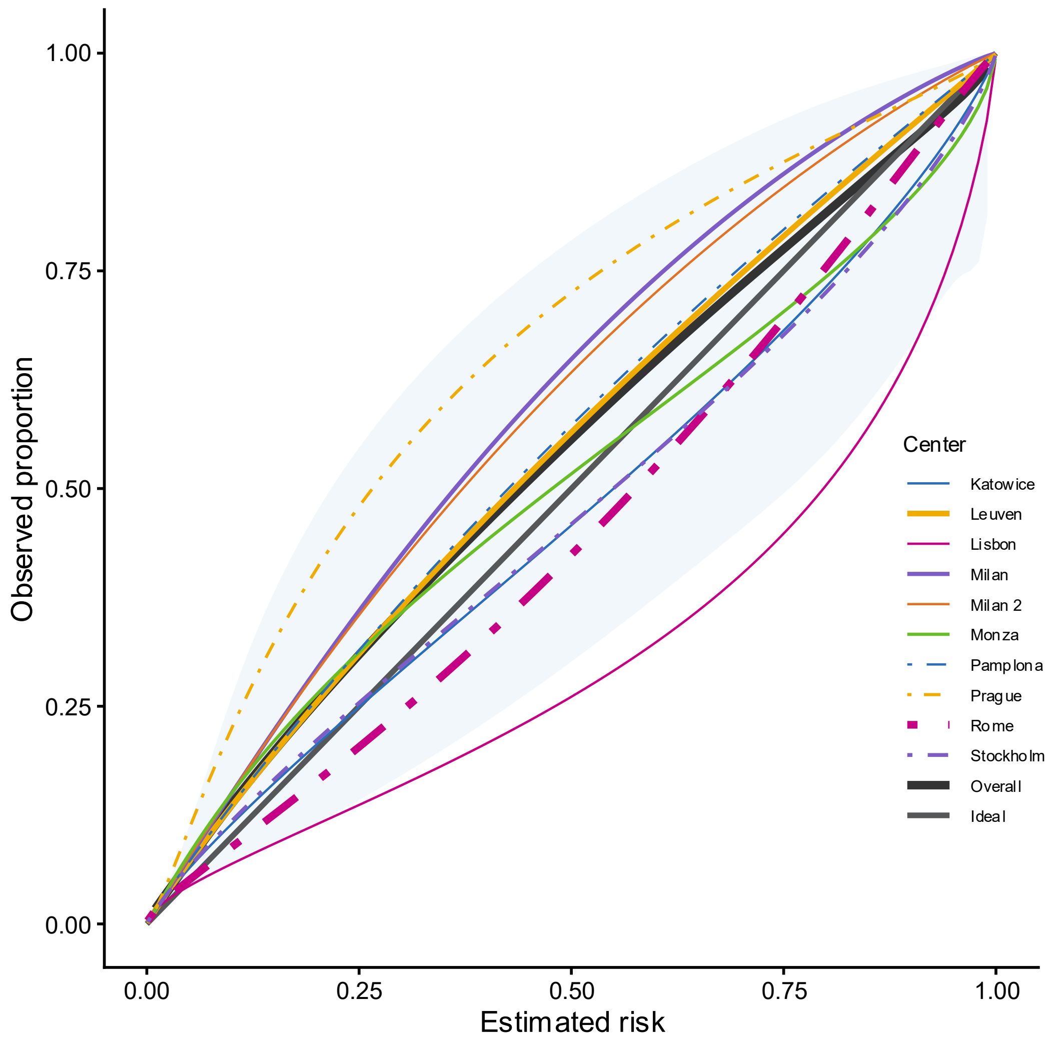

**Figure S71. Calibration plot for two step strategy with ADNEX2 ignoring CA125 in oncology centres (n = 4375). Overall calibration curves calculated taking clustering into account and shaded area indicating 95% prediction interval. Line width is proportional to sample size.**

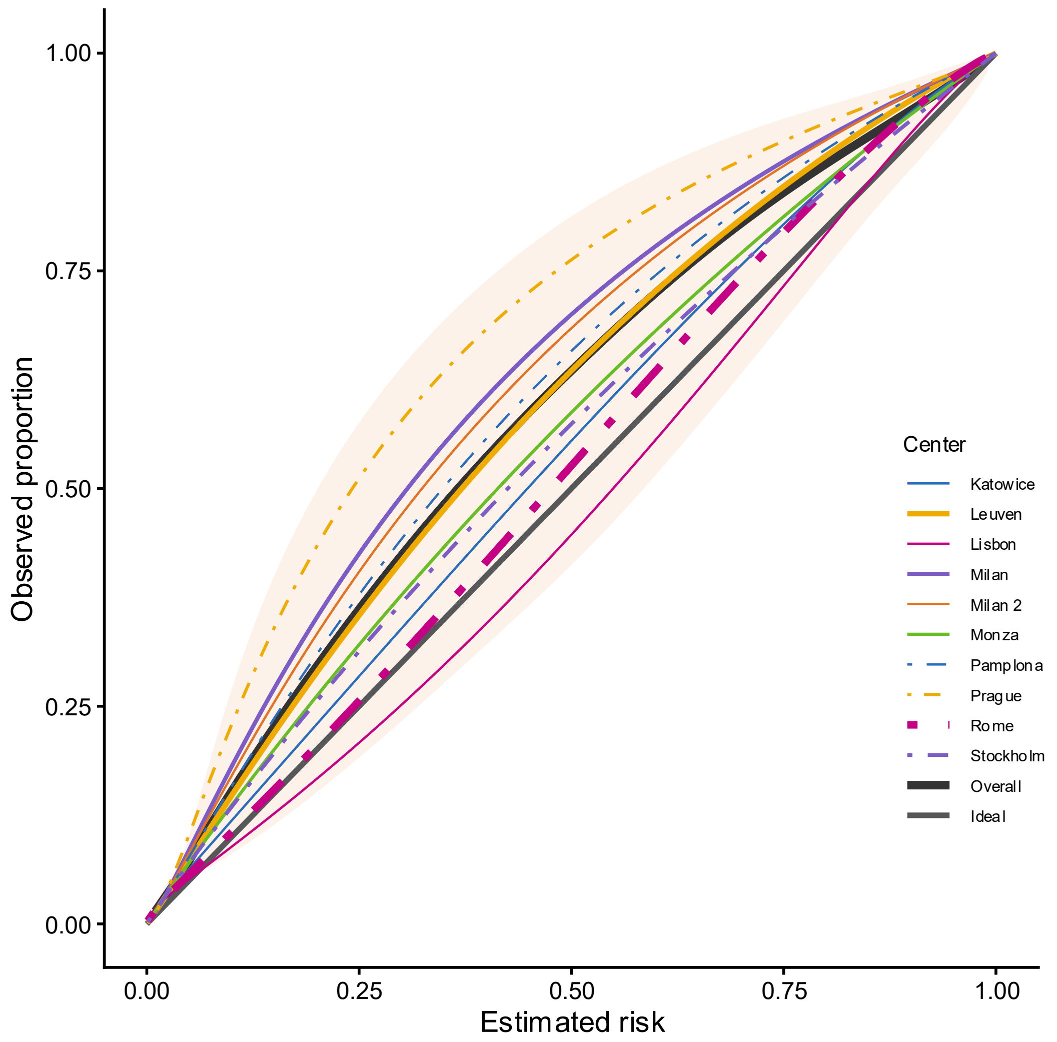

**Figure S72. Calibration plot for two step strategy with ADNEX withCA125 if available in oncology centres (n = 4375). Overall calibration curves calculated taking clustering into account and shaded area indicating 95% prediction interval. Line width is proportional to sample size.**

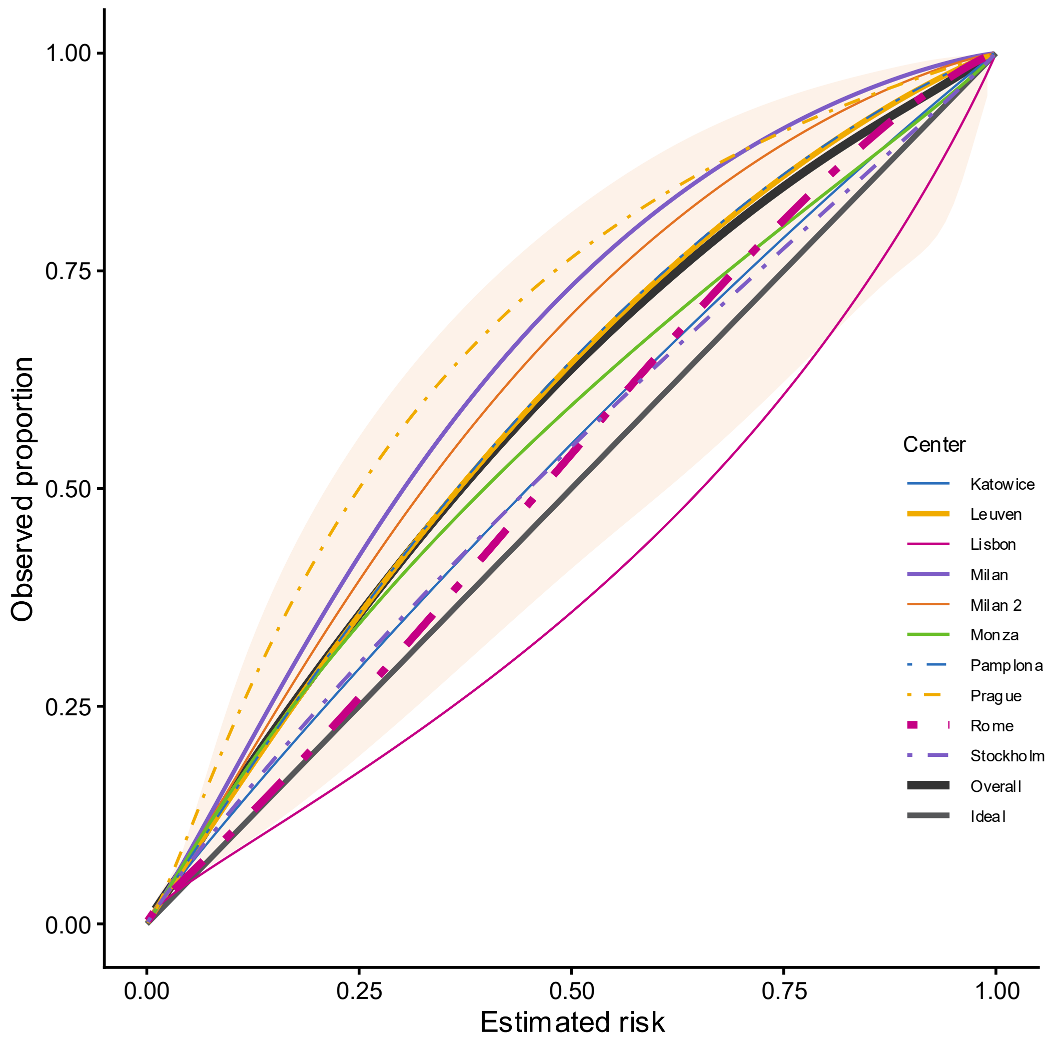

**Figure S73. Calibration plot for two step strategy with ADNEX ignoring CA125 in oncology centres (n = 4375). Overall calibration curves calculated taking clustering into account and shaded area indicating 95% prediction interval. Line width is proportional to sample size.**

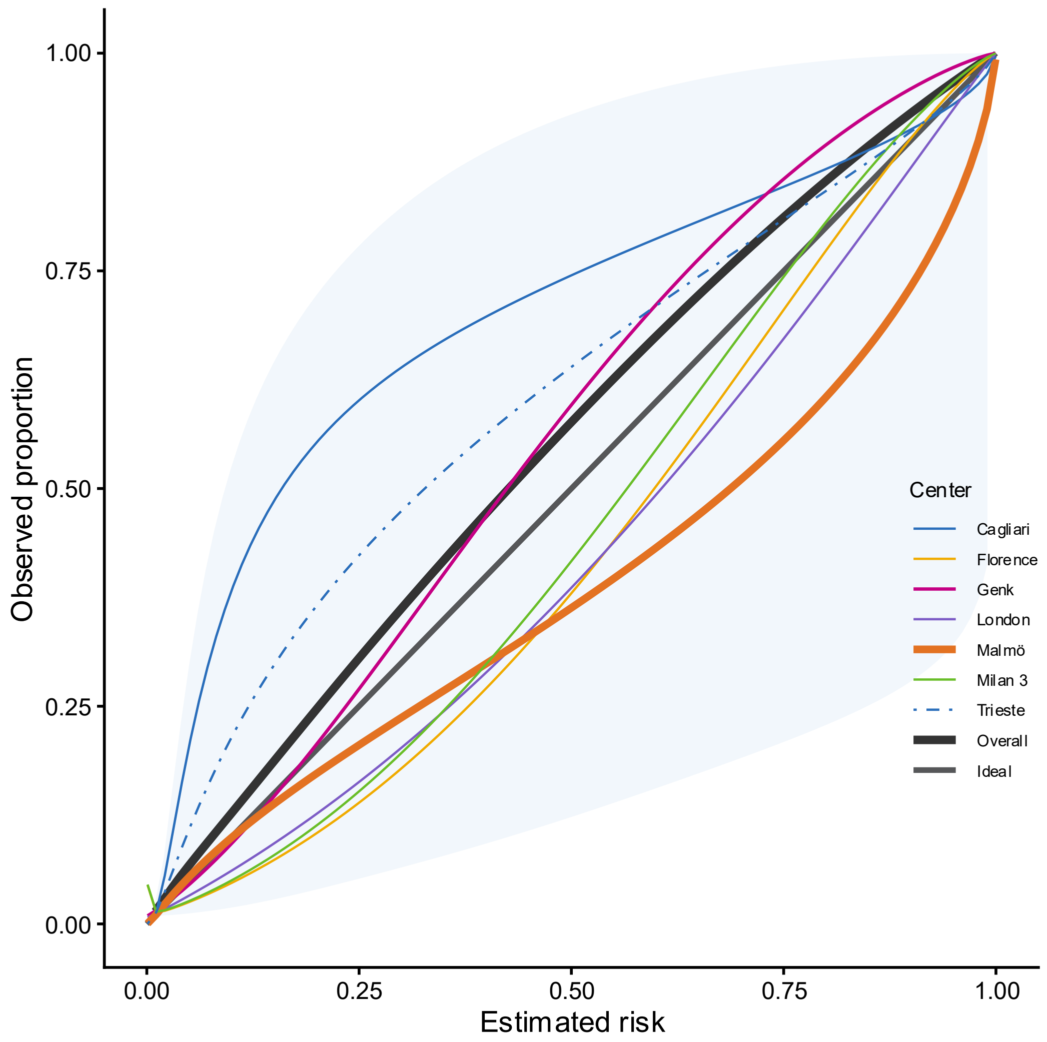

**Figure S74. Calibration plot for two step strategy with ADNEX2 in non-oncology centres (n = 2472). Overall calibration curves calculated taking clustering into account and shaded area indicating 95% prediction interval. Line width is proportional to sample size.**

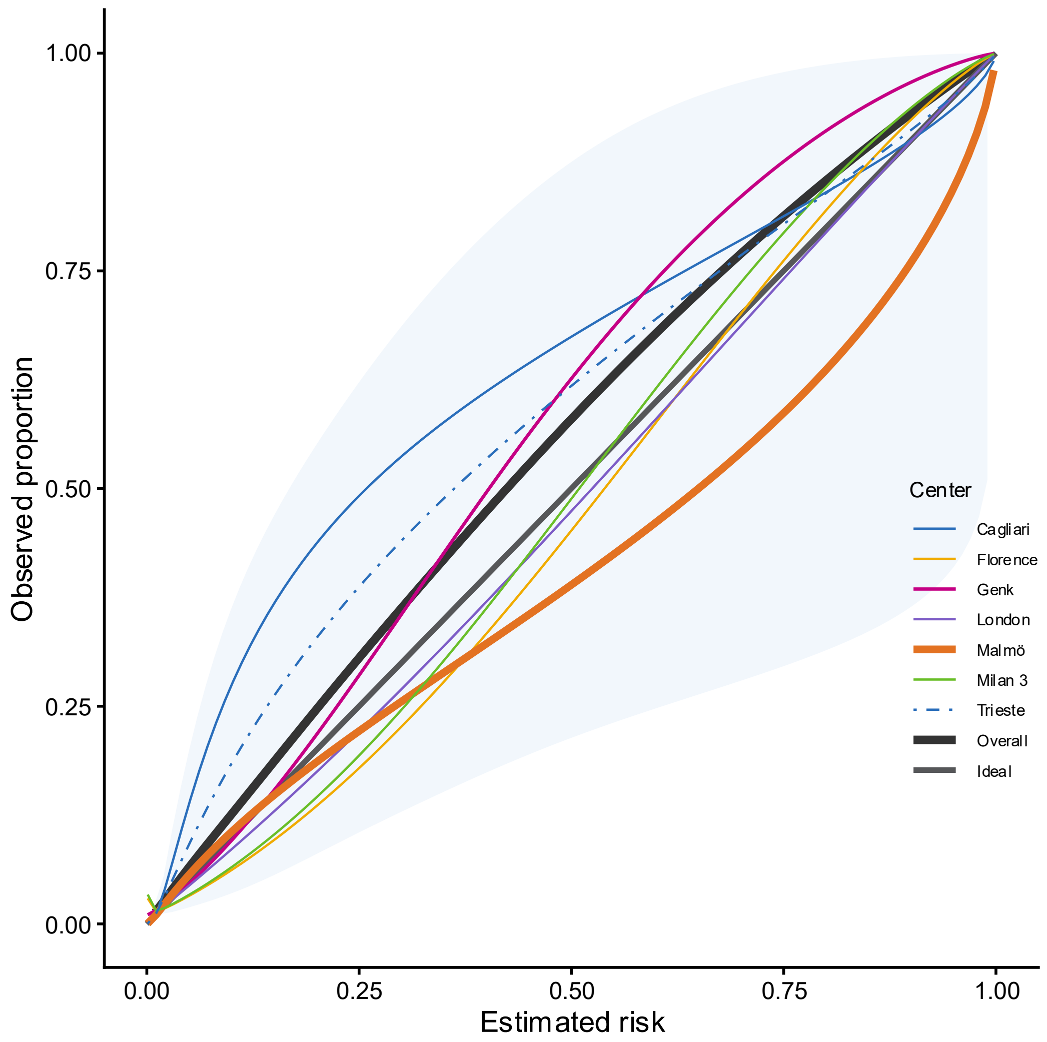

**Figure S75. Calibration plot for two step strategy with ADNEX2 ignoring CA125 in non-oncology centres (n = 2472). Overall calibration curves calculated taking clustering into account and shaded area indicating 95% prediction interval. Line width is proportional to sample size.**

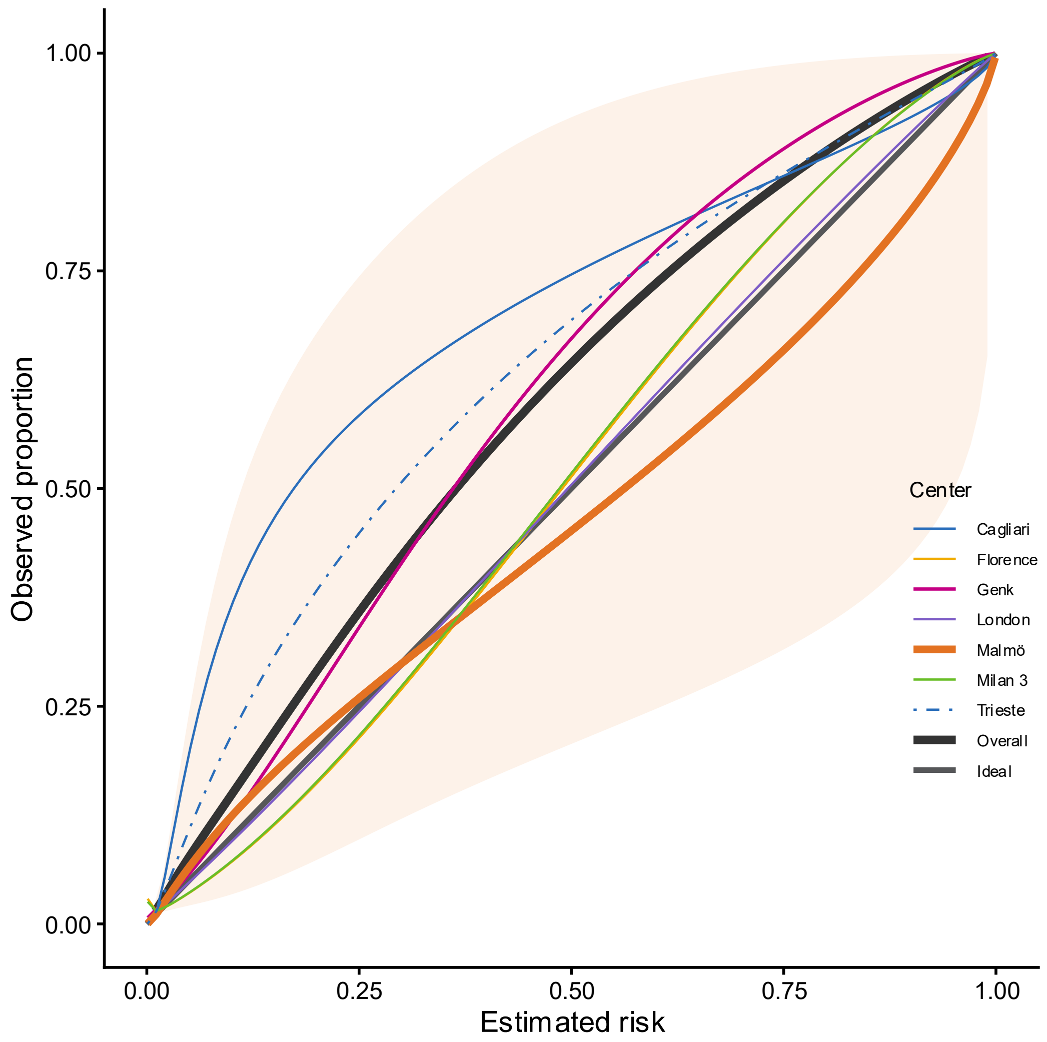

**Figure S76. Calibration plot for two step strategy with ADNEX in non-oncology centres (n = 2472). Overall calibration curves calculated taking clustering into account and shaded area indicating 95% prediction interval. Line width is proportional to sample size.**

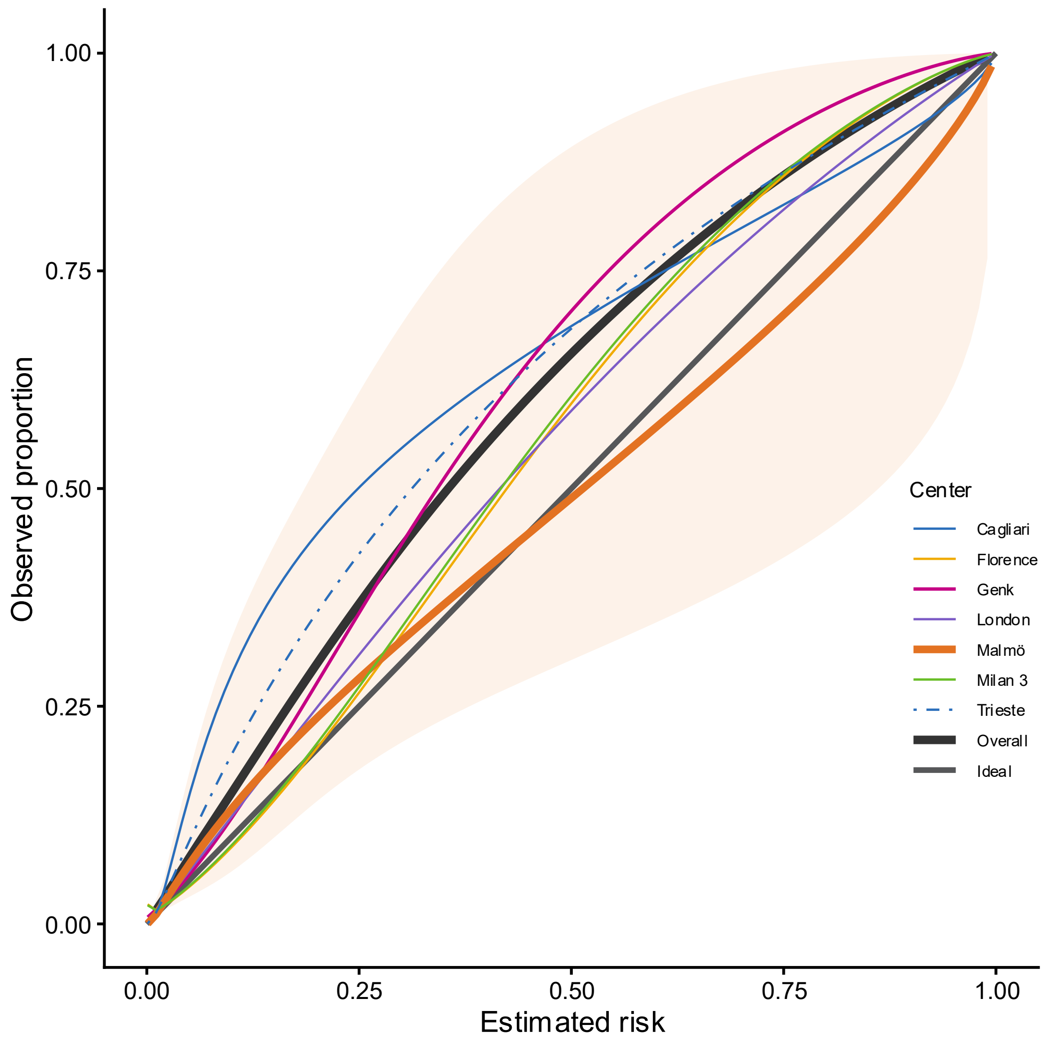

**Figure S77. Calibration plot for two step strategy with ADNEX ignoring CA125 in non-oncology centres (n = 2472). Overall calibration curves calculated taking clustering into account and shaded area indicating 95% prediction interval. Line width is proportional to sample size.**

#### S5.4 CLASSIFICATION

**Table S18. Sensitivity and specificity by subgroup for two step strategy with ADNEX2 and ADNEX**

| | | **2SS with ADNEX2** | | **2SS with ADNEX** | | |
|---|---|---|---|---|---|---|
| | Threshold | Sensitivity | Specificity | Sensitivity | Specificity | CA125 |
| Premenopausal | 1% | 0.961 (0.914 - 0.983) | 0.608 (0.540 - 0.671) | 0.957 (0.912 - 0.980) | 0.612 (0.547 - 0.674) | If available |
| | | 0.965 (0.928 - 0.984) | 0.604 (0.539 - 0.666) | 0.965 (0.927 - 0.984) | 0.613 (0.549 - 0.673) | Ignored |
| | 5% | 0.925 (0.879 - 0.955) | 0.825 (0.782 - 0.862) | 0.926 (0.882 - 0.955) | 0.843 (0.802 - 0.878) | If available |
| | | 0.931 (0.887 - 0.958) | 0.821 (0.778 - 0.858) | 0.927 (0.881 - 0.956) | 0.839 (0.797 - 0.874) | Ignored |
| | **10%** | **0.895 (0.849 - 0.928)** | **0.876 (0.845 - 0.901)** | **0.879 (0.828 - 0.917)** | **0.896 (0.863 - 0.922)** | **If available** |
| | | **0.905 (0.861 - 0.936)** | **0.875 (0.844 - 0.901)** | **0.889 (0.839 - 0.925)** | **0.894 (0.863 - 0.919)** | **Ignored** |
| | 20% | 0.820 (0.766 - 0.864) | 0.922 (0.897 - 0.941) | 0.775 (0.707 - 0.831) | 0.940 (0.916 - 0.958) | If available |
| | | 0.838 (0.788 - 0.878) | 0.916 (0.892 - 0.936) | 0.772 (0.708 - 0.826) | 0.935 (0.912 - 0.953) | Ignored |
| | 30% | 0.749 (0.681 - 0.807) | 0.947 (0.925 - 0.963) | 0.701 (0.631 - 0.763) | 0.963 (0.943 - 0.977) | If available |
| | | 0.754 (0.686 - 0.812) | 0.942 (0.921 - 0.958) | 0.701 (0.621 - 0.771) | 0.956 (0.935 - 0.971) | Ignored |
| | 40% | 0.693 (0.619 - 0.759) | 0.960 (0.939 - 0.974) | 0.655 (0.583 - 0.721) | 0.970 (0.948 - 0.983) | If available |
| | | 0.704 (0.624 - 0.773) | 0.956 (0.935 - 0.970) | 0.641 (0.558 - 0.716) | 0.968 (0.948 - 0.981) | Ignored |
| Postmenopausal | 1% | 0.979 (0.960 - 0.989) | 0.403 (0.336 - 0.473) | 0.981 (0.964 - 0.990) | 0.376 (0.308 - 0.448) | If available |
| | | 0.979 (0.961 - 0.988) | 0.393 (0.327 - 0.463) | 0.980 (0.964 - 0.989) | 0.374 (0.307 - 0.446) | Ignored |
| | 5% | 0.966 (0.939 - 0.981) | 0.671 (0.599 - 0.736) | 0.965 (0.933 - 0.982) | 0.653 (0.579 - 0.721) | If available |
| | | 0.962 (0.935 - 0.978) | 0.647 (0.580 - 0.709) | 0.961 (0.929 - 0.979) | 0.625 (0.550 - 0.695) | Ignored |
| | **10%** | **0.948 (0.909 - 0.971)** | **0.770 (0.701 - 0.827)** | **0.949 (0.909 - 0.972)** | **0.765 (0.694 - 0.824)** | **If available** |
| | | **0.949 (0.911 - 0.971)** | **0.753 (0.682 - 0.813)** | **0.948 (0.905 - 0.972)** | **0.755 (0.676 - 0.819)** | **Ignored** |
| | 20% | 0.919 (0.858 - 0.955) | 0.847 (0.781 - 0.895) | 0.906 (0.836 - 0.947) | 0.856 (0.806 - 0.895) | If available |
| | | 0.913 (0.858 - 0.948) | 0.833 (0.765 - 0.885) | 0.907 (0.845 - 0.946) | 0.849 (0.791 - 0.893) | Ignored |
| | 30% | 0.886 (0.812 - 0.933) | 0.883 (0.838 - 0.916) | 0.861 (0.785 - 0.913) | 0.894 (0.853 - 0.924) | If available |
| | | 0.890 (0.822 - 0.934) | 0.872 (0.822 - 0.909) | 0.870 (0.802 - 0.918) | 0.885 (0.841 - 0.918) | Ignored |
| | 40% | 0.850 (0.770 - 0.906) | 0.907 (0.871 - 0.934) | 0.829 (0.754 - 0.884) | 0.920 (0.889 - 0.943) | If available |
| | | 0.858 (0.786 - 0.908) | 0.902 (0.863 - 0.931) | 0.813 (0.742 - 0.868) | 0.911 (0.883 - 0.934) | Ignored |
| Surgically managed within 120 days | 1% | 0.979 (0.951 - 0.991) | 0.407 (0.334 - 0.484) | 0.977 (0.950 - 0.990) | 0.409 (0.338 - 0.484) | If available |
| | | 0.979 (0.955 - 0.990) | 0.401 (0.330 - 0.477) | 0.977 (0.952 - 0.989) | 0.413 (0.343 - 0.485) | Ignored |
| | 5% | 0.962 (0.942 - 0.976) | 0.617 (0.534 - 0.694) | 0.960 (0.934 - 0.977) | 0.640 (0.558 - 0.714) | If available |
| | | 0.963 (0.939 - 0.978) | 0.588 (0.499 - 0.671) | 0.960 (0.932 - 0.977) | 0.613 (0.527 - 0.693) | Ignored |
| | **10%** | **0.943 (0.912 - 0.963)** | **0.723 (0.649 - 0.787)** | **0.936 (0.902 - 0.959)** | **0.744 (0.670 - 0.807)** | **If available** |
| | | **0.945 (0.916 - 0.964)** | **0.708 (0.633 - 0.773)** | **0.937 (0.901 - 0.960)** | **0.734 (0.659 - 0.797)** | **Ignored** |
| | 20% | 0.898 (0.843 - 0.936) | 0.812 (0.744 - 0.866) | 0.873 (0.807 - 0.918) | 0.837 (0.784 - 0.879) | If available |
| | | 0.898 (0.849 - 0.933) | 0.799 (0.731 - 0.853) | 0.869 (0.807 - 0.914) | 0.827 (0.769 - 0.873) | Ignored |
| | 30% | 0.845 (0.771 - 0.898) | 0.866 (0.809 - 0.907) | 0.816 (0.744 - 0.872) | 0.890 (0.841 - 0.924) | If available |
| | | 0.853 (0.782 - 0.904) | 0.846 (0.787 - 0.892) | 0.820 (0.747 - 0.875) | 0.874 (0.829 - 0.908) | Ignored |

| | | 2SS with ADNEX2 | | 2SS with ADNEX | | |
|---|---|---|---|---|---|---|
| | Threshold | Sensitivity | Specificity | Sensitivity | Specificity | CA125 |
| | 40% | 0.809 (0.732 - 0.868) | 0.889 (0.841 - 0.924) | 0.779 (0.707 - 0.838) | 0.911 (0.871 - 0.939) | If available |
| | | 0.814 (0.740 - 0.870) | 0.883 (0.836 - 0.918) | 0.764 (0.690 - 0.825) | 0.900 (0.861 - 0.929) | Ignored |
| Oncology centres | 1% | 0.987 (0.974 - 0.994) | 0.458 (0.360 - 0.559) | 0.986 (0.973 - 0.993) | 0.454 (0.356 - 0.555) | If available |
| | | 0.985 (0.973 - 0.992) | 0.455 (0.357 - 0.555) | 0.985 (0.973 - 0.992) | 0.456 (0.359 - 0.556) | Ignored |
| | 5% | 0.972 (0.954 - 0.983) | 0.704 (0.639 - 0.762) | 0.974 (0.957 - 0.984) | 0.703 (0.635 - 0.763) | If available |
| | | 0.971 (0.953 - 0.982) | 0.691 (0.619 - 0.754) | 0.973 (0.953 - 0.984) | 0.687 (0.607 - 0.758) | Ignored |
| | **10%** | **0.957 (0.936 - 0.971)** | **0.787 (0.730 - 0.834)** | **0.955 (0.935 - 0.969)** | **0.789 (0.733 - 0.836)** | **If available** |
| | | **0.958 (0.936 - 0.972)** | **0.777 (0.717 - 0.827)** | **0.956 (0.936 - 0.970)** | **0.781 (0.722 - 0.831)** | **Ignored** |
| | 20% | 0.925 (0.900 - 0.944) | 0.853 (0.810 - 0.888) | 0.914 (0.894 - 0.931) | 0.870 (0.832 - 0.900) | If available |
| | | 0.924 (0.898 - 0.944) | 0.841 (0.794 - 0.880) | 0.912 (0.889 - 0.931) | 0.862 (0.823 - 0.894) | Ignored |
| | 30% | 0.891 (0.869 - 0.910) | 0.892 (0.859 - 0.918) | 0.867 (0.843 - 0.887) | 0.909 (0.880 - 0.931) | If available |
| | | 0.892 (0.859 - 0.918) | 0.884 (0.847 - 0.914) | 0.868 (0.836 - 0.894) | 0.900 (0.870 - 0.923) | Ignored |
| | 40% | 0.860 (0.832 - 0.884) | 0.913 (0.886 - 0.934) | 0.831 (0.808 - 0.851) | 0.932 (0.909 - 0.950) | If available |
| | | 0.862 (0.823 - 0.893) | 0.909 (0.880 - 0.933) | 0.820 (0.790 - 0.847) | 0.926 (0.903 - 0.944) | Ignored |
| Non-oncology centres | 1% | 0.918 (0.749 - 0.977) | 0.606 (0.541 - 0.668) | 0.921 (0.756 - 0.978) | 0.590 (0.522 - 0.654) | If available |
| | | 0.919 (0.748 - 0.977) | 0.597 (0.535 - 0.656) | 0.927 (0.764 - 0.980) | 0.588 (0.518 - 0.654) | Ignored |
| | 5% | 0.909 (0.859 - 0.943) | 0.840 (0.794 - 0.878) | 0.901 (0.852 - 0.935) | 0.846 (0.804 - 0.881) | If available |
| | | 0.888 (0.812 - 0.936) | 0.828 (0.786 - 0.864) | 0.888 (0.824 - 0.931) | 0.834 (0.794 - 0.867) | Ignored |
| | **10%** | **0.853 (0.796 - 0.895)** | **0.894 (0.857 - 0.922)** | **0.837 (0.781 - 0.881)** | **0.910 (0.877 - 0.934)** | **If available** |
| | | **0.868 (0.816 - 0.907)** | **0.888 (0.854 - 0.915)** | **0.842 (0.789 - 0.884)** | **0.905 (0.873 - 0.930)** | **Ignored** |
| | 20% | 0.735 (0.645 - 0.809) | 0.944 (0.911 - 0.965) | 0.678 (0.609 - 0.740) | 0.952 (0.929 - 0.968) | If available |
| | | 0.760 (0.694 - 0.816) | 0.937 (0.908 - 0.958) | 0.691 (0.623 - 0.751) | 0.948 (0.925 - 0.964) | Ignored |
| | 30% | 0.622 (0.517 - 0.717) | 0.966 (0.942 - 0.980) | 0.600 (0.518 - 0.676) | 0.971 (0.953 - 0.982) | If available |
| | | 0.656 (0.582 - 0.724) | 0.957 (0.934 - 0.972) | 0.622 (0.561 - 0.679) | 0.964 (0.946 - 0.976) | Ignored |
| | 40% | 0.583 (0.505 - 0.658) | 0.973 (0.954 - 0.984) | 0.561 (0.488 - 0.631) | 0.980 (0.961 - 0.990) | If available |
| | | 0.613 (0.546 - 0.676) | 0.970 (0.950 - 0.982) | 0.550 (0.481 - 0.617) | 0.977 (0.958 - 0.987) | Ignored |

#### S5.5 CLINICAL UTILITY

**Table S19. Clinical utility for deciding whom to refer to specialized oncological care of two step strategy with ADNEX or ADNEX2 across subgroups.**

| | | | 2SS with ADNEX2 | | | 2SS with ADNEX | | |
|---|---|---|---|---|---|---|---|---|
| **Subgroup** | **CA125** | **Threshold** | **NB (95% Crl; PI)** | **$\Delta NB$** | **P-useful** | **NB (95% Crl; PI)** | **$\Delta NB$** | **P-useful** |
| Premenopausal | BDs only | 0.01 | 0.13 (0.08-0.20; 0.01-0.57) | -0.0013 | 69% | 0.13 (0.08-0.20; 0.01-0.57) | -0.0013 | 69% |
| | If available | 0.01 | 0.13 (0.08-0.20;0.01-0.57) | 0.0021 | 69% | 0.13 (0.08-0.20;0.01-0.56) | 0.002 | 67% |
| | If available | 0.02 | 0.13 (0.08-0.20;0.01-0.56) | 0.0078 | 81% | 0.13 (0.08-0.20;0.01-0.56) | 0.0074 | 79% |
| | If available | 0.03 | 0.12 (0.07-0.19;0.01-0.55) | 0.0123 | 85% | 0.13 (0.07-0.19;0.01-0.54) | 0.0123 | 86% |
| | If available | 0.04 | 0.12 (0.07-0.19;0.01-0.54) | 0.0194 | 91% | 0.12 (0.07-0.19;0.01-0.54) | 0.0201 | 91% |
| | If available | 0.05 | 0.12 (0.07-0.19;0.01-0.53) | 0.0282 | 94% | 0.12 (0.07-0.19;0.01-0.53) | 0.0287 | 95% |
| | If available | **0.10** | **0.11 (0.06-0.18;0.00-0.53)** | **0.0693** | **96%** | **0.11 (0.06-0.18;0.01-0.52)** | **0.0688** | **96%** |
| | If available | 0.15 | 0.11 (0.06-0.17;0.00-0.52) | 0.1069 | 96% | 0.10 (0.06-0.16;0.00-0.51) | 0.1032 | 97% |
| | If available | 0.20 | 0.10 (0.05-0.16;0.00-0.50) | 0.0989 | 96% | 0.10 (0.05-0.16;0.00-0.49) | 0.0959 | 97% |
| | If available | 0.25 | 0.09 (0.05-0.15;-0.00-0.49) | 0.0929 | 96% | 0.09 (0.05-0.14;0.00-0.45) | 0.0883 | 97% |
| | If available | 0.30 | 0.09 (0.04-0.14;-0.00-0.46) | 0.0862 | 96% | 0.09 (0.05-0.14;0.00-0.43) | 0.086 | 98% |
| | If available | 0.35 | 0.08 (0.04-0.13;-0.00-0.43) | 0.0812 | 96% | 0.08 (0.04-0.13;0.00-0.41) | 0.0816 | 98% |
| | If available | 0.40 | 0.08 (0.04-0.13;-0.00-0.41) | 0.0763 | 96% | 0.08 (0.05-0.13;0.00-0.38) | 0.0802 | 98% |
| | Ignored | 0.01 | 0.13 (0.08-0.20;0.01-0.56) | 0.0021 | 69% | 0.13 (0.08-0.20;0.01-0.56) | 0.0022 | 70% |
| | Ignored | 0.02 | 0.13 (0.08-0.20;0.01-0.55) | 0.0075 | 82% | 0.13 (0.08-0.20;0.01-0.56) | 0.0069 | 78% |
| | Ignored | 0.03 | 0.13 (0.07-0.19;0.01-0.55) | 0.0129 | 85% | 0.13 (0.08-0.19;0.01-0.54) | 0.0132 | 84% |
| | Ignored | 0.04 | 0.12 (0.07-0.19;0.01-0.55) | 0.0202 | 90% | 0.12 (0.07-0.19;0.01-0.54) | 0.0199 | 90% |
| | Ignored | 0.05 | 0.12 (0.07-0.19;0.01-0.54) | 0.0283 | 94% | 0.12 (0.07-0.19;0.01-0.54) | 0.0284 | 94% |
| | Ignored | **0.10** | **0.11 (0.06-0.18;0.01-0.53)** | **0.0704** | **96%** | **0.11 (0.07-0.18;0.01-0.52)** | **0.0701** | **96%** |
| | Ignored | 0.15 | 0.11 (0.06-0.17;0.00-0.52) | 0.108 | 96% | 0.10 (0.06-0.17;0.00-0.50) | 0.1049 | 97% |
| | Ignored | 0.20 | 0.10 (0.05-0.16;-0.00-0.50) | 0.0998 | 96% | 0.09 (0.05-0.15;0.00-0.48) | 0.0938 | 96% |
| | Ignored | 0.25 | 0.09 (0.05-0.15;-0.00-0.48) | 0.0921 | 96% | 0.09 (0.05-0.14;0.00-0.46) | 0.0885 | 97% |
| | Ignored | 0.30 | 0.09 (0.04-0.14;-0.00-0.47) | 0.0853 | 95% | 0.08 (0.04-0.14;-0.00-0.45) | 0.0832 | 97% |
| | Ignored | 0.35 | 0.08 (0.04-0.13;-0.01-0.44) | 0.0802 | 95% | 0.08 (0.04-0.13;0.00-0.42) | 0.0821 | 97% |
| | Ignored | 0.40 | 0.08 (0.04-0.13;-0.01-0.43) | 0.0766 | 95% | 0.08 (0.04-0.12;0.00-0.39) | 0.0755 | 98% |
| Postmenopausal | BDs only | 0.01 | 0.28 (0.18-0.40; 0.03-0.79) | 0.0007 | 48% | 0.28 (0.18-0.40; 0.03-0.79) | 0.0007 | 48% |
| | If available | 0.01 | 0.28 (0.18-0.39;0.03-0.78) | -0.0013 | 36% | 0.28 (0.18-0.39;0.03-0.78) | -0.0015 | 34% |
| | If available | 0.02 | 0.27 (0.18-0.38;0.03-0.77) | 0.0026 | 66% | 0.27 (0.18-0.38;0.03-0.76) | 0.0018 | 61% |
| | If available | 0.03 | 0.27 (0.17-0.38;0.03-0.77) | 0.0077 | 80% | 0.27 (0.18-0.38;0.03-0.75) | 0.0067 | 79% |
| | If available | 0.04 | 0.27 (0.17-0.38;0.02-0.77) | 0.0132 | 88% | 0.27 (0.17-0.38;0.03-0.76) | 0.0126 | 87% |
| | If available | 0.05 | 0.27 (0.17-0.38;0.02-0.77) | 0.0179 | 89% | 0.27 (0.17-0.38;0.02-0.76) | 0.0173 | 90% |
| | If available | **0.10** | **0.26 (0.16-0.37;0.01-0.75)** | **0.0469** | **96%** | **0.26 (0.16-0.37;0.02-0.74)** | **0.0473** | **96%** |
| | If available | 0.15 | 0.25 (0.15-0.36;0.01-0.73) | 0.0861 | 97% | 0.25 (0.15-0.36;0.01-0.73) | 0.0842 | 96% |
| | If available | 0.20 | 0.24 (0.14-0.35;0.00-0.73) | 0.1283 | 96% | 0.23 (0.14-0.35;0.00-0.73) | 0.1249 | 96% |
| | If available | 0.25 | 0.23 (0.13-0.34;-0.00-0.72) | 0.1786 | 95% | 0.22 (0.13-0.33;-0.00-0.72) | 0.1732 | 96% |
| | If available | 0.30 | 0.22 (0.13-0.33;-0.00-0.71) | 0.2201 | 96% | 0.22 (0.12-0.32;-0.00-0.71) | 0.2155 | 96% |
| | If available | 0.35 | 0.21 (0.11-0.32;-0.01-0.70) | 0.2077 | 94% | 0.21 (0.12-0.32;-0.00-0.70) | 0.2081 | 96% |
| | If available | 0.40 | 0.20 (0.11-0.31;-0.01-0.70) | 0.203 | 94% | 0.20 (0.11-0.30;-0.00-0.69) | 0.1991 | 96% |
| | Ignored | 0.01 | 0.28 (0.18-0.39;0.03-0.78) | -0.0017 | 33% | 0.28 (0.18-0.39;0.03-0.78) | -0.0017 | 32% |
| | Ignored | 0.02 | 0.27 (0.18-0.38;0.03-0.77) | 0.0022 | 63% | 0.27 (0.18-0.38;0.03-0.76) | 0.0016 | 61% |
| | Ignored | 0.03 | 0.27 (0.17-0.38;0.03-0.76) | 0.0069 | 79% | 0.27 (0.18-0.38;0.03-0.75) | 0.0062 | 78% |
| | Ignored | 0.04 | 0.27 (0.17-0.38;0.03-0.76) | 0.0123 | 87% | 0.27 (0.17-0.38;0.03-0.75) | 0.0109 | 86% |
| | Ignored | 0.05 | 0.27 (0.17-0.38;0.02-0.76) | 0.0171 | 90% | 0.27 (0.17-0.37;0.03-0.75) | 0.0165 | 90% |
| | Ignored | **0.10** | **0.26 (0.16-0.37;0.02-0.75)** | **0.0464** | **95%** | **0.26 (0.16-0.36;0.02-0.74)** | **0.0464** | **95%** |
| | Ignored | 0.15 | 0.24 (0.15-0.35;0.01-0.73) | 0.083 | 96% | 0.24 (0.15-0.35;0.01-0.73) | 0.0815 | 96% |
| | Ignored | 0.20 | 0.24 (0.14-0.34;0.00-0.73) | 0.1249 | 96% | 0.23 (0.14-0.34;0.00-0.73) | 0.1247 | 96% |
| | Ignored | 0.25 | 0.23 (0.13-0.34;0.00-0.72) | 0.1781 | 96% | 0.22 (0.13-0.33;-0.00-0.72) | 0.1732 | 95% |
| | Ignored | 0.30 | 0.22 (0.12-0.33;-0.00-0.71) | 0.2191 | 95% | 0.22 (0.12-0.32;-0.00-0.71) | 0.2154 | 96% |
| | Ignored | 0.35 | 0.21 (0.12-0.32;-0.01-0.70) | 0.2085 | 95% | 0.20 (0.11-0.31;-0.00-0.70) | 0.2038 | 96% |
| | Ignored | 0.40 | 0.20 (0.11-0.31;-0.01-0.69) | 0.2025 | 95% | 0.19 (0.10-0.29;-0.01-0.68) | 0.1889 | 95% |
| Operated within 120 days | BDs only | 0.01 | 0.35 (0.24-0.47; 0.05-0.82) | -0.003 | 55% | 0.35 (0.24-0.47; 0.05-0.82) | -0.003 | 55% |
| | If available | 0.01 | 0.35 (0.24-0.47;0.05-0.81) | -0.001 | 37% | 0.35 (0.24-0.47;0.05-0.81) | -0.0012 | 36% |

| | | | 2SS with ADNEX2 | | | 2SS with ADNEX | | |
|---|---|---|---|---|---|---|---|---|
| Subgroup | CA125 | Threshold | NB (95% Crl; PI) | $\Delta NB$ | P-useful | NB (95% Crl; PI) | $\Delta NB$ | P-useful |
| | If available | 0.02 | 0.35 (0.24-0.47;0.05-0.82) | 0.0012 | 58% | 0.35 (0.24-0.47;0.05-0.81) | 0.0008 | 55% |
| | If available | 0.03 | 0.34 (0.23-0.46;0.04-0.81) | 0.0021 | 60% | 0.34 (0.24-0.46;0.05-0.80) | 0.0017 | 58% |
| | If available | 0.04 | 0.34 (0.23-0.46;0.04-0.80) | 0.0057 | 71% | 0.34 (0.23-0.45;0.04-0.80) | 0.0059 | 72% |
| | If available | 0.05 | 0.34 (0.23-0.45;0.04-0.80) | 0.01 | 78% | 0.34 (0.23-0.45;0.04-0.79) | 0.0102 | 78% |
| | If available | **0.10** | **0.32 (0.21-0.44;0.03-0.79)** | **0.0314** | **85%** | **0.32 (0.22-0.44;0.04-0.78)** | **0.0306** | **85%** |
| | If available | 0.15 | 0.31 (0.20-0.43;0.03-0.78) | 0.0602 | 91% | 0.31 (0.20-0.42;0.02-0.77) | 0.0559 | 91% |
| | If available | 0.20 | 0.30 (0.19-0.41;0.02-0.77) | 0.093 | 92% | 0.29 (0.18-0.41;0.01-0.76) | 0.0864 | 92% |
| | If available | 0.25 | 0.29 (0.18-0.40;0.01-0.76) | 0.1332 | 94% | 0.27 (0.17-0.38;0.02-0.73) | 0.1239 | 93% |
| | If available | 0.30 | 0.27 (0.17-0.38;0.01-0.74) | 0.1806 | 95% | 0.27 (0.17-0.38;0.02-0.72) | 0.1764 | 94% |
| | If available | 0.35 | 0.26 (0.16-0.37;0.01-0.72) | 0.2373 | 94% | 0.26 (0.16-0.36;0.01-0.71) | 0.2376 | 96% |
| | If available | 0.40 | 0.25 (0.15-0.36;0.00-0.71) | 0.2484 | 95% | 0.25 (0.15-0.35;0.01-0.70) | 0.2461 | 97% |
| | Ignored | 0.01 | 0.35 (0.24-0.47;0.05-0.82) | -0.0013 | 34% | 0.35 (0.24-0.47;0.05-0.81) | -0.0011 | 38% |
| | Ignored | 0.02 | 0.35 (0.24-0.47;0.05-0.81) | 0.0008 | 56% | 0.35 (0.24-0.46;0.05-0.81) | 0.0003 | 52% |
| | Ignored | 0.03 | 0.34 (0.23-0.46;0.05-0.81) | 0.0025 | 63% | 0.35 (0.24-0.46;0.05-0.80) | 0.0026 | 63% |
| | Ignored | 0.04 | 0.34 (0.23-0.46;0.05-0.80) | 0.0061 | 73% | 0.34 (0.23-0.46;0.05-0.79) | 0.0049 | 69% |
| | Ignored | 0.05 | 0.34 (0.23-0.46;0.04-0.80) | 0.0094 | 77% | 0.34 (0.23-0.45;0.04-0.79) | 0.0095 | 78% |
| | Ignored | **0.10** | **0.32 (0.21-0.44;0.03-0.79)** | **0.0313** | **87%** | **0.32 (0.22-0.44;0.04-0.78)** | **0.0309** | **84%** |
| | Ignored | 0.15 | 0.31 (0.20-0.42;0.02-0.78) | 0.0587 | 91% | 0.31 (0.20-0.42;0.03-0.77) | 0.0558 | 92% |
| | Ignored | 0.20 | 0.29 (0.19-0.41;0.02-0.76) | 0.0912 | 93% | 0.29 (0.18-0.40;0.01-0.76) | 0.0839 | 91% |
| | Ignored | 0.25 | 0.28 (0.18-0.40;0.01-0.76) | 0.1313 | 94% | 0.27 (0.17-0.39;0.01-0.74) | 0.1233 | 93% |
| | Ignored | 0.30 | 0.27 (0.16-0.39;0.00-0.74) | 0.1761 | 94% | 0.26 (0.16-0.38;0.01-0.73) | 0.1721 | 94% |
| | Ignored | 0.35 | 0.26 (0.15-0.37;0.00-0.73) | 0.2354 | 95% | 0.25 (0.15-0.36;0.01-0.72) | 0.234 | 95% |
| | Ignored | 0.40 | 0.25 (0.15-0.36;0.00-0.71) | 0.2491 | 95% | 0.24 (0.14-0.34;0.01-0.69) | 0.2351 | 95% |
| Oncology centers | BDs only | 0.01 | 0.31 (0.21-0.42; 0.07-0.69) | 0.0005 | 52% | 0.31 (0.21-0.42; 0.07-0.69) | 0.0005 | 52% |
| | If available | 0.01 | 0.31 (0.21-0.42;0.07-0.69) | -0.0001 | 49% | 0.31 (0.21-0.42;0.07-0.69) | -0.0002 | 47% |
| | If available | 0.02 | 0.31 (0.20-0.42;0.07-0.69) | 0.0028 | 69% | 0.30 (0.20-0.42;0.07-0.69) | 0.0022 | 66% |
| | If available | 0.03 | 0.30 (0.20-0.41;0.06-0.69) | 0.0066 | 80% | 0.30 (0.20-0.41;0.07-0.68) | 0.0062 | 79% |
| | If available | 0.04 | 0.30 (0.20-0.41;0.06-0.69) | 0.0121 | 88% | 0.30 (0.20-0.41;0.06-0.68) | 0.0123 | 88% |
| | If available | 0.05 | 0.30 (0.20-0.41;0.06-0.69) | 0.0176 | 91% | 0.30 (0.20-0.41;0.06-0.68) | 0.0179 | 92% |
| | If available | **0.10** | **0.29 (0.18-0.40;0.05-0.68)** | **0.0461** | **95%** | **0.29 (0.18-0.40;0.05-0.68)** | **0.0458** | **94%** |
| | If available | 0.15 | 0.28 (0.18-0.39;0.04-0.68) | 0.0827 | 96% | 0.28 (0.18-0.39;0.04-0.67) | 0.0819 | 96% |
| | If available | 0.20 | 0.27 (0.17-0.38;0.03-0.66) | 0.123 | 96% | 0.27 (0.17-0.38;0.04-0.66) | 0.1232 | 96% |
| | If available | 0.25 | 0.26 (0.16-0.37;0.02-0.65) | 0.1715 | 96% | 0.26 (0.16-0.36;0.04-0.63) | 0.1677 | 97% |
| | If available | 0.30 | 0.25 (0.15-0.36;0.02-0.63) | 0.229 | 97% | 0.25 (0.16-0.35;0.03-0.61) | 0.2262 | 97% |
| | If available | 0.35 | 0.24 (0.14-0.35;0.01-0.62) | 0.2402 | 97% | 0.24 (0.15-0.34;0.03-0.60) | 0.2406 | 98% |
| | If available | 0.40 | 0.23 (0.14-0.34;0.01-0.60) | 0.2336 | 97% | 0.23 (0.15-0.33;0.03-0.58) | 0.2328 | 98% |
| | Ignored | 0.01 | 0.31 (0.21-0.42;0.07-0.70) | -0.0006 | 43% | 0.31 (0.21-0.42;0.07-0.69) | -0.0005 | 44% |
| | Ignored | 0.02 | 0.31 (0.21-0.41;0.07-0.69) | 0.0028 | 70% | 0.30 (0.20-0.42;0.07-0.69) | 0.0019 | 63% |
| | Ignored | 0.03 | 0.30 (0.20-0.41;0.06-0.69) | 0.0063 | 78% | 0.30 (0.20-0.41;0.06-0.69) | 0.0057 | 76% |
| | Ignored | 0.04 | 0.30 (0.20-0.41;0.06-0.69) | 0.0115 | 87% | 0.30 (0.20-0.41;0.06-0.69) | 0.0113 | 86% |
| | Ignored | 0.05 | 0.30 (0.20-0.41;0.06-0.69) | 0.0171 | 90% | 0.30 (0.20-0.41;0.06-0.69) | 0.0173 | 90% |
| | Ignored | **0.10** | **0.29 (0.18-0.40;0.05-0.68)** | **0.0457** | **95%** | **0.29 (0.18-0.40;0.05-0.68)** | **0.0455** | **94%** |
| | Ignored | 0.15 | 0.28 (0.17-0.39;0.04-0.67) | 0.0806 | 96% | 0.28 (0.17-0.39;0.04-0.67) | 0.0807 | 95% |
| | Ignored | 0.20 | 0.27 (0.16-0.38;0.03-0.66) | 0.121 | 96% | 0.27 (0.17-0.38;0.03-0.66) | 0.121 | 96% |
| | Ignored | 0.25 | 0.26 (0.16-0.37;0.02-0.65) | 0.17 | 96% | 0.26 (0.16-0.37;0.02-0.64) | 0.1681 | 96% |
| | Ignored | 0.30 | 0.25 (0.15-0.36;0.01-0.64) | 0.2264 | 96% | 0.25 (0.15-0.36;0.02-0.62) | 0.2237 | 96% |
| | Ignored | 0.35 | 0.24 (0.14-0.35;0.01-0.62) | 0.2402 | 96% | 0.24 (0.15-0.34;0.02-0.60) | 0.2387 | 97% |
| | Ignored | 0.40 | 0.23 (0.14-0.34;0.01-0.60) | 0.2338 | 96% | 0.23 (0.14-0.33;0.02-0.58) | 0.2271 | 98% |
| Non-Oncology centers | BDs only | 0.01 | 0.09 (0.04-0.18; 0.00-0.47) | -0.0018 | 61% | 0.09 (0.04-0.18; 0.00-0.47) | -0.0018 | 61% |
| | If available | 0.01 | 0.09 (0.04-0.17;0.00-0.45) | -0.0001 | 55% | 0.09 (0.04-0.17;0.00-0.45) | -0.0002 | 54% |
| | If available | 0.02 | 0.09 (0.04-0.17;0.00-0.45) | 0.0072 | 76% | 0.09 (0.04-0.17;0.00-0.45) | 0.0067 | 75% |
| | If available | 0.03 | 0.09 (0.03-0.17;0.00-0.45) | 0.013 | 81% | 0.09 (0.03-0.17;0.00-0.45) | 0.0132 | 82% |
| | If available | 0.04 | 0.08 (0.03-0.17;0.00-0.45) | 0.0206 | 86% | 0.08 (0.03-0.17;0.00-0.45) | 0.0206 | 86% |
| | If available | 0.05 | 0.08 (0.03-0.17;-0.00-0.45) | 0.0291 | 88% | 0.08 (0.03-0.16;-0.00-0.45) | 0.0289 | 88% |
| | If available | **0.10** | **0.07 (0.03-0.16;-0.01-0.43)** | **0.0733** | **90%** | **0.07 (0.03-0.15;-0.00-0.42)** | **0.0737** | **91%** |
| | If available | 0.15 | 0.07 (0.02-0.14;-0.01-0.39) | 0.0691 | 90% | 0.06 (0.02-0.14;-0.01-0.38) | 0.0643 | 90% |
| | If available | 0.20 | 0.06 (0.02-0.13;-0.01-0.36) | 0.0616 | 89% | 0.06 (0.02-0.12;-0.01-0.35) | 0.0552 | 90% |
| | If available | 0.25 | 0.05 (0.02-0.12;-0.01-0.33) | 0.055 | 89% | 0.05 (0.02-0.11;-0.01-0.31) | 0.0516 | 90% |
| | If available | 0.30 | 0.05 (0.01-0.10;-0.02-0.28) | 0.0497 | 89% | 0.05 (0.02-0.10;-0.01-0.27) | 0.049 | 91% |

| | | | 2SS with ADNEX2 | | | 2SS with ADNEX | | |
|---|---|---|---|---|---|---|---|---|
| Subgroup | CA125 | Threshold | NB (95% Crl; PI) | $\Delta NB$ | P-useful | NB (95% Crl; PI) | $\Delta NB$ | P-useful |
| | If available | 0.35 | 0.05 (0.01-0.10;-0.02-0.27) | 0.0452 | 88% | 0.05 (0.02-0.09;-0.01-0.25) | 0.047 | 92% |
| | If available | 0.40 | 0.04 (0.01-0.09;-0.04-0.25) | 0.0429 | 87% | 0.05 (0.02-0.09;-0.01-0.21) | 0.0461 | 94% |
| | Ignored | 0.01 | 0.09 (0.04-0.17;0.00-0.45) | 0.0001 | 56% | 0.09 (0.04-0.17;0.00-0.45) | 0.0001 | 56% |
| | Ignored | 0.02 | 0.09 (0.04-0.17;0.00-0.44) | 0.0062 | 74% | 0.09 (0.04-0.17;0.00-0.44) | 0.0063 | 74% |
| | Ignored | 0.03 | 0.08 (0.03-0.17;0.00-0.44) | 0.0133 | 83% | 0.09 (0.04-0.17;0.00-0.44) | 0.0137 | 83% |
| | Ignored | 0.04 | 0.08 (0.03-0.16;0.00-0.44) | 0.021 | 87% | 0.08 (0.03-0.16;0.00-0.43) | 0.0203 | 86% |
| | Ignored | 0.05 | 0.08 (0.03-0.16;-0.00-0.44) | 0.0289 | 89% | 0.08 (0.03-0.16;0.00-0.43) | 0.0285 | 89% |
| | Ignored | **0.10** | **0.08 (0.03-0.16;-0.00-0.43)** | **0.0743** | **91%** | **0.07 (0.03-0.15;-0.00-0.41)** | **0.0742** | **92%** |
| | Ignored | 0.15 | 0.07 (0.02-0.14;-0.01-0.41) | 0.0677 | 90% | 0.06 (0.02-0.13;-0.00-0.38) | 0.0644 | 91% |
| | Ignored | 0.20 | 0.06 (0.02-0.13;-0.01-0.37) | 0.0617 | 90% | 0.06 (0.02-0.12;-0.01-0.34) | 0.0553 | 90% |
| | Ignored | 0.25 | 0.05 (0.01-0.12;-0.02-0.34) | 0.0548 | 89% | 0.05 (0.02-0.11;-0.01-0.31) | 0.0514 | 91% |
| | Ignored | 0.30 | 0.05 (0.01-0.11;-0.02-0.30) | 0.0492 | 88% | 0.05 (0.02-0.10;-0.01-0.28) | 0.0485 | 91% |
| | Ignored | 0.35 | 0.04 (0.01-0.10;-0.02-0.29) | 0.0448 | 87% | 0.05 (0.02-0.09;-0.02-0.26) | 0.0467 | 92% |
| | Ignored | 0.40 | 0.04 (0.01-0.09;-0.04-0.27) | 0.0438 | 86% | 0.04 (0.01-0.08;-0.02-0.22) | 0.0424 | 92% |

BDs: Benign descriptors; NB: Net Benefit; Crl: Credible interval; PI: Prediction interval, $\Delta NB$: Delta net benefit.CA125 ignored means not including CA125 results in the model; If available means CA125 values are used in the model if available.

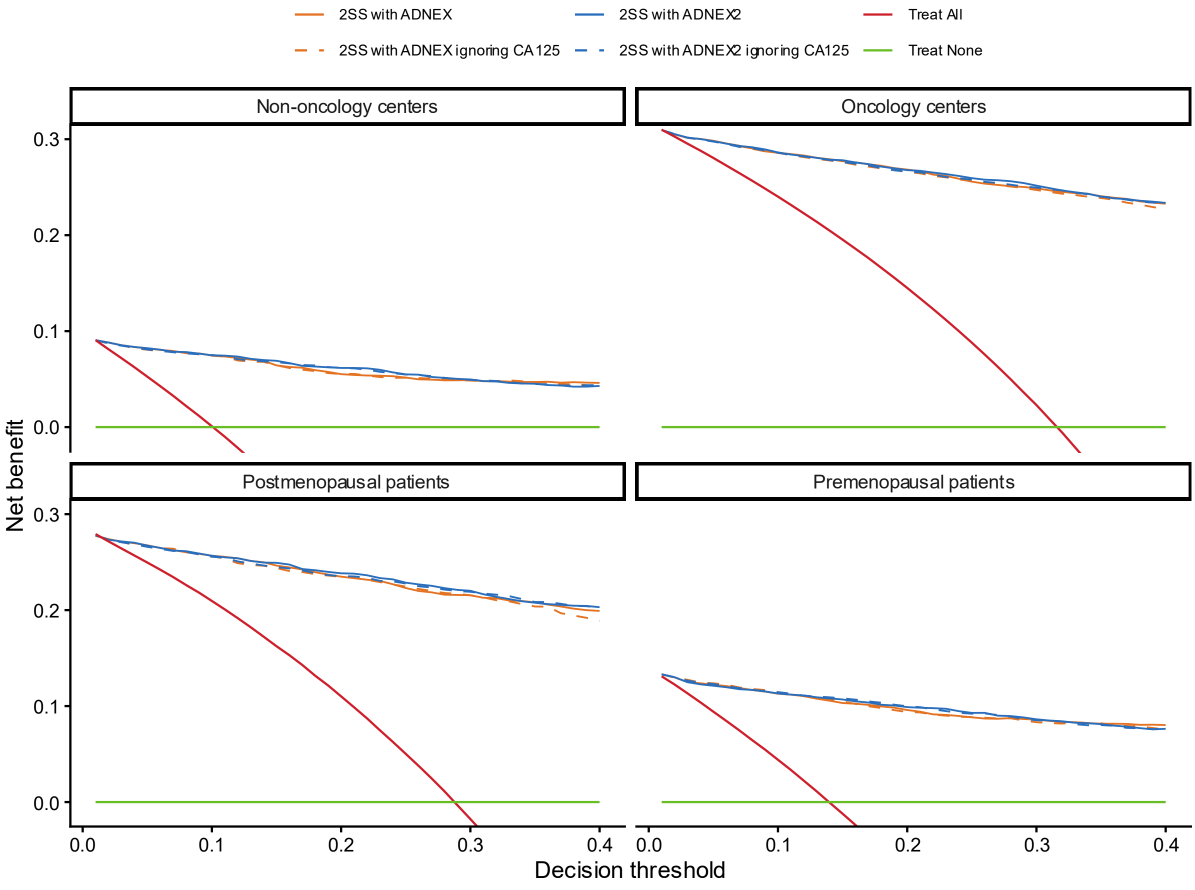

**Figure S78. Decision curve analysis for two-step strategy with ADNEX2 and ADNEX in all patients by subgroup.**

### S6. TRIPOD+AI CHECKLIST

From Collins et al. [20]

#### S6.1 TRIPOD+AI ABSTRACT CHECKLIST

| SECTION | ITEM | CHECKLIST ITEM | REPORTED ON |
|---|---|---|---|
| TITLE | 1 | Identify the study as developing or evaluating the performance of a multivariable prediction model, the target population, and the outcome to be predicted | Title page |
| BACKGROUND | 2 | Provide a brief explanation of the healthcare context and rationale for developing or evaluating the performance of all models | Abstract (Objective) |
| OBJECTIVES | 3 | Specify the study objectives, including whether the study describes model development, evaluation, or both | Abstract (Objective) |
| METHODS | 4 | Describe the sources of data | Abstract (Design, Setting, and Participants) |
| METHODS | 5 | Describe the eligibility criteria and setting where the data were collected | Abstract (Setting / Participants) |
| METHODS | 6 | Specify the outcome to be predicted by the model, including time horizon of predictions in case of prognostic models | Abstract (Objective) |
| METHODS | 7 | Specify the type of model, a summary of the model-building steps, and the method for internal validation | Abstract (Interventions) |
| METHODS | 8 | Specify the measures used to assess model performance (eg, discrimination, calibration, clinical utility) | Abstract (Main outcome measures) |
| RESULTS | 9 | Report the number of participants and outcome events | Abstract (Results) |
| RESULTS | 10 | Summarise the predictors in the final model | Abstract (Interventions) |
| RESULTS | 11 | Report model performance estimates (with confidence intervals) | Abstract (Results) |
| DISCUSSION | 12 | Give an overall interpretation of the main results | Abstract (Conclusions) |
| REGISTRATION | 13 | Give the registration number and name of the registry or repository | Abstract (Trial registration) |

#### S6.2 TRIPOD+AI CHECKLIST

| SECTION/TOPIC | ITEM | CHECKLIST ITEM | REPORTED ON |
|---|---|---|---|
| TITLE | 1 | Identify the study as developing or evaluating the performance of a multivariable prediction model, the target population, and the outcome to be predicted | Title |
| ABSTRACT | 2 | See TRIPOD+AI for Abstracts checklist | See S6.1 |
| INTRODUCTION (BACKGROUND) | 3a | Explain the healthcare context (including whether diagnostic or prognostic) and rationale for developing or evaluating the prediction model, including references to existing models | Section 1 (Introduction) |
| | 3b | Describe the target population and the intended purpose of the prediction model in the context of the care pathway, | Section 1 (Introduction) |

| | | | |
|---|---|---|---|
| | | including its intended users (e.g., healthcare professionals, patients, public) | |
| | 3c | Describe any known health inequalities between sociodemographic groups | Section 4.2 (Strengths and limitations) |
| **INTRODUCTION (OBJECTIVES)** | 4 | Specify the study objectives, including whether the study describes the development or validation of a prediction model (or both) | Section 1 (Introduction) |
| **METHODS (DATA)** | 5a | Describe the sources of data separately for the development and evaluation datasets (e.g., randomised trial, cohort, routine care or registry data), the rationale for using these data, and representativeness of the data | Section 2.1 (Design and participants) |
| | 5b | Specify the dates of the collected participant data, including start and end of participant accrual; and, if applicable, end of follow-up | Section 2.1 (Design and participants) |
| **METHODS (PARTICIPANTS)** | 6a | Specify key elements of the study setting (e.g., primary care, secondary care, general population) including the number and location of centres | Section 2.1 (Design and participants), Table S1 |
| | 6b | Describe the eligibility criteria for study participants | Section 2.1 (Design and participants) |
| | 6c | Give details of any treatments received, and how they were handled during model development or evaluation, if relevant | Section 2.2 (Procedures and variable measurements) |
| **METHODS (DATA PREPARATION)** | 7 | Describe any data pre-processing and quality checking, including whether this was similar across relevant sociodemographic groups | Section 2.4 (Data cleaning and preparation), Supp S1 (Exclusion of centres) |
| **METHODS (OUTCOME)** | 8a | Clearly define the outcome that is being predicted and the time horizon, including how and when assessed, the rationale for choosing this outcome, and whether the method of outcome assessment is consistent across sociodemographic groups | Section 2.3 (Outcome), Table 1 |
| | 8b | If outcome assessment requires subjective interpretation, describe the qualifications and demographic characteristics of the outcome assessors | Section 2.2 (Procedures and variable measurements) , Section 2.3 (Outcome) |
| | 8c | Report any actions to blind assessment of the outcome to be predicted | Section 2.2 (Procedures and variable measurements), Section 2.3 (Outcome) |
| **METHODS (PREDICTORS)** | 9a | Describe the choice of initial predictors (e.g., literature, previous models, all available predictors) and any pre-selection of predictors before model building | Section 2.5 (Statistical analysis) |
| | 9b | Clearly define all predictors, including how and when they were measured (and any actions to blind assessment of predictors for the outcome and other predictors) | Section 2.2 (Procedures and variable measurements), Section 1 (Introduction), Supp S2.2, Supp S7 |
| | 9c | If predictor measurement requires subjective interpretation, describe the qualifications and demographic characteristics of the predictor assessors | Section 2.2 (Procedures and variable measurements) |
| **METHODS (SAMPLE SIZE)** | 10 | Explain how the study size was arrived at (separately for development and evaluation), and justify that the study size was sufficient to answer the research question. Include details of any sample size calculation | Section 2.5 (Statistical analysis), Supp S2.1 (Sample size), Tables S2, S3 |
| **METHODS (MISSING DATA)** | 11 | Describe how missing data were handled. Provide reasons for omitting any data | Section 2.5 (Statistical analysis), Supp S2.2 (Missing data handling) |
| | 12a | Describe how the data were used (e.g., for development and evaluation of model performance) in the analysis, | Section 2.5 (Statistical analysis) |

| | | | |
|---|---|---|---|
| METHODS (ANALYTICAL METHODS) | | including whether the data were partitioned, considering any sample size requirements | |
| | 12b | Depending on the type of model, describe how predictors were handled in the analyses (functional form, rescaling, transformation, or any standardisation). | Section 2.5 (Statistical analysis), Supp S2.3.1 (Model updating) |
| | 12c | Specify the type of model, rationale, all model-building steps, including any hyperparameter tuning, and method for internal validation | Section 2.5 (Statistical analysis), Supp S2.3.1 (Model updating) |
| | 12d | Describe if and how any heterogeneity in estimates of model parameter values and model performance was handled and quantified across clusters (e.g., hospitals, countries). See TRIPOD-Cluster for additional considerations | Section 2.5 (Statistical analysis), Supp S2.3 (Methodological details) |
| | 12e | Specify all measures and plots used (and their rationale) to evaluate model performance (e.g., discrimination, calibration, clinical utility) and, if relevant, to compare multiple models | Section 2.5 (Statistical analysis), Supp S2.3 (Methodological details) |
| | 12f | Describe any model updating (e.g., recalibration) arising from the model evaluation, either overall or for particular sociodemographic groups or settings | Section 2.5 (Statistical analysis), Supp S2.3.1 (Model updating) |
| | 12g | For model evaluation, describe how the model predictions were calculated (e.g., formula, code, object, application programming interface) | Section 2.5 (Statistical analysis), Section 2.2 (last paragraph), Supp S2.3.1 |
| **METHODS (CLASS IMBALANCE)** | 13 | If class imbalance methods were used, state why and how this was done, and any subsequent methods to recalibrate the model or the model predictions | N/A |
| **METHODS (FAIRNESS)** | 14 | Describe any approaches that were used to address model fairness and their rationale | Section 4.2 (Strengths and limitations) |
| **METHODS (MODEL OUTPUT)** | 15 | Specify the output of the prediction model (e.g., probabilities, classification). Provide details and rationale for any classification and how the thresholds were identified | Section 1 (Introduction), Section 4.4 (Results in context) |
| **METHODS (TRAINING VERSUS EVALUATION)** | 16 | Identify any differences between the development and evaluation data in healthcare setting, eligibility criteria, outcome, and predictors | Supp S4.1, Table S10 |
| **METHODS (ETHICAL APPROVAL)** | 17 | Name the institutional research board or ethics committee that approved the study and describe the participant-informed consent or the ethics committee waiver of informed consent | Section 2.1 (Design and participants) |
| **OPEN SCIENCE (FUNDING)** | 18a | Give the source of funding and the role of the funders for the present study | Section 5 (Open science - Funding) |
| **OPEN SCIENCE (CONFLICTS OF INTEREST)** | 18b | Declare any conflicts of interest and financial disclosures for all authors | Section 5 (Open science - Conflicts of interest) |
| **OPEN SCIENCE (PROTOCOL)** | 18c | Indicate where the study protocol can be accessed or state that a protocol was not prepared | Section 5 (Open science - Protocol) |
| **OPEN SCIENCE (REGISTRATION)** | 18d | Provide registration information for the study, including register name and registration number, or state that the study was not registered | Abstract, Section 5 (Open science - Protocol) |
| **OPEN SCIENCE (DATA SHARING)** | 18e | Provide details of the availability of the study data | Section 5 (Open science - Data sharing) |

| | | | |
|---|---|---|---|
| **OPEN SCIENCE (CODE SHARING)** | 18f | Provide details of the availability of the analytical code | Section 5 (Open science - Code sharing) |
| **PATIENT & PUBLIC INVOLVEMENT** | 19 | Provide details of any patient and public involvement during the design, conduct, reporting, interpretation, or dissemination of the study or state no involvement. | Section 2.7 (Patient and public involvement) |
| **RESULTS (PARTICIPANTS)** | 20a | Describe the flow of participants through the study, including the number of participants with and without the outcome and, if applicable, a summary of the follow-up time. A diagram may be helpful. | Section 3 (Results), Figure 1 |
| | 20b | Report the characteristics overall and, where applicable, for each data source or setting, including the key dates, key predictors (including demographics), treatments received, sample size, number of outcome events, follow-up time, and amount of missing data. A table may be helpful. Report any differences across key demographic groups. | Section 3 (Results), Table 1, Table 2, Supp S4.1 (Tables S4-S9), Supp S5.1 (Table S16) |
| | 20c | For model evaluation, show a comparison with the development data of the distribution of important predictors (demographics, predictors, and outcome). | Supp S4.1, Table S10 |
| **RESULTS (MODEL DEVELOPMENT)** | 21 | Specify the number of participants and outcome events in each analysis (e.g., for model development, hyperparameter tuning, model evaluation) | Section 3 (Results), Table 1, Table 2, Figure 1, Supp S4.1 (Tables S6-S7) |
| **RESULTS (MODEL SPECIFICATION)** | 22 | Provide details of the full prediction model (e.g., formula, code, object, application programming interface) to allow predictions in new individuals and to enable third-party evaluation and implementation, including any restrictions to access or re-use (e.g., freely available, proprietary) | Section 2.6 (Future independent external validation) |
| **RESULTS (MODEL PERFORMANCE)** | 23a | Report model performance estimates with confidence intervals, including for any key subgroups (e.g., sociodemographic). Consider plots to aid presentation. | Section 3.2 (ADNEX2), Table 3, Figures 2-3, Supp S4.4, S4.5, S4.6, S4.7, Supp S4.3 (Table S11), Supp S5 (Tables S17-S19, Figures S47-S78) |
| | 23b | If examined, report results of any heterogeneity in model performance across clusters. See TRIPOD-Cluster for additional details. | Section 3.2 (ADNEX2), Table 3, Supp S4.4 (Figures S12-S24), Supp S4.5 (Figures S25-S36), Supp S4.3 (Table S11), Supp S4.5 (Table S13), Supp S4.7 (Table S15), Supp S5 (Figures S47-S77, Table S19) |
| **RESULTS (MODEL UPDATING)** | 24 | Report the results from any model updating, including the updated model and subsequent performance | Section 3.2 (ADNEX2) |
| **DISCUSSION (INTERPRETATION)** | 25 | Give an overall interpretation of the main results, including issues of fairness in the context of the objectives and previous studies | Section 4 (Discussion) |
| **DISCUSSION (LIMITATIONS)** | 26 | Discuss any limitations of the study (such as a non-representative sample, sample size, overfitting, missing data) and their effects on any biases, statistical uncertainty, and generalizability | Section 4.2 (Strengths and limitations) |
| **DISCUSSION (USABILITY)** | 27a | Describe how poor quality or unavailable input data (e.g., predictor values) should be assessed and handled when implementing the prediction model | Section 4.4 (Results in context) |
| | 27b | Specify whether users will be required to interact in the handling of the input data or use of the model, and what level of expertise is required of users | Section 2.2 (Procedures and variable measurements) |
| | 27c | Discuss any next steps for future research, with a specific view to applicability and generalizability of the model | Section 4.5 (Future research) |

### S7. STUDY PROTOCOL

### International Ovarian Tumour Analysis (IOTA) Phase 5

#### A multicentre study to examine the short and long term outcomes of the conservative management of benign-looking adnexal masses and the pre-operative characterisation of ovarian tumours

STUDY CO-ORDINATORS

Tom Bourne, Lil Valentin, Dirk Timmerman

Contact details:
Dirk Timmerman, MD, PhD
Department of Obstetrics and Gynaecology, University Hospitals Leuven,
Herestraat 49, B-3000 Leuven, BELGIUM.
Telephone: + 32 16 344201 (office) Fax: + 32 16 344205
+ 32 16 344215 (secretary)


STEERING COMMITTEE

Dirk Timmerman (University Hospitals, KU Leuven)
Tom Bourne (Imperial College, London)
Antonia C. Testa (Università Cattolica di Sacro Cuore, Roma)
Lil Valentin (University of Lund / Malmö)
Ben Van Calster (KU Leuven)
Sabine Van Huffel (ESAT-SISTA, KU Leuven)
Ignace Vergote (University Hospitals, KU Leuven)

Summary

The medium to long term behaviour of benign-looking adnexal masses that do not undergo surgery is unknown. It is possible for these masses to undergo malignant transformation, rupture or torsion. Furthermore they may undergo changes in volume and/or morphology that may or may not predict any of these behaviours. To date, no research has rigorously investigated the long-term behaviour of such masses. Consequently, there are no evidence-based guidelines on the optimal management of the majority of adnexal tumours. It is therefore not surprising that clinical practice is highly variable, with some clinicians preferring to operate on virtually any mass. When a clinician decides not to operate, the time intervals selected for follow up scans is often arbitrarily chosen. On the other hand, we do have some convincing data to suggest that simple cysts are rarely malignant and so it is generally thought that operating on these common tumours is probably not necessary and simply increases costs and morbidity. Developing new insights into the natural history of benign looking conservatively managed ovarian masses would potentially change the management of thousands of women, by avoiding surgery or even further surveillance for some and detecting cancer earlier or even preventing it for others.

In this international multicentre study IOTA phase 5 we aim to develop the optimal evidence-based algorithm for the management of all adnexal tumours in order to improve the detection of ovarian cancer while at the same time reducing the number of unnecessary operations. At least three thousand patients with an adnexal mass will undergo an ultrasound examination and if no operation is needed they will be followed up for at least 5 years. At each visit the investigator will assess the tumour and decide whether surgery is necessary based on the available information and local protocols. Survival and logistic regression analysis will be used to develop decision aids to assist clinicians in making decisions regarding surgery and follow up.

**Relation with other IOTA studies**

The **IOTA study (International Ovarian Tumour Analysis)** is a multicentre collaborative project for the pre-operative characterisation of ovarian tumours..

**IOTA phase 1:** The **first phase** of IOTA was conducted between 1999 and 2002. Several new mathematical models were developed based on the prospectively collected data of 1066 patients with a persisting adnexal tumour from 9 European centres (1). Between 2002 and 2005 three centres continued the prospective collection in order to be able to perform an internal validation of mathematical models developed in IOTA phase 1. In this so-called IOTA **phase 1b** study a dataset of 507 new patients was prospectively collected in 3 out of the 9 original IOTA centres (2). All models proved to perform excellently with areas under the ROC curves of more than 0.94.

**IOTA phase 2:** The **second phase** of IOTA consisted of an external validation of the models and this was conducted between 2005 and 2007. The diagnostic algorithms were prospectively validated on 1938 patients with adnexal tumours in 19 centres in Belgium, Italy, UK, Sweden, Poland, Czech Republic, Canada, and China (3). A first analysis showed that overall performance of the logistic regression models was excellent (area under the ROC curve 0.94). We concluded that a subgroup of “uncertain” tumours needs a reliable second stage test in order to help even experienced ultrasound examiners.

**IOTA phase 3:** The **third phase** of the IOTA study started in 2010.

The aim was to validate the added value of mathematical models as new diagnostic tool in the prediction of ovarian cancer in clinical practice in centres that were involved in IOTA phase 1 or 2. It is a temporal validation of IOTA mathematical models as a first stage examination. However in cases where the prediction is unreliable, we aim to further improve the predictive performance of this diagnostic tool with second stage tests, such as new sets of tumor markers, proteomics and three-dimensional Power Doppler ultrasonography.

**IOTA phase 4:** Randomised controlled trial in 7 London hospitals. Clinical implementation of IOTA logistic regression models LR2 vs. the routinely used Risk of Malignancy Index. Assessment of efficacy, referral pattern and costs.

Introduction

There is very little evidence on which to base a recommendation on how apparently benign looking adnexal masses should be managed. Because the natural history of such adnexal masses is not known, and because of the fear of "missing" ovarian cancer, many adnexal masses are currently surgically removed, even if they do not manifest any signs of malignancy. This is not optimal, because every surgical procedure is associated with risks of both short-term and long-term complications, for example pulmonary embolism, deep vein thrombosis, and bowel perforation or obstruction (4). Furthermore we do not know if benign ovarian lesions impact on fertility, although we know that surgery on ovaries may cause adhesions, which in turn may cause infertility, chronic pelvic pain and bowel obstruction. We do not know the frequency of these complications, nor do we know how often benign adnexal masses are associated with complications such as torsion if they are not removed. There is some evidence, however, that expectant management of presumed ovarian dermoid cysts less than 6 cm is safe and does not seem to interfere with pregnancy or delivery (5-7). The available data also suggest that expectant management of simple cysts less than 5 cm in post-menopausal women is a safe strategy (8,9).

On the other hand ovarian cancer is associated with a high mortality rate and significant morbidity. It is the fifth leading cause of cancer-related deaths (10). The lifetime risk of developing ovarian cancer is around 1 in 50 to 1 in 70 (11,12). Every year more than 200,000 new cases are diagnosed worldwide. The disease has a poor prognosis, with five-year relative survival strongly depending on disease stage (12,13). For example, Cancer Research UK reports survival rates of 73% for stage I versus 16% for stage IV ovarian cancer.

New diagnostic strategies to decrease mortality are needed, as treatment advances have not decreased mortality over the past 20 years (14). Effective screening programmes may help, but current candidate tests remain unsatisfactory (15). A crucial issue is that ovarian cancer is typically asymptomatic in its early stages. Screening algorithms have generally resulted in high sensitivity at the cost of a large number of false positives. As a result a large number of surgical interventions are made in order to find relatively few cancers. However although one study has suggested that there is no benefit in removing benign ovarian tumors (16), there are no conclusive data to inform us regarding the long term behaviour of presumed benign ovarian cysts left in situ. Should benign cysts have malignant potential, then a policy of removing such masses may have a significant impact on mortality from this disease.

Research directed towards the use of diagnostic tests and models to predict malignancy in ovarian tumours has focused on masses that have been subsequently surgically removed in order to provide a clear histological end point. Clinicians decide whether to operate on an ovarian mass depending on a number of factors. These may include the subjective characterisation of the mass using ultrasound, the use of simple models such as the risk of malignancy index, the age of the patient, the serum CA 125 level and the presence or absence of symptoms such as pain. The management of cysts that are not removed surgically is not evidence based and often subject to wide variation. In the absence of rigorous follow up data, we do not know how many false negative results for cancer are associated with these cysts, or if they sometimes undergo malignant transformation. We will only gain this knowledge by long term systematic follow up of a large cohort of ovarian cysts.

A number of studies have focused on the prediction of malignancy in surgically removed masses (for overviews, see references 2,17-19). There is strong scientific evidence that subjective evaluation of a mass using ultrasound by an experienced examiner is a very good method for discriminating between benign and malignant adnexal masses (2,3,20-21), and that a correct histological diagnosis can be suggested on the basis of ultrasound findings in many cases (22,23). We have previously established the International Ovarian Tumor Analysis (IOTA) group to develop and validate prediction models based on large, multi-centre datasets with standardised definitions and data collection procedures (25). The aim was to develop robust models to predict malignancy that performed well, and were widely generalisable. In doing so we aimed to overcome the shortcomings of earlier studies such as small sample sizes, single centre recruitment, and lack of standardised data collection. These models (1, 25-27) successfully passed temporal and external validation (2,3,28). Following these validation studies, we selected two logistic regression models for further study. The first (LR1) is a model with 12 predictors, the second (LR2) contains only six predictors. Even in postmenopausal women conservative management and sonographic follow up of incidental unilocular and multilocular cysts <7cm may be a valuable option (29).

#### Objectives

The general aim of this study is the development of the optimal algorithm for the management of all adnexal masses. This can be broken down into different specific objectives: 1) to study the occurrence of complications such as rupture, torsion, or malignancy in patients with benign looking conservatively treated masses; 2) to test the published IOTA diagnostic models for predicting that a mass is malignant at first visit or benign (either on the basis of histology following surgery or by the absence of malignant features on an ultrasound scan one year after the initial visit), and to predict complications (e.g. ocurrence of malignancy and other) during long-term follow-up; 3) to investigate factors that may be related to the need for surgery during long-term follow up; 4) to study the natural history of conservatively treated benign looking masses and to establish descriptive curves of the longitudinal changes seen in parameters from conservatively managed benign tumors (e.g. change in diameter, size of any solid component, number of papillations, or color score). We hope these curves will allow us to determine if any particular growth pattern is associated with complications or malignancy

Related to these four objectives, we aim to carry out the following analyses: 1) descriptive analysis of complications overall and by participating center (anonymised) and overall Kaplan-Meier curves for the need for surgery among benign looking masses; 2) estimation of discriminatory ability of LR1 for malignancy at the initial visit using the c-index and ROC curves and for complications during long-term follow-up using the hazard ratio and c-index within the context of survival analysis; 3) survival analysis to investigate predictors of the need for surgery during long-term follow-up of non-operated masses; 4) the development of longitudinal curves of the changes seen in the characteristics of non-operated masses using longitudinal analysis techniques such as mixed models and functional linear discriminant analysis (FLDA).

Methods

**Study design**

International multicenter prospective observational cohort study

**Eligible for inclusion**

Any woman at least 18 years old with an adnexal mass.

Any mass with benign ultrasound morphology may be suitable for conservative management.

Pregnant patients can be included, but their data will be analysed separately.

**Exclusion Criteria**

.

Cysts that are deemed to be clearly physiological and less than 3 cm in maximum diameter are not eligible for inclusion.

Any cyst with features of malignancy is excluded from the conservative management

The denial or withdrawal of oral informed consent

**Official approval by the Ethics Committee**

The multicentre project IOTA phase 5 will be submitted to the Ethics Committee of the University Hospitals Leuven as main investigating centre as well as in each participating centre.

The study will be performed in accordance with generally accepted standards of Good Clinical Practice and the investigators will adhere to all applicable laws and regulations governing the conduct of clinical trials, including but not limited to the ICH Harmonized Tripartite Guidelines for Good Clinical Practice and the Declaration of Helsinki (2008).

**Insurance policy**

This multicentre international study is initiated by the University Hospitals Leuven, Belgium. Each participating centre outside Belgium is fully responsible for patient care within its own hospital in agreement with local laws. Each centre is also responsible for all legal aspects of patient care and for its own insurance for all matters related to this study.

**Financial Support**

The IOTA phase 3 project is supported by an Applied Biomedical Research grant (Toegepast Biomedisch Onderzoek, TBM) from the Flanders Institute for Scientific and Technological Research: IWT Flanders, Belgium (IWT-TBM 070706). This grant covers costs of central data collection, proteomic analysis, analysis of new tumour markers and statistical analyses. For IOTA phase 5 we received a research grant for a doctoral researcher by the Flemish Fund for Scientific Research (FWO Vlaanderen 06260, IOTA5).

There is no financial compensation for principal investigators nor patients.

**Definition of benign ultrasound morphology**

This is defined on the basis of subjective assessment of ultrasound findings by an experienced ultrasound examiner. Only lesions where the ultrasound examiner is certain or almost certain that the lesion is benign can be managed conservatively. The management of benign masses will be decided according to local protocols.

**Number of study patients and recruitment period**

This is an observational study and therefore a sample size cannot be calculated. We aim to collect at least 3000 women with an adnexal mass and at least 1000 women with an adnexal mass managed conservatively. We plan an initial recruitment period of eighteen months. Patients will be followed up for at least 5 years, unless surgical intervention is necessary.

**Follow-up**

Ultrasound (and clinical) follow up will be organised by the ultrasound examiner who entered the patient into the study. Follow-up will be after 3 months (maximal range 1-4 months), 6 months (maximal range 4-8 months) and then every 12 months (maximal range 10-14 months).

Although measurement of serum CA 125 levels is encouraged, it is not mandatory for inclusion in the study. If CA125 is measured it should be recorded in the study screen and preferably measured on each visit.

**Duration of follow-up**

A yearly analysis will be carried out in order to evaluate acute complications. The duration of follow up will be for at least 5 years and is not limited as long as the patient is compliant with the study and the study is ongoing.

Departments (e.g. radiology departments) that are not involved with clinical decision making about plans for follow up or surgery cannot participate in the full IOTA 5 study. In these centres data can be prospectively collected as an observational study. Only patients with appropriate outcome measures (i.e. follow up ultrasonography after one or more years or patients with complete details of clinical history or surgical procedures) will be included in any statistical analysis.

**Collection of clinical data**

Family history: Number of first degree relatives with ovarian cancer (0-...) Medical history: Personal history of ovarian cancer and breast cancer

Age (years)

Previous hysterectomy (yes/no) Previous oophorectomy (yes/no)

Contraception (drop down list) (None/oral combined contraceptive pill/progestogen only pill/ patch/vaginal ring/Mirena coil/copper IUD) Hormonal therapy (yes, no).

Is the patient currently wishing to conceive? (yes/no) Menopausal status (pre- or postmenopausal)

History of subfertility? Yes/no

History of ovarian stimulation for subfertility? Yes/no

<u>For ALL patients before menopause two extra questions pop up:</u>

Patient is currently pregnant? (No/Yes)

Patient became pregnant during the last year? (No/Yes)

If Yes: Outcome of pregnancy (it should be possible to enter more than one date, should there be more than one pregnancy during follow-up):

Ongoing pregnancy

Miscarriage; date: …

Ectopic; date: …

Termination; date: …

Delivery; date: …

Complications from the lesion during pregnancy? No/Yes (pop up list):

Acute pain

Chronic pain

Suspected torsion

Infection

Haemorrhage related to the cyst

Cyst rupture

Required surgery

Other, please specify: ….

Complications from the lesion during delivery? No/Yes (pop up list):

Acute pain

Suspected torsion

Haemorrhage

Cyst rupture

Obstructed labour

Malpresentation (e.g. breech or unstable lie)

Other, please specify: ….….

**Ultrasound examination**

A standardized ultrasound examination following the IOTA protocol is carried out.

All ultrasound variables are included in the dedicated software. In the database 0 always means NO and 1 always means YES.

The adnexal lesion is that part of an ovary or of an adnexal mass that is judged by ultrasonography to be not consistent with normal physiology. This can be a persistent unilocular cyst, surrounded by normal looking ovarian stroma with some follicles. In this case the whole ovary containing the cyst is the 'ovary', whereas the unilocular cyst is the 'lesion'. Both are measured and the cyst is described as being 'unilocular' and not 'unilocular-solid'. In other cases the lesion is separate from the ovary (e.g. hydrosalpinx). Again, both ovary and lesion are measured separately. In other cases no normal ovarian stroma is seen. In these cases the lesion and the ovary are undistinguishable and the measurement of lesion and ovary will be the same.

Measurements (in mm): The ovary in two perpendicular planes

The lesion in two perpendicular planes

The volume of the tumor is calculated from the three diameters in two perpendicular planes

The presence of ascites (*i.e.fluid outside* the pouch of Douglas) is noted (yes/no).

Fluid in the pouch of Douglas is measured in the sagittal plane (the largest anteroposterior diameter is given).

(see Figure)

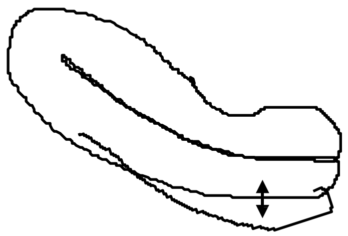

An incomplete septum (as seen in hydrosalpinges) is defined as a thin strand of tissue running across the cyst cavity from one internal surface to the contralateral side, butis not complete in some scanning planes. If a cyst only has incomplete septa, it is unilocular, despite the fact that in certain sections the cyst appears to be multilocular.

Solid means echogenicity suggesting the presence of tissue (e.g. the myometrium, the ovarian stroma, myomas, fibromas). Blood clots and the presence of solid tissue can be distinguished by looking for internal movement when gently pushing the structure with the transducer. The presence of blood flow (with the appropriate color Doppler settings) is diagnostic for solid tissue. The absence of flow is not definitive . In cases of doubt the lesion should be classified as solid.

Solid papillary projections are defined as any solid projections into the cyst cavity from the cyst wall greater than or equal to 3 mm in height

Base

Height

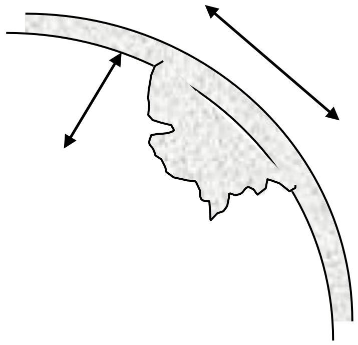

If it is unsure whether solid papillary projections or an incomplete septum are present, the 'worse case scenario' is used. E.g. 'cogwheel excrescences' and 'beads-on-a-string' (as seen in hydrosalpinges) should be classified as papillary excrescences if their height is greater than or equal to 3 mm. The 'white ball' in a dermoid (i.e. Rokitansky node), should not be classified as a solid papillary projection.

*The 'sludge' on the internal walls of endometriotic cysts is not regarded as a papillary projection. In these cases the internal walls are usually 'irregular'.*

The number of separate papillary projections is noted (1/2/3/more).

The presence of flow within some of these projections is noted (yes/no).

Solid papillary projections are described as being 'smooth' or 'irregular' (e.g. cauliflower-like). In some cases it is difficult to judge whether it is a papillary projection and from which point to measure the projection. In these cases it may be helpful to use an imaginary line as shown in the following schematic drawing:

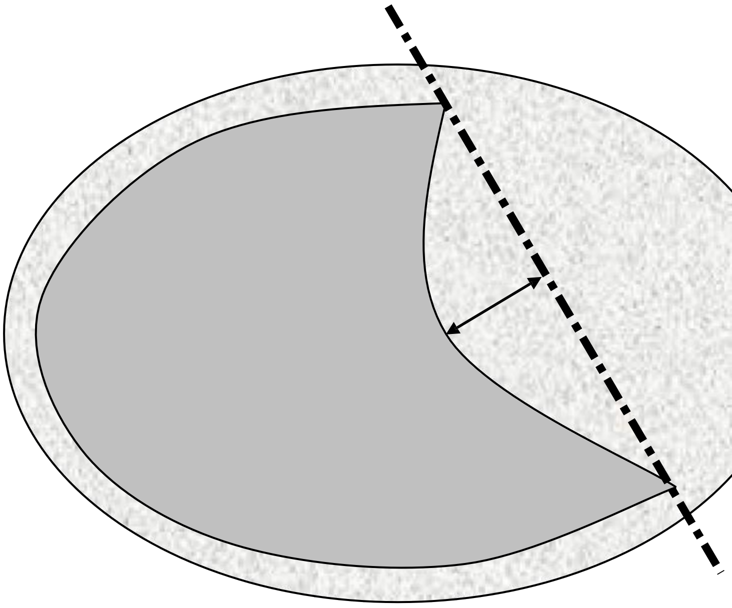

All lesions are qualitatively classified into one of 5 categories:

unilocular (a unilocular cyst without septa and without solid parts or papillary structures). Normal ovarian stroma is not regarded as 'solid' (e.g. a peritoneal cyst, containing a normal ovary, is unilocular and not unilocular-solid).

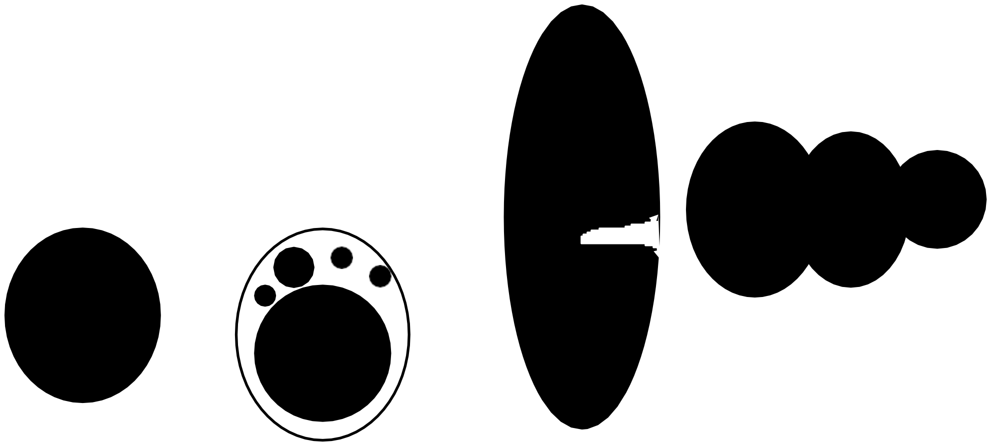

incomplete septum; e.g. in hydrosalpinx)

unilocular cyst with solid component (a unilocular cyst with a measurable solid component or at least one papillary structure). This category may include pyo- or hydrosalpinges with the so-called 'beads-on-a-string' or 'cogwheel' appearance if ≥ 3 mm. If the solid part contains very

small cysts the mass might be unilocular-solid (see below).

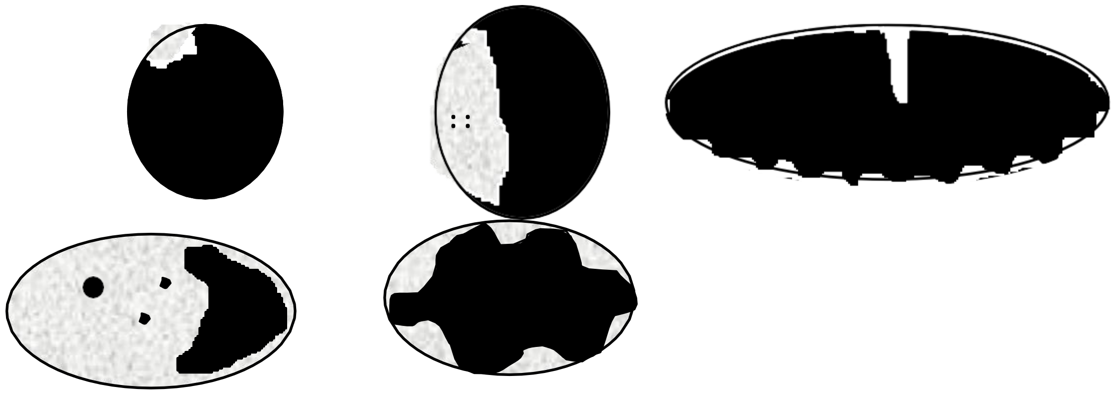

multilocular (a cyst with at least one septum but no measurable solid components or papillary projections). The 'lesion' is measured as indicated by the arrows.

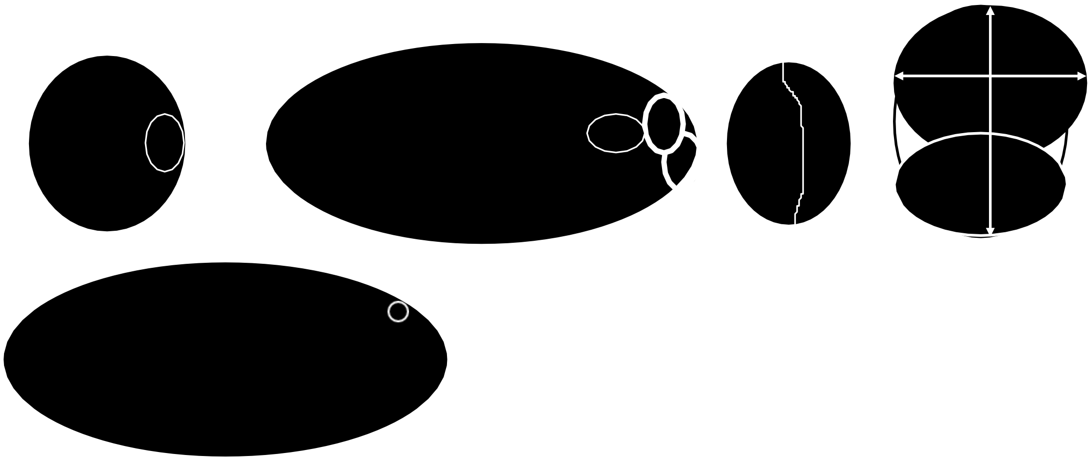

<u>multilocular with solid component</u> (a multilocular cyst with a measurable solid component or at least one papillary structure)

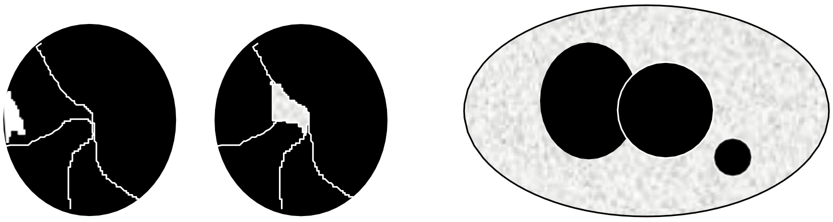

<u>solid</u> (a tumour where the solid components comprise 80% or more of the tumour when assessed in a two-dimensional section).

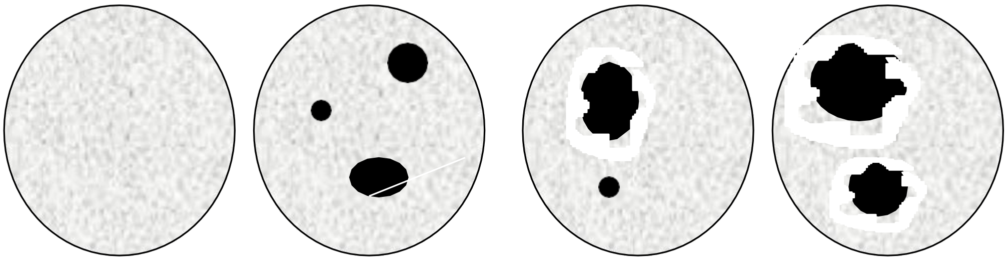

(solid tumour with an irregular cyst wall) A solid tumour may contain papillary projections protruding into the small cysts.

#### Quantitative assessment of morphology

In cystic-solid tumours the largest solid component is measured separately (in three perpendicular planes). The solid component is noted as being smooth or irregular (e.g. cauliflower-like). In some cases a solid papillary projection is the largest solid component and thus the papillary projection is recorded both as papillary projection and as solid component.

The internal wall is also noted as being smooth or irregular.

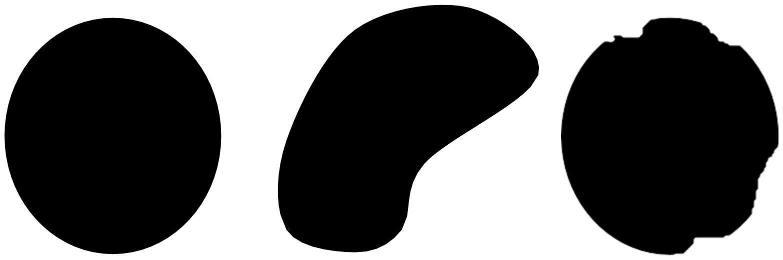

Smooth Smooth Irregular

If there is a solid papillary projection, then the wall is irregular by definition.

The external wall of tumors are not examined unless they are solid.

In cases of solid tumours the description of the internal wall being smooth or irregular is usually not applicable but the outline of the tumour is described as smooth or irregular.

If there is any irregularity in either the inner wall of any cyst or in the outer wall of a solid tumour or on the surface or echogenicity of a solid component, the lesion is described as ‘irregular’.

The dominant feature of the cystic contents is described as anechoic (black), low-level echogenic (homogeneous low level echogenic as seen in mucinous tumours), ‘ground glass’ appearance (homogeneously dispersed echogenic cystic contents, as often seen in endometriotic cysts), hemorrhagic (with internal thread-like structures, representing fibrin strands; it is possible to describe the echogenicity as star-shaped, cobweb-like or jelly-like) or mixed echogenic (as often seen in teratomas) (see images attached).

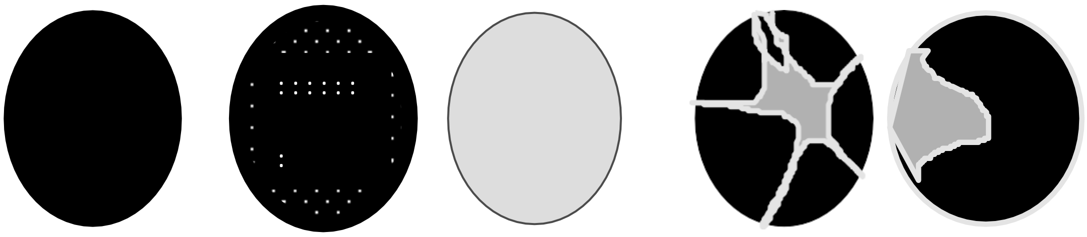

Anechoic Low level Ground glass Hemorrhagic Hemorrhagic

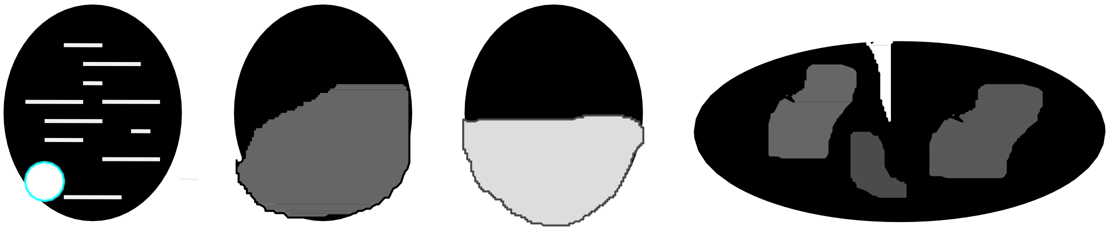

Mixed Mixed (old blood-fluid level Mixed (e.g. abscess) or fat-fluid level)

The presence of acoustic shadows, defined as loss of acoustic echo behind a sound-absorbing structure, is noted as well. Solid tumours are identified by the appearance of the internal texture, by the absence of internal movement when moving the transducer or by colour Doppler imaging (presence of central flow).

In solid tumours the dominant feature of any cystic contents is described only if it can be assessed.

‘Ovarian crescent sign’, defined as the presence of normal ovarian tissue adjacent to an adnexal tumour. (“absent” or “present”, mandatory new variable for phase 3 and 5)

Ultrasound evidence of metastases (e.g. “omental cake” or peritoneal tumoural implants). (“absent” or “present”, mandatory new variable for phase 3 and 5)

#### Colour Doppler imaging and blood flow indices

Subsequently, the entire tumor is surveyed by CDI. The power, gain and pulse repetition frequency are initially adjusted for maximum sensitivity of low blood flow states. The lowest velocity signals

are filtered out by gradually increasing the pulse repetition frequency and flow analysis is concentrated on the highest velocity signals. A subjective semiquantitative assessment of the amount of blood flow (area and colour scale) within the septa, cyst walls, or solid tumor areas is made: a score of 1 is given when no blood flow can be found in the lesion; a score of 2 is given when only a small amount of flow can be detected; 3 is given when moderate flow is present and 4 is given when the adnexal mass appears highly vascular with marked blood flow using colour Doppler (abundant flow). This colour score refers only to the colour Doppler image and not to Doppler shift spectrum. It is given for the tumour as a whole (not for a solid part or a septum only, but for the whole tumour). Multiple photographic prints are made of relevant structures and Doppler signals.

#### Quality control

Several informative images or volumes of all adnexal masses should be made. Preferably, these are stored digitally. Photographs or video are acceptable as well.

#### Subjective assessment

After ultrasonographic examination of the mass the investigator gives his subjective assessment of the mass:

A: Malignant or benign or borderline?

B: Probability of malignancy: 1 = benign

(=level of certainty) 2 = probably benign

3 = uncertain

4 = probably malignant

5 = malignant

C: Self impression: presumed histological diagnosis (e.g. dermoid, serous cystadenoma, endometrioma, abscess…)

#### Surgical intervention

Surgery is performed according to local protocols. The reason for surgery, e.g. symptoms (pain, discomfort or pressure symptoms), raised serum CA125 levels or changes in the morphology or volume of the mass is recorded in the study screen.

#### Study screen

An astraia study screen will be used which will permit the entry of **multiple scans** per patient.

All centres will receive the **IOTA 5 study screen**.

At initial set up centres can choose between the two options:

Full IOTA 5 study (with planning of appropriate follow up and conservative management whenever feasible).

Observational study only (e.g. in radiology departments that are not involved in management decisions)

**Recorded variables (**entered in the **astraia study screen**).

**Patient data**

(click one option from list below)

New patient with diagnosis of adnexal mass

New patient who was already in follow up in your centre for adnexal mass before she was enrolled to the IOTA 5 study. How many months in follow up? …

Follow up scan of patient that is already enrolled to the IOTA 5 study before

**Ultrasound**

Spontaneous resolution of the adnexal mass (no further details are entered)

Adnexal mass present (fill in all variables below)

12 variables described in LR 1:

Age

personal history of ovarian cancer

personal history of breast cancer

Max diameter of lesion

Max diameter of solid component

Presence of ascites

Presence of blood flow within papillary projection

Irregular internal cyst walls

Presence of a purely solid tumour

Colour score (1/2/3/4)

Presence of acoustic shadows

Current hormonal therapy

Presence of pain during the examination

as well as simple rules, RMI and other variables:

Type of tumour (unilocular/unilocular-solid/multilocular /multiloc-solid/solid)

Ovarian crescent sign

Cyst content

Incomplete septum

Mobility: mobile/reduced mobility/completely fixed

Number of locules

Number of papillations (0/1/2/3/more)

Size of ovary

Bilateral tumour

Evidence of metastases

Menopausal status: premenopausal/postmenopausal

Serum CA 125 result (not mandatory)

Symptoms during the last year before ultrasound scan (multiple options are possible)

Pelvic pain

Postmenopausal bleeding

increased abdominal size

persistent abdominal distention (bloating)

appetite loss

constipation

diarrhoea

urinary urgency

urinary frequency

weight changes

dyspareunia

Other: please specify: ….

*For centres participating at the full IOTA 5 study only:* Suggested management recorded by examiner: "What type of management do you propose for this patient based on ultrasound and clinical data?"

Conservative management without follow up

Conservative management with follow up as specified in the protocol

Surgery by a gynaecologist or general surgeon

Surgery by an oncological surgeon

**Current status of the patient (this new tab should come before the tab "Histology"). A fixed query could be made to automatically ask the investigator about the status of all patients that were not rescanned 55 weeks after their previous scan as soon as the investigator opens the IOTA 5 study screen.**

Lost to follow up (no other pop-up)

Patient stopped participating to the study (please specify why : …..) (no other pop-up)

Patient withdrew her consent (data cannot be used for statistical analysis and no reason is aked) (no other pop-up)

Surgery performed (pop-up of fields about operation below)

<u>If Surgery</u>

Date of operation:

Type of operation: cystectomy or oophorectomy or staging etc

Laparotomy with vertical incision

Laparotomy with horizontal incision

Operative laparoscopy

Diagnostic laparoscopy

Primary chemotherapy

Indication for operation (more than one possibility may be thicked)

Suspicion of malignancy based on ultrasound

Suspicion of malignancy based on other information if so what?

Malignancy cannot be excluded

Acute pain

Chronic pain

Suspected torsion

Fertility concerns

Patient request

Increase in size of the tumour

Change in morphology of the tumour

Increase in CA 125 level

Indicated by other imaging technique (CT, MRI…)

Other doctor recommended operation. Please specify the reason: …

“en passent” removal of the mass when patient was operated for another indication

Other: Specify: ….

Findings at operation (more than one option may be ticked)

No complications of the tumour

Torsion

Rupture

Inflammation/Infection

Adhesions

Bleeding from tumour

Metastatic cancer

Other complications of tumour: specify: ….

Other non-gynaecological pathology (e.g. appendicitis): specify: ….

Complications during operation (within one week of surgery). (more than one option may be ticked) :

Conversion from laparoscopy to laparotomy

Bowel perforation

Bleeding requiring transfusion

Embolism, deep venous thrombosis

Wound Infection

Peritonitis

Other: Specify: ….

Histological diagnosis (pop up list as before) with open text area ("Details")

<u>If follow up examination:</u>

Complications of adnexal mass: no /yes

If yes : date of first complication: ….

And pop up list (you can select more than one):

Acute pain

Chronic pain

Suspected torsion

Infection

Haemorrhage

Rupture

Other, please specify: ….

Death: date

Death directly or indirectly relatd to adnexal mass?

cause of death (

Autopsy findings: specify: ….

Not needed for IOTA phase 5: second stage tests

#### Consent / information leaflet

Information leaflets are at the discretion of the participating centres. Approval of the local Ethical Committee for clinical studies is necessary.

#### Serum tumour markers

Serum CA 125 measurements or other tumour markers are performed locally, using a CA 125 II immunoradiometric assay

#### Tissue collection

Preferably the whole tumour should be removed. However, representative biopsies may be sufficient (e.g. in advanced ovarian cancer or endometrioma).

#### Tumour classification

Tumours are classified according to the criteria recommended by the International Federation of Gynecology and Obstetrics (FIGO). In malignant tumours the degree of differentiation is included.

#### Statistical analysis

For the first objective, a descriptive analysis of complications on all data and stratified for participating center (anonymized) will be performed, as well as an overall Kaplan-Meier curve of 'complication free survival'. Complication free survival is defined as the time to the need for surgery during long-term follow-up. Patients that did not need surgery at the end of the follow-up period are right censored at the time of the last examination.

For the second objective, the discriminatory ability of LR1 and the polytomous model to detect malignancy at the initial visit will be assessed. To this end, the logit of the risk of malignancy given by LR1, logit(LR1), is used to predict malignancy using logistic regression, and the performance assessed with the odds ratio, the c-index and a ROC curve. Linearity of the effect of logit(LR1) will be assessed using spline functions. This analysis is planned one year after the end of the inclusion phase of the trial. Non-operated masses will be classified as benign at the initial visit if there is an absence of a clinical diagnosis of cancer after 1 year of follow up. In addition, the ability of LR1 to predict complications during long-term follow-up will be assessed. This will be done using Cox proportional hazards regression with the logit(LR1) as predictor of complication free survival. Performance will be assessed using the hazard ratio and the c-index within the context of survival analysis. Linearity of the effect of logit(LR1) will be investigated using Schoenfeld residuals.

For the third objective, a multivariable survival analysis will be undertaken using Cox proportional hazards regression or more complex alternatives based on support vector machines (30). The models will be penalized to prevent overfitting, given that not enough events (i.e., patients that need surgery during long term follow-up) are expected for the number of available predictor variables. Internal validation will be assessed through bootstrapping rather than a split of the data in training and test sets (31).

For the fourth objective, longitudinal normative curves will be derived of the changes seen in the characteristics of non-operated masses. Longitudinal analysis techniques such as mixed models, longitudinal support vector machines, and functional linear discriminant analysis (FLDA) (32-34).

#### Study supervision

Central supervision: the Steering Committee is responsible for the protocol, quality control, interim analyses of the data and final analysis and reporting of the study.

Local supervision: the Principal Investigators are responsible for the data collection in their centres.

Dirk Timmerman is responsible for the co-ordination of the overall IOTA project and the contact between the centres.

Sabine Van Huffel and Bart De Moor are responsible for the data management and the development of new algorithms, in collaboration with Ben Van Calster, Lieveke Ameye and Kirsten Van Hoorde.

**Publication policy**

The steering committee is responsible for publication of the data in scientific journals. As such the members are co-authors in all resulting clinically relevant papers, to which they made significant contributions. By the time of the final analysis the principal investigators have to have contributed at least 50 cases to the study. They are co-authors, according to the number of patients they contributed to the study (depending on the journal's restriction of the number of co-authors) on condition that they contribute to writing the papers and read and approve the final version.

Purely mathematical papers without clinical relevance related to the study data are published by S. Van Huffel, B. De Moor and co-workers at ESAT with reference to the IOTA group and the inclusion of as many as possible of the clinical contributors.

The Katholieke Universiteit Leuven represented by its department K.U.LEUVEN RESEARCH & DEVELOPMENT, having its office in 3000 Leuven, Minderbroedersstraat 8A – box 5105, Belgium, VAT number BE 419.052.173 holds intellectual property rights that might result from the IOTA project.

**References (Study protocol)**

Timmerman D, Testa AC, Bourne T, Ferrazzi E, Ameye L, Konstantinovic ML, Van Calster B, Collins WP, Vergote I, Van Huffel S, Valentin L. Logistic regression model to distinguish between the benign and malignant adnexal mass before surgery: a multicenter study by the International Ovarian Tumor Analysis Group. J Clin Oncol 2005; 23:8794-8801.

Van Holsbeke C, Van Calster B, Testa AC, Domali E, Lu C, Van Huffel S, Valentin L, Timmerman D. Prospective internal validation of mathematical models to predict malignancy in adnexal masses: results from the Internation Ovarian Tumor Analysis study. Clin Cancer Res 2009; 15:684-691.

Timmerman D, Van Calster B, Testa AC, Guerriero S, Fischerova D, Lissoni AA, Van Holsbeke C, Fruscio R, Czekierdowski A, Jurkovic D, Savelli L, Vergote I, Bourne T, Van Huffel S, Valentin L. Ovarian cancer prediction in adnexal masses using ultrasound based logistic regression models: a temporal and external validation study by the IOTA group. Ultrasound Obstet Gynecol 2010; 36(2):226-34.

Menon U, Gentry-Maharaj A, Hallett R, Ryan A, Burnell M, Sharma A, Lewis S, Davies S, Philpott S, Lopes A, Godfrey K, Oram D, Herod J, Williamson K, Seif MW, Scott I, Mould T, Woolas R, Murdoch J, Dobbs S, Amso NN, Leeson S, Cruickshank D, McGuire A, Campbell S, Fallowfield L, Singh N, Dawnay A, Skates SJ, Parmar M, Jacobs I. Sensitivity and specificity of multimodal and ultrasound screening for ovarian cancer, and stage distribution of detected cancers: results of the prevalence screen of the UK Collaborative Trial of Ovarian Cancer Screening(UKCTOCS). Lancet Oncol. 2009 Apr;10(4):327-40.

Hoo WL, Yazbek J, Holland T, Mavrelos D, Tong EN, Jurkovic D. Expectant management of ultrasonically diagnosed ovarian dermoid cysts: is it possible to predict outcome? Ultrasound Obstet Gynecol. 2010 Aug;36(2):235-40.

Caspi B, Appelman Z, Rabinerson D, Zalel Y, Tulandi T, Shoham Z. The growth pattern of ovarian dermoid cysts: a prospective study in premenopausal and postmenopausal women. Fertil Steril. 1997 Sep;68(3):501-5. PubMed PMID: 9314922.

Caspi B, Levi R, Appelman Z, Rabinerson D, Goldman G, Hagay Z. Conservative management of ovarian cystic teratoma during pregnancy and labor. Am J Obstet Gynecol. 2000 Mar;182(3):503-5. PubMed PMID: 10739498.

Valentin L, Akrawi D. The natural history of adnexal cysts incidentally detected at transvaginal ultrasound examination in postmenopausal women. Ultrasound Obstet Gynecol. 2002 Aug;20(2):174-80.

Valentin L, Skoog L, Epstein E. Frequency and type of adnexal lesions in autopsy material from postmenopausal women: ultrasound study with histological correlation. Ultrasound Obstet Gynecol. 2003 Sep;22(3):284-9.

Jemal A, Siegel R, Ward E, Hao Y, Xu J, Thun MJ. Cancer statistics 2009. CA Cancer J Clin 2009; 59:225-249.

Cancer Research UK a. UK ovarian cancer incidence statistics. http://info.cancerresearchuk.org/cancerstats/types/ovary/incidence/ (accessed Oct 7, 2009).

Horner MJ, Ries LAG, Krapcho M, Neyman N, Aminou R, Howlader N, Altekruse SF, Feuer EJ, Huang L, Mariotto A, Miller BA, Lewis DR, Eisner MP, Stinchcomb DG, Edwards BK (eds.) SEER Cancer statistics review 1975-2006, NCI, Bethesda, MD.

Cancer Research UK b. UK Ovarian cancer survival statistics. http://info.cancerresearchuk.org/cancerstats/types/ovary/survival/ (accessed Oct 7, 2009).

Engel J, Eckel R, Schubert-Fritschle G, et al. Moderate progress for ovarian cancer in the last 20 years: prolongation of survival, but no improvement in cure rate. Eur J Cancer 2002; 38:2435-45.

Vergote I, Amant F, Ameye L, Timmerman D. Screening for ovarian carcinoma: not quite there yet (editorial). Lancet Oncology 2009; 10:308-309.

Crayford TJB, Campbell S, Bourne TH, Rawson HJ, Collings WP. Benign ovarian cysts and ovarian cancer: a cohort study with implications for screening. Lancet 2000; 355:1060-1063.

Timmerman D. The use of mathematical models to evaluate pelvic masses: can they beat an expert operator? Best Pract Res Clin Obstet Gynaecol 2004; 18:91-104.

Van Holsbeke C, Van Calster B, Valentin L, Testa AC, Ferrazzi E, Dimou I, Lu C, Moerman Ph, Van Huffel S, Vergote I, Timmerman D. External validation of mathematical models to distinguish between benign and malignant adnexal tumors: a multicenter study by the International Ovarian Tumor Analysis Group, Clin Cancer Res 2007; 13:4440-4447.

Geomini P, Kruitwagen R, Bremer GL, Cnossen J, Mol BW. The accuracy of risk scores in prediting ovarian malignancy: a systematic review. Obstet Gynecol 2009; 113:384-394.

Valentin L, Jurkovic D, Van Calster B, Testa A, Van Holsbeke C, Bourne T, Vergote I, Van Huffel S, Timmerman D. Adding a single CA 125 measurement to ultrasound imaging performed by an experienced examiner does not improve preoperative discrimination between benign and malignant adnexal masses. Ultrasound Obstet Gynecol. 2009 Sep;34(3):345-54. PubMed PMID: 19585547.

Van Calster B, Timmerman D, Bourne T, Testa AC, Van Holsbeke C, Domali E, Jurkovic D, Neven P, Van Huffel S, Valentin L. Discrimination between benign and malignant adnexal masses by specialist ultrasound examination versus serum CA-125. J Natl Cancer Inst. 2007 Nov 21;99(22):1706-14. Epub 2007 Nov 13. PubMed PMID: 18000221.

Valentin L. Pattern recognition of pelvic masses by gray-scale ultrasound imaging: the contribution of Doppler ultrasound. Ultrasound Obstet Gynecol. 1999 Nov;14(5):338-47.

Sokalska A, Timmerman D, Testa AC, Van Holsbeke C, Lissoni AA, Leone FP, Jurkovic D, Valentin

L. Diagnostic accuracy of transvaginal ultrasound examination for assigning a specific diagnosis to adnexal masses. Ultrasound Obstet Gynecol. 2009 Oct;34(4):462-70.

Timmerman D, Valentin L, Bourne TH, Collins WP, Verrelst H, Vergote I. Terms, definitions and measurements to describe the sonographic features of adnexal tumors: a consensus opinion from the International Ovarian Tumor Analysis (IOTA) group. Ultrasound Obstet Gynecol 2000; 16:500-505.

Van Calster B, Timmerman D, Lu C, Suykens JAK, Valentin L, Van Holsbeke C, Amant F, Vergote I, Van Huffel S. Preoperative diagnosis of ovarian tumors using Bayesian kernel-based methods. Ultrasound Obstet Gynecol 2007; 29:496-504.

Van Calster B, Timmerman D, Nabney IT, Valentin L, Testa AC, Van Holsbeke C, Vergote I, Van Huffel

S. Using Bayesian neural networks with ARD input selection to detect malignant ovarian masses prior to surgery. Neural Comput Applic 2008; 17:489-500.

Ameye L, Valentin L, Testa AC, Van Holsbeke C, Domali E, Van Huffel S, Vergote I, Bourne T, Timmerman D. A scoring system to differentiate malignant from benign masses in specific ultrasound-based subgroups of adnexal tumors. Ultrasound Obstet Gynecol 2009; 33:92-101.

Van Holsbeke C, Van Calster B, Bourne T, Melis GB, Testa AC, Guerriero S, Fruscio R, Lissoni AA, Czekierdowski A, Savelli L, Van Huffel S, Valentin L, Timmerman D. External validation of diagnostic models to predict the risk of malignancy in adnexal masses. Submitted.

Leone FP, Crepaldi A, Marciante C, Cetin I. Sonographic follow-up of unilocular >5 cm and multilocular ovarian cysts <7 cm in post-menopausal women: preliminary results. OC21.05. Ultrasound Obstet Gynecol 2011; 38 (Suppl. 1): 40.

Van Belle V, Pelckmans K, Suykens JAK, Van Huffel S. Additive survival least squares support vector machines. Stat Med 2010**;** 29:296-308.

Steyerberg EW, Harrell FE Jr, Borsboom GJ, Eijkemans MJ, Vergouwe Y, Habbema JDF. J Clin Epidemiol 2001; 54:774-781.

Verbeke G, Molenberghs G. Linear mixed models for longitudinal data. Springer: New York, 2000.

Luts J, Molenberghs G, Verbeke G, Van Huffel S, Suykens JAK. A mixed effects least squares support vector machine model for classification of longitudinal data. Internal Report 10 -249, ESAT-SISTA, K.U.Leuven (Leuven, Belgium), 2010.

James GM, Hastie TJ. Functional linear discriminant analysis for irregularly sampled curves. J R Stat Soc B 2001;63:533–550.
